# A core outcome set for adult general ICU patients

**DOI:** 10.1101/2024.05.29.24308094

**Authors:** Maj-Brit Nørregaard Kjær, Camilla Rahbek Lysholm Bruun, Anders Granholm, Morten Hylander Møller, Bodil Steen Rasmussen, Camilla Bekker Mortensen, Lone Musaeus Poulsen, Thomas Strøm, Eva Laerkner, Anne Craveiro Brøchner, Trine Haberlandt, Anne-Marie Gellert Bunzel, Louise Stenbryggen Herløv, Anna Holm, Praleene Sivapalan, Stine Estrup, Maria Cronhjort, Anna Schandl, Jon Henrik Laake, Kristin Hofsø, Fredrike Blokzijl, Frederic Keus, Carmen Andrea Pfortmueller, Marlies Ostermann, Jade M Cole, Matt P Wise, Wojciech Szczeklik, Anna Wludarczyk, Tomas Jovaiša, Maurizio Cecconi, Martin Ingi Sigurdsson, Marek Nalos, Johanna Hästbacka, Marja Mäkinen, Naomi Hammond, Edward Litton, Kimberley Haines, Sheila Nainan Myatra, Bharath Kumar Tirupakuzhi Vijayaraghavan, Kavita Yadav, Vivekanand Jha, Balasubramanian Venkatesh, Ingrid Egerod, Anders Perner, Marie O Collet

## Abstract

**Purpose:** Randomised clinical trials should ideally use harmonised outcomes that are important to patients and to facilitate meta-analyses and ensuring generalisability. Core outcome sets for specific subsets of ICU patients exist, e.g., respiratory failure, delirium, and COVID-19, but not for ICU patients in general. Accordingly, we aimed to develop a core outcome set for adult general ICU patients.

**Methods:** We developed a core outcome set in Denmark following the Core Outcome Measures in Effectiveness Trials (COMET) Handbook. We used a modified Delphi consensus process with multiple methods design, including literature review, survey, semi-structured interviews, and discussions with initially five Danish research panels, involving adult ICU survivors, family members, clinicians, and researchers. The core outcome set was internationally validated in local panels in 14 countries and revised accordingly.

**Results:** We identified 329 published outcomes, of which 50 were included in the 264 participant Delphi survey. After 82 semi-structured survey participant interviews no additional outcomes were added. The first survey round was completed by 249 (94%) participants, and 202 (82%) contributed to the final third round. The initial core outcome set comprised six core outcomes. International validation involved 217 research panel members and resulted in the final core outcome set of survival, free of life support, free of delirium, out of hospital, health-related quality of life, and cognitive function.

**Conclusions:** We developed and internationally validated a core outcome set with six core outcomes to be used in research, specifically clinical trials involving adult general ICU patients.

## Introduction

Outcomes assessed in randomised clinical trials (RCTs) should ideally be important to patients and used consistently across trials to facilitate comparisons, generalisability, and valid meta-analyses to inform clinical guidelines [1, 2]. Acutely admitted intensive care unit (ICU) patients have a high mortality [3–5], hence this outcome is frequently used and often hypothesised to be affected by the interventions assessed in clinical trials in the ICU setting [6]. However, ICU survivors often report experiencing persistent physical, cognitive, and mental impairments [7–9], aspects that are not covered by survival alone.

Outcome choices and definitions in critical care RCTs vary substantially, as illustrated by a recent scoping review revealing 103 distinct outcomes for assessing functional, neurological, and cognitive aspects and 29 distinct outcomes for health-related quality of life (HRQoL) [10]. The inconsistency in choices of outcomes and the definitions across RCTs involving ICU populations presents a challenge for comparison of trial results and evidence synthesis [10–13]. Therefore, it is important to establish a standardised approach to outcomes and measurements in [11, 14–16]. Involvement of patients, family members, clinicians, and researchers adds substantial value to discussions and prioritisation of outcomes [17–19].

Several core outcomes set (COS) have been developed for specific subsets of the ICU population, such as acute respiratory failure, delirium, or COVID-19, or for interventions, e.g., rehabilitation. However, there is no COS for the broad population of adult general ICU patients [7, 20–23], who do not fit into specific subsets, and may have a range of impairments.

The Intensive Care Platform Trial (INCEPT, www.incept.dk) will focus on assessing frequently used interventions in acutely ill ICU patients and has a need for a COS for the general ICU patient.

The aim of this study was to develop and internationally validate a generic COS for adult general ICU patients, regardless of the intervention assessed in the trials and involving a diversity of patients, family members, clinicians, and researchers with lived ICU experience.

## Methods

We conducted a study using multiple methods design to develop a COS. Our approach incorporated a modified Delphi consensus process, and integrated a literature review, surveys, semi-structured interviews, and international external validation. The work took place between February 2021 and February 2024; the study was registered in the Core Outcome Measures in Effectiveness Trials (COMET) database (https://comet-initiative.org/Studies/Details/1882), and conducted following a published protocol [24]. The COS was developed in accordance with the COMET initiative [11] and reported according to the Core Outcome Set-STAndards for Reporting (COS-STAR) Statement [25], and Guidance for Reporting Involvement of Patients and the Public short form [26] (checklists in the electronic supplementary material (ESM)).

In line with the publicly registered protocol amendment [27], we conducted an international (external) validation of the initial COS. This involved international sites from the Collaboration for Research in Intensive Care (CRIC) network and its collaborators. The international validation process was initiated in November 2023 and completed in February 2024.

## Participants

### Stakeholder involvement

The steering committee (listed in author contributions) established research panels in Denmark to facilitate the involvement of persons with lived experience being adult ICU survivors (patients), family members, multi professional health care workers (clinicians), and researchers. The Danish research panels actively participated in condensing outcomes identified in the literature review. They were involved in developing survey questions, ensuring their cognitive validity through initial and inter-survey evaluations. Additionally, international research panels were established for the international validation (Table S1, ESM).

### Survey and interview

We invited patients, family members, clinicians, and researchers from Denmark to participate in the Delphi survey and/or interviews. We aimed to include 400 participants: 100 patients, 100 family members and 200 multi professional ICU clinicians and researchers in the survey [24]. In addition, we targeted a total of 90 individual interviews: 30 patients, 30 family members, and 30 ICU clinicians or researchers. We recruited ICU survivors and family members directly in ICUs and via other ongoing research projects [24]. Clinicians and researchers were recruited via the steering committees’ clinical- and research network. The sampling of participants aimed at balancing the distribution considering age, sex, ethnicities, admission type, risk of mortality above and below 25% (defined by the Simplified Mortality Score for the Intensive Care Unit (SMS- ICU) score [28]), ICU length of stay, and time since ICU stay (details in ESM). We collected demographic and clinical data for the survey participants and interviewees (Table S2-S3, ESM) [24].

### Process

Figure 1 illustrates the process for developing a COS involving a modified Delphi consensus process (step 1- 4) and an additional internationally validation of the COS (step 5-6).

**Figure 1.**
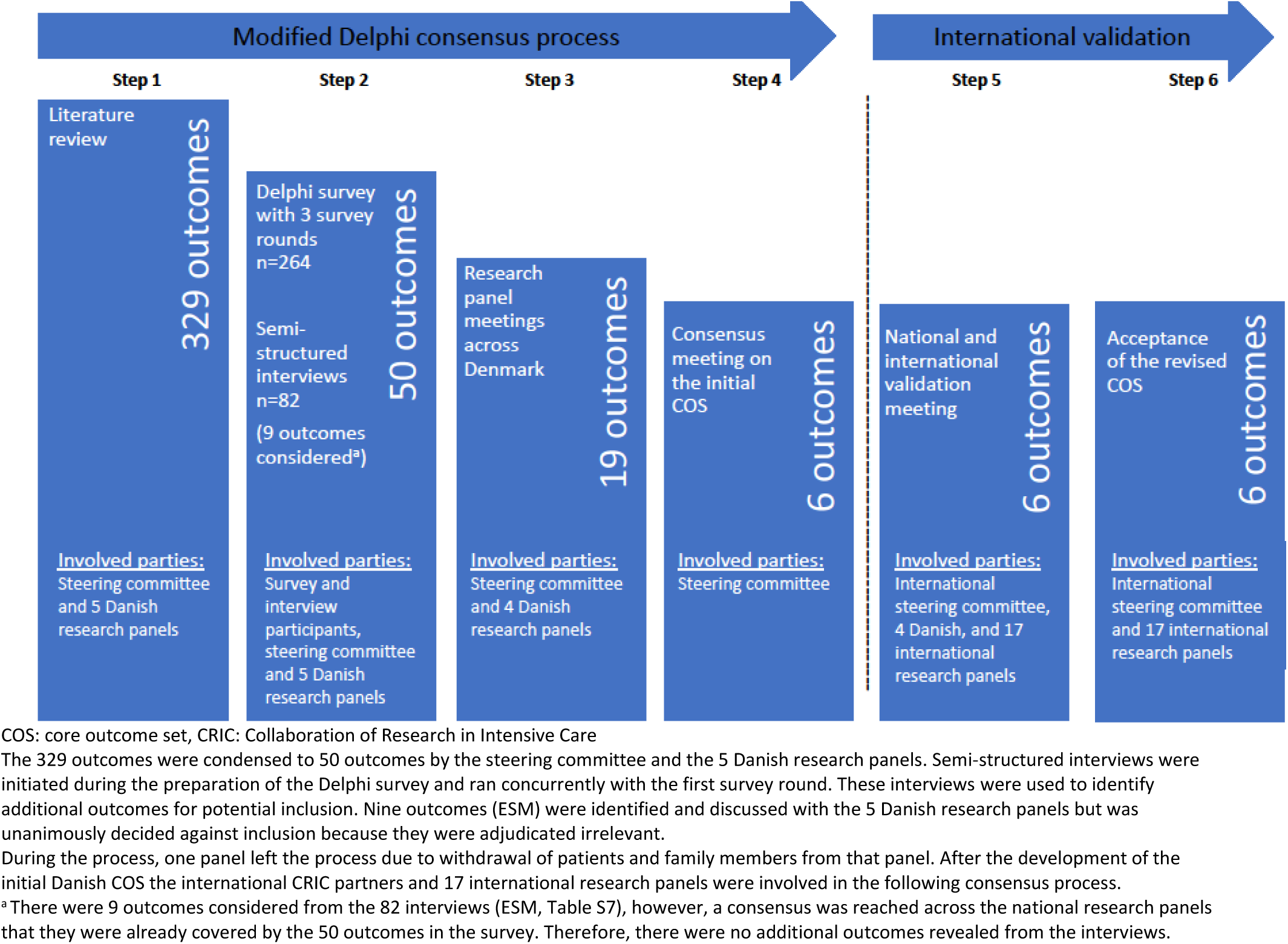

**Figure 2.**
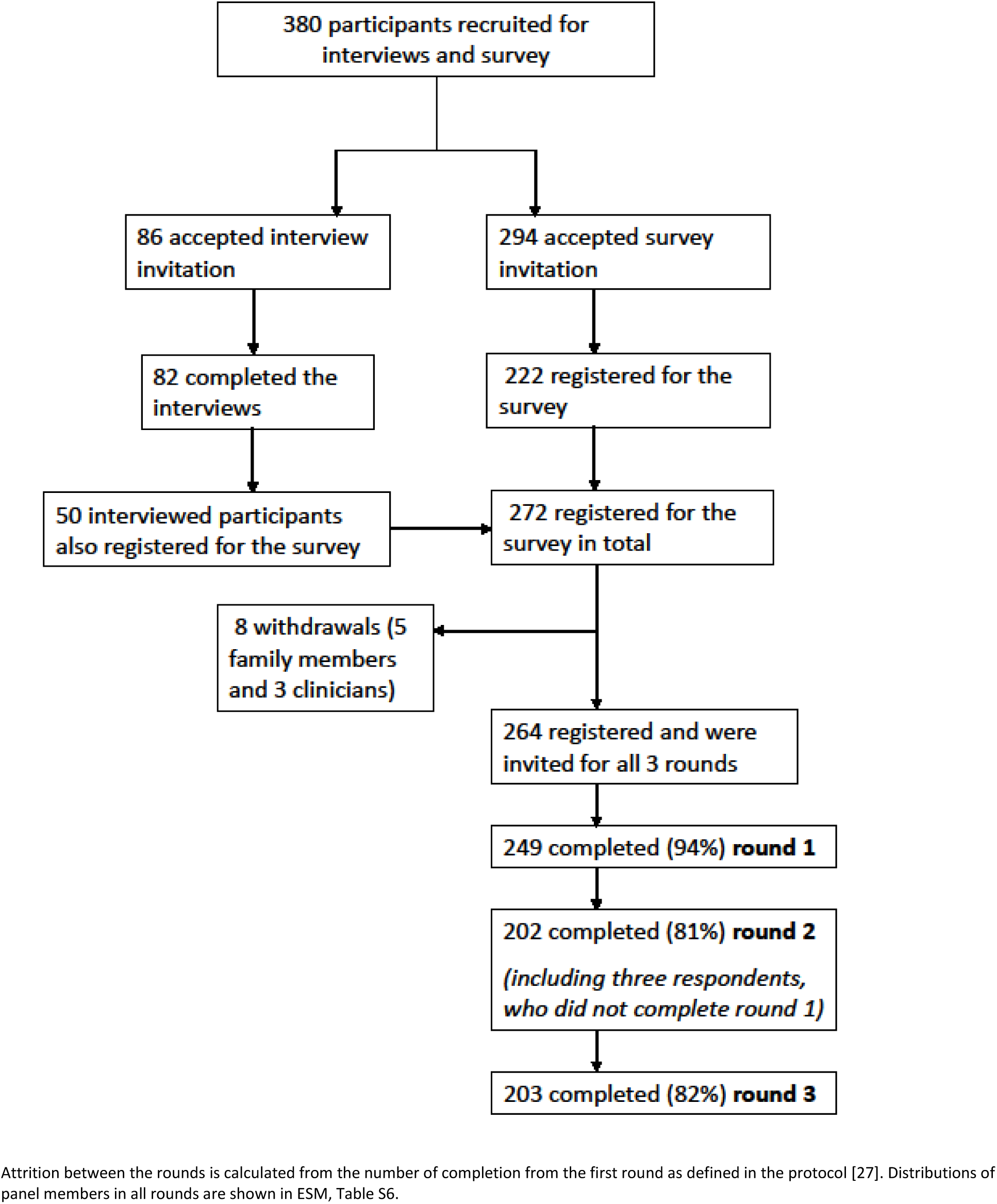
Flow diagram for the participants being interviewed and registered for the Delphi survey.

#### Step 1

Two authors (MNK and SE) independently and in duplicate conducted an initial literature review to identify relevant outcomes, results were compared and merged to one list of outcomes (search string and full list of outcomes in the ESM, Table S4) [24].

We conducted multiple meetings in Danish research panels to identify important outcomes, using the nominal group technique with face-to-face discussion in small groups aiming for immediate consensus [29]. These meetings helped to identify the most essential outcomes to inform the survey [29].

#### Step 2

Meetings were held between survey rounds to reach consensus on removal and inclusion of outcomes suggested in the first survey round. Participants in the Delphi survey rated the importance of an outcome on a Likert scale from 1-9 [11]. Ratings were categorised as follows: 1-3 the outcome was considered ‘not important’, 4-6 ‘important, but not critical’, and 7-9 ‘critical’ [24].

### Consensus definition

Within each stakeholder group (patients, family members, clinicians, and researchers) consensus for an outcome was achieved when two criteria were met: 1) ≥70% of the participants rated the outcome as ‘critical’ (a score ≥7), and 2) no more than ≤15% of the participants rated the outcome ‘not important’ (≤3) [11, 24]. In the second and third survey rounds, participants were provided with their own score, the score from all four groups, and a summary score.

#### Step 3

Following the final survey round, four research panels (the fifth research panel withdrew due to no patients in the panel at the time) met to reach consensus on the most important outcomes.

#### Step 4

Subsequently, the steering committee convened to obtain consensus on the initial COS following review of comments on several suggested outcomes from the research panels.

#### Step 5

The international steering committee (listed in author contributions) familiarised themselves with the modified Delphi consensus process. This involved studying the condensed outcomes in the survey, reviewing the survey results, and understanding the rationale behind the initial COS (ESM, page 23). Investigators from each country established local research panels (Figure 1, Step 5). International research panels were briefed on the initial COS and its underlying rationale. Utilising the nominal group technique with face-to-face discussion in smaller groups [29], all countries discussed each single core outcome and reached a consensus on whether to adopt them as they were, adapt them (i.e., accept with changes), or reject them. Minutes from the meetings are presented in the ESM Table S5. Two online meetings, due to time zones, including the international steering committee, was convened on January 30, 2024, to address all issues raised during the international validation process.

### Consensus definition

For the international validation, a predefined cut-off value for consensus was not established. However, the protocol amendment outlined, a transparent process for all participating countries, ensuring that any issues raised during the consensus process discussions were documented [27].

#### Step 6

The steering committee considered the feedback from the international research panels and revised the wording of the COS accordingly. The revised COS was then reassessed by the Danish and international research panels and approved.

### Analysis

For the qualitative analysis, the semi-structured interviews were transcribed (Microsoft Word 2016, Microsoft Corporation, Redmond, Washington), and were then reviewed by the research team (MNK, CRLB, AMGB, AH, MOC) who identified potentially new outcomes. All survey rounds in the Delphi consensus process were delivered electronically using the web based DelphiManager software version 5.0 (COMET, University of Liverpool, Liverpool; www.comet-initiative.org/delphimanager). For statistical analyses, we used R (R Core Team, Foundation for Statistical Computing, Vienna, Austria) version 4.2.2, and Microsoft Excel 2016 (Microsoft Corporation, Redmond, Washington).

We used descriptive statistics to report the population, ratings, and responses of survey rounds with frequencies and proportions for categorical data and medians and interquartile ranges for numeric data. Less than 20% of data were missing. As per protocol, missing data were handled by carrying the last observation forward (further details in the ESM, Table S6) [24].

### Ethics and consent

All Danish participants in the survey and interviews provided written informed consent either by signature (interviews) or ticking confirmed consent in the survey registration. All data were handled confidentially, and participants could withdraw their consent anytime. The Danish Data Protection Agency and Ethical Committee for the Capital Region waived the need for ethical committee assessment (H-21010116).

## Results

### Participants

A total of 380 participants took part in the Delphi survey, of whom 90 were invited for interviews by quota sampling [30]; 82 completed the interviews. The inclusion flow and response rate are presented in Figure 2 and characteristics of the survey participants are presented in Table 1 (all characteristics are presented in ESM, Table S2-S3).

**Table 1.** Characteristics of stakeholders in the Delphi survey and the semi-structured interviews.

| Characteristics of stakeholders for the first round of the Delphi survey (n=264) |  |  |  |  |
| --- | --- | --- | --- | --- |
|  | Patients<br>n=65 | Family members<br>n=49 | Clinicians<sup>e</sup><br>n=136 | Researchers<sup>f</sup><br>n=14 |
| Age (years) | 62 (50 to 70) | 59 (47 to 65) | 46 (39 to 52) | 41 (37 to 48) |
| Sex (female) | 21 (32%) | 34 (69%) | 93 (68%) | 9 (64%) |
| Other ethnicity than Danish<sup>a</sup> | 4 (6%) | 1 (2%) | 5 (4%) | 3 (21%) |
| Admission type |  |  |  |  |
| - Surgical | 29 (47%) | - | - | - |
| - Medical | 33 (53%) | - | - | - |
| SMS-ICU<sup>b</sup> at ICU admission<sup>c</sup> | 20 (13.5 to 22.0) | - | - | - |
| - <17 point | 23 (37%) | - | - | - |
| - ≥17 point | 40 (63%) | - | - | - |
| Self-reported critical illness severity<sup>d</sup> | 10.0 (8.5 to 10.0) | - | - | - |
| ICU length of stay (days)<sup>c</sup> | 9.0 (6.0 to 22.5) | - | - | - |
| Time since ICU stay (months)<sup>c</sup> | 14 (3 to 20) | - | - | - |
| Assistance to fill in the survey | 11 (17%) | - | - | - |
| Family to a deceased patient | - | 5 (10%) | - | - |
| Characteristics of stakeholders participating in the semi-structured interviews (n=82) |  |  |  |  |
|  | Patients<br>n=27 | Family members<br>n=25 | Clinicians<sup>e</sup><br>n=26 | Researchers<sup>f</sup><br>n=4 |
| Age (years) | 60 (50 to 68) | 54 (41 to 60) | 45 (43 to 48) | 43 (38 to 48) |
| Sex (female) | 13 (48%) | 18 (72%) | 21 (81%) | 4 (100%) |
| Other ethnicity than Danish<sup>a,g</sup> | 1 (4%) | 1 (4%) | 3 (10%) | 0 (0%) |
| Admission type |  |  |  |  |
| - Surgical<sup>b</sup> | 13 (48%) | - | - | - |
| - Medical<sup>b</sup> | 14 (52%) | - | - | - |
| SMS-ICU<sup>b</sup> at ICU admission<sup>c</sup> | 19 (15 to 22) | - | - | - |
| - <17 points | 10 (37%) | - | - | - |
| - ≥17 points | 17 (63%) | - | - | - |
| Self-reported critical illness severity<sup>d</sup> | 9 (8 to 10) | - | - | - |
| ICU length of stay (days)<sup>c</sup> | 9 (7 to 25) | - | - | - |
| Time since ICU admission (months)<sup>c</sup> | 11 (4 to 17) | - | - | - |
| Family to a deceased patient | - | 3 (12%) | - | - |

ICU: intensive care unit, SMS-ICU: The Simplified Mortality Score for the Intensive Care Unit [28].
Numeric data are presented as medians with interquartile ranges, categorical data as counts with percentages.
<sup>a</sup> Other than Danish origins covers both 'immigrants' (born abroad and neither parent being a Danish citizen or born in Denmark) and 'descendants' (born in Denmark and neither of the parents is a Danish citizen or born in Denmark) (Statistics Denmark, www.dst.dk). Danish origin means at least one parent born in Denmark with Danish citizenship (Statistics Denmark, www.dst.dk).
<sup>b</sup> We aimed for a balanced distribution of patients with higher mortality risk ≥17 (predicted 90-day mortality risk of 25.3% and above) vs. lower risk <17 (predicted 90-day mortality risk of 22.8% and below) [24, 28]. Further details in the methods section.
<sup>c</sup> These variables were obtained from the patients' medical records. For the survey there were two patients with missing medical record data. One withdrew consent, one where there were only paper medical records, which couldn't be located. No missing data for patients interviewed.
<sup>d</sup> A numeric score (0-10 where 0 is non-critical and 10 is most critical). From either the patients' view or from the view of family members to ICU patients.
<sup>e</sup> Multi professional clinicians cover doctors, nurses, physiotherapists, occupational therapists, and others (ESM, Table S2 and S3).
<sup>f</sup> Multi professional researchers (ESM, Table S2 and S3).
<sup>g</sup> Missing data for 2 patients, 1 family member, and 1 clinician.

**Table 2.**
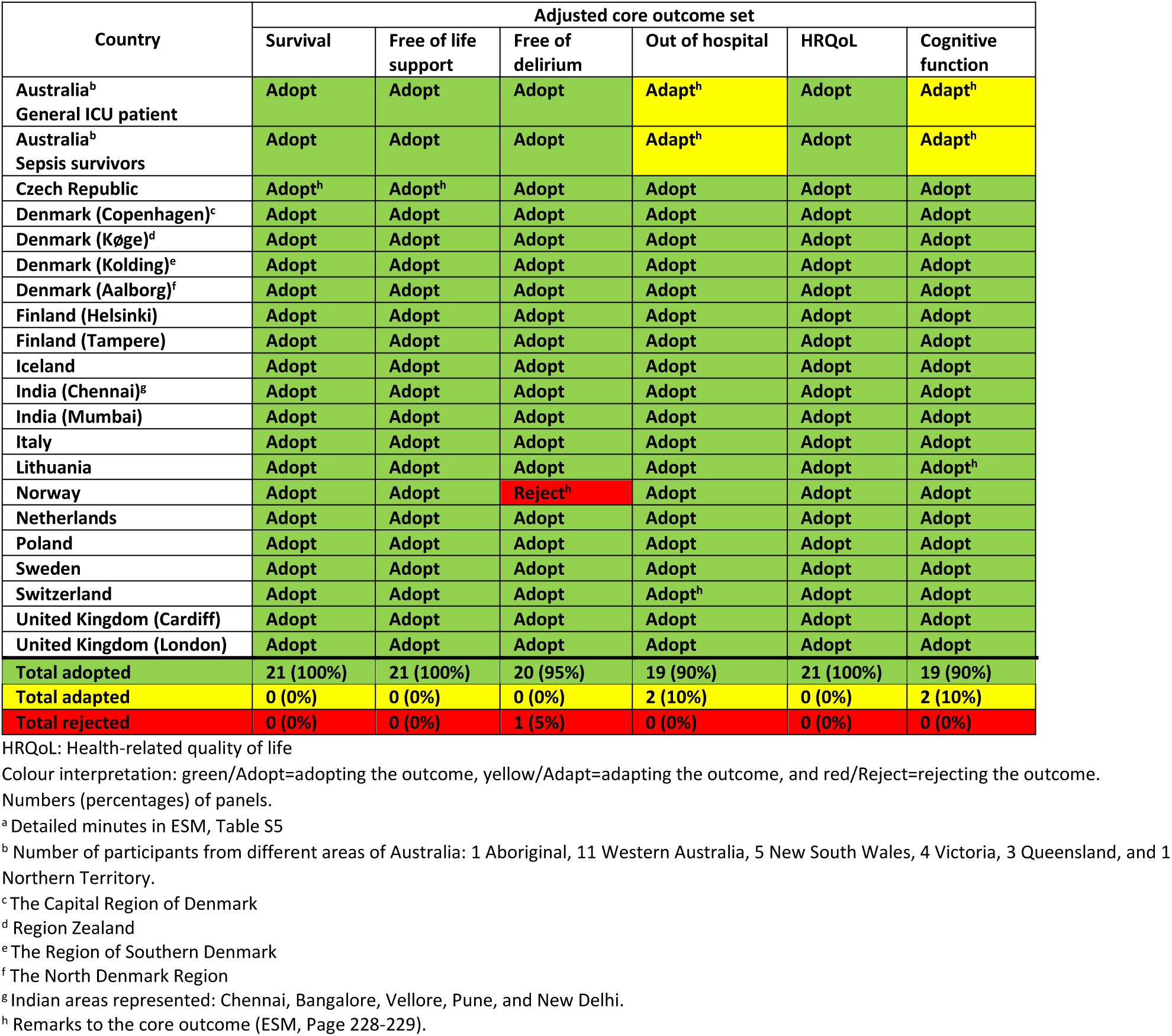
International validation of the core outcome set across the participating countries.

A total of 217 research panel members participated in the development of the COS (ESM, Table S1). The 4 research panels in Denmark had a total of 39 members (47 in the 5 initial panels) consisting of 8 patients, 5 family members, 17 clinicians, and 9 researchers. The 17 international research panels had a total of 178 members consisting of 46 patients, 26 family members, 60 clinicians, and 46 researchers (ESM, Table S4).

### Step 1

First, the 329 outcomes from the literature review were screened for duplicates by MNK, CBM, PS, SE, LMP, EL, and MOC and with thematic analysis the outcomes were condensed in categories and translated to Danish. The Danish research panels reached consensus on condensing the outcomes 50 outcomes, more details in the ESM.

### Step 2

The 82 interviews provided 9 additional outcomes to be considered (ESM, Table S7), but the Danish research panels concluded that they were already covered by the initial 50 outcomes.

In response to comments received during the first survey round and uncertainties raised by our Danish research panels, we adjusted the second survey round while maintaining the same set of 50 outcomes (ESM, Table S8). For all three rounds, we generated graphs depicting the scores on the Likert scale from 1-9 for each outcome and distributed them to all survey participants (Figure S1-S150, ESM).

Overall, the 50 outcomes were all considered important for inclusion (Figure S151, ESM). There was more variation among clinicians compared with patients and family members (Figure S152, ESM). The largest decline in response rates from the first to the final round was among family members and clinicians (Table S6, ESM).

### Step 3

Informed by the survey findings, the Danish research panels convened at separate consensus meetings. Each panel, prioritized between 9 and 13 outcomes for inclusion in the COS (Figure 1 and ESM, Table S8). After removing overlaps, a total of 19 unique outcomes emerged. One additional outcome, suggested by one research panel, was included to be assessed by the steering committee for the initial COS.

### Step 4

From the 19 outcomes, the steering committee agreed on six core outcomes, including the four that were deemed ‘critical’ by all Danish panels, thus defining the initial, Danish COS (Figure 3). Other outcomes were discussed, such as ‘health-economic consequences of a treatment in ICU’, ‘discharge location after hospitalisation’, and particularly ‘overall well-being’. ’Well-being’ may be considered part of HRQoL after ICU, and there was consensus not to include the additional discussed outcomes in the COS (ESM, Table S9).

**Figure 3.**
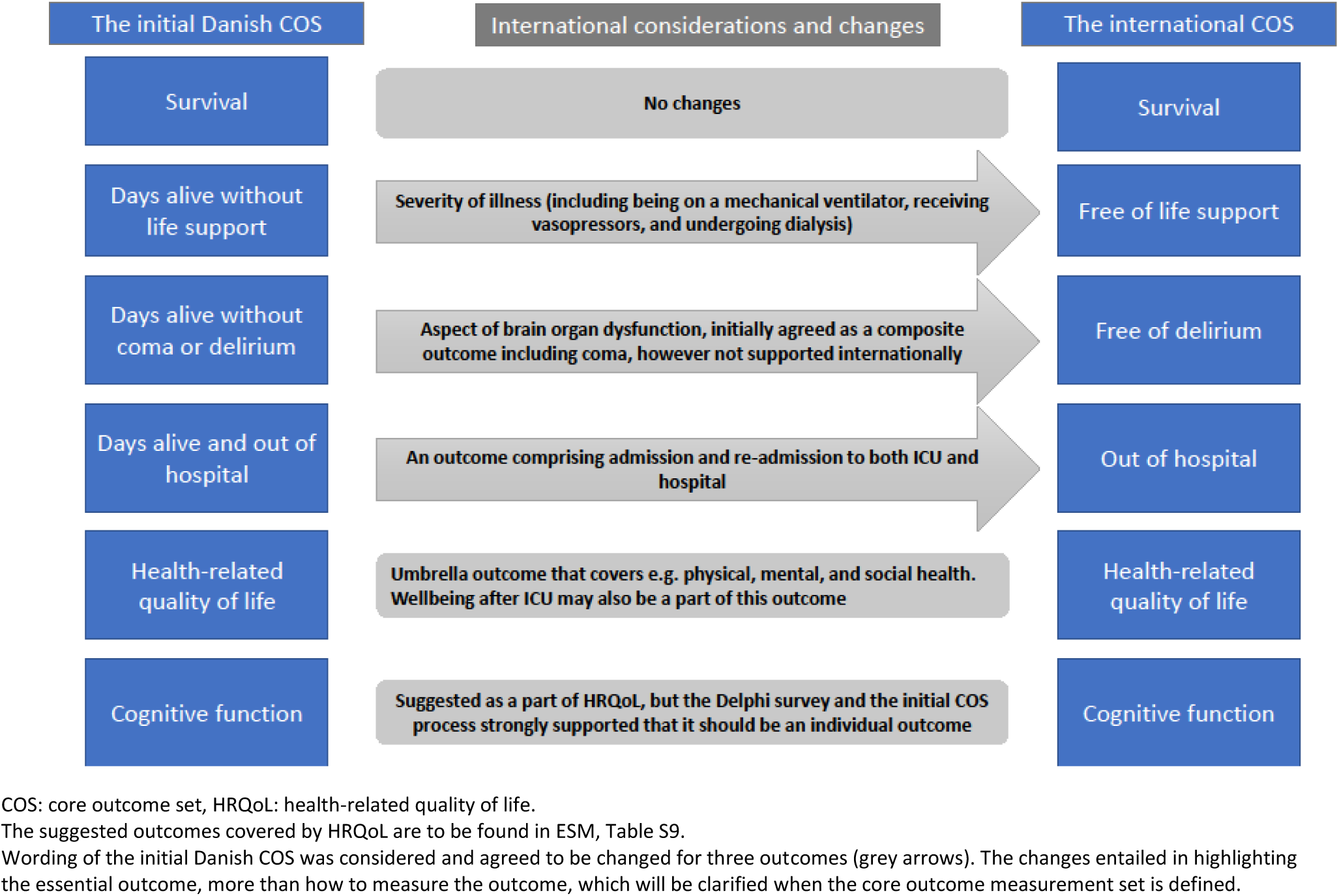
The initial Danish COS and the adjusted international COS.

### Step 5

Seventeen research panels in 13 countries (excluding Denmark) validated the initial COS (ESM, Table S1 and Table S10). Detailed minutes from all international research panels are provided in ESM, Table S5.

Three core outcomes were generally accepted: ‘survival’, ‘free of life support’, and ‘HRQoL’. During the international validation process, discussions were centred on the other initial core outcomes. It was considered for ‘days alive without coma or delirium’ that coma and delirium could not reasonably be amalgamated into a single outcome measure, and it was difficult to define coma and assess delirium, with delirium being a fluctuating mental state. ‘Days alive out of hospital’, and ‘cognitive function’ were rejected by one panel (5%), while three panels (14%) suggested that both outcomes should be adapted. ‘Days alive out of hospital’ should incorporate returning to home or previous level of function and ‘cognitive function’ could be considered as a domain of the HRQoL outcome.

The initial COS was revised based on the minutes from the international consensus meeting. An updated COS was then presented for feedback from the Danish and international research panels (Table 2, Figure 3).

### Step 6

The final internationally validated COS included survival, free of life support, free of delirium, out of hospital, HRQoL, and cognitive function. The COS reached unanimous consensus on ‘survival’, ‘free of life support’, and ‘health-related quality of life’. The remaining three outcomes had at least 90% consensus. For ‘out of hospital’, two panels (10%) recommended adapting it to include ‘returning to pre-ICU condition’.

One panel (5%) rejected ‘free of delirium’ because of practicalities and tools around assessing delirium (ESM, Page 228). Regarding ‘cognitive function’, two panels (10%) recommended adaptation to include this measure in HRQoL (Table 2, Figure 3).

## Discussion

We employed a multiple methods study incorporating a 3-round modified Delphi survey and an international validation process. This resulted in consensus of 6 core outcomes for adult, general ICU patients being: ‘survival’, ‘free of life support’, ‘free of delirium’, ‘out of hospital’, ‘HRQoL’, and ‘cognitive function’.

The inclusion of ‘survival’, ‘HRQoL’, and ‘cognitive function’ align with the COS for acute respiratory failure developed by Needham and colleagues [23, 31, 32]. However, our COS diverges by not including ‘muscle and/or nerve function’ and ‘pulmonary function’, likely relevant for patients with acute respiratory failure [32]. Conversely, our COS includes ‘free of life support’, ‘free of delirium’, and ‘out of hospital’, which were not included by Needham et al. Other core outcomes, including pain, mental health, and physical function, were all discussed during our consensus meetings. Although, ultimately not included in our COS, these will be considered when developing the subsequent core outcome measurement set.

Delirium was a critical outcome during all three rounds of the Delphi survey involving various stakeholders. In the international validation, it was highlighted due to its frequent occurrence and its burdensome nature for ICU patients, family members, and clinicians. It was acknowledged that delirium is a fluctuating state that can be challenging to detect [33]. Rose and colleagues developed a COS specifically for research interventions that aim to treat or prevent delirium in critically ill patients, underscoring the importance of this outcome [34].

The usability of our COS is somewhat constrained until the next step, which involves exact definitions, including the selection of appropriate instruments for their measurement and the timing of their assessments [11, 24]. Our future development of a standardised core outcome measurement set will also adhere to the COMET Handbook and COnsensus-based Standards for the selection of health status Measurement Instruments (COSMIN) initiatives [11, 35] and will involve the established research panels.

### Strengths and limitations

The strengths of this study. First, a diverse stakeholder involvement, which includes patients, family, and clinicians and researchers. Second, we actively involved stakeholders throughout the entire process and ensured well-balanced consensus meetings using nominal group technique [29]. Third, from the outset, we proactively planned for stakeholder inclusion within the modified Delphi process [24]. Fourth, the semi-structured interviews represented an opportunity to potentially reveal outcomes that had not been identified in our literature search. Fifth, we adhered rigorously to the modified Delphi process as described in the COMET Handbook. We also published the protocol before study commencement, following relevant COS-STAR recommendations [11]. Sixth, the Delphi process exhibited a high response rate without the necessity for response imputation. However, distinguishing between COS and the core outcome measurement set (instruments) posed a challenge during the consensus discussions, as some core outcomes definitions are influenced by how they are assessed. Seventh, we opted to enhance the COS relevance by an international validation process, encouraged by commenting on the published protocol to incorporate a broader context [27, 36]. This additional step aimed to strengthen the applicability of the initial COS. Consensus discussions in native languages further improved the applicability and generalisability of the COS. Furthermore, we anticipate that the international research panels will continue contributing to research in intensive care, as proven successful in other medical specialties [37].

Limitations of this study. First, the modified Delphi consensus process was confined to Denmark. Second, the decision to involve international collaborators in the validation process was made *post hoc*, following the completion of the Danish COS as outlined in the protocol [24]. We subsequently, registered the amendment to the protocol before conducting the international validation [27]. This approach offered a practical way of involving multiple international sites in the consensus development of a COS even though the panels may not have been fully representative of their countries. Third, convincing colleagues in other countries to the use of the COS may be challenged based on what is considered important in clinical practice and practise variations. In the context of randomised trials, random allocation will reduce this challenge. Additionally, we collaborated with a diverse array of countries, each with its unique case mix and economic circumstances, enhancing the generalisability of our findings. Despite varied perspectives and robust discussions during the consensus process, the final COS emerged consistently across all countries, underscoring the rigor of its development.

### Conclusions

We have developed and internationally validated a COS for ICU patients in general with six core outcomes. These are ‘survival’, ‘free of life support’, ‘free of delirium’, ‘out of hospital’, ‘health-related quality of life’, and ‘cognitive function’, and we encourage its use in future research, specifically clinical trials in the ICU setting.

## Data Availability

All data produced in the present work are contained in the manuscript

## Acknowledgements

We thank all 22 research panels who took part in the development of this core outcome set for their time and efforts.

## From the Danish national research panels

From the **Capital Region of Denmark** we thank Rasmus Hansen, Michael Piil Petersen, and Kent Bering (patients, Tine Piil Petersen, and Maria Høpner (family members), and Nicolai Haase (medical doctor). From the **Region Zealand** we thank Jan Müller (patient), Listbeth Christiansen (ICU nurse), Lars Nebrich (intensivist), and Laura Gandrup Kølby (occupational therapist). From the **North Denmark Region** Hans Henrik Svane (patient), Majbritt Svane (family member), Ruth Bliksted (physiotherapist), Karin Rehnholt Pedersen (ICU nurse), Maria Anker Thyø (intensivist), Heidi Yde Herrig (occupational therapist), Stine, Rom Vestergaard (ICU nurse). From the **Region of Southern Denmark** Tatjana Pihlkjær (physiotherapist), Jette West Larsen (ICU nurse), and Ib Højer Witt (patient), and Peter Martin Hansen, Jens Michelsen (medical doctor), Christine Haugaaard (ICU nurse), Mads Bisgaard Borup (physiotherapist), and Mette Bang Jørgensen (patient), Morten Borup (medical doctor), Henning Nielsen (Patient), Anne-Marie Nielsen (Family member). Leme Lehmkuhl from the Department of Anaesthesiology and Intensive Care, OUH Svendborg for recruiting patients and relatives for the modified Delphi survey.

## We specifically thank all international research panels and collaborators

From the research panels in **Australia:** Henry Hamelink, Sharon Knapp, Matt Maiden, Pamela McArdle, Peter McArdle, Alea Mclean, Jing Ning, Anne Marie Palermo, Sue Pellicano, Abbey Spinelli, Matt Spinelli, Lisa Van Der Lee, and Jake Dye (sepsis survivors, Regional/Victoria), Anthony Delaney (intensivist, Metro/New South Wales), Kirsten Thompson (sepsis coordinator/policy, Regional/Northern Territory), Frances Bass (research nurse, Metro/New South Wales), Matthew Ames (sepsis survivor, Metro/Queensland), Diane Ames (family perspective, Metro/Queensland), Amy Freeman-Sanderson (ICU Speech Pathologist, Metro/New South Wales), Andrew Udy (intensivist, Metro/Victoria), Brett Abbenbroek (ICU nurse, Metro/Queensland), Alex Poole (ICU nurse/researcher, Metro/Queensland), Priya Nair (intensivist, Metro/New South Wales), Simon Finfer (intensivist, Metro/New South Wales).

From the research panel in **Czech Republic:** Stepan Marecek, Nikola Musilova, Romana Musilova, Daniel Nalos, Mrs Hana Modrova (patients, and family members). Katerina Rusinova and Martin Balik (medical doctors).

From the research panels in **Finland:** Hanna Lahti, Hemmo Lahti, Heidi Kiiltomäki, Simo Hentonen, Ville Jalkanen (Tampere) and Pirkka Pekkarinen, Marjaana Tiainen, Leena Saarinen, Merja Heitto, Jarmo Heitto (Helsinki).

From the research panel in **Iceland:** Jakob V Finnbogason (ICU patient), Sigríður G Árnadóttir (relative), Árni Már, Ólöf S Sigurðardottir, and Hrönn Birgisdottir (ICU nurses), Sigurbergur Karason (ICU physician).

**From the research panel in India (Mumbai):** JV Divatia (Intensivist) Srinivas Samavedam (Intensivist) Rajiv Shah (ICU patient), Poonam Kharde (family member), Anjana Rane {Nurses). Others do not wish to be named.

**From the research panel in India (Chennai):** Bhuvana Krishna (Intensivist), JV Peter (Intensivist), H. Aishwarya (Respiratory therapist), Ram Rajagopalan (Intensivist), Vinitha Vijay (Nurse), Shree Lekha Madhav (Family member), Manikandan Ganeshan (Patient), Sarasa Mani (Patient), and Nandkumar N (Family member).

From the research panel in **Italy:** Giuseppe Bianco, Mariano Aresu, Viktoriya Kravchuk, Salvatore Ruggeri, and Barbara Gramegna (Patients) and Elena Costantini, Luca Carenzo, Daniela Elli (Department. of Anaesthesiology and Intensive Care, Humanitas Research Hospital, Milan) and Massimo Antonelli (Department. of Anesthesiology and Intensive Care Medicine, Gemelli Hospital, Rome) and Giacomo Grasselli (Dipartimento de Anestesia, Milan and Department. of Pathophysiology and Transplantation, Policlinico Maggiore, Milan).

From the research panels in **Lithuania:** Mindaugas Serpytis, Ingrida Lisauskiene, Jokubas Stanaitis, Sarunas Judickas, Albinas Kalimavicius From the research panels in **the Netherlands:** Siep Bos (Patient representative), Otto Polling (Patient representative), and Elza van den Berg (Research nurse).

From the research panels in **Norway:** Tom Rosenvinge (Patient representative, Mogreina), Celine Semb Hagen Linde (Patient representative, Fornebu), and Roar Magne Norheim (Patient representative, Risør), and Kristina Struksnes Fjone (Physiotherapist/Researcher, Oslo), Reidar Kvåle (Clinician and Professor, Bergen), Pål Klepstad (Clinician and Professor, Trondheim).

From the research panels in **Poland** thanks to The Polish research panel and Ryszard Gawda (medical doctor).

From the research panels in **Sweden** thanks to The Swedish core outcome set panel members.

From the research panels in **Switzerland:** Marie-Madlen Jeitziner, Matthias Exl, Daniela Bertschi (Department. of Intensive Care, Inselspital, University Hospital Bern and University of Bern) Sabrina Grossenbach (Department. of Physiotherapy, Inselspital, University Hospital Bern and University of Bern).

From the research panels in **United Kingdom**: PPI representatives Sheila Brooks, Debra Williams, Steven- Lee Dyton-Thomas, David Wynn Morgan, Julian Williams (Cardiff, Wales) and Clinicians Dr Sarka Furmanova, Dr Julie Highfield, Gemma Jones, Rachael Parker, Dr Gareth Scholey, Paul Twose (University Hospital of Wales). PPI representatives Chantal Davies, Danny Castell, Simon Kennedy, Del London (London, UK) Ms Gill Radcliffe, Ms Jo Gilroy, Ms Fabiola DAmato, Dr Joel Meyer, Ms Rebeka Wright (Guy’s and St Thomas’ Hospital, London). The validation process was supported by the Intensive Care Society UK.

## Author contributions

Conceptualisation: MNK, AG, MHM, AP, and MOC.

Data curation: MNK, CRLB, AG, MHM, BSR, CBM, TS, EL, ACB, TH, AMGB, LSH, AH, PS, SE, MC, AS, JHL, KH, FB, CP, MO, JMC, MPW, WS, AW, TJ, MC, MIS, MN, JH, MM, NH, EL, KH, SNM, BKTV, VJ, and MOC.

Formal analysis: MNK, and AG.

Funding acquisition: AG, MHM, AP, and MOC.

Investigation: all authors.

Methodology: MNK, AG, MHM, and MOC.

Project administration: in Denmark: MNK, CLRB, and MOC; internationally: MNK and CLRB.

Resources: MNK, CRLB, AG, MHM, BSR, CBM, TS, EL, ACB, TH, AMGB, LSH, AH, MC, AS, JHL, KH, FB, CP, MO, JMC, MPW, WS, AW, TJ, MC, MIS, MN, JH, MM, NH, EL, KH, SNM, BKTV, KY, VJ, and MOC.

Supervision: AG, MHM, AP, and MOC.

Validation: MNK, CRLB, AG, MHM, BSR, CBM, TS, EL, ACB, TH, AMGB, LSH, AH, PS, SE, MC, AS, JHL, KH, FB, CP, MO, JMC, MPW, WS, AW, TJ, MC, MIS, MN, JH, MM, NH, EL, KH, SNM, BKTV, KY, VJ, and MOC.

Visualization: MNK, and AG. Writing – first draft: MNK.

Writing – reviewing & editing: all authors.

### Steering committee, authors

MNK, CRLB, AG, BSR, CBM, LMP, TS, EL, ACB, TH, AMGB, LSH, IE, and MOC.

### International CRIC partners, authors

MC, AS, JHL, KH, FB, CP, MO, JMC, MPW, WS, AW, TJ, MC, MIS, MN, JH, MM, NM, EL, KH, SNM, VH, BV, and BKTV.

### International steering committee, authors

MNK, CRLB, AG, BSR, CBM, LMP, TS, EL, ACB, TH, AMGB, LSH, IE, MOC, MC, AS, JHL, KH, FB, CP, MO, JMC, MPW, WS, AW, TJ, MC, MIS, MN, JH, MM, NM, EL, KH, SNM, VH, BV, KY, and BKTV.

