## Supplemental material for "A core outcome set for adult general ICU patients"

**Electronic Supplementary Material (ESM)**

### Contents

Additional methodological details ... 3

Stakeholder involvement ... 3

Developing the initial core outcome set ... 3

Consensus process ... 6

The additional international validation ... 7

Analysis and reporting ... 7

Ethics ... 8

Table S1. Distribution of panel members in categories and professions across countries for the research panels ... 9

Table S2. Characteristics of stakeholders registered for the first round of the Delphi survey (n=264) ... 10

Table S3. Characteristics of stakeholders participating in the semi-structured interviews (n=82) ... 12

Search strategy for the initial list of outcomes ... 14

Table S4. Raw list of outcomes obtained from the search string ... 15

Rationale behind the initial core outcome set ... 23

Table S5. Summary minutes from all participating countries holding an international validation meeting ... 33

Table S6. Distribution of stakeholders registered for the Delphi survey ... 67

Table S7. Outcomes derived from the semi-structured interviews ... 68

Table S8. The 50 outcomes in the survey including help-text ... 69

Figure S1-S50. Graphs for the first survey round ... 74

Figure S51-S100. Graphs for the second survey round ... 124

Figure S101-150. Graphs for the third survey round ... 174

Figure S151. Heatmap showing the three Delphi survey round responses for all stakeholders ... 224

Figure S152. Heatmap showing the three Delphi survey round responses by stakeholder groups ... 225

Table S9. Outcomes related to health-related quality of life ... 226

Table S10. Results from the international validation meetings across the participating countries ... 227

Comments to the adjusted core outcomes ... 228

Table S11. Core Outcome Set-STandards for Reporting: The COS-STAR Statement Checklist ... 230

Table S12 Guidance for Reporting Involvement of Patients and the Public short form: The GRIPP2-SF checklist ... 232

References ... 234

### Additional methodological details

#### Stakeholder involvement

The steering committee (composed of authors MNK, CRLB, AG, BSR, CBM, LMP, TS, EL, ACB, TH, AMGB, LSH, IE, and MOC) for the initial core outcome set (COS) covers multi professional clinicians and researchers with knowledge of the intensive care unit (ICU) environment and critically ill patients. The steering committee established initially 5 research panels in 4 out of 5 regions in Denmark [1, 2], consisting of 10 patients, 5 family members, 19 multi professional clinicians, and 9 researchers in total. The research panels functioned as advisory boards in relation to the process with developing the COS. Thus, the research panels were involved in several research stages with a nominal group technique: the design, recruitment strategy, consensus, and dissemination in a broad sense from, e.g., information materials, interview guides, and wording in the survey [2].

#### Developing the initial core outcome set

##### *Identification of outcomes*

First step was a literature review obtaining outcomes from COS developed to the critically ill patient (Figure 1 in the manuscript, Step 1). This step was conducted to obtain initial knowledge of outcomes relevant for the critically ill population. The search string and all listed outcomes are to be found in the end of the electronic supplementary material (ESM) (Figure 1).

##### *Interviews*

Interviews were conducted concurrently with the design of the survey and the first survey round. Semi-structured individual interviews were carried out with patients, family members, clinicians, and researchers to identify any outcomes overlooked in the literature search [1]. Quota sampling was employed to ensure a balanced distribution of characteristics and especially for patients we ensured distribution of patients with predicted mortality above and below 25%, and furthermore we used a matrix with variables important to consider, with the goal of achieving a diverse distribution of [1, 3]:

- Age (young: 18 to 50 years old, middle: 51 to 75 years old, and old: >75 years old)
- Time after ICU discharge (1-2 weeks, 6 weeks or in a follow up clinic, or 1 year)
- Sex (female, male)
- Ethnicities (other than Danish origins covers ‘immigrants’ born abroad and neither of the parents is a Danish citizen or born in Denmark and ‘descendants’ born in Denmark and neither of the parents is a Danish citizen or born in Denmark

- Admission type (surgical or medical)
- SMS-ICU (low <17 point or high mortality risk ≥17). The Simplified Mortality Score for the Intensive Care Unit (SMS-ICU) is a severity score that predicts 90-day mortality with higher points meaning higher risk of mortality [4]. The range of points is 0-42. SMS-ICU assesses the severity of illness, which could influence the perspective of the importance of outcomes for a COS. Therefore, we aimed for a balanced distribution of patients with higher mortality risk ≥17 (predicted 90-day mortality risk of 25.3% and above) vs. lower risk <17 (predicted 90-day mortality risk of 22.8% and below) [4]
- ICU length of stay (<7 days or ≥7 days)

Interviews were conducted either by phone or through physical attendance, based on the preferences of the informants. Three interview guides were developed to encompass all stakeholders invited for interviews: one for patients, one for family members, and one for clinicians and researchers [1]. These guides underwent cognitive validation by Danish research panels, focusing on the structure, the directness of questions considering potential emotional stress, and whether they addressed the intended research questions.

Although all interviews were reviewed for outcomes not covered in our literature search, the framework analysis of the interviews will not be included in this study. The interviews were carried out by MNK, CRLB, EL, SRV, CBM, AH, MOC, and three former members of the steering committee using validated interview guides published in the protocol [1].

##### *Construction of survey*

The first step involved a literature review, utilising a search string (in this ESM) from a previous systematic review regarding COS for critical illness and recovery (Figure 1, Step 1) [1, 5]. From this search, we obtained a raw list of outcomes considered for developing a COS. We independently and in duplicate (MNK and SE) obtained 329 outcomes listed alphabetically in the protocol and in page 9 in this ESM [1]. MNK, CBM, PS, SE, LMP, EL, and MOC removed duplicates and used thematic analysis to condense the outcomes in categories. Then MNK, CRLB, PS, and MOC translated all outcomes to Danish and made mind maps with a central category in the middle and associated outcomes around it. Some categories contained overlapping outcomes to break up all outcomes into smaller pieces to be presented for the Danish research panels.

The second step was to present all the categories for the Danish research panels. We used the nominal group technique, with face-to-face discussion in small groups aiming for an instant result [6], to discuss, reallocate and further condense to essential outcomes as many were overlapping ending up with 50 outcomes (Figure 1, Step 2).

The third step was determining how to formulate survey questions that covered the essential outcomes and to construct the survey in DelphiManager (COMET, University of Liverpool, Liverpool) using their default layouts and graphics.

The fourth step was for the research panels to pilot test the survey to ensure intelligibility and face validity, aiming to prevent attrition resulting from a lack of understanding. This was achieved through the use of the think aloud technique, testing the DelphiManager software [7].

##### *Recruitment for the survey*

We aimed to recruit 400 participants for the survey, 100 patients, 100 family members, and 200 clinicians and researchers [1]. We used convenience sampling recruiting both patients and family members via long-term follow-up of trial participants, outpatient clinics, the general ward, ICU patient cafés, and hand out invitations (flyers). Clinicians and researchers were identified in the authors affiliations by the research network, Collaboration of Research in Intensive Care (CRIC), and comprised multi professional participants. Informants were also invited for the survey.

We aimed for a comprehensive representation of patients in the survey as well following the same matrix as for the interview covering the following variables obtained and shown in the table illustrating characteristics of the patients in the Delphi survey [1]:

- Age (young: 18 to 50 years old, middle-aged: 51 to 75 years old, and old: >75 years old)
- Time after ICU discharge (1-2 weeks, 6 weeks or in a follow up clinic, or 1 year)
- Sex (female, male)
- Ethnicities as described above
- Admission type (surgical or medical)
- SMS-ICU as described above
- ICU length of stay (<7 days or ≥7 days)

##### *Survey round 1*

In round 1, we recorded demographic data according to the protocol [1], and assigned all consenting registrations a unique identifier.

After the first survey round, the research panels held a consensus meeting where the results from the first round was summarised and the survey participants suggested additional outcomes, and participants' comments were reviewed. There was unanimous agreement not to add any additional outcomes, but the comments added by survey participants required revision of the questions and revision of parts of the help text provided. Also, there was consensus not to remove any outcomes from the survey, as outlined in the 'consensus process' section.

##### *Additional two survey rounds*

We performed two additional rounds as the survey was revised. For precision, we included the wording 'assessing' or 'investigating' as an introduction of 31 outcomes. Additionally, for six questions related to health economics, well-being, blood samples, delirium, thirst, and dyspnoeic conditions, we included examples or provided more details to clarify the inquiries. Furthermore, we expanded the help text in six outcomes. Predefined, only those registered for the first survey round were able to participate in the following two round(s) [1]. No changes were made between round 2 and 3. One national research panel were convened between the rounds 2 and 3 to review the scores of the outcomes, determining whether there was consensus to remove any from the survey.

##### *Retention strategy*

We supplied all invited participants with an e-mail address and a hot line for any questions related to the survey. Each round defaulted to a three-week duration, with weekly notifications sent out to remind participants to complete the survey. The final round was extended to four weeks when feasible. Patients and family members received phone calls to inquire if they required assistance in using the DelphiManager software. Additional information on the retention strategy can be found in the protocol [1].

#### Consensus process

After the completion of the survey rounds, four research panels individually conducted a consensus meeting to review the findings and reach consensus on their final proposal for a COS. MNK and CRLB visited all four panels to ensure a consistent consensus process starting with a summarisation of the results (MNK) and maintained consistent minutes (CRLB). The research panels were tasked with discussing the survey results and determining which outcomes should be considered core outcomes, thereby defining their perspective on a COS. The research panels identified between 9 and 13 outcomes, with some overlap, ultimately resulting in a total of 19 outcomes (Figure 1).

The summaries from all 4 meetings, the survey results, and the 19 outcomes were presented to the steering committee informing their consensus meeting across Denmark, with the goal of reaching consensus on an initial COS with an accessible number of outcomes [7]. The detailed description of the meeting and the rationale behind the revealed 6 outcomes are to be found in this ESM page 23.

#### The additional international validation

The countries within the Collaboration of Research in Intensive Care (CRIC) network were informed about the pragmatic international validation of the initial COS and agreed to participate in this study. The participating countries included Australia, the Czech Republic, Finland, Iceland, India, Italy, Lithuania, New Zealand, Norway, Poland, Sweden, Switzerland, the Netherlands, the United Kingdom, and the United States of America.

International validation necessitated at least one research panel from each participating country. Using the nominal group technique, these panels deliberated on the initial COS and its applicability in their national context. The protocol amendment for this process was written and registered before the first international validation meeting was convened [8]. During the validation meetings, each country provided insights on whether they could adopt the outcome(s) as they were, whether adaptions were needed for their country, or whether they rejected the outcome(s). Meeting facilitators were briefed on the process behind the initial COS, the rationale for it, and were equipped with a slideshow to ensure a consistent presentation of the meeting's purpose. All necessary information was provided to guide the facilitators in discussions involving new outcomes that may have suggested or previously discussed. Detailed minutes were documented to ensure a transparent process and discussion.

Following completion of all international validation meetings, all authors gathered online for a consensus meeting. They deliberated and tried to reach an agreement on whether the initial COS could be universally applicable for all participating countries, whether adaption was necessary, or if new outcomes should be added, akin to the extended version as demonstrated by Fink et al. [9].

#### Analysis and reporting

Characteristics were recorded from informants who consented to be interviewed and from survey participants who consented for registration in the DelphiManager software for the first survey round. Few characteristics were acquired from patients' medical journals following consent (for further information see the protocol) [1].

##### *Attrition between rounds*

We had more than 5% attrition, which prompted the utilisation of the *last observation carried forward* approach for rounds 2 and 3 in cases where patients did not respond to previously rated items. The attrition was 19% between round 1 and 2, as well as round 1 and 3 as attrition was calculated from those completing the first round. Consequently, we present the complete cases with last observation carried forward as planned [1].

##### *Statistics*

We used descriptive statistics to report the populations and responses of the survey rounds, presenting numbers and percentages of respondents. Regarding the characteristics of the participants, categorical data were expressed using numbers and percentages, while the numeric data were represented by medians with interquartile ranges (IQRs). Missing data were handled with last observation carried forward and with percentages for missingness presented as previously described [1].

#### Ethics

Participants who underwent interviews provided written informed consent, while survey participants indicated their consent by ticking the 'confirming consent' box in the survey. They were informed of the voluntary nature of participation before any additional responses were collected. Data were handled confidentially and anonymously without using their names, and participants had the option to withdraw their consent at any time. We sought approval for all participating hospitals and the Danish Data Protection Agency to ensure compliance with the General Data Protection Regulation (GDPR). The Ethical Committee for the Capital Region of Denmark waived the need for ethical approval (H-21010116). For the international validation, Iceland was obliged to obtain the Ethics Board Permission with informed consent from participants, and the United Kingdom (London) obtained institutional approval from the Guy's & St Thomas' Hospital London.

Subject confidentiality was rigorously upheld by participating investigators, research staff, and the sponsoring institution. No information concerning the study, or the data will be released to any unauthorized third party without prior written approval from the steering committee.

In the case of the DelphiManager software, anonymity for all participants was ensured through individual unique identifiers, and data were stored in a secure location.

#### Table S1. Distribution of panel members in categories and professions across countries for the research panels

| Country | Number of panels[a] | Patients | Family members | Doctors | Nurses | Other[f] | Researchers | Total |
|---|---|---|---|---|---|---|---|---|
| **Australia** | 2 | 5 | 5 | (5) | 3 (5) | (2) | 11 | 24 |
| **Czech Republic** | 1 | 2 | 2 | 2 | 0 | 0 | 2 | 8 |
| **Denmark** | 4 | 8 | 5 | 6 (4) | 6 (5) | 5 | 9 | 39 |
| **Finland[c]** | 2 | 7 | 3 | 3(1) | 0 | 0 | 6 | 19 |
| **Iceland** | 1 | 1 | 1 | (2) | 2 (1) | 0 | 3 | 7 |
| **India[d]** | 2 | 4 | 4 | 10 (1) | 2 | 2 | 2 | 24 |
| **Italy** | 1 | 3 | 2 | 2(3) | 1 | 1 | 3 | 12 |
| **Lithuania** | 1 | 1 | 1 | 3 | 2 | 0 | 3 | 10 |
| **Norway** | 1 | 2 | 0 | 1 (3) | (1) | (1) | 6 | 9 |
| **Netherlands** | 1 | 4 | 2 | 2 | 2 | 0 | 2 | 12 |
| **Poland** | 1 | 1 | 1 | 2 | 1 | 0 | 2 | 7 |
| **Sweden** | 1 | 5 | 1 | 2 | 2 | 2 | 0 | 12 |
| **Switzerland** | 1 | 2 | 2 | 1 (1) | (2) | (1) | 4 | 9 |
| **United Kingdom[e]** | 2 | 9 | 2 | 3 (1) | 3 (1) | 6 | 2 | 25 |
| **Total** | 21 | 54 | 31 | 37 | 24 | 16 | 55 | 217 |

All data entered are numeric. No missing data. Brackets refer to the proportion of researchers.
This table contains all the countries who have held a national or international validation meeting with a local research panel(s) contributing to the consensus of the core outcome set.
[a] Number of research panels established in the single country.
[b] Australian areas represented: Aboriginals, Western Australia, New South Wales, Victoria, Queensland, and Northern Territory. One panel was focused on the general intensive care unit patient and the other panel on sepsis survivors.
[c] One panel from Tampere and one panel from Helsinki.
[d] Indian areas represented: Chennai, Bangalore, Vellore, Pune, New Delhi, and Mumbai.
[e] One panel from Wales and one from England.
[f] Other consisted of multi professional stakeholders as e.g., physiotherapists, occupational therapists.

#### Table S2. Characteristics of stakeholders registered for the first round of the Delphi survey (n=264)

| | Patients | Family members | Clinicians | Researchers[g] |
|---|---|---|---|---|
| | n=65 | n=49 | n=136 | n=14 |
| **Characteristics** | | | | |
| **Age (years)** | 62 (50 to 70) | 59 (47 to 65) | 46 (39 to 52) | 41 (37 to 48) |
| **Sex (female)** | 21 (32%) | 34 (69%) | 93 (68%) | 9 (64%) |
| **Region of residence:** | | | | |
| **-The Capital Region of Denmark** | 19 (29%) | 13 (27%) | 30 (22%) | 6 (43%) |
| **-Region Zealand** | 6 (9%) | 6 (12%) | 22 (16%) | 2 (14%) |
| **-Central Denmark Region** | 12 (19%) | 9 (18%) | 19 (14%) | 2 (14%) |
| **-The North Denmark Region** | 14 (22%) | 10 (20%) | 35 (26%) | 4 (29%) |
| **-The Region of Southern Denmark** | 14 (22%) | 11 (22%) | 30 (22%) | 0 (0%) |
| **Other ethnicity than Danish[a]** | 4 (6%) | 1 (2%) | 5 (4%) | 3 (21%) |
| **Admission type** | | | | |
| - **Surgical** | 29 (47%)[f] | - | - | - |
| - **Medical** | 33 (53%)[f] | - | - | - |
| **SMS-ICU[b] at ICU admission[c]** | 20 (13.5 to 22.0)[f] | - | - | - |
| - **<17 points** | 23 (37%)[f] | - | - | - |
| - **≥17 points** | 40 (63%)[f] | - | - | - |
| **Self-reported critical illness severity[d]** | 10.0 (8.5 to 10.0) | - | - | - |
| **ICU length of stay (days)[c]** | 9.0 (6.0 to 22.5)[f] | - | - | - |
| **Time since ICU admission (months)[c]** | 14 (3 to 20)[f] | - | - | - |
| **Assistance to fill in the survey** | 11 (17%) | - | - | - |
| **Family to a deceased patient** | - | 5 (10%) | - | - |
| **Clinical experience (years)** | - | - | 18 (11 to 24) | 12 (6 to 20) |
| **Employed in ICU (experience in years)** | - | - | 10 (3 to 15) | 5 (1 to 10) |
| **Professional group** | | | | |
| - **Medical doctors** | - | - | 64 (47%) | 8 (57%) |
| - **Nurses** | - | - | 52 (38%) | 5 (36%) |
| - **Physiotherapist** | - | - | 11 (8%) | 1 (7%) |
| - **Occupational therapist** | - | - | 8 (6%) | 0 (0%) |
| - **Other** | - | - | 1 (1%) | 0 (0%) |
| **Research experience (years)** | - | - | 0 (0 to 5) | 7 (5 to 8) |
| - **Medical doctors** | - | - | 4 (0 to 10) | 5.5 (4.5 to 7.3)[h] |
| - **Nurses** | - | - | None | 8 (5 to 8)[i] |
| - **Physiotherapist** | - | - | 0 (0 to 1) | 10 (10 to 10)[j] |
| - **Occupational therapist** | - | - | None | - |
| - **Other[e]** | - | - | - | - |
| **Time doing research[f]** | | | | |
| - **Full time** | - | - | - | 9 (64%) |
| - **Part time** | - | - | 12 (9%) | 4 (29%) |
| - **Free time** | - | - | 49 (37%) | 1 (7%) |
| - **No research** | - | - | 71 (54%) | - |
| **Highest obtained academic degree** | | | | |
| - **Not bachelor** | - | - | 11 (8%) | - |
| - **Bachelor** | - | - | 48 (35%) | - |
| - **Master** | - | - | 50 (37%) | 7 (50%) |
| - **PhD or higher degree** | - | - | 27 (20%) | 7 (50%) |
| **Employment** | | | | |
| - **University hospital** | - | - | 118 (87%) | 13 (93%) |
| - **Non-university hospital** | - | - | 15 (11%) | 1 (7%) |
| - **Other** | - | - | 3 (2%) | - |

SMS-ICU: The Simplified Mortality Score for the Intensive Care Unit[4], ICU: intensive care unit.
Numeric data are presented as medians with interquartile ranges, categorical data as counts with percentages.
[a] Danish origin has at least one parent born in Denmark with Danish citizenship (Statistics Denmark, www.dst.dk). Other than Danish origins covers both 'immigrants' born abroad and neither of the parents is a Danish citizen or born in Denmark and 'descendants' born in Denmark and neither of the parents is a Danish citizen or born in Denmark (Statistics Denmark, www.dst.dk).
[b] Patients with higher mortality risk ≥17 (predicted 90-day mortality risk of 25.3% and above) vs. lower risk <17 (predicted 90-day mortality risk of 22.8% and below) [4].
[c] These variables were obtained from the patients' medical records. For the survey there were two patients with missing medical record data. One withdrew consent, one where there were only paper medical records, which couldn't be located.
[d] Self-reported severity of illness assessed on a numeric score (0-10 where 0 is non-critical and 10 is most critical). From either the patients' view or from the view of family members to ICU patients.
[e] No other professions with research experience than listed.
[f] There were 4 clinicians with missing data for the question concerning time doing research.
[g] Researchers covers clinician-researchers and researchers from other professional groups
[h] There were two missing among doctors.
[i] There were three missing among nurses.
[j] There were two missing among physiotherapists.

#### Table S3. Characteristics of stakeholders participating in the semi-structured interviews (n=82)

| | Patients (n=27) | Family members (n=25) | Clinicians (n=26) | Researchers(n=4)[h] |
|---|---|---|---|---|
| **Characteristics** | | | | |
| **Age (years)** | 60 (50 to 68) | 54 (41 to 60) | 45 (43 to 48) | 43 (38 to 48) |
| **Sex (female)** | 13 (48%) | 18 (72%) | 21 (81%) | 4 (100%) |
| **Region of residence:** | | | | |
| **- The Capital Region of Denmark** | 6 (22%) | 6 (24%) | 5 (19%)[j] | 2 (50%) |
| **-Region Zealand** | 3 (11%) | 3 (12%) | 4 (15%)[j] | 0 (0%) |
| **-Central Denmark Region** | 6 (22%) | 6 (24%) | 4 (15%)[j] | 1 (25%) |
| **-The North Denmark Region** | 7 (26%) | 6 (24%) | 6 (23%)[j] | 1 (25%) |
| **-The Region of Southern Denmark** | 5 (19%) | 4 (16%) | 6 (23%)[j] | 0 (0%) |
| **Other ethnicity than Danish[a,b]** | 1 (4%) | 1 (4%) | 3 (12%) | 0 (0%) |
| **Admission type** | | | | |
| - **Surgical** | 13 (48%) | - | - | - |
| - **Medical** | 14 (52%) | - | - | - |
| **SMS-ICU[c] at ICU admission[c]** | 19 (15 to 22) | - | - | - |
| - **<17 points** | 10 (37%) | - | - | - |
| - **≥17 points** | 17 (63%) | - | - | - |
| **Self-reported critical illness severity[e]** | 9 (8 to 10) | - | - | - |
| **ICU length of stay (days)[d]** | 9 (7 to 25) | - | - | - |
| **Time since ICU admission (months)[d]** | 11 (4 to 17) | - | - | - |
| **Family to a deceased patient** | - | 3 (12%) | - | - |
| **Clinical experience (years)** | - | - | 19 (14 to 23) | 13 (5 to 22) |
| **Employed in ICU (experience in years)** | - | - | 12 (5 to 16) | 7 (3 to 11) |
| **Professional group** | | | | |
| - **Medical doctors** | - | - | 12 (46%) | 1 (25%) |
| - **Nurses** | - | - | 9 (35%) | 4 (75%) |
| - **Physiotherapist** | - | - | 3 (12%) | 0 (0%) |
| - **Occupational therapist** | - | - | 2 (7%) | 0 (0%) |
| - **Other[f]** | - | - | 0 (0%) | 0 (0%) |
| **Research experience (years)** | - | - | 0 (0 to 6)[g] | 8 (3 to 12) |
| - **Medical doctors** | - | - | 6 (0 to 10)[g] | 4 (4 to 4) |
| - **Nurses** | - | - | 0 (0 to 0)[g] | 10 (7 to 13) |
| - **Physiotherapist** | - | - | 0 (0 to 7)[g] | 0 (0 to 0) |
| - **Occupational therapist** | - | - | 0 (0 to 0)[g] | 0 (0 to 0) |
| - **Other[f]** | - | - | 0 (0 to 0)[g] | 0 (0 to 0) |
| **Time doing research** | | | | |
| - **Full time** | - | - | 0 (0%)[g] | 4 (100%) |
| - **Part time** | - | - | 3 (81%)[g] | 0 (0%) |
| - **Free time** | - | - | 6 (23%)[g] | 0 (0%) |
| - **None** | - | - | 16 (62%)[g] | |
| **Highest obtained academic degree** | | | | |
| - **Bachelor** | - | - | 5 (19%) | 0 (0%) |
| - **Master** | - | - | 14 (54%) | 3 (75%) |
| - **PhD or higher** | - | - | 7 (27%) | 1 (25%) |
| - | | | | |
| **Employment** | | | | |
| - **University hospital** | - | - | 26 (100%) | 4 (100%) |
| - **Non-university hospital** | - | - | 0 (0%) | 0 (0%) |
| - **University** | - | - | 0 (0%) | 0 (0%) |
| - **Other** | - | - | 0 (0%) | 0 (0%) |

SMS-ICU: The Simplified Mortality Score for the Intensive Care Unit[4], ICU: intensive care unit.

Numeric data are presented as medians with interquartile ranges, categorical data as counts with percentages.
[a] Danish origin has at least one parent born in Denmark with Danish citizenship (Statistics Denmark, www.dst.dk). Of missing data were 2 patients, 2 family member, and 1 clinician. Other than Danish origins covers 'immigrants' born abroad and neither of the parents is a Danish citizen or born in Denmark and 'descendants' born in Denmark and neither of the parents is a Danish citizen or born in Denmark (Statistics Denmark, www.dst.dk).
[b] Missing data for 2 patients, 1 family member, and 1 clinician.
[c] Patients with higher mortality risk ≥17 (predicted 90-day mortality risk of 25.3% and above) vs. lower risk <17 (predicted 90-day mortality risk of 22.8% and below) [4].
[d] These variables were obtained from the patients' medical records. No missing data.
[e] Self-reported severity of illness assessed on a numeric score (0-10 where 0 is non-critical and 10 is most critical). From either the patients' view or from the view of family members to ICU patients.
[f] No other professions than already listed.
[g] There were 1 clinician with missing data for the question concerning region of residence, clinical experience, and time doing research.
[h] Researchers covers clinician-researchers and researchers from other professional groups

#### Search strategy for the initial list of outcomes

Search strategies for publications on critical care-related core outcome sets from the systematic review conducted by Dinglas et al.[5] The search was conducted 1 May 2021.

Search strategy for PubMed:

("acute care"[tw] OR "asystole"[tw] OR "brain injuries, traumatic"[mesh] OR "burn unit"[tw] OR "burn units"[mesh] OR "burn units"[tw] OR "cardiac arrest"[tw] OR "cardiac arrests"[tw] OR "cardiac death"[tw] OR "cardiac surgeries"[tw] OR "cardiac surgical procedures"[mesh] OR "cardiac surgery"[tw] OR "cardiopulmonary arrest"[tw] OR "cardiopulmonary arrests"[tw] OR "cardiopulmonary death"[tw] OR "coma"[mesh] OR "coma"[tw] OR "comas"[tw] OR "comatose"[tw] OR "coronary care unit"[tw] OR "coronary care units"[mesh] OR "coronary care units"[tw] OR "critical care"[mesh] OR "critical care"[tw] OR "H D U "[tw] OR "HDU"[tw] OR "HDUs"[tw] OR "heart arrest"[mesh] OR "heart arrest"[tw] OR "heart arrests"[tw] OR "heart death"[tw] OR "heart surgeries"[tw] OR "heart surgery"[tw] OR "high dependency unit"[tw] OR "high dependency units"[tw] OR "I C U "[tw] OR "I T U"[tw] OR "ICU"[tw] OR "ICUs"[tw] OR "intensive care units"[mesh] OR "intensive care units, neonatal"[mesh] OR "intensive care units, pediatric"[mesh] OR "intensive care"[tw] OR "intensive therapy unit"[tw] OR "intensive therapy units"[tw] OR "ITU"[tw] OR "ITUs"[tw] OR "mechanical ventilation"[tw] OR "PICU"[tw] OR "PICUs"[tw] OR "P I C U "[tw]OR "recovery room"[mesh] OR "respiratory care unit"[tw] OR "respiratory care units"[mesh] OR "respiratory care units"[tw] OR "sepsis"[mesh] OR "sepsis"[tw] OR "septic"[tw] OR "TBI"[tw] OR "TBIs"[tw] OR "traumatic brain injuries"[tw] OR "traumatic brain injury"[tw] OR "ventilated"[tw] OR "ventilator"[tw] OR "ventilators"[tw]) AND ("critical care outcomes"[mesh] OR "critical care outcome"[tw] OR "critical care outcomes"[tw] OR "core outcome"[tw] OR "core outcomes"[tw] OR "core set"[tw] OR "core sets"[tw] OR "core measure"[tw] OR "core measures"[tw] OR "core domain"[tw] OR "core domains"[tw])

Search strategy for Embase:

('acute care':ti,ab,kw OR 'asystole':ti,ab,kw OR 'traumatic brain injury'/exp OR 'burn unit':ti,ab,kw OR 'burn unit'/exp OR 'burn units':ti,ab,kw OR 'cardiac arrest':ti,ab,kw OR 'cardiac arrests':ti,ab,kw OR 'cardiac death':ti,ab,kw OR 'cardiac surgeries':ti,ab,kw OR 'heart surgery':ti,ab,kw OR 'cardiac surgery':ti,ab,kw OR 'cardiopulmonary arrest':ti,ab,kw OR 'cardiopulmonary arrests':ti,ab,kw OR 'cardiopulmonary death':ti,ab,kw OR 'coma'/exp OR 'coma':ti,ab,kw OR 'comas':ti,ab,kw OR 'comatose':ti,ab,kw OR 'coronary care unit':ti,ab,kw OR 'coronary care unit'/exp OR 'coronary care units':ti,ab,kw OR 'critical care':ti,ab,kw OR 'H D U ':ti,ab,kw OR 'HDU':ti,ab,kw OR 'heart arrest'/exp OR 'heart arrest':ti,ab,kw OR 'heart arrests':ti,ab,kw OR 'heart death':ti,ab,kw OR 'heart surgeries':ti,ab,kw OR 'heart surgery':ti,ab,kw OR 'high dependency unit':ti,ab,kw OR 'high dependency units':ti,ab,kw OR 'I C U ':ti,ab,kw OR 'I T U':ti,ab,kw OR 'ICU':ti,ab,kw OR 'ICUs':ti,ab,kw OR 'intensive care'/exp OR 'intensive care unit'/exp OR 'neonatal intensive care unit'/exp OR 'pediatric intensive care unit'/exp OR 'intensive care':ti,ab,kw OR 'intensive therapy unit':ti,ab,kw OR 'intensive therapy units':ti,ab,kw OR 'ITU':ti,ab,kw OR 'ITUs':ti,ab,kw OR 'mechanical ventilation':ti,ab,kw OR 'PICU':ti,ab,kw OR 'PICUs':ti,ab,kw OR 'P I C U ':ti,ab,kw OR 'recovery room'/exp OR 'respiratory care unit':ti,ab,kw OR 'respiratory care units':ti,ab,kw OR 'sepsis'/exp OR 'sepsis':ti,ab,kw OR 'septic':ti,ab,kw OR 'TBI':ti,ab,kw OR 'TBIs':ti,ab,kw OR 'traumatic brain injuries':ti,ab,kw OR 'traumatic brain injury':ti,ab,kw OR 'ventilated':ti,ab,kw OR 'ventilator':ti,ab,kw OR 'ventilators':ti,ab,kw) AND ('critical care outcome'/exp OR 'critical care outcome':ti,ab,kw OR 'critical care outcomes':ti,ab,kw OR 'core outcome':ti,ab,kw OR 'core outcomes':ti,ab,kw OR 'core set':ti,ab,kw OR 'core sets':ti,ab,kw OR 'core measure':ti,ab,kw OR 'core measures':ti,ab,kw OR 'core domain':ti,ab,kw OR 'core domains':ti,ab,kw)

#### Table S4. Raw list of outcomes obtained from the search string

- Abnormal time of temperature during infection
- Accidental extubation
- Acute physiology and chronic health evaluation (APACHE II) score
- Acute Respiratory Distress Syndrome (ARDS)
- Additional medication for agitation
- Adverse events
- Adverse events
- Alanine aminotransferase (ALT)
- All-cause mortality
- Anaemia
- Antibiotic therapy
- Antipyretic rate
- Anxiety
- Any return of spontaneous circulation (ROSC)
- Acute physiology and chronic health evaluation (APACHE IV) scorre
- Appetite
- At hospital discharge is independence in functional level established matching home environment
- Biochemical outcomes
- Blood gas analysis
- Blood oxygen saturation
- Blood pressure
- Blood routine test
- Body composition
- Body composition tests (ultrasound/anthropometry)
- Boredom
- Brain function
- Calcitonin
- Cardiovascular disease
- Catecholamine infusion
- Change in activities of daily living
- Change in downstream marker (e.g. interleukin-6),
- Changes in relationships
- Chest computed tomography (CT)
- Chest pain
- Chest x-ray examination
- Chloroquine blood concentration
- Circulatory function
- Clearance time of fatigue
- Clinical workload
- Cluster of differentiation (CD)-4 and CD8 T-cells count
- Coagulation function
- Coagulation outcomes
- Cognition

- Cognition
- Cognitive function and symptoms
- Complete blood count (CBC)
- Confusion, uraemia, respiratory rate, blood pressure, age ≥65 years (CURB-65) pneumonia severity score
- Consciousness
- Constipation
- Contamination of packs and glass bottles
- Cost-effectiveness
- Costs (cost-effectiveness)
- Cough
- C-reactive protein (CRP)
- CT of hip
- Curative effects of traditional Chinese medicine (TCM) syndrome
- Days of immobility (bed-bound)
- Days of sedation
- D-dimer
- Decreased quality of life
- Decreased walking distance
- Delirium duration*
- Delirium incidence*
- Delirium prevalence*
- Delirium resolution*
- Delirium severity*
- Delirium type*
- Delirium*
- Delivery of care
- Demand for first aid measurement
- Diabetes
- Diarrhoea
- Difficulties concentrating
- Difficulties in activity of daily living (ADL)
- Difficulties returning to work/employment
- Disability
- Discharge location
- Disease severity
- Disseminated intravascular coagulation (DIC) score
- Dizziness
- Duration (d) of extracorporeal membrane oxygenation (ECMO)
- Duration (d) of oxygen inhalation
- Duration (d) of supplemental oxygenation
- Duration (d)/frequency of medical ventilation
- Duration of antibiotics treatment
- Duration of hospital stay
- Duration of ICU stay

- Duration of intubation
- Duration of weaning from mechanical ventilation?
- Dysphagia
- Electrocardiogram (ECG)
- Emotional well-being
- Endurance
- Erythrocyte sedimentation rate
- Exercise capacity and intensity
- Fatigue
- Feeding intolerance
- Feeling isolated
- Fever
- Financial impact on patient
- Financial impact on the individual
- Financial impact on the patient
- Finger oxygen improvement rate
- Flexion contractures and heterotopic ossification
- Frequency of clinical improvement (turning to ordinary or recovery)
- Frequency of hospital discharge
- Frequency of ICU admission
- Frequency of interstitial pneumonia
- Frequency of multi organ dysfunction syndrome (MODS)
- Frequency of other sequelae
- Frequency of shock
- Gas production
- Gastrointestinal discomfort
- Gastrointestinal function and symptoms
- Genetic factors/biomarkers
- Global Organ Dysfunction/Multi-organ failure
- Gratitude
- Headache
- Health care resource use
- Healthcare resource utilization (e.g., doctors' visits, need for further intervention)
- Health-related quality of life
- Health-related quality of life
- Hearing impairment
- Heart function
- Heart rate
- Hobbies
- Hospital/ICU-free survival
- Hospital-acquired infection
- Hospitalisation cost
- Hospitalisation/length of stay
- Human leukocyte antigen – DR isotype (HLA-DR)

- ICU free days
- Interleukin (Il-10)
- Interleukin (Il-4)
- Interleukin (Il-6)
- Interleukin (Il-8)
- Immunity and antibodies
- Immunoglobin
- Impact on family and/or caregivers
- Impact on family, caregivers or both
- Improvement of aerobic capacity
- Improvement of functional exercise capacity
- Improvement of physical function
- Improvement of respiratory muscle strength
- Improvement of skeletal muscle strength
- Incidence of antibiotic treatment
- Incidence of hypoxia
- Incidence of intensive care unit admission
- Incidence of medical complications during hospitalisation
- Incidence of shock
- Incidence of Ventilator associated pneumonia
- Incontinence
- Infections
- Intensive care unit acquired weakness
- Intensive care unit length of stay
- Interferon y
- Interleukin (Il)-2
- Intracranial haemorrhage
- Jaw thrust manoeuvre
- Joint pain
- Kidney function
- Lack of physical strength
- Length of hospital stay
- Length of hospital stay
- Less independence
- Level of virus antibodies in blood sample
- Life participation
- Liquid balance
- Liver function
- Loneliness/isolation
- Long-term mortality rate
- Loss of taste
- Lowest oxygen saturation (SpO2) during intubation
- Lung scarring (fibrosis)
- Lymphocyte count

- Lymphocyte count
- Major bleeding
- Major organ failure
- Management of complex medication regimens
- Mechanical ventilation
- Mechanical ventilation duration
- Memory loss
- Mental health/emotional functioning
- Metabolic complications
- Mobility
- Mortality
- Move to nursing home
- MRI of hip
- Muscle and/or nerve function
- Muscle function, nerve function or both
- Muscle pain
- Myocardial enzymes
- Myoglobin
- National Early Warning Score (NEWS)2 score
- Nausea/vomiting
- Neurologic recovery
- Neuromuscular function
- Nightmares
- Nutrition
- Organ failure
- Other infection
- Outcomes; respiratory, thoracic and mediastinal outcomes
- Overall muscle strength
- Oxygen saturation
- Pain
- Pressure of arterial oxygen to fractional inspired oxygen concentration (PaO2/FiO2)-ratio
- Participation
- Patient-centred morbidity rate (health related quality of life (HRQoL), time course of return to premorbid function)
- Percentage of cases returning to critical care
- Percentage of cases turning to severe or critical
- Physical function
- Physical symptoms
- Physiologic consequences (extent or time to resolution of study entry criteria etc., alterations in individual
  physiologic variables – e.g., pressure of arterial oxygen to fractional inspired oxygen concentration (PO2/FiO2)-ratio,
  mean arterial pressure (MAP))
- Physiological or clinical (e.g., cardiac outcomes; nervous system-related outcomes)
- Physiological response to exercise
- Post-intensive care syndrome (PICS) screening at hospital discharge

- Pneumonia
- Pneumonia severity index (PSI) score
- Pneumothorax
- Post intensive care syndrome (PICS) for family (PICS-F) before hospital discharge (PICS for family)
- Post-traumatic stress disorder
- Presence of delirium* while in hospital
- Progressing to the critical stage
- Progressing to the severe stage
- Proinflammatory cytokines
- Proportion of patient without cough
- Proportion of patient without wheezing
- Proportion of patients without fatigue
- Proportion of patients without fever
- Proportion of patients without sputum
- Proportion of pt. with negative SARS-CoV-2
- Psychological state
- Pulmonary function and symptoms
- Pulmonary function; lung function
- Pulmonary rehabilitation
- Rate of ALT recovery
- Rate of creatine kinase (CK) recovery
- Rate of mycobacteria (MB) recovery
- Rate of no requiring supplemental oxygen
- Rate of patient receiving systematic corticosteroids
- Rate of preventing mild to moderate patients from shifting to severe patients
- Rate of progressing to final stage
- Readmission/control visits
- Reintubation
- Remission rate of respiratory symptoms
- Renal function
- Resilience
- Respiratory function
- Respiratory muscle function
- Respiratory muscle function
- Respiratory muscle strength
- Respiratory rate
- Respiratory rate
- Respiratory support
- Return to work or prior activities
- Routine urine culture test
- Sadness/depression
- Satisfaction with life or personal enjoyment
- Score of traditional Chinese medicine (TCM) symptoms
- Self-efficacy/management

- Sepsis/septic shock
- Sequential organ failure assessment (SOFA) score
- Sexual function
- Sexual function and symptoms
- Short tempered
- Shortness of breath
- Shortness of breath
- Short-term morbidity rate
- Short-term mortality rate
- Single organ dysfunction/single organ failure
- Sleep
- Sleep function and symptoms
- Social roles
- Social roles, activities or relationships
- Successful extubation
- Survival
- Survival after leaving ICU
- Survived event
- Susceptibility to repeated infections
- Sustained return of spontaneous circulation (ROSC)
- Swallowing function and symptoms
- Temperature
- The proportion of inpatients
- The rate of discontinuations due to adverse events
- The time when the condition became worse
- Therapeutic target (alteration in target (e.g. reduced tumour necrosis factor (TNF)))
- Time delirium and coma free days
- Time delirium free
- Time of ALT recovery
- Time of Mb recovery
- Time of Mb recovery
- Time of severe acute respiratory syndrome coronavirus 2 ribonucleic acid (SARS-CoV-2 RNA) turns to negative
- Time of using assisted breathing
- Time to 2019-nCoV real time – polymerase chain reaction (RT-PCR) negativity
- Time to CD4 t cell recovery
- Time to CD8 cell recovery
- Time to clinical improvement (TTCI)
- Time to cough disappearance
- Time to cough reported as mild
- Time to CRP recovery
- Time to defervescence (the subsidence of a fever)
- Time to delirium onset
- Time to dyspnoea reported as absent
- Time to gastrointestinal symptoms reported as absent

- Time to hospital discharge
- Time to national early warning score (NEWS2) of lower or equal to 2 maintained for 24 hours
- Time to release for isolation
- Time to resolution
- Time to treatment failure
- Tinnitus
- Total costs
- Tracheostomy
- Troponin
- Tube placement (time needed or success)
- Tumour necrosis factor (TNF)-alfa
- Type of residence
- Type of residence
- Use of antipsychotic medications
- Use of non-invasive ventilation (NIV)
- Ventilator free days
- Ventilators parameters
- Viral load or clearance
- Vision impairment
- Walking distance
- Weakness
- Weight loss
- Wishing to have died

#### Rationale behind the initial core outcome set

*(This is the original and unedited text version shared with all collaborators)*

The consensus process for developing the primary core outcome set (COS) in Denmark is depicted (Figure 1) and shortly explained below. Initially, 329 outcomes were identified from a literature search. Duplicates or overlapping outcomes were removed and remaining outcomes were aggregated in categories. From these categories 50 outcomes were found to be representative. We also performed 87 semi-structured interviews to ensure that the list covered all outcomes before creating the survey, which it did. The 50 outcomes informing the modified Delphi survey were narrowed to 19 by the four research panels. Additionally, our national stakeholder group further reduced the 19 outcomes to 6.

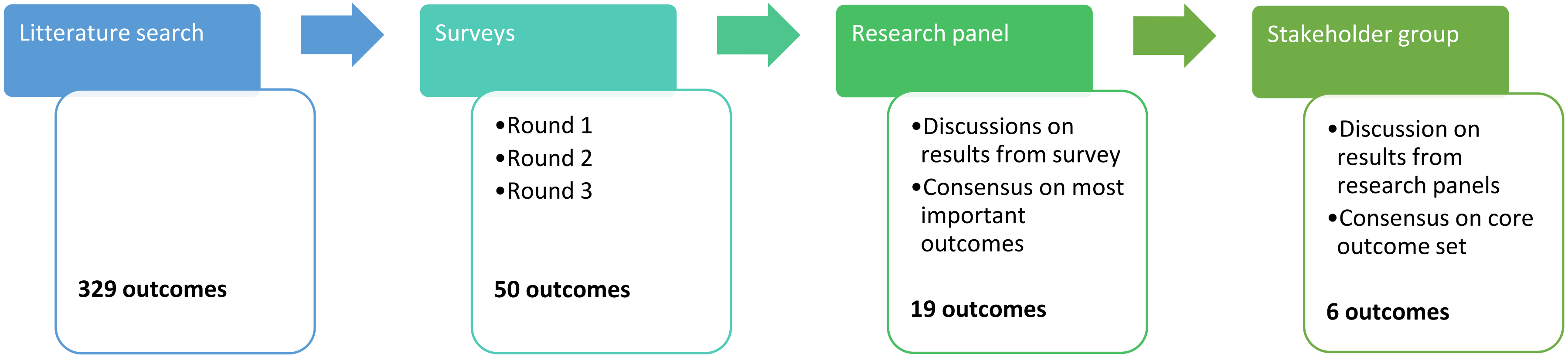


Figure 1. Consensus process for developing the primary COS

#### Survey

To determine which outcomes should be assessed across all clinical trials, patients, relatives, clinicians, and researchers were invited to participated in a survey consisting of three rounds. In each survey round, participants were required to grade the importance of 50 questions representing a specific outcome from 1-9, further categorized as *not important* (1-3), *important, but not critical* (4-6), and *critical* (7-9). Participants were instructed not to evaluate the questions in relation to one another, but to independently assess each question.

Each question was introduced with the phrase "How important is it to record or measure x?" Overall, the questions in the three survey rounds were the same. During the first round it was possible for participants to suggest unrevealed outcomes. These suggestions were presented to the research panels; however, no additions were made. We made a few changes in the wording of the questions and help-text between round 1 and 2 to clarify the understanding, and only minor changes between round 2 and 3. When answering the survey, participants were shown the results of the previous rounds with the aim of fostering consensus regarding critical outcomes. The heat-map (Figure 2) suggests that the process has triggered some changes along the way, while other outcome ratings were consistent.

The results also showed that patients and relatives participating in the survey did not rate any outcomes as not important, and only a few outcomes were rated not important by the clinicians and researchers. Thus, it wasn't possible to exclude any outcomes based on the survey alone.

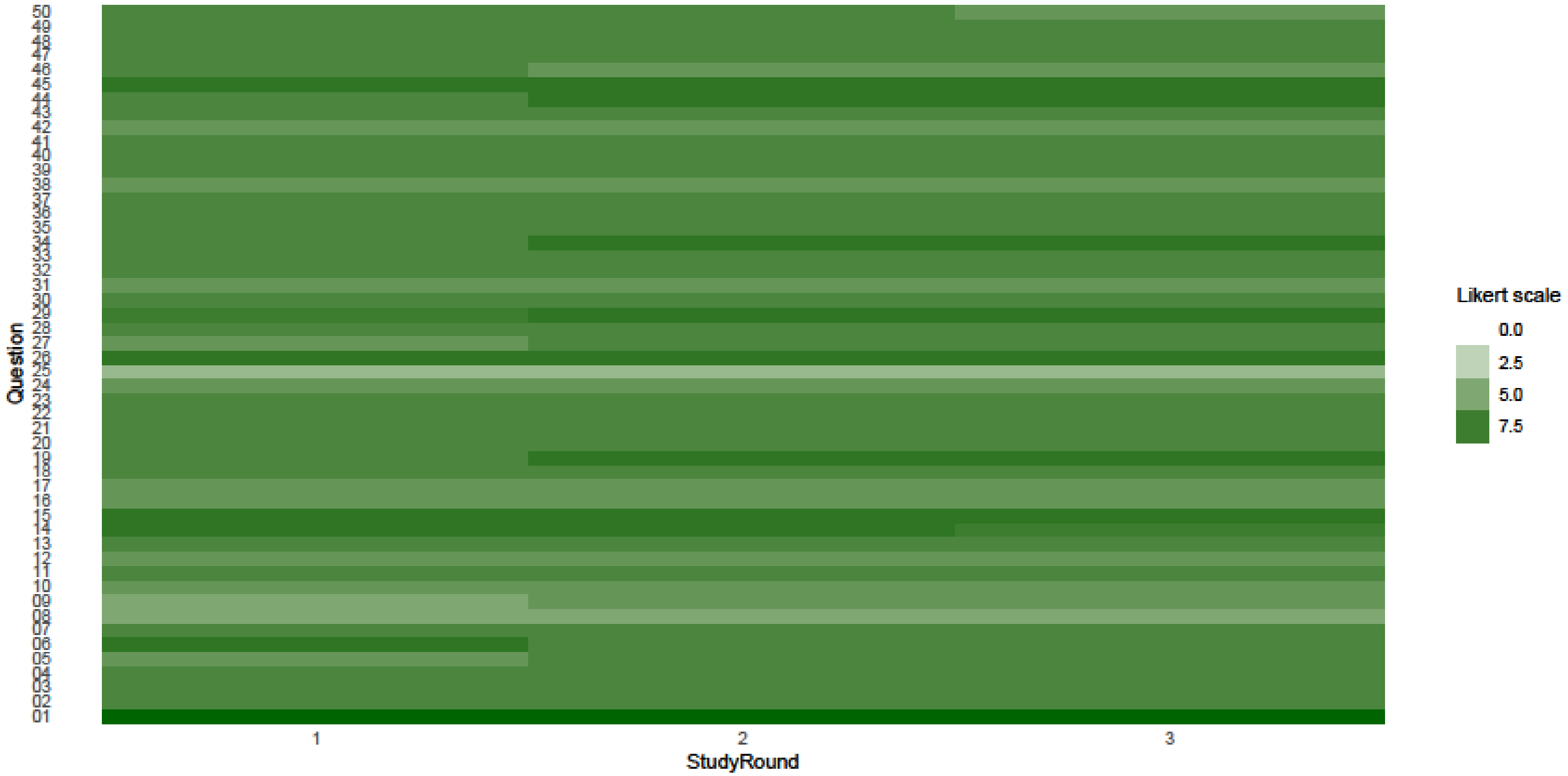


Figure 2. Heat map

All participants rated the 50 outcomes ("Question") on a Likert scale from 1-9, where 1-3 is 'not important', 4-6 'important, but not critical', and 7-9 'critical' to include this outcome. This was rated for each survey round (StudyRound) nr 1, 2, and 3. Greener represents more important outcome.

#### Research panels

Results of the survey rounds were presented to four research panels comprising patients, family members, clinicians, and researchers (31 people in total). The task of the research panel was to narrow down and sort through what outcomes they could agree were critical outcomes. Unlike the survey participants, the research panels had to consider the individual outcomes in relation to each other. Whether one outcome was more important than another.

As a result of this process, the four research panels ended up rating between 9 to 13 outcomes as critical outcomes to assess across all clinical trials. The cumulative number of critical outcomes reached 19, as they varied across the different research panels (Table 1). In several cases, the research panels included multiple outcomes under an inclusive outcome. This includes patients' well-being, health-related quality of life after ICU discharge, patients' independent performance of activities of daily living after ICU discharge, and mental health after ICU discharge.

As for well-being, the original survey question addressed the patients' overall well-being. However, all research panels found that 'well-being' referred to the patients' well-being during their stay in the ICU. Well-being outside the ICU was considered included in health-related quality of life after ICU discharge.

Finally, all panels reached a consensus that three outcomes were not important and were excluded from the further consensus process. These three were: 1) whether to measure blood values within normal limits while in ICU, 2) whether to record vital signs within normal limits while in ICU, and 3) whether to record new medications prescribed for the patient after ICU discharge.

Table 1. The cumulative number of the 19 critical outcomes across the four research panels

| Outcomes |
| --- |
| 1. Survival |
| 2. ICU length of stay |
| 3. Hospital length of stay |
| 4. ICU readmission rate during a hospital stay |
| 6. Patients' overall well-being |
| 7. Health economic consequences of a particular treatment in ICU |
| 11. Duration of mechanical ventilation while in ICU |
| 12. Duration of pharmacologic circulatory support while in ICU |
| 14. Duration of sedation while in ICU |
| 15. Duration of delirium while in ICU |
| 19. Experience pain while in ICU |
| 23. Discharge location after hospitalization |
| 26. Health-related quality of life after ICU discharge |
| 30. Family caregivers' experienced quality of life after the ICU stay |
| 34. Independently perform activities of daily living after ICU discharge |
| 37. Experience newly onset shortness of breath after ICU discharge |
| 39. New onset of speaking troubles after ICU discharge |
| 45. Mental function (cognitive function) after ICU discharge |
| 51. Psychological health after ICU discharge (new outcome added during the research panel consensus meetings) |

The colors refer to the number of panels who have agreed on the importance of each outcome. Dark green is 4/4 panels, light green 3/4 panels, yellow 2/4 panels, and orange 1/4 panels.

#### Stakeholder group

The national stakeholder group involved in the development of the COS for the general ICU population gathered to define the primary COS. The stakeholder group was asked to consider the results of the research panel discussions in relation to whether an outcome would be too difficult to define or measure (methodological considerations), whether an outcome would be too resource consuming (feasibility considerations), and whether an outcome targeted the entire heterogeneous population. Thus, the stakeholder group had to consider a broader range of factors influencing COS consensus than both research panels and survey participants. There was no predetermined maximum number of outcomes, but it was discussed that both Cochrane and GRADE recommend a maximum of 7 outcomes in a COS.

As a starting point for the meeting, the results of the research panels discussions were presented. Subsequently, the debate was centered on the 19 outcomes chosen as critical outcomes by the research panels. This discussion led to consensus on 6 critical outcomes defining the primary COS (Figure 3). The rationale behind each outcome is explained below.

Figure 3. Primary COS

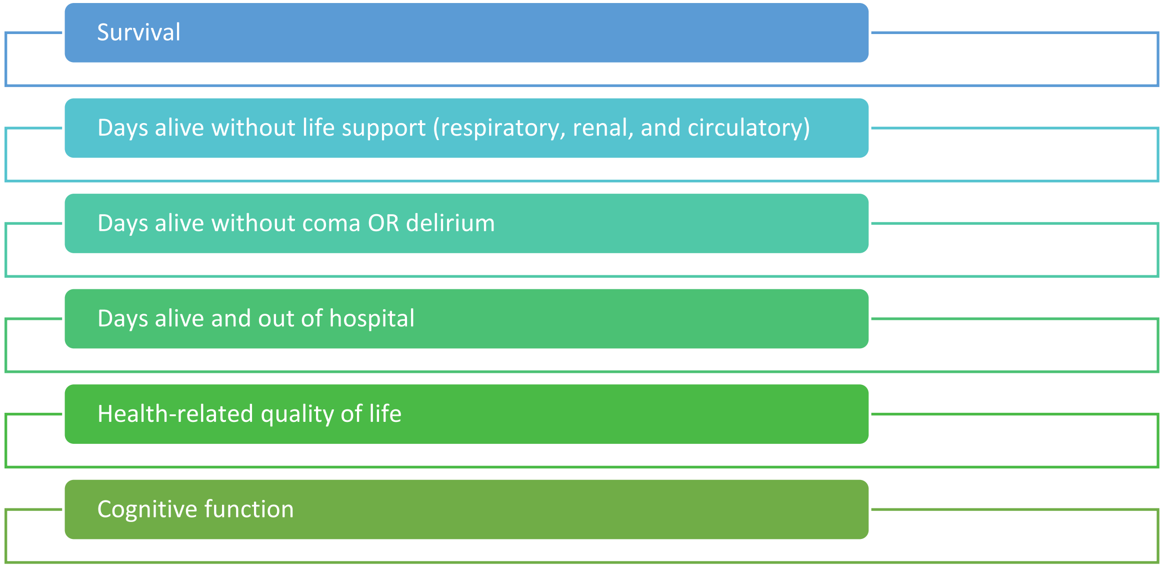

#### Rationale behind each outcome

##### Survival

The original question in the survey was "How important is it to record ICU survival?" The stakeholder group agreed that ICU survival was not an optimal outcome due to the considerable variation in the definition of intensive care units worldwide. However, survival was considered as a critical outcome among all participants in both survey, research panels, and the stakeholder group. But the outcome 'survival' is to be recorded at predetermined timepoints.

##### Days alive without life support (respiratory, renal, and circulatory)

This outcome expresses organ failure and disease severity, consisting of various elements from the survey, including the duration of mechanical ventilation, circulatory support, and dialysis. Each intervention represents a different aspect of organ failure. The stakeholder group considered days alive without life support as a better measure of severity compared to time in the hospital, readmission to the ICU, or time in intensive care. For example, days in intensive care could be synonymous with the various treatments we offer our patients." For both mechanical ventilation, circulatory support, and dialysis, it was discussed whether different practices could affect the use of the outcome. Given substantial variations, the degrees of treatment need to be defined. However, this should occur after external validation, as relevant comments may influence the definition.

##### Days alive and out of hospital (DAAOOH)

Admission time at hospital and at ICU was discussed as well as re-admissions, but there may be large variation in practice. When is a patient admitted to an ICU and when is the patient discharged, this could be according to resources, when looking across countries. Therefore, DAAOOH may be a more neutral and covering outcome, which is also well known as a trial outcome. The elements of DAAOOH like length of hospital stay/ICU stay and re-admissions is graded critical according to the participants and is integrated in DAAOOH.

##### Health-related quality of life (HRQoL)

All participants agree that HRQoL is a critical outcome. HRQoL covers many outcomes (Table 3) including the patient's physical, mental, and social health. It was discussed that well-being may be a part of HRQoL. Also, it was discussed if post intensive care syndrome (PICS) could constitute the overarching outcome to cover both HRQoL and cognitive function. But the stakeholder group agreed that this outcome is not that well performing as a core outcome, yet, and there is uncertainty about how to assess it. All the underlying outcomes will be taken into consideration when deciding what tool to use for assessing HRQoL. Whether HRQoL should be assessed at fixed timepoint is also to be discussed.

**Days alive without coma OR delirium**

Both duration of coma and duration of delirium were rated as critical outcomes in 3/4 research panels. As well as respiratory failure, delirium is a prominent organ failure among many ICU patients. The stakeholder group discussed whether delirium and coma could be included in the aggregated outcome Days alive without life support. One argument against doing this, was that it would be easier to compare future data with data from previous trials. Also, life support refers to interventions, whereas delirium or coma refers to a syndrome or condition.

**Cognitive function**

This outcome was not much debated, due to consensus. However, it was pointed out, that the outcome must be measured at a fixed timepoint.

#### Other outcomes not included in the primary COS

**6.Patients' overall well-being.** The degree of well-being was critical for three out of four panels as a core outcome, but due to its broad definition and not being realistic to assess in all trials (though the comfort-scale may be close), it was not included as a core outcome. Several outcomes were found to be related to well-being, with well-being serving as an umbrella term that encompasses outcomes such as pain, shortness of breath and anxiety during ICU admission (Table 2).

**7.Health economic consequences of a particular treatment in ICU.** This outcome was considered to comprehensive to evaluate in all clinical trials. Depending on the tool assessing health-related quality of life this outcome may be optional to calculate.

**19.Experience pain while in ICU.** Is a subjective outcome and can be difficult to assess if the patients can not cooperate. However, this outcome is covered by the well-being outcome and must be considered important when defining measurement tools for well-being.

**23.Discharge location after hospitalisation**

**30.Family caregivers' experienced quality of life after the ICU stay.** Is considered as a part of the patients' HRQoL. Caregivers' experienced quality of life may have a mutual impact on the patients HRQoL.

**34.Independently perform activities of daily living after ICU discharge.** Regarded as a part of the patients' HRQoL.

**37.Experience new onset of shortness of breath after ICU discharge.**

**39.New onset of speaking troubles after ICU discharge.** Regarded as a part of the patients' HRQoL.

**51(new outcome). Psychological health after ICU discharge (new outcome added during the ´consensus meetings).** Regarded as a part of the patients' HRQoL.

Table 2. Outcomes related to well-being

| |
|---|
| 16. Nausea and vomiting while in ICU |
| 17. Thirst while in ICU |
| 18. Short of breath while in ICU |
| 19. Pain while in ICU |
| 20. Anxiety while in ICU |
| 21. Sleep while in ICU |
| 22. Ability to communicate while in ICU |

Table 3. Outcomes related to health-related quality of life

| |
|---|
| 24. Record patients' outpatient or general practitioner visits related to the long-term effects of intensive care |
| 27. Newly onset tiredness or fatigue, unrelieved by sleep or rest, after ICU discharge |
| 28. Newly onset pain after ICU discharge |
| 29. Resume their usual roles in daily life after ICU discharge |
| 32. Record whether patients after ICU discharge, compared to before, have the same labour market attachment |
| 34. Independently perform activities of daily living after ICU discharge |
| 35. Independently able to get dressed after ICU discharge |
| 36. Mobile after ICU discharge |
| 38. New onset speaking troubles after ICU discharge |
| 39. New onset of swallowing difficulties after ICU discharge |
| 40. Bowel function after ICU discharge |
| 41. Urinary function after ICU discharge |
| 42. Sexual function after ICU discharge |
| 43. Senses: vision, hearing, touch, smell, and taste after ICU discharge |
| 44. Physical strength to perform activities of daily living after ICU discharge |
| 46. Loneliness after ICU discharge |
| 48. Anxiety after ICU discharge |
| 49. Depression after ICU discharge |

The colors refer to the number of panels who have agreed on the importance of each outcome. Dark green is 4/4 panels, light green 3/4 panels, yellow 2/4 panels, and orange 1/4 panels

#### Table S5. Summary minutes from all participating countries holding an international validation meeting

| | **Survival** | **Number of days alive without life support** | **Number of days alive without coma or delirium** | **Number of days alive out of hospital** | **Health-related quality of life** | **Cognitive function** | **Additional** | **Evaluation/ conclusion** |
|---|---|---|---|---|---|---|---|---|
| **Australia** summary for both the general ICU patient and the sepsis patients<br><br>(*Summary separated in the following two rows)* | There was near unanimous agreement to accept this measure, acknowledging that it should not be the only measure and that the measurement time point is important and may vary between studies. | Accepting this measure was supported, but with a substantial minority favouring adapting the measure with further specification, including for example adapting the measure to specify relation to life support machines and ensuring that each component is also considered individually. | There was no support for accepting this measure and a majority favoured rejecting the measure. Coma and Delirium were not considered equivalent or appropriate to include in a single measure. | This measure was generally supported. A consistent theme was that it was being at home and not simply out of hospital that was important. Another consistent theme was that being at home was considered important by ICU survivors but that this outcome measure was questions by clinicians and researchers as more a resource than patient-centred measure. | This measure was unanimously supported, noting the need to be able to compare to quality of life prior to intensive care and that quality of life valued by one person may be different to another. | There was no support to accept this measure. Predominant view to reject based on cognitive function being an essential component of quality of life. | The main domain considered important but not currently included in the proposed core outcome set were measures of psychosocial wellbeing, contribution to society and place in community, and return to work. | |
| **Australia, New South Wales** summary for sepsis patient | Survival is important. Shouldn't be the sole outcome - the time point is equally important. Median to longer term survival (60/90days – 6/12mths). Stable differential timepoint for a trial, longer time to results if main trial follow up outcome is too long, also | May not be sensitive enough. Very blunt instrument. Very difficult to implement. Quality of the data is varied across hospitals (delirium) Patient perspective – delirium was very impactful and linked with PTSD – would want this as an outcome or adapted within the cognitive outcomes. | May not be sensitive enough. Very blunt instrument. Very difficult to implement. Quality of the data is varied across hospitals (delirium) Patient perspective – delirium was very impactful and linked with PTSD – would want this as | Supportive but more detail required – what is considered out of hospital. Place of discharge – is it the same as the place prior to admission? Days alive and at home perhaps a better description. Measuring this outcome is hard. Analysis hard. We | All agreed this outcome was important and should be part of the minimum standard BUT the caveat is how we measure it and with what instrument. Sepsis survivors put forward that the measures for this outcome may be different for sepsis vs general | Most thought this outcome was limited to only one component of psychological function and if cognition is to be included separately to HRQoL, so should the other psychological components. Discussion to expand to psychological | | |

| | Survival | Number of days alive without life support | Number of days alive without coma or delirium | Number of days alive out of hospital | Health-related quality of life | Cognitive function | Additional | Evaluation/ conclusion |
|---|---|---|---|---|---|---|---|---|
| | increases loss to follow-up, trial practically and costs. Reference to Marshall study All agreed as a minimal outcome. | Most agreed this outcome shouldn't be part of a minimal core outcome set. Adapt delirium under cognition or HRQoL – downstream effects may be picked up with HRQoL and qualitative studies. 9 voted to reject – 3 to adapt into the cognitive or HRQoL outcome. | an outcome or adapted within the cognitive outcomes. Most agreed this outcome shouldn't be part of a minimal core outcome set. Adapt delirium under cognition or HRQoL – downstream effects may be picked up with HRQoL and qualitative studies. 9 voted to reject – 3 to adapt into the cognitive or HRQoL outcome. | don't know enough about it. Is this about resource use rather than patient centred outcomes. Should be picking up the patient outcomes in other outcomes. Potentially very varied within health systems 10 agreed with the outcome but that it should be adapted to not be out of hospital (i.e., days alive and at home). 1 rejected. Created a lot of discussion that this outcome is used a lot but perhaps is more about resource use than patient centredness. | critical illness. Consultation from sepsis survivors required. Important in trials to recognise that caregivers' outcomes are important and should be incorporated in to the follow up. All agreed with this as a core outcome (with the caveat that it is important to have stakeholders discuss what will be measured and with what instrument. Sepsis may be different to general critical illness) | function and not only cognitive function, i.e., include depression, anxiety, PTSD and psychosocial health. Wellbeing, communication, cognitive communication was also thought to be important to be included in this outcome. Delirium to be adapted in this outcome. Adapt. All agreed that this outcome should be adapted to include the broader psychological and psychosocial functioning (which includes cognition). Cognitive function should not be the only measure included in a COS. | | |
| **Australia Perth, Western Australia + virtual** Summary for the general ICU patient | High degree of consensus that this outcome is 'key' and that survival is the main objective of intensive care treatment on which other outcomes are contingent. Issued considered included | General agreement that this is an important outcome measure but that it should specify that life support related to machines (e.g. breathing machines, pumps for continuous | Predominant view to adapt as coma and delirium considered to be very different entities and not appropriate to measure as a single outcome by attendees due to a | Predominant view to adapt, primarily recognising that it is essential that home is important and recognised, not other places outside of | A unanimous view that this was an important core outcome measure, noting the need to be able to compare to quality of life prior to intensive care and that quality of life | Predominant view to reject based on cognitive function being an essential component of quality of life. | Physical function outcome measure (e.g. ability to achieve independent mobilisation) Look at the patients ability to return to occupation/school Contribution to society- Would be | |

| | Survival | Number of days alive without life support | Number of days alive without coma or delirium | Number of days alive out of hospital | Health-related quality of life | Cognitive function | Additional | Evaluation/ conclusion |
|---|---|---|---|---|---|---|---|---|
| | survival versus quality of life, the need for defined and potentially multiple measurement time points and the challenging terminology of 'survival' as a long-term outcome in relation to intensive care treatment (i.e. can survival still relate to intensive care long after discharge?).<br><br>Quotes, comments, and themes:<br>If the patient does not survive, the rest of the COS are not relevant; the pt may survive ICU but may not survive their hospital admission, "it is important to look at survival long-term not just ICU/hospital survival"<br>"Survival is a very important aspect to look at"<br>To also look at the quality of survival; if a pt survives but has poor function, this is important to know<br>Questions about the timepoint in which this COS measuring; | intravenous infusion of medicines), and that should specify new life support to differentiate from patients with pre-existing needs for machines including dialysis.<br><br>Quote, comments, and themes:<br>It would also be important to have a measure of the impact on the body "Should life support specify machines?"<br>Important to have this definition defined, "what do we mean by life support?"<br>This core outcome measure is vague and needs clarification<br>In terms of ECMO this is an "Important measure"<br>Multiple opinions to adapt this COS to "life support machines"<br>Not all machines may apply for all research projects | number of factors including the very different causes (medically induced or illness induced) of coma, that delirium may have more experiential impact on ICU patients than coma, and that long-term outcomes are more important than this short-term outcome.<br><br>Quote, comments, and themes:<br>This core outcome needs specification; what do we mean by coma (induced or not induced); not sure is coma should be stated together with delirium<br>May not be very valid for research trials as certain clinicians' sedate patients more than others; there are going to be a lot of variables between practice of sedation depending on each clinician; physiological function is more | hospital like a nursing home or rehabilitation facility.<br><br>Quote, comments, and themes:<br>This core outcome is important as it also links with survival; alive and out of hospital is important but it is also important to look at how much time a patient has spent at home; cannot look at this in isolation because it will not give enough information<br>Going forward we probably have more transition of hospital to community-based healthcare, how will this affect this core outcome measure? | valued by one person may be different to another.<br><br>Quote, comments, and themes:<br>"This core outcome will be very important", nurses tend to establish a relationship with a patient so it would be nice to know how the patient is, are they living the life they want to?<br>"An important measure; to see if a patient can get their life back on track is very meaningful to see"<br>Does quality of life need to compare pre-ICU admission to post ICU?<br>Which tool will be used to assist with this measurement? If we don't have a tool that everyone agrees on it will be hard to compare outcomes globally | <br>Quote, comments, and themes:<br>It would be better to measure functional impairment than cognition itself. Cognition is multifactorial May be better to look at whether a patient can go back to work/study/school rather than what is their memory like?<br>Would this core outcome exclude people who had brain surgery?<br>Important to consider people who had cognitive surgery therefore may need to adapt COS<br>Patients who had their cognitive function measured found it stressing and confronting; patients said they felt like being tested and was very anxious, felt "down" that they did not do well on test<br>To measure how the patient is prior vs post ICU, cognitive function | important to know if patient is able to return to work, can't provide for family, or the extend they are able to contribute to community<br>Important to also look at patient wishes, survival may not be a good thing for every patient<br>Supporting network: Would be good to what social support the patient may have access to.<br>Important to look at study tools used in research to best capture these outcome | |

| | Survival | Number of days alive without life support | Number of days alive without coma or delirium | Number of days alive out of hospital | Health-related quality of life | Cognitive function | Additional | Evaluation/ conclusion |
|---|---|---|---|---|---|---|---|---|
| | to look at survival itself is very vague, which timepoint in ICU would be appropriate to see a correlation<br>Questions on how consistent ICU missions are at each hospital, and how this might influence outcome measures<br>Hospitals may not all have the same written mission statement; these COS is to be used for research purposes to determine if care has helped the pt to become better; main purpose is to determine what outcomes are important to the patient that should be monitored whilst conducting a research project<br>Thoughts to adapt this COS and will depend on a timepoint; a longitudinal approach is good; to look at not just one timepoint but multiple timepoints (3M,6M,12M)<br>Unsure as this COS may be important to families but may not be important to the | | important than delirium and may need to be a part of a separate core outcome<br>Delirium can be more impacting than being in coma; first 24h out of coma can be a worse experience than coma.<br>“Delirium is a huge consideration, it was the sickest and worst during my admission”<br>Measure of delirium is more important as it is the main issue that is causing a patient to stress; days out of ICU and in ward traumatic with delirium<br>Coma and delirium are two separate things, these outcomes needs to be separated<br>There are other things that have effect on a patient’s long-term outcome; not sure if this core outcome should only focus on delirium and not the other factors | | | could fall under quality of life<br>Patients performed a cognitive test and found it confronting, test could be affected by multiple factors such as quality of sleep, may have had a stressful morning. Patients felt as though they may have done better if the call was scheduled on another day, there may be variability on health status when reporting on measures<br>This core outcome may not be relevant; cognitive function could be part of quality of life | | |

| | Survival | Number of days alive without life support | Number of days alive without coma or delirium | Number of days alive out of hospital | Health-related quality of life | Cognitive function | Additional | Evaluation/ conclusion |
|---|---|---|---|---|---|---|---|---|
| | pt; may be difficult to adapt<br>Survival is important but not sure if it should be a COS; "cannot put everybody in same basket", what may the quality of life be if the pt survives but is left with cognitive impairment<br>Survival is important in ICU, but survival as a COS may be a "waste of a measure", "Survival should be an assumed outcome," and if the pt survives we should measure other things relating how well did they survive<br>Questions regarding if the pt survived well, why is there a post survival measurement? It was confusing for pts when they have survived but studies look at mortality within 3yrs-5yrs<br>It is important to look at survival to determine whether an intervention is safe | | | | | | | |
| **Czech Republic** | Was accepted by 100% of panel members but survival with | Days alive without life support (respiratory, renal, and circulatory) – six | Days alive without coma OR delirium – outcome accepted by all | Days alive and out of hospital – outcome was again not as | Health-related quality of life – the panel considered | Cognitive function – outcome was accepted by 100% of panel members, | | Overall everyone in the meeting felt it was successful and very |

| | Survival | Number of days alive without life support | Number of days alive without coma or delirium | Number of days alive out of hospital | Health-related quality of life | Cognitive function | Additional | Evaluation/ conclusion |
|---|---|---|---|---|---|---|---|---|
| | meaningful cognitive outcome was favoured. | out of eight panel members accepted. The outcome was not so important for patients as long as comfort, cognitive function, communication and hope were present. Outcome was important for clinicians and researchers, family members. E.g. length of ventilation not considered important as long as dyspnoea was not experience. (I'd rather be ventilated longer, earlier intubation or later extubation than to experience dyspnoea) Comfort included important aspect of the ability to communicate with staff and family – this helped to avoid/limit anxiety. Pain was also included but could be tolerated as long as other aspects, in particular, communication were attended to. | and considered more important than outcome | important for patients as long as comfort, cognitive function, communication and hope were present. Outcome was important for clinicians and researchers, family members. | four domains of QoL Physical – less important for patients, family members more important for clinicians and researchers Psychological – more important for patients, even more for family members Non-religious spirituality – important to be able to integrate ICU stay and critical illness into the context of one's life Social – family presence and a degree of self sufficiency important aspect It was noted that physical domain is not as important for patient and families as the other 3 domains of QoL | psychological health considered very important but very inter-individual. | | important to discuss these issues and all agreed that new information and perspectives came to light. They were all happy to continue the discussions in the future. |
| **Finland (Helsinki)** | It was discussed and unanimously stated | It was discussed and unanimously stated | It was discussed and unanimously stated that the | It was disc38art and unanimously stated that the | It was discussed and unanimously stated that the | It was discussed and unanimously stated that the | It was discussed and it was unanimously concluded that 6 | What worked well was that participants were |

| | Survival | Number of days alive without life support | Number of days alive without coma or delirium | Number of days alive out of hospital | Health-related quality of life | Cognitive function | Additional | Evaluation/ conclusion |
|---|---|---|---|---|---|---|---|---|
| | that the variable is important. Comments; Patient: It is absolutely important that this is measured. However, it has felt strange when, after intensive care, a survey was sent home, asking about being alive, and not being more interested in health. Doctor: It is important to include this variable because it can be clearly measured. Death is a significant outcome in intensive care. Doctor: Surviving the intensive care unit alive is important, but this alone is not enough, you have to look at how the patients survive holistically. Patient: If the person is satisfied, the treatment has gone well | that the variable is important. Comments; Doctor: Hospital and doctor-centric measure, commits the patient to the intensive care unit. It is significant in terms of ICU performance and resources. Doctor: In terms of resources, it is significant from the point of view of costs to the society. Patient: With vital function support treatments, the patient can live normally even outside the hospital. Doctor: The criteria for starting and stopping ventilator treatment and other life support in studies should be the same in all countries. Patient: But do the outcome variables work in every country with different income levels. | variable is important. Doctor, researcher: Unconsciousness can be due to sedation or injury Doctor: Delirium is very critical in intensive care. It is very welcome that this is raised and included in the variables. Patient: It is good that this is taken into account, the patient's delirium or coma is also, very difficult for the relatives. Doctor: Intensive care environment predisposes to delirium, drugs can also cause it. | variable is important. However, it should also be specified how to define outside the hospital (rehabilitation, supported housing, hospital bed department, etc.) Comments Patient. This is a significant treatment outcome variable. Doctor: For many, it is important to return home. It should be defined what means outside the hospital. Patient: It is important to return home or to where you came from. | variable is important. The quality of life measure should always include physical, psychological and social quality of life. Comments Doctor: Quality of life is routinely measured after intensive care, looking at different areas of life, and general health (on a scale of 0–100). Patient: It can be difficult to define the perceived state of health, or to relate the assessment of the state of health to one's own normal condition. Doctor: The survey is always subjective, a person can have functional limitations and still be satisfied with their quality of life. Patient: The quality of life indicator should also take into account relatives | variable is important. It was considered whether adverse events in intensive care should also be taken as a separate core outcome variable Comments Doctor: Will there be a problem if we screen. In this case, there should be a readiness to refer to further investigation if necessary. Doctor: If cognition deteriorates, it affects everything about recovery, quality of life and the patient's loved ones. | core outcome variables of intensive care can be considered suitable for Finnish conditions. | eager to discuss and all thought this was interesting and important. There were no feelings of difficulties to speak out one´s opinions e.g. due to hierarchy or other obstacles, the environment at both library rooms was relaxed. The structured approach by using the standardised power point presentation (translated to Finnish) was good base for the introductory phase. The video was also good to have. Things that were slightly challenging: In both meetings it was sometimes necessary to repeat that the focus is in general ICU research and comparison of the study groups It is challenging for the lay people to fully understand the principles of |

| | Survival | Number of days alive without life support | Number of days alive without coma or delirium | Number of days alive out of hospital | Health-related quality of life | Cognitive function | Additional | Evaluation/ conclusion |
|---|---|---|---|---|---|---|---|---|
| | | | | | Patient: It would be good if there was a customized self-care questionnaire for each intensive care patient | | | clinical studies (randomization etc), when explained only shortly<br>Some outcomes are essentially measured within a given time limit (days alive without...) and it might have become easier to grasp if the tools would have been already chosen (QoL, cognition etc)<br>Sometimes the discussion drifted to the patients own recovery process that they wanted to share,, but that was allowed.<br>For me as a coordinator in the first meeting it was slightly challenging not to interfere too much with my own opinion. I felt that I could have stressed the importance of delirium more and maybe interfered, but I chose not to, for keeping the authenticity. |
| **Finland (Tampere)** | It was unanimously agreed that survival | This was generally perceived slightly | J.H. first explained delirium to the | All: This was largely discussed | Patients: This should be | Patients and family members: | Patients and family members suggested | Same as for Finland (Helsinki) |

| | Survival | Number of days alive without life support | Number of days alive without coma or delirium | Number of days alive out of hospital | Health-related quality of life | Cognitive function | Additional | Evaluation/ conclusion |
|---|---|---|---|---|---|---|---|---|
| | is an important outcome and belongs to the core outcome set. Doctor: It has been the traditional measure of outcome and it is easy to measure and absolute of nature Doctor: It is a condition that has to be fulfilled for all the other outcomes to be achieved Patients and family members agreed to including this outcome in the COS. | less clear. Coordinator explained the concept and the benefits in study populations with high mortality when mortality is a competing event. And it was explained that this outcome always is measured within a certain time period. Doctor: This outcome requires that the criteria of initiating and ending a certain treatment need to be well defined. We also discussed the impact of different centres and their practices in this context. Doctors and patients: This may not be so much of importance to the patient. Doctor and researcher-doctor: But it may be important when calculating the benefit to the organization, economics, and thus benefit to society. Doctors: The life support is actually close to a measure | panel. The concept of coma was a bit difficult, and we discussed whether it covers both pharmacologically induced and non-pharmacological coma. This may have caused some confusion. The patients shared their experiences, both had experienced delirium. One of them had had delirium with only positive content in the delusions. The patients and family members thought that delirium is important but were not sure whether it should be included in the COS. Doctor and doctor-researcher: Delirium is important but can be slightly less easy to detect in many cases. They said that it is a good outcome in some studies but were | from the aspect of what out of hospital means. What will be counted as out of hospital, for example in cases when healthcare institutions are still involved. Patients: The patients want to go home, and sometimes this requires extra care that can be provided from healthcare providers. Doctors: So this depends on the systems in each participating center. Conclusion: This was however considered an important outcome and to be included. | interpreted carefully, whether there are some new unrelated problems or is the result reflecting problems related to intensive care. Coordinator: Explained that this is a composite and that when comparing groups, both groups should have equal amounts of "external causes" in addition to the ICU related causes. Patients: How would cultural differences be taken into account? Patients and family members: It is important to know that quality of life can also improve due to different values a�er critical illness. Doctors and researcher: The validity of this measure is dependent of which tool is going to be chosen | This is a very important outcome to be included in the COS, it41articts many aspects of life. Doctors and researchers: Agree We discussed a li�le how this can be measured and how the baseline situation is affecting the results. Conclusion: It was unanimously agreed that this should be included in the COS. | that the well-being of family members should also be studied as an outcome. | |

| | Survival | Number of days alive without life support | Number of days alive without coma or delirium | Number of days alive out of hospital | Health-related quality of life | Cognitive function | Additional | Evaluation/ conclusion |
|---|---|---|---|---|---|---|---|---|
| | | that determines the need of ICU care. Patient and family member: How well does this outcome describe recovery? Patient and family member: In our experience, discharge from ICU may also be an impairment, because the sudden drop of the time when personnel are available. This was discussed and well understood. We also discussed whether the number of readmissions would be influencing this outcome. Conclusion: Consensus was reached that this outcome is important and should be in the COS | slightly hesitant to have it included in all studies. On the other hand, they pointed out that this is also an organ dysfunction, and therefore important. Conclusion: There was some hesitance in accepting this outcome to the COS. | | and what is the timing of the assessment. Conclusion: This was accepted conditionally. | | | |
| **Iceland** | All individuals in panel agreed that this was an important outcome, both for the patient as well as the clinicians. No barriers for measurement in Iceland | All individuals in panel agreed that this was an mportant outcome, both for clinicians and patients. No barriers for measurement in Iceland. Discussions revolved around timing of assessment, and how various support would need to be | All individuals in panel agreed that this was an important outcome, both for clinicians and patients. No barriers for measurement in Iceland. Discussion revolved around the varying aspects of delirium | All individuals in panel agreed that this was an important outcome, both for clinicians and patients. No barriers for measurement in Iceland. Discussion revolved around the importance of | All individuals in panel agreed that this was an important outcome, both for clinicians and patients. No barriers for measurement in Iceland. Also that this is frequently not measured, and would be a big | No comments | No comments | There was a lively discussion amongst all participants, and all agreed that the outcome set was very patient-centred – which was positively regarded by all. There was some discussion about a lack of outcomes |

| | Survival | Number of days alive without life support | Number of days alive without coma or delirium | Number of days alive out of hospital | Health-related quality of life | Cognitive function | Additional | Evaluation/ conclusion |
|---|---|---|---|---|---|---|---|---|
| | | clearly defined – as some organ support is possible outside of the ICU depending on hospital resources. | and its fluctuating course, where a person might classify as having or not having delirium based on time of assessment. This would increase inaccuracy of this measurement, and mandates that the measurement is very clearly defined. | defining what "out of hospital means" – outpatient rehabilitation, nursing home etc. Also that this outcome not only measures the ICU treatment but also the hospital treatment and rehabilitation, dependent on timing of assessment. | benefit of a more global adoption of core outcome set. | | | describing more closely the ICU course of participants, but overall felt that at least three of the outcomes could reflect this. For things to improve, there was a discussion whether readmission rate within a specific timeframe would be beneficial. In general, some examples of potential measurements would have benefitted the discussion and ensured that all stakeholders had a mutual understanding of what individual outcome could mean. These examples were provided to facilitate understanding when prompted by the primary investigator for Iceland, emphasizing that the exact timing and choice of tools to measure were |

| | Survival | Number of days alive without life support | Number of days alive without coma or delirium | Number of days alive out of hospital | Health-related quality of life | Cognitive function | Additional | Evaluation/ conclusion |
|---|---|---|---|---|---|---|---|---|
| | | | | | | | | without the scope of the meeting. |
| **India (Chennai)** | Survival was explained (not just ICU survival) and explained that the length of survival is still debated but the consensus has been that it is one of important outcomes. From the perspective of intensive care patient, when they go into ICU, they are concerned about their life. Ultimately what does matter, including for the family member is whether they are going to come out alive from ICU. There is no doubt that mortality/survival is going to be a very valid outcome. Survival is a hard outcome compared to other outcomes that can be manipulated. For eg. days in ICU (dependent on intensivist if he wants to keep patient for an extra day or bed availability in the ward). These are | It was explained what the outcome means through examples and asked if anyone has any doubts regarding the outcome understanding and opened floor for discussion. There was confusion even amongst intensivists. The person must be alive and free from any kind of support. If the person has lived for 15 days without support and then passes on, then their days are counted as zero. Another point is that sometimes patient's family members insist on taking the patient back home for providing a peaceful end (DAMA, LAMA etc.), in trials we report them as alive, but we really don't know what happened to them. We should keep these in mind while discussing this COS. It may be important to know the severity of the health condition; based on | It was discussed that delirium and Coma should be covered in the mental health component of post ICU evaluation? It was clarified that in this case the outcome was being measured while in hospital or until a prespecified time point (to be determined). This could be a part of being free of organ support. As clinician it is our goal to keep the patient as awake as possible and limit the harmful impact of sedation. That's why this outcome is considered separately. This can be modified by clinician so I am not convinced that it can be an outcome. In ICU everything we do is directed towards the brain. It is not directly supported, but | Going to hospital and turned into vegetative state was discussed, I am still alive and out of hospital but that's not the quality of life I would want. So, its shades of grey. It takes a toll on care giver as well. We should have scales like GOSC or RANKIN Score where its not black and white but shades of grey. Having a metric that incorporates quality of life would make the outcome more relevant. It may be statistically important in clinical trials but in GOSC the story would be different. As a core outcome parameter, I am little hesitant to say this is essential. We are going to cover the health-related quality of life in the next outcome. | This outcome talks about the several aspects of health physical health, social health, mental health, and cognitive health, after discharge from ICU. Time points are not defined. The parameter should also consist of how nutrition improves post critical care to be added to HR-QoL. Three aspects to be considered, viz it should be easily measurable, it should be repeatable, and it should be replicable. | No disagreements. **Consensus was achieved on adopting this outcome.** | It was discussed whether financial consequences/health economic consequences should be mentioned here. It is more relevant in Indian setting. Can we have access to all the other outcomes? They might be more relevant to us in Indian setting. | |

| | Survival | Number of days alive without life support | Number of days alive without coma or delirium | Number of days alive out of hospital | Health-related quality of life | Cognitive function | Additional | Evaluation/ conclusion |
|---|---|---|---|---|---|---|---|---|
| | subjective, soft outcomes and may be manipulated but survival can't be manipulated. It is definitely important, at what timepoint, that may be dialogued further. As a caregiver, lay person, for me, survival is one of the reasons we agree for trials, even without knowing much of background about it. We understand that the patient needs it. Survival is the 'must have' outcome. She was reminded of the incident where doctors told her husband is a cat on the wall, at that point of time, it was the survival which was most important thing to consider. Bhuvana: Survival is the most important outcome for any patient. When it comes to ICU, it's even more important. We can always think about the timeframe. It is a patient centred outcome and important for doctors as well. | that, we can decide if the outcome is important. Also, this outcome is important because being alive and hooked up to machines affects the quality of life and survival as the patient may deteriorate. There should be a corollary when we talk about ventilation because hi-flow nasal therapy which is not conventionally classified as ventilator. There would be patients on this kind of support. How do we classify those patients? Second point is, patients with low blood pressure, we have to personalize targets. We say that is the MAP is above 65, we cut off vasopressor support, irrespective of whether we think the patient needs a higher map or not. This is a grey zone. It was summarized that there is lack of clarity about the non-invasive type of respiratory support, and whether the | indirectly. It is a patient centric outcome. We may need to go in details to refine the outcome so that it is measured appropriately. VJ: It reflects the special value that is placed by the stakeholders on brain function. It's important to hear from patients and caregivers whether they think it is an independent and important outcome. Shree Lekha: It is obvious that I would want my family member to be communicating to me because it makes me believe that they are recuperating successfully, despite the machines attached. Doctors may give reassurance, but the blink of the eye or wave of hand is a good sign for me emotionally. Does the parameter have | It is an essential outcome as a single measure. Of course, quality of life can be looked at separately. It is important to know that you can be alive and survive outside hospital for a longer period. Most of us are confusing between the technical issues related to analysis vs what is relevant to the patient and their families and clinicians. This outcome is also going to capture the resources that's used and trauma to the patient and their families and to the healthcare systems. Days alive does take these factors into account. The 3rd and 4th outcomes are overlapping but not necessarily identical parameters. Getting into hospital again | | | | |

| | Survival | Number of days alive without life support | Number of days alive without coma or delirium | Number of days alive out of hospital | Health-related quality of life | Cognitive function | Additional | Evaluation/ conclusion |
|---|---|---|---|---|---|---|---|---|
| | **Consensus was achieved on adopting.** | definition was broad enough to include them.<br>Besides the technical definition, the outcome is quite relevant from anyone's perspective. Other than living in the ICU, living without support is a very valuable outcome to have. I would rather be alive without a ventilator than with it. It is a valuable endpoint.<br>Apart from the statistical point of view which I didn't understand, I feel it is very important outcome to have.<br>It was asked if we could add on with some categorization for stable and unstable patients, like patients who require large support, a stable patient who doesn't need inotrope support?<br>It was explained that in this outcome they are trying to club all the support together. Three key types of support - dialysis support, breathing support | without sedation? BK clarified, in the text of outcome, the impact of sedation is mentioned.<br>It is important, and patient centered but how will we measure it? We may have to put clauses like, without sedation.<br>VJ: If we measure something badly, it ceases to be useful outcome, even if we prioritise it. We hope that trial conduct will be standardized, and randomization would take care of some of the variations. We should look at the importance of outcome, keep in mind the challenges of measurement and ensure the standardization of the trial.<br>We are not looking at it from patient's comfort or their family's point of view, we are looking at it from the technicalities of | and again is not only going to affect quality of life but so many other parameters like health economics, people end up spending more due to repeated hospitalizations. We must keep in mind that the outcome should be measurable and be practical, otherwise the value goes down. This outcome makes sense and is important.<br>I also agree that it is an important and must have outcome.<br>**Consensus was achieved on adopting this outcome.** | | | | |

| | Survival | Number of days alive without life support | Number of days alive without coma or delirium | Number of days alive out of hospital | Health-related quality of life | Cognitive function | Additional | Evaluation/ conclusion |
|---|---|---|---|---|---|---|---|---|
| | | and blood pressure support. If a new treatment is being tested, does it keep you free of all these supports?<br>Needed clarification if we need to consider all three supports? In that case, it is going to be a bit tough. Others had similar concerns because someone may require renal therapy for longer time and clubbing them with someone on respiratory support. Second how do you account for someone who is on chronic kidney dialysis support? Will they be considered being on organ support? As an outcome is it good, no problem, but the nitty-gritty have to be sorted. To counter Urvi's concern, I suppose randomization will address these choices.<br>We have lapsed into how to measure the outcomes which is different from the fact that whether we think the outcome is important or not. I | measurements which we clearly need to ignore. Someone who doesn't need sedation and is not delirious is obviously recovering and that is a good indicator of the success of the treatment. (Probably sedation was given more because the treatment was ineffective). So it is a good outcome to have. I do have reservations about the definitions and appropriateness of measurements of each outcome, but I am staying out of it completely and trying to understand if it's a valuable endpoint. We need to look at the outcomes from this perspective. **Consensus was achieved for adopting this outcome.** | | | | | |

| | Survival | Number of days alive without life support | Number of days alive without coma or delirium | Number of days alive out of hospital | Health-related quality of life | Cognitive function | Additional | Evaluation/ conclusion |
|---|---|---|---|---|---|---|---|---|
| | | suppose we did agree that the outcome is important and that there are some challenges and subjectivity in measuring it as it is not a hard outcome to some extent, and we hope that randomization will take care of those uncertain variables. It was concluded that all agree that this is an important outcome and there are no disagreements on the outcome itself; of course there are few challenges in measuring it. **Consensus was achieved on adopting.** | | | | | | |
| **India (Mumbai)** | Both patients did not value survival if it was on life support Physiotherapist and Occupational Therapist said survival without quality of life had no meaning For the doctors and researchers and family, this was the most important outcome | All agreed that this was important | Patients and nurses valued this the most | Only the 2 patients felt this was most important Family views- How does it matter where you are, if you can't eat, walk, or have cognitive dysfunction. Difficult to manage for the family if out of hospital, with no | Physiotherapist, Occupational therapist, and nurses valued this the most | Patients and relatives valued cognitive function the most Physiotherapist, Occupational therapist, and nurses valued this the most Doctors felt it was important, but not most important | | |

| | Survival | Number of days alive without life support | Number of days alive without coma or delirium | Number of days alive out of hospital | Health-related quality of life | Cognitive function | Additional | Evaluation/ conclusion |
|---|---|---|---|---|---|---|---|---|
| | | | | health care support (paid from pocket mostly in India) | | | | |
| **Lithuania** | Discussion on this point started with defining the difference of ICU survival and hospital survival. Small proportion of participants felt that ICU survival per se is of limited value and focus should be shifted to hospital survival. Since clinical studies often focus on entirety of clinical episode, thus if positive outcome in ICU is not followed by positive hospital, the initial success is of limited value to patients and research. Proponents of ICU survival as a core outcome listed five key points for consideration: 1.It allows to focus on processes, treatments and interventions carried out on ICU, thus may be of relevance to mixed general ICU patient population, when clinical trials often study general approaches (for | Panel debated definition provided in the COS presentation. Discussion focused on whether using this as a definition of disease severity is appropriate or whether definition should define this outcome more in terms of recovery (or completeness of recovery) from critical illness. This aspect is also supported by the middle section of definition (comparing this to such outcomes as time in hospital or ICU readmissions). Panel agreed that initiation and discontinuation of organ support is likely to be a lot less variable, than other outcomes. Panel suggested that additional standard explanation regarding ambulatory organ support should be included with definition of this | Debating this as a composite outcome caused the widest variation in opinions and some difficulty in reaching consensus. Panel agreed that both coma and delirium reflect CNS failure and its severity, but somewhat struggled with the concept of combining both in a single outcome. There was a consensus that both contribute to increased hospital length of stay and worsening of other key outcomes. There was an opinion from nursing and patient representatives, that both coma and delirium result in significantly increased care requirements and may have significant negative impact on carers, | Panel was unanimous in adopting this as one the most important outcomes. There was only limited discussion regarding this outcome due to consensus. Further discussion focused on benefits of reporting as many of the core outcomes as possible as this would provide a more complete picture of patient outcomes. | Panel agreed that this is critically important outcome and unanimously supported its adoption. Discussion followed on methodological aspects. As there are several scoring and assessment systems, trials may use different ones, which would make it more difficult to compare results, even though the outcome may appear the same. Panel felt that it would be preferable to have recommendations on which scoring systems should be used (for example SF-36 for general population, Glasgow Outcome Scale Extended for patients with neurological injury). Panel acknowledged that traditional assessment of HRQoL | Panel agreed that this outcome is critically important and has clear links to HRQoL. However, there were multiple uncertainties regarding methodology, consistency, tools, time-points. Acceptable degree of cognitive decline may be dependent on multiple factors, including patient preferences, family, social and system-wide resources. Healthcare and social care systems with more extensive resources may accept different levels of acceptable cognitive decline in comparison to those where long-term care is predominantly depended on family members, privately funded or private | | |

| | Survival | Number of days alive without life support | Number of days alive without coma or delirium | Number of days alive out of hospital | Health-related quality of life | Cognitive function | Additional | Evaluation/ conclusion |
|---|---|---|---|---|---|---|---|---|
| | example fluid balance, sedation, ventilation, etc.), rather than treatments aimed at specific pathology.<br>2.Patient representatives suggested that even limited survival after ICU may be of benefit to the patient, especially if cognitive function is preserved. Some examples included terminal conditions diagnosed during the same ICU episode and survival after ICU gave patients and their families much needed time to come to terms with the diagnosis and poor short-term outcome.<br>3.While ICU care is likely to have some variation, but the range of this variation is likely to be relatively minor in majority of European countries (possibly to a comparable range of the outcome variances amongst ICUs in the same country). However, there is likely to be | outcome. Specific example is intermittent chronic dialysis. In cases of irrecoverable renal failure after critical illness, there is a time point when dialysis dependence is established, and patients transition to ambulatory renal replacement. After this time point dialysis reflects chronic health condition and is no longer representative of acute disease severity.<br>This led to the next discussion point regarding completeness of recover. Sever panel members felt very strongly about importance of measuring impact of our interventions on the likelihood of organ recovery.<br>Panel agreed that this could be primary (or key secondary outcome) in selected trials investigating new treatments and interventions.<br>There was some discussion about ambulatory | especially family members who look after their relatives with significant neurological disability.<br>There was some disagreement on the value and impact of this core outcome on decision making. Some felt that duration of coma and delirium is an important factor in their decision making, but there were those who made very clear distinction between the two stating that prediction of coma or delirium would have very different weight in interpretation of research results and decision making.<br>Overall it was agreed that days without coma or delirium could be used as a surrogate marker for survival with good neurological function. There was an absolute consensus, that | | (patient/family interviews at fixed time points) is very resource intense, therefore concerns were raised regarding feasibility of testing this in majority of clinical trials.<br>Panel also commented on the fact that due to nature of critical illness, background information of HRQoL is seldom available, therefore it is hardly ever feasible to make before and after comparisons of HRQoL. Some opinions were expressed on the use of secondary healthcare data (for example prolonged need of care, repeated hospital admissions, new diagnoses or new regular treatments after ICU admission, or similar indicators) could be used as surrogate indicators of | insurance funded long-term care. In many cases absence of close relatives may be deciding factor. Therefore, major variations are likely even amongst the EU member states, even more so worldwide, thus limiting general applicability of this outcome. Similarly to HRQoL, issues were raised in regard to lack of baseline information and limited scope of before and after comparison and also resources required to measure this outcome in clinical trials.<br>Panel felt that wider adoption of this core outcome would depend on recommended tools, simplicity of use and applicability in variety of systems/countries with varying long-term care arrangements. | | |

| | Survival | Number of days alive without life support | Number of days alive without coma or delirium | Number of days alive out of hospital | Health-related quality of life | Cognitive function | Additional | Evaluation/ conclusion |
|---|---|---|---|---|---|---|---|---|
| | significant variation in post-ICU care, especially with regards to provision of nursing care (nurse to patient ratio), rehabilitation, physiotherapy, speech and language, psychology, etc. Therefore, ICU mortality is likely to be a more standardised and comparable outcome, then hospital mortality.<br>4.Follow-up from point 2 was raised by several panel members - intensivists may have limited scope of initiating significant change in post-ICU care, thus if clinical trial protocols use hospital mortality as a primary outcome, there may be increasing pressure to standardise not only ICU, but post-ICU management as well. This may reduce the number of units participating in multi-centre trials.<br>5.It was stated that hospital mortality following ICU | respiratory support in select conditions (end-stage motor-neurone disease) and ambulatory cardiovascuolar support (ventricular assist devices) but it was felt that these issues are less likely be the subject of general ICU research.<br><br>Overall panel recommended to adopt this outcome. | could be one of the most important outcomes. This position was supported by all participants. However, there is a challenge to this approach, as this definition does not cover patients with debilitating neurological injury, but who are neither in coma nor delirium. For example, patients with persistent GCS score of 12-14 (spont eye opening, obey simple commands, unable to communicate, respond). While this is not a large cohort, it is not insignificant. While days alive with favourable neurological condition was mentioned as a better option, panel did acknowledge that standardising such definition is more difficult and may be prone to various biases. Panel agreed that | | HRQoL. While these indicators lack fidelity provided by patient interviews, such data can be collated from already existing sources (health and social insurance or similar governmental bodies), thus significantly reducing resources needed and could be more uniformly applied. Panel also acknowledged, that there may be significant legislative variations in EU member states (and even more so worldwide), thus reducing its uniform applicability. Panel also discussed subjective versus objective perception of HRQoL. Patient representatives felt that same degree of disability may be perceived as acceptable to | | | |

| | Survival | Number of days alive without life support | Number of days alive without coma or delirium | Number of days alive out of hospital | Health-related quality of life | Cognitive function | Additional | Evaluation/ conclusion |
|---|---|---|---|---|---|---|---|---|
| | survival can fall into two categories – expected and unexpected deaths. Focusing solely on hospital mortality, may reduce our ability to focus on preventable post-ICU deaths and design prevention systems. Panel debated fixed time points for ICU mortality. General consensus was that it should be measured as *de facto* ICU and hospital mortality, irrespective of ICU/Hospital length of stay. Panel was not in favour of arbitrary time points (such as 28-day mortality). Some examples were provided from evidence of cardiothoracic ICU research, that demonstrated significant differences between 30 and 90 day mortality and measurable proportions of those deaths arose from post 30-day, but in-ICU mortality. Fixed time points might be | | incorporation of cognitive function in this outcome would be important, especially in research projects, that do not use COS outcome 6 (cognitive function). | | some and unacceptable to others. Some examples were provided by clinical colleagues on patients who adapted well to disability following critical illness, resulting in acceptable quality of life in-spite of significantly reduced objective HRQoL scores. Panel advocated wider adoption of subjective QoL evaluation tools to capture above mentioned aspects. | | | |

| | Survival | Number of days alive without life support | Number of days alive without coma or delirium | Number of days alive out of hospital | Health-related quality of life | Cognitive function | Additional | Evaluation/ conclusion |
|---|---|---|---|---|---|---|---|---|
| | greater value when measuring long-term outcomes (for example 6, 12 months, 5 years).<br><br>Overall panel recommended to adopt this outcome. | | | | | | | |
| **Norway** | There was broad agreement that survival is a central goal in the treatment of critically ill patients and an important outcome in clinical studies. Everyone agreed that it should be included in the dataset. There was a discussion about when it is most appropriate to define the time of death without the group reaching a conclusion. This will be discussed later. | There was a debate whether “days alive outside the intensive care unit” could replace the proposed elements of organ support treatment, but we concluded that the proposed endpoint is important, necessary, easy to measure, reflects resource use in the intensive care unit, and provides a fairly good insight into patient´s severity of illness. | This can be perceived as a measure of central nervous system dysfunction during the intensive care stay, and input from patients clearly emphasized that delirium is a strongly undesirable experience. It is therefore essential to minimize intensity and duration of delirium if possible. Furthermore, it was pointed out that delirium as a symptom is an important prognostic factor for the mental health in survivors. Thus, there was broad agreement about the importance of delirium as a phenomenon, but | In 4 out of 6 proposed outcomes, survival is part of the endpoint. There was broad agreement that survival was adequately addressed in points 1 and 2. The group therefore saw no significant added value in including this endpoint. On the contrary, there was broad agreement that this endpoint could be removed in favour of other important endpoints that have been omitted from the proposed COS (see below). The group pointed out significant variation in the organization of intensive care units and | The group considered this to be very important endpoint, highly relevant, and essential to include in a COS. Throughout the discussion, it was emphasized very clearly that the endpoint is poorly specified in the Danish proposal, and therefore, some apprehension regarding the further process regarding its precise content, as well as concern that later choice of instruments to measure HRQoL may limit or broaden the scope of this domain. | Similarly, the group unanimously agreed that cognitive function is a very important endpoint. Much time was spent discussing how cognitive function can be measured in two ways (subjectively and objectively), and that results from previous studies may be difficult to compare. Nevertheless, the group wanted to retain this endpoint. | The group believed that the draft had significant shortcomings. Two absent issues were highlighted: physical function and mental health. Both elements represent central symptom domains in the “post-intensive care syndrome” (PICS), and the group considers it a clear weakness in the proposed COS that these have been omitted. Although HRQoL might possibly include both elements, the group agreed that both aspects should be specifically addressed as endpoints in a COS. | The group has conducted a meeting in accordance with the guidelines provided. The group had a good and open discussion about the 6 proposed endpoints. The group agrees with inclusion of endpoints 1, 2, 5, and 6 but rejects endpoints 3 and 4. Furthermore, the group believes that a common dataset should specifically include physical function and mental health and therefore calls for this inclusion. |

| | Survival | Number of days alive without life support | Number of days alive without coma or delirium | Number of days alive out of hospital | Health-related quality of life | Cognitive function | Additional | Evaluation/ conclusion |
|---|---|---|---|---|---|---|---|---|
| | | | the group was critical of including it as a separate endpoint in a COS. The following issues were raised: a) the combination of delirium and coma is artificial as these are two distinct phenomena; b) delirium is a fluctuating mental state and will therefore often be difficult to capture; c) systematic monitoring of delirium is labour-intensive, not performed in all intensive care units, and will therefore be difficult to implement without extraordinary resources. The group was also critical of placing delirium in a context with the number of days alive, as survival is already implemented in other endpoints. | hospitals in different parts of the country, which may affect the validity and consistency of such data. | | | | |
| **Netherlands** | No need for discussion, clear for | There was a discussion because of the choice for | This is a very important outcome because | Outcome of great value. It is important to | All participants agreed that this outcome is very | Very important from the patient's perspective | We discussed a lot of the outcomes that were mentioned in | It worked well that we took our time for the |

| | Survival | Number of days alive without life support | Number of days alive without coma or delirium | Number of days alive out of hospital | Health-related quality of life | Cognitive function | Additional | Evaluation/ conclusion |
|---|---|---|---|---|---|---|---|---|
| | all participating panel members. | 'days alive without life support' instead of days on the ventilator or days with ECMO etc. We discussed that, in the end, you could still calculate the outcome 'days on a ventilator''. However, seen from the patient' perspective it is more important to measure the days without life support instead of days on life support as it is more based on the question; what do you contribute to the patient's life? What really matters (and what the treatment is meant for) is the days alive without support. The participants also mentioned that it is important to determine a cut-off point for each study. | it can affect recovery after the ICU (and may possibly result in long-term complaints) | measure this outcome as days alive and discharged from the hospital because there is a lot of variation in the length of stay among the different hospitals (in the ICU and on the wards as well) | important. We discussed what HRQoL really means and how this can be measured. But also which outcomes are related to HRQoL as it seems to be a collection of outcomes (well-being, daily functioning etc) – more like an umbrella term | because cognitive functioning can have a significant impact on daily life and is not part of the HRQoL-questionnaires | the 19 outcomes list including performing daily activities, well-being, pain and discomfort in the ICU etcetera but agreed that some of these outcomes are less important because they are inconvenient for a short period of time (i.e., discomfort, thirst, pain during ICU-admittance) and also that HRQoL covers a lot of these outcomes including well-being, stress and daily functioning. The panel did not come up with any additional outcomes. | introduction, to get to know each other a little bit. Some participants did not know what to expect so it was really helpful to show them the YouTube video and to take enough time to introduce the benefits of a panel and the COS. We took a lot of time for this first meeting, in a place far from the ICU which also worked well. Other than that, apart from an emergency power test, there were no distractions in the form of phones ringing or people leaving or entering the room. Everyone was also present in civilian clothes, which lowered the threshold between professionals and patients.<br>If I have to mention something that could be improved, it is the emergency power test, but these tests are only planned shortly in |

| | Survival | Number of days alive without life support | Number of days alive without coma or delirium | Number of days alive out of hospital | Health-related quality of life | Cognitive function | Additional | Evaluation/ conclusion |
|---|---|---|---|---|---|---|---|---|
| | | | | | | | | advance and fortunately the power was back after fifteen minutes. Also, the attendees were prepared for the emergency testing by the moderator. |
| **Poland** | The panel members agreed that this was a universal, relevant endpoint that was quite obvious from all points of view. The issue of timepoints was raised | The experts noted that this outcome requires a clear definition. Is it ICU-limited support or long-term support? The problem of end stage renal disease/dialysis was noticed (supporting kidney function in the long term is not as burdening as supporting the respiratory system) and the fact that often discharge from intensive care of a patient who requires respiratory or renal support is possible, unlike supporting the circulatory system | Very important from the point of view of the patient's families.<br><br>The definition of coma and delirium – potentially difficult to define due to its continuous nature (wide spectrum of disorders).<br><br>An56articipt indicator of the quality of care. | Requires defining what does out of hospital mean? How to approach long-term care and rehabilitation facilities?<br><br>Also, the issue of timepoints was raised | Potential difficulty in the assessing the outcome due to lack of "post-ICU" outpatient clinics.<br>In the panel's opinion, a very important outcome, but with numerous limitations – different tools, lots of aspects. Quite complex outcome.<br>A large group of patients with communication problems (aphasia, paresis) – is the caregiver's assessment important then?<br>The issue of the care-givers quality of life was also mentioned at this point. | As in the point above, possible difficulties in the assessment were mentioned, depending somewhat on the time point at which the assessment will be made. The moment of discharge from the ICU, the moment of discharge from the hospital – less difficulties. Distant time point – is it possible and reliable to remotely assess cognitive functions (e.g. by telephone?)<br>It seems necessary to use the same (universal?) tools to make this outcome objective and comparable | | It seems that each panel member understood the purpose of the meeting and was able to present their point of view on specific aspects. The opinions complemented each other and it is indeed very important that people from various interested groups participated in such a panel.<br><br>It was mentioned in the presentation that "*Frequency and tool will be discussed in another meeting, not here. Definitions are provided in the notes*" but we found it not sufficient for our discussion. Different panel |

| | Survival | Number of days alive without life support | Number of days alive without coma or delirium | Number of days alive out of hospital | Health-related quality of life | Cognitive function | Additional | Evaluation/ conclusion |
|---|---|---|---|---|---|---|---|---|
| | | | | | | | | members understood some outcomes differently, in their own style, and the lack of clear definitions or the time and circumstances in which they would be assessed made discussions somewhat difficult, especially regarding two last outcomes.<br><br>To sum up, the meeting went smoothly and the discussions were interesting and developing. For the future, we think it would be useful if at least one of the panel members had been involved in an earlier stage of COS creation or had some type of training/mentoring from the previous stakeholders. |
| **Sweden** | All participants agreed on that survival should be included in a core outcome set. One patient emphasized that "survival is important, but one | There was a discussion regarding the definition of life support " is for example intermittent dialysis and high flow oxygen included?" | The ICU survivors (former patients) firmly stated that they wished that duration of delirium was as short as possible, since it may save | This might be important from a financial perspective but not from a patient perspective. It is an outcome that | All participants agreed on that this was an important outcome. However, it was highlighted that the measurement should preferably | The57articipantts stated that cognitive function is important to be able to manage daily life, to work, and is a part of one's personality. | The 57articipantts suggested adding psychological function (51) to the core outcome set. One patient stated that you may have good HRQL even | The joint discussion was audio recorded, which provided us an opportunity to return to the discussion in case there were |

| | Survival | Number of days alive without life support | Number of days alive without coma or delirium | Number of days alive out of hospital | Health-related quality of life | Cognitive function | Additional | Evaluation/ conclusion |
|---|---|---|---|---|---|---|---|---|
| | does not want to end up as a vegetable". | Circulatory support is not as burdensome for the patient as respiratory support might be. Yet, circulatory support is essential for survival. In the end it was concluded that all days without life support could potentially reduce the risk of complications and time of recovery. There might be a difference in duration of life support because of cultural differences. In Sweden it is common to terminate life support in desolate cases, which is not the case in all countries. | patients (and family) the trauma of being exposed to hallucinations, worry and anxiety, which potentially can mitigate the risk of PTSD. They stated that losing "mental anchor to reality" was by far the worst experience. It was perceived as positive to include delirium as it will lead to more focus on the evaluation of delirium. "Dare to ask" about delirium was highlighted. One patient shared that his hallucinations fluctuated and remained even after discharge from hospital. However, delirium may be difficult to measure, as it varies over time. Being in coma may sometimes be inevitable and was therefore not necessarily considered to be an adverse outcome. There was a discussion regarding whether | depends on the resources in the health system rather than being a patient-centered outcome. All patients expressed that they had been discharged from hospital too early due to lack of resources. Also, at home patients may receive extensive support both from health care and from next of kin. Being in an acceptable condition (good shape) and able to manage daily activities independently, were more important to the patients compared to an (too) early discharge. Plus, that many other factors, except recovery, do influence the time point of discharge. We were hesitant to adopt this outcome and spend a lot of time on this | be adapted to intensive care patients (and not to a general population) and such questionnaire should have a distinct differentiation between physical and psychological health. We also discussed if the physical function should also be estimated by objective measures or whether self-rated function is sufficient. The patients stated that psychological well being should be emphasized more than in existing HRQL questionnaires, as it is very important for their recovery. | One patient expressed that cognitive recovery is indicative of returning to the person you were before the illness or injury. It was also questioned whether mental function and cognitive function are equivalent. | though you are physically disabled, but it is impossible if you have detrimental psychological symptoms. An argument to add this was that HRQL is self-reported, and that there are many objective ways of measuring psychological function that might be overlooked. | uncertainties about differences of opinions. We believe it was important to highlight that other outcomes were not less important in clinical practice and these outcomes were related to randomized controlled studies only. Providing concrete examples of what can be evaluated in RCTs also helped to guide the discussion. |

| | Survival | Number of days alive without life support | Number of days alive without coma or delirium | Number of days alive out of hospital | Health-related quality of life | Cognitive function | Additional | Evaluation/ conclusion |
|---|---|---|---|---|---|---|---|---|
| | | | coma should be a key core outcome variable (or not), and that there is a difference between illness induced coma and medically induced sedation. One patient expressed that he believed that his hallucinations were due to the sedative drugs he received, and that he would never accept being sedated again. | discussion and returned to the question several times. After a lengthy discussion, we agreed that we will adopt the outcome as it might be important to measure this outcome in Sweden where health care resources are sparse. We suggest to clarify if "Days alive out of hospital" includes discharge to rehabilitation centres, and nursing homes or only applies to home. | | | | |
| Switzerland | Patients:<br>very important outcome, however it is also very important what comes after, survival is not the only goal, meaning full living is key<br><br>Relatives:<br>the first step, most important, primary aim and focus during the first phase of treatment. Relatives | Nurses:<br>Patients without these support measures are usually not on the ICU anymore<br><br>Inverse relationship between devices/types of devices and relatives' distress is well proven in the literature<br><br>Physicians: | Patients:<br>Important outcome<br>Very powerful and bad "dreams", thought to be real, even thereafter for a time it was not clear what was real and what was not<br>Impact on personal well-being huge<br>Nurses: | Adjustment:<br>" Days alive and return to previous living situation"<br>Importance to go "home" (wherever or whatever this is)<br><br>"Home is where the heart is" – meaning that it is one of the primary aims of ICU care to | (discussed after survival because of the question when survival is meaningful)<br>Patients:<br>Very important outcome, time points difficult to define as undulating wave. Psychological quality of life more important than physical. Professional post- | Physician:<br>More objectively measurable<br><br>Coping with cognitive dysfunction is very individual and also the level of dysfunction that is acceptable to patients<br><br>Relatives: | Mobility: should it be assessed separately.<br>Independency/self-efficacy in ADL:<br>Being able to be self-sufficient or be care independent is a key ability/wish for all groups and runs very strongly in Swiss society<br><br>Difficult to assess and very individual regarding what is | The meeting went very well, and the atmosphere was good and productive. Everybody agreed to be available for further meetings to discuss research matters in the future. One challenge was that to give everybody sufficient "space" to voice opinions the session was a |

| | Survival | Number of days alive without life support | Number of days alive without coma or delirium | Number of days alive out of hospital | Health-related quality of life | Cognitive function | Additional | Evaluation/ conclusion |
|---|---|---|---|---|---|---|---|---|
| | often are not aware what the outcome with survival may entail on the long run (i.e., disability, length of the recovery process etc)<br><br><u>Physician/nurses:</u> survival primary aim/focus in the first phase, thereafter, question of "meaning full living" becomes more important. When measured is important<br><br><u>Physiotherapist:</u> survival first, all other considerations come next, potentially a question of when measured (after ICU discharge maybe not, but maybe yes after 1 year)<br><br>Further discussion: *"meaning-full survival"*<br>Patients: difficult to define, many aspects go in to this<br><br>Relatives: entails several aspects, difficulty to support patients, especially with long-term | Indicates the trajectory of recovery and is thus importance<br><br>Also, an indirect measure of effectiveness of treatment<br><br><u>Physiotherapist:</u> Shows how sick a patient is<br><br>May has implications on quality of life and potential for recovery<br><br><u>Relatives:</u><br>Important as it signifies a sort of progress "having less devices/medication". Less dependency. The less invasive the better, thus liberation form organ-support is important<br><br><u>Patients:</u><br>All the devices are not memorable. Took little notice of pharmacological support. The only thing that was memorable was the inability to speak and being taken off | Patients often suffer from very powerful and long episodes of hyperactive delirium<br><br>Strain on nurses/health-care professionals considerable, i.e., hard to watch<br><br>Needs a lot of resources<br><br>Most important variable in this core outcome set<br><br>Important for more targeted resource use (i.e. physiotherapists)<br><br><u>Relatives:</u><br>Very difficult to cope with: A) to see relative so in distress B) being in conflict with patients as patients often think that they are not understood by relatives<br><br>Time to wait until the patient is out of coma is important<br><br><u>Physiotherapist:</u> | enable the patient to return to the pre-ICU living situation<br><br>It is also important to measure how many days are spent in (health) care institutions (rehabilitation, nursing home, home for the elderly) post ICU stay, not only the hospital | ICU care, would help tremendously, as the "normal" doctors do not understand the diseases and their potential long-term consequences (physically, psychologically) Multi-modal assessment (social, financial, psychological, re-integration) necessary<br><br><u>Nurses:</u><br>In combination with survival the most important outcome, if we knew the health-related quality of life resulting of an ICU stay, the discussion about survival would sometime be redundant<br><br>The question is whether relatives are included in this outcome<br><br><u>Relatives:</u><br>Important outcome | Important outcome<br><br>Difficult to cope with for relatives because "disability" is not visible<br><br>Level of cognitive dysfunction is evaluated differently by patients and relatives, difficulty to draw a clear line to health-care related quality of life<br><br>Very subjective as somebody can happy with major cognitive dysfunction, but the relatives may be very distressed by the same level of dysfunction "patient not being him/herself anymore"<br><br><u>Patients:</u><br>Especially micro-cognitive deficits and tiredness are very difficult to cope with as often not accepted by society | acceptable and what not<br><br>Part of the panel feels that this is part of health-care related quality of live, however the panel feels that it should be assessed separately to give the Issue more strength<br><br>outcome should include financial, working and social aspects of life<br><br><u>Family outcomes (functioning, resilience, or mental health (wellbeing, psychological distress):</u><br>More integrative view on outcomes including relatives, care givers and patients (i.e. quality of life, cognitive decline)<br>Take the view of the society in consideration (resource use, costs) | bit longer than anticipated (4 hours) and the patients (both with remaining concentration issues) were very tired at the end. |

| | Survival | Number of days alive without life support | Number of days alive without coma or delirium | Number of days alive out of hospital | Health-related quality of life | Cognitive function | Additional | Evaluation/ conclusion |
|---|---|---|---|---|---|---|---|---|
| | consequences of critical illness (physical, financial, psychological)<br><br>Probably summarized in health-care related quality of life | the respirator for suctioning | Very important<br><br>The earlier the patient is out of coma/delirium the earlier one can start to work with the patients/start rehabilitation<br><br><u>Physician:</u><br>Important outcome, because rehabilitative measures can only start after the patient is out of coma/delirium<br><br>Puts a lot of strain on health-care personnel “mental load”<br><br>Prolongs the duration of treatment and has detrimental effects if prolonged (cognitive decline, muscular waste etc)<br><br>Further discussion: Should coma and delirium be separated?<br>• short discussion | | Emphasis should be on psychological outcomes<br><br>Include the relatives viewpoint, respectively their quality of life<br><br><u>Physician:</u><br>Many different factors<br><br>Doubtful that current assessment tools are inclusive enough as focused on physical problems, the instruments should include psychological, financial, social and well-being factors<br><br><u>Physiotherapist:</u><br>Very important outcome measured factors should be tailored to what is important to patients e.g., driving a car is not assessed in any tool.<br>**Remark:**<br>New tools are necessary | Very difficult as also very much is dependent on the situation, there are situations (a lot of noise, tiredness) where cognitive function is worse than in other situations, this needs to be incorporated in the measurement of this outcome<br><br>The “how” it is measured is difficult<br><br><u>Nurses:</u><br>Important outcome<br><br>Subjective and objective aspects of quality of life, not only health related quality of life<br><br>Important also for regulatory purposes/social security (i.e., to get financial support from the state)<br><br><u>Physiotherapist:</u><br>Important outcome | | |

| | Survival | Number of days alive without life support | Number of days alive without coma or delirium | Number of days alive out of hospital | Health-related quality of life | Cognitive function | Additional | Evaluation/ conclusion |
|---|---|---|---|---|---|---|---|---|
| | | | • both sort of in a continuum<br><br>panel feels that both put the same strain on all groups and should be evaluated together | | Multi-modal assessment (social, financial, psychological, re-integration) necessary<br>Professional post-ICU care necessary for standardization<br>Include the relative's viewpoint or their quality of life | HRQoL is more important than cognitive, there is a lot of common ground, should we only measure the former? However we feel that it should be included in the HRQoL category if new multi-modal assessment tools are developed that include social, financial, psychological, re-integration etc | | |
| UK Cardiff, Wales | Essential outcome – if you are not recording survival how can any of the others be examined – ICM Consultant, Relative, Physiotherapist and SALT.<br>"This is key. Once I was aware of my situation all I was focused on was surviving and getting out of intensive care "– Previous patient.<br>"Survival must be measured or how will you know whether treatments for patients are working "– Current patient. | "I feel this is massively important. The side effects, and the physical and mental impacts that come with needing each machine or treatment are huge and can really affect how you cope each day when you are a patient. This outcome should definitely be looked at and measured because its' really important to patients" – Current patient.<br>"Each time I had to go back on the ventilator, or needed dialysis again, it was such a blow to me | "I feel this is massively important. The side effects, and the physical and mental impacts that come with needing each machine or treatment are huge and can really affect how you cope each day when you are a patient. This outcome should definitely be looked at and measured because its' really important to patients" – Current patient. | "ICU Consultants should know if a patient is readmitted back to hospital, even if they don't need to come back to the ICU, because if the patient survives ICU but they are constantly in and out of hospital and their health is terrible, is it worth it?" – Relative<br>"This is important to measure – if a patient survives ICU but has chronic health problems should this be | Generated the most amount of discussion. Previous and current patients confirmed experience of each of the 6 outcomes described on the slide and felt very strongly that these were vitally important to be measured. All shared experiences along with the two relatives about things that had affected their HRQOL both in and out of the ICU. Putting up the slide that showed | Also generated a lot of discussion. All parties felt that this was a vitally important outcome to measure and that regaining cognitive function as quickly as possible was vital to patients and their relatives wellbeing and quality of life.<br>"The temporary disruption to my cognitive function while in the ICU I can deal with now I'm past it. But had I not been able to recover my cognitive function sufficiently to be | | We felt it went very well and had positive feedback from all members of the panel.<br>We were lucky to have a very good panel from a variety of backgrounds but all with experience of Intensive Care. They were all talkative and happy to contribute which generated lots of discussion. The informal setting helped to facilitate this.<br>We can't think of anything that didn't work well |

| | Survival | Number of days alive without life support | Number of days alive without coma or delirium | Number of days alive out of hospital | Health-related quality of life | Cognitive function | Additional | Evaluation/ conclusion |
|---|---|---|---|---|---|---|---|---|
| | "If we can survive the ICU admission, we've at least got a shot at further recovery once we leave the ICU, so initial survival must be recorded, the difficulties afterwards can be captured in other research" – Previous patient.<br>Dates for measuring survival are very important and should be specified in the COS. FICM recommend that 1 year should be the maximum time frame for recording survival – Psychologist, Physiotherapist. | and my family. But each time I came off the ventilator or the dialysis machine I felt like I was doing better and this pushed me on to survive and try my best to get better: - Previous patient.<br>"When he had to go back on the ventilator or kept needing the drugs to keep his blood pressure up, I wanted to know if this meant he was never going to get better. I couldn't get any information on the internet or from asking others and so if this was something that was measured in this COS, I may have been able to find that information out myself – its important information to have and should be measured" – Relative.<br>Very important to measure these parameters because it helps us with treatment plans and predicting trajectory of illness – ICU Consultants, | "Each time I had to go back on the ventilator, or needed dialysis again, it was such a blow to me and my family. But each time I came off the ventilator or the dialysis machine I felt like I was doing better and this pushed me on to survive and try my best to get better: - Previous patient.<br>"When he had to go back on the ventilator or kept needing the drugs to keep his blood pressure up, I wanted to know if this meant he was never going to get better. I couldn't get any information on the internet or from asking others and so if this was something that was measured in this COS, I may have been able to find that information out myself – its important information to have and should | considered a bad outcome and should we be considering the socio-economic burden on the patient, their family and society?" – ICU Consultant<br>The Physiotherapist and the Psychologist felt that the number of days and number of occasions admitted back into hospital should be measured separately. | the 6 outcomes related to the well-being helped direct the discussion.<br><br>All parties agreed that this was the most important of all the 6 COS they were being asked to discuss.<br>Key statements were "This is the purpose, getting patients a decent quality of life both in and out of the ICU should be the aim everyone is working towards"<br>"It's about a life **saved** vs a life **lived"** | able to return to my life before I was admitted to the ICU, I can't say in all honesty that I would have wanted to survive, especially if I had been left with a significant cognitive deficit" – Previous patient.<br>All the healthcare professionals discussed how objectively reporting on cognitive function could provide evidence for providing more resources towards helping regain maximum cognitive function, e.g. preventing delirium, giving treatments that do the least amount of damage to cognitive function, providing more therapy involvement while in ICU from SALT, Physio and OT. | | and when we asked some members of the panel if there was anything they thought could have gone better or could be added/improved no suggestions were given, all gave positive feedback from their experience of participating. |

| | Survival | Number of days alive without life support | Number of days alive without coma or delirium | Number of days alive out of hospital | Health-related quality of life | Cognitive function | Additional | Evaluation/ conclusion |
|---|---|---|---|---|---|---|---|---|
| | | Physiotherapist, SALT | be measured" – Relative.<br>Very important to measure these parameters because it helps us with treatment plans and predicting trajectory of illness – ICU Consultants, Physiotherapist, SALT | | | | | |
| UK London, England | | | | | Health related quality of life should include "mental well-being"; if it is not included, it should be added as a separate core outcome | | | Consensus:<br>Quality of life and functional independence after ICU were considered very important; for some patients, this was more important than survival<br>Burden and impact on family after ICU is important but also **during** ICU stay<br>Not considered to be very important:<br>- duration of mechanical ventilation - duration of pharmacological cardiovascular support<br>-health economics<br><br>Some panel members were |

| | Survival | Number of days alive without life support | Number of days alive without coma or delirium | Number of days alive out of hospital | Health-related quality of life | Cognitive function | Additional | Evaluation/ conclusion |
|---|---|---|---|---|---|---|---|---|
| | | | | | | | | surprised to see that "mental health" was not included in the top 19 outcomes<br><br>**Primary Core Outcome set**<br>Consensus:<br>agreement with key outcomes<br>Quality of life very important<br>Quality of life more important than days alive without organ support<br>Some panel members felt that "mental well-being" should be included if possible<br><br>**Overall conclusion:**<br><br>Adoption of all core outcomes apart from "days alive without organ support" (patients felt that they were asleep when they were on mechanical ventilation and therefore not aware of the exact duration of organ support) |

#### Table S6. Distribution of stakeholders registered for the Delphi survey

| Stakeholders | Registered for the survey | Respondents round 1[b] | Respondents round 2[b,c] | Respondents round 3[b,c] |
|---|---|---|---|---|
| | 264 | 249 (94%) | 202 (81%) | 203 (82%) |
| **Patients** | 65 | 61 | 43 | 48 |
| **Family members** | 49 | 46 | 35 | 34 |
| **Clinicians** | 136 | 129 | 110 | 107 |
| **Researchers[a]** | 14 | 13 | 14 | 14 |

[a] Researchers conducting research in the ICU setting or in adjacent populations of critically ill patients.

[b] Respondents completing the survey.

[c] For round 2 there were 208 respondents, but 6 without completing the survey, and for round 3 there were 207 respondents, and 4 not completing the survey. Due to less than 20% attrition between round 1 and 2, as well as round 1 and 3 we present the complete cases with last observation carried forward as planned [1].

#### Table S7. Outcomes derived from the semi-structured interviews

- **How important is it to record new diagnoses after ICU admission?**
- **How important is it to measure hope?**
- **How important is it to record balance as an ability?**
- **How important is it to avoid infections?**
- **Time until returning home**
- **Number of inter- and intra-hospital transfers**
- **How important is it to quickly remove invasive tubes and lines?**
- **Resource utilization for personnel providing intervention/treatment?**
- **Withdrawal of therapy**

#### Table S8. The 50 outcomes in the survey including help-text

These are the questions raised for the 50 outcomes including the help text for the 3rd round.

| | OUTCOME | HELP TEXT |
|---|---|---|
| 1 | How important is it to record ICU survival? | Your response is only regarding ICU survival - not life after ICU or long-term survival. |
| 2 | How important is it to record the ICU length of stay? | Your response is only regarding the importance of recording number of days in ICU. |
| 3 | How important is it to record the hospital length of stay? | Your response is only regarding the importance of recording the number of days in hospital. |
| 4 | How important is it to record the ICU readmission rate during a hospital stay? | Your response is only regarding the importance of recording ICU readmission - not the reason why. |
| 5 | How important is it to record hospital readmission? | Your response is only regarding the importance of recording hospital readmission - not the reason why. |
| 6 | How important is it to record patients' overall well-being? | Well-being is a combination of feeling good and functioning well, including the feeling of safety, harmony, joy, and satisfaction, as well as having positive, meaningful social relationships. |
| 7 | How important is it to measure the health economic consequences of a particular treatment in ICU? | Please consider the importance of recording how society gets the best value for the money. |
| 8 | How important is it to measure the environmental consequences of a particular treatment in ICU? | Please consider the importance of recording the environmental consequences of a treatment, e.g., in the form of the $CO_2$ footprint. |
| 9 | How important is it to measure whether blood values are within normal limits while in ICU? | Please consider the importance of recording whether blood tests taken in ICU are in the expected range. |
| 10 | How important is it to record whether vital signs are within normal limits while in ICU? | Please consider the importance of recording whether e.g., heart rate, blood pressure, temperature, respiratory rate, and oxygen saturation are within normal limits while in ICU. |

| | | |
|---|---|---|
| **11** | How important is it to record the duration of mechanical ventilation while in ICU? | Please consider the importance of recording how long a patient needs artificial respiratory assistance while in ICU. |
| **12** | How important is it to record the duration of pharmacologic circulatory support while in ICU? | Please consider the importance of recording how long a patient needs pharmacologic circulatory support while in ICU. |
| **13** | How important is it to record the duration of dialysis while in ICU? | Please consider the importance of recording how long a patient needs to have artificial blood cleansing by dialysis due to kidney failure while in ICU. |
| **14** | How important is it to record the duration of sedation while in ICU? | Please consider the importance of recording how long a patient needs to be sedated while in ICU. |
| **15** | How important is it to record the duration of delirium while in ICU? | Delirium is an acute confusional state characterized by inattention, incoherent thinking and speech, and altered level of consciousness - many experience not being themselves and that events are not real (hallucinations). |
| **16** | How important is it to record whether patients experience nausea and vomiting while in ICU? | Please consider the importance of recording whether a patient experiences nausea and vomiting while in ICU. |
| **17** | How important is it to record whether patients experience thirst while in ICU? | Please consider the importance of recording whether the patient experiences thirst while in ICU. |
| **18** | How important is it to record whether patients are short of breath while in ICU. | Please consider the importance of recording whether the patient experiences shortness of breath while in ICU. |
| **19** | How important is it to record whether patients experience pain while in ICU? | Please consider the importance of recording whether the patient is pain free while in ICU. |
| **20** | How important is it to record whether patients experience anxiety while in ICU? | Please consider the importance of recording whether the patient experiences fear or anxiety while in ICU. |
| **21** | How important is it to record patients' sleep while in ICU? | Please consider the importance of recording whether the patient gets any sleep while in ICU. |
| **22** | How important is it to record patients' ability to communicate while in ICU? | Please consider the importance of whether the patient is able to exchange information or express meaning by verbal/nonverbal communication or augmentative and alternative communication devices while in ICU. |

| | | |
|---|---|---|
| **23** | How important is it to record patients' discharge location after hospitalization? | Please consider the importance of recording the discharge location, e.g., home, care home, rehabilitation, or other. |
| **24** | How important is it to record patients' outpatient or general practitioner visits following hospitalization? | Please consider the importance of recording the number of follow-up visits conducted as a consequence of undesired effects of intensive care and hospitalization. |
| **25** | How important is it to record whether new medications are prescribed for the patient after ICU discharge? | Please consider the importance of recording whether the patient needs to take new medications as a consequence of the ICU stay? |
| **26** | How important is it to record patients' health-related quality of life after ICU discharge? | Please consider the importance of recording the patient's self-reported life situation from the perspective of goals, expectations, and worries, including affluence, environment, and physical and psychological health. |
| **27** | How important is it to record whether patients experience newly onset tiredness or fatigue, unrelieved by sleep or rest, after ICU discharge? | Please consider the importance of whether the patient experiences tiredness, weakness, or lack of energy, unrelieved by sleep, after ICU discharge? |
| **28** | How important is it to record whether patients experience newly onset pain after ICU discharge? | Please consider the importance of recording the patient's pain after ICU discharge. |
| **29** | How important is it to record whether patients resume their usual roles in daily life after ICU discharge? | The usual role in daily life is the individual's function, characteristics or behaviour in different situations, e.g., in private life, work life, or leisure time, as before the ICU stay. |
| **30** | How important is it to record family caregivers' experienced quality of life after the ICU stay? | Please consider the importance of recording the family caregiver's view of own life situation viewed in relation to the individual's goals, expectations and worries, including affluence, environment, and physical, psychological and mental health. |
| **31** | How important is it to record patients' financial capabilities after, compared to before, the ICU stay? | Please consider the importance of the patient's financial independence after ICU discharge, e.g., by return to work, right to sick leave, or early retirement, or by not needing to take a loan to maintain the standard of living. |
| **32** | How important is it to record whether patients after ICU discharge, compared to before, have the same labour market attachment. | Please consider the importance of recording the patient's labour market attachment after ICU discharge, if the patient was attached to the labour market before. |

| | | |
|---|---|---|
| **33** | How important is it to record patients' physical impediments such as scars, wounds, or ostomy after ICU discharge? | Please consider the importance of recording whether patients have impediments such as scars, or poor wound healing after e.g., pressure sores, “throat tubes” (tracheotomy), or ostomy on the stomach, etc. |
| **34** | How important is it to record whether patients independently perform activities of daily living after ICU discharge? | Please consider the importance of recording the patient’s ability to independently perform activities of daily living, such as personal care, shopping, cooking, driving and other daily chores. |
| **35** | How important is it to record whether patients are independently able to get dressed after ICU discharge? | Please consider the importance of recording whether the patient is independently able to dress after ICU discharge. |
| **36** | How important is it to record whether patients are mobile after ICU discharge? | Please consider the importance of recording whether a patient is independently able to move about after ICU discharge, e.g., get out, drive, bicycle, shop, go for at walk or take the stairs. |
| **37** | How important is it to record whether patients experience newly onset shortness of breath after ICU discharge? | Please consider the importance of recording whether the patient experiences being breathless or short of breath after ICU discharge. |
| **38** | How important is it to record whether patients have newly onset trouble speaking after ICU discharge? | Please consider the importance of recording whether the patient experiences e.g., hoarseness, voice fatigue, sore throat, lump in the throat, need to clear the throat, loss of voice, or continuous cough related to the ICU stay. |
| **39** | How important is it to record whether patients have newly onset difficulty swallowing after ICU discharge? | Please consider the importance of recording whether a patient to a greater or lesser extent has difficulty chewing and swallowing after ICU discharge. Some avoid coarse rye bread and red meat, while others require minced food and thick liquids. |
| **40** | How important is it to record patients' bowel function after ICU discharge? | Bowel function is the patient's ability to maintain normal digestion after ICU discharge and ability to eat normally without nausea and vomiting and have usual stool frequency. |
| **41** | How important is it to record patients' urinary function after ICU discharge? | Please consider the importance of recording whether the patient has normal bladder control without the need for urinary aids and without involuntary urination. |
| **42** | How important is it to record the patient's sexual function after ICU discharge? | Sexual function is the ability to receive and give sexual pleasure. |

| | | |
|---|---|---|
| **43** | How important is it to record patients' senses: vision, hearing, touch, smell, and taste after ICU discharge? | Please consider the importance of recording the patient's ability to se, hear, touch, smell and taste. |
| **44** | How important is it to record whether patients have the physical strength to perform activities of daily living after ICU discharge? | Please consider the importance of recording whether the patient has the strength to perform basic physical needs such as eating, drinking, using the toilet or holding a telephone after ICU discharge. |
| **45** | How important is it to record patients' mental function (cognitive function) after ICU discharge? | Mental function includes concentration, attention, memory, problem solving, learning, sense of direction or spatial orientation. |
| **46** | How important is it to record whether patients experience loneliness after ICU discharge? | Please consider the importance of recording whether the patient feels involuntarily alone (isolated or abandoned) after ICU discharge. |
| **47** | How important is it to record whether patients are diagnosed with posttraumatic stress syndrome (PTSD) after ICU discharge? | PTSD is a diagnosis describing a stress response to a traumatic event where the individual experiences flashbacks and tries to avoid things or situations that are a reminder of the event. Many survivors experience irritability and have trouble sleeping, concentrating or remembering. |
| **48** | How important is it to record whether patients experience anxiety after ICU discharge? | Please consider the importance of recording whether a patient experiences anxiety preventing the performance of normal daily activities. |
| **49** | How important is it to record whether patients experience depression after ICU discharge? | Please consider the importance of recording whether a patient is less happy, lacks joy of life, is more tired than usual, or feels that life is unmanageable. |
| **50** | How important is it to record whether patients experience nightmares after ICU discharge? | Please consider the importance of recording whether a patient experiences unpleasant, scary or anxiety provoking dreams. |

### Figure S1-S50. Graphs for the first survey round

#### Figure S1: How important is survival?

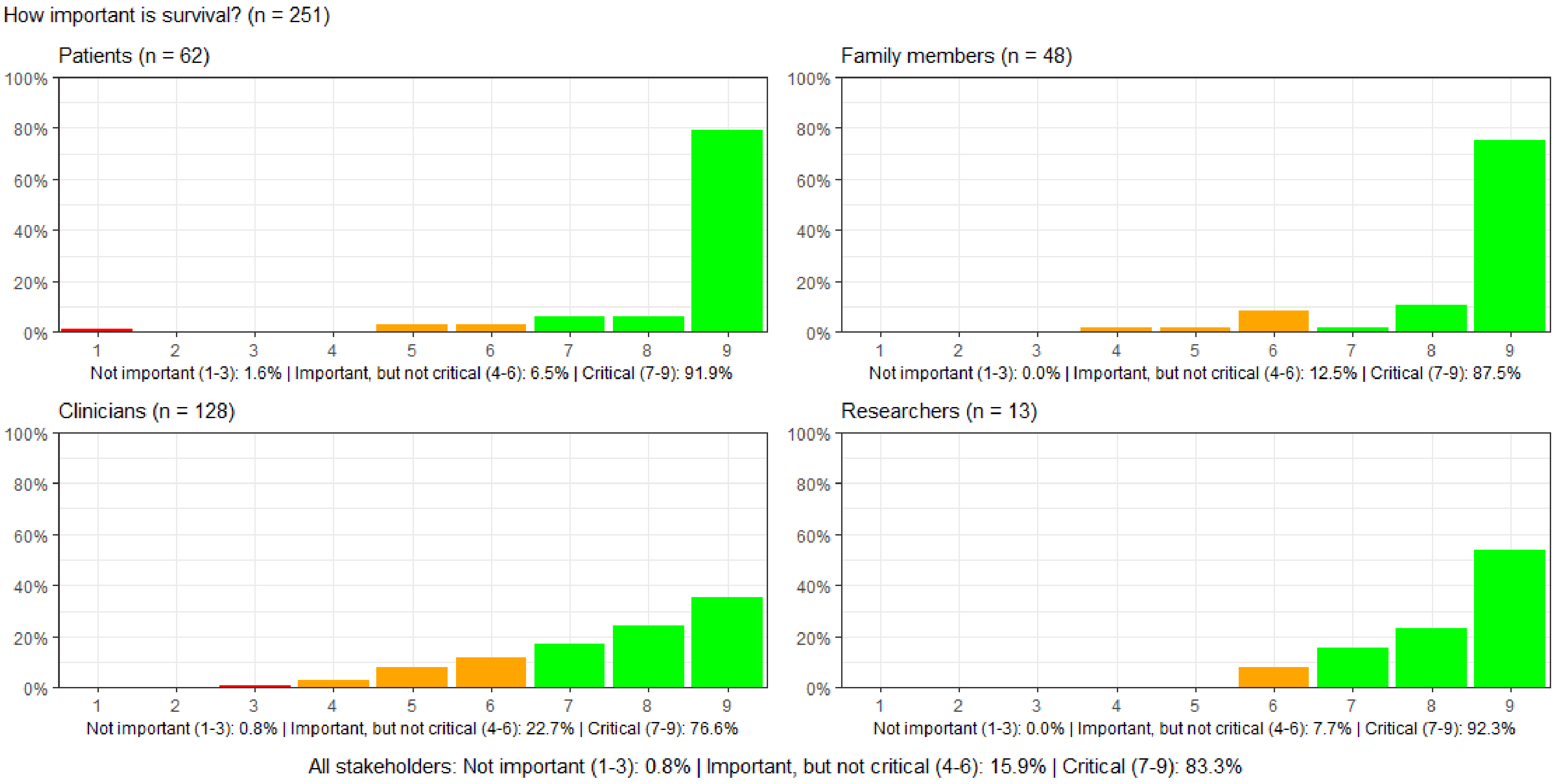

#### Figure S2: How important is the ICU length of stay?

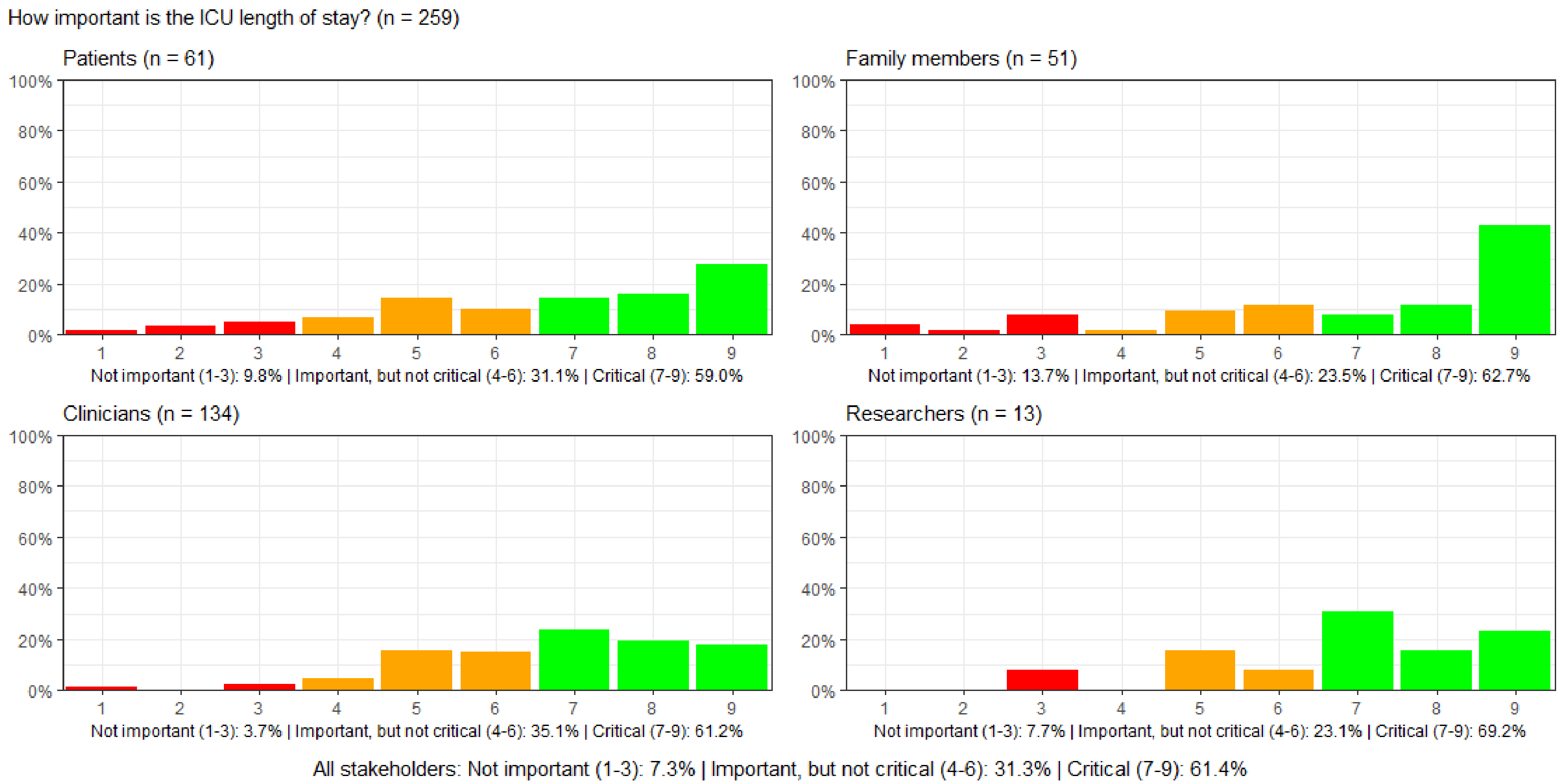

#### Figure S3: How important is the hospital length of stay?

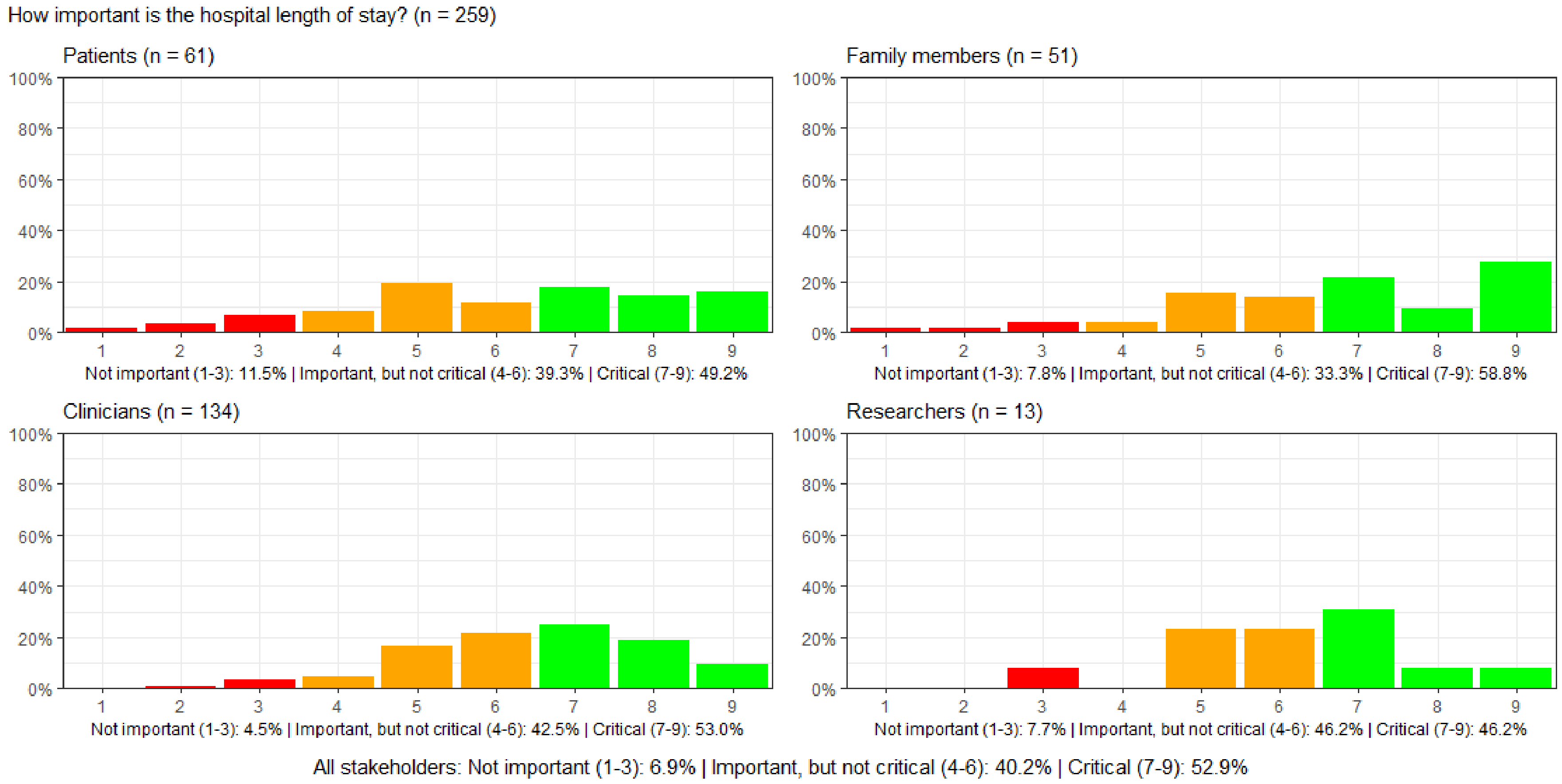

#### Figure S4: How important is it whether one is readmitted to ICU?

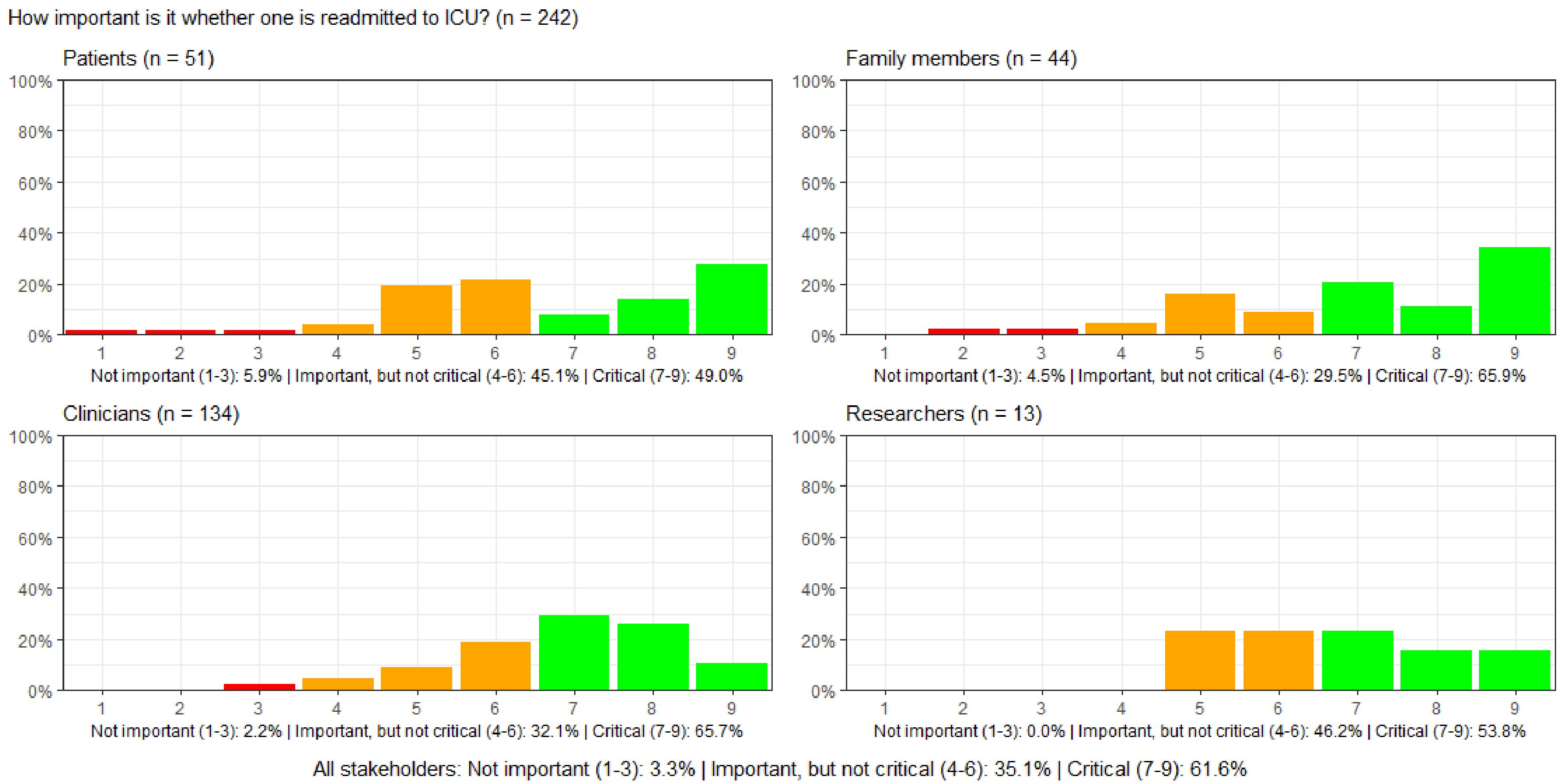

#### Figure S5: How important is it whether one is readmitted to the hospital?

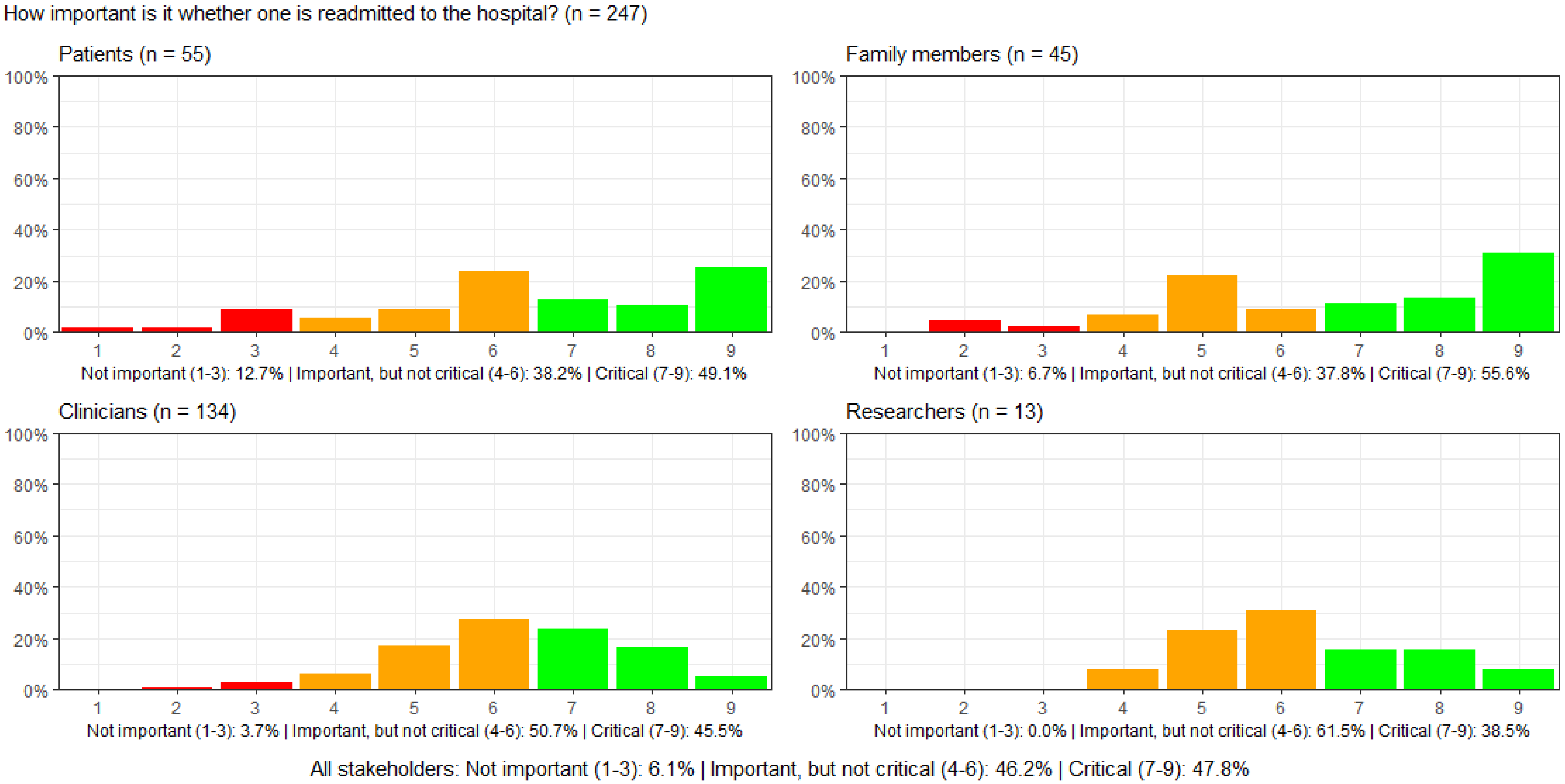

#### Figure S6: How important is it to have an overall sense of well-being?

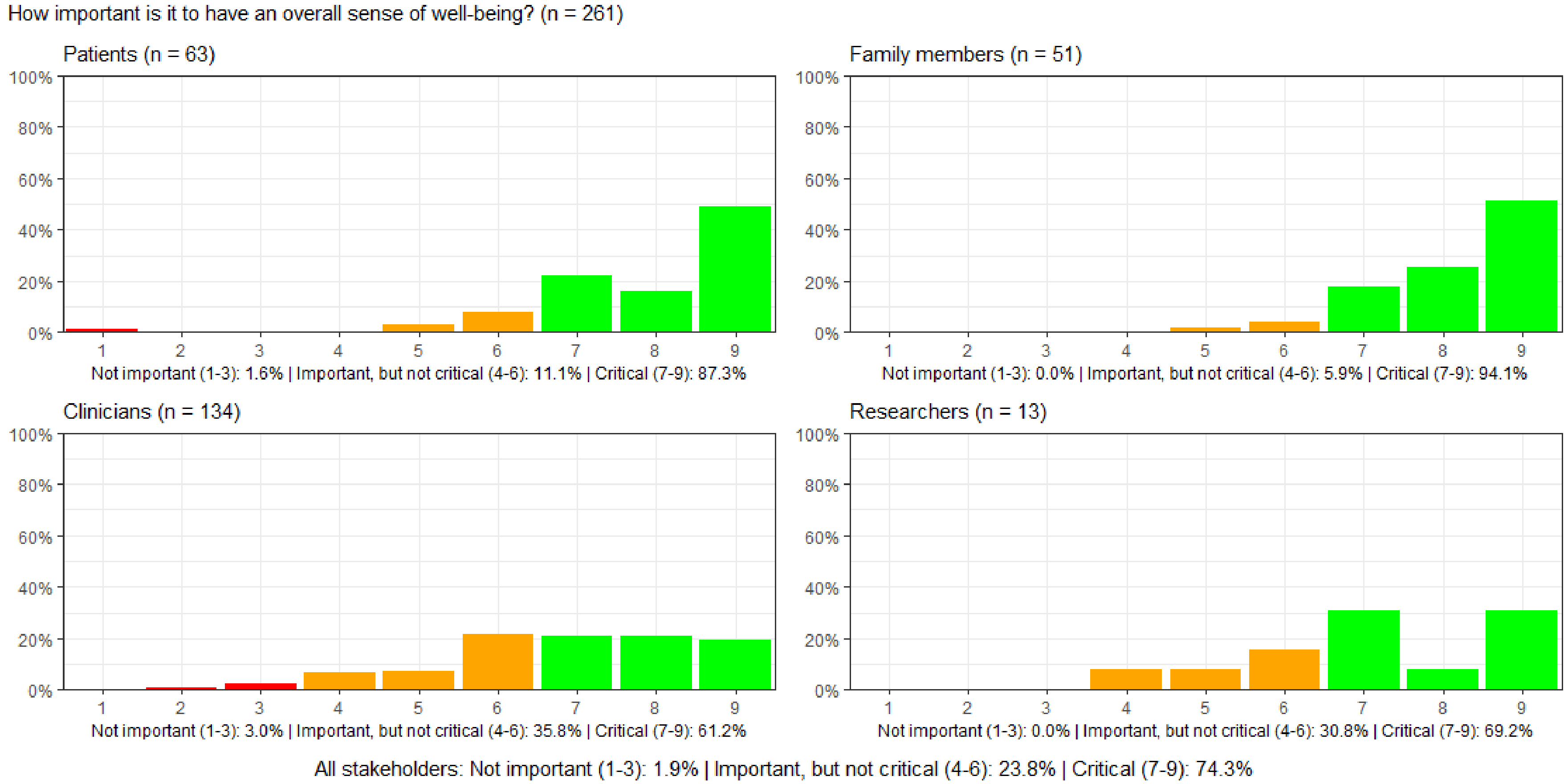

#### Figure S7: How important is a health economic measurement of the specific treatment in relation to the outcome?

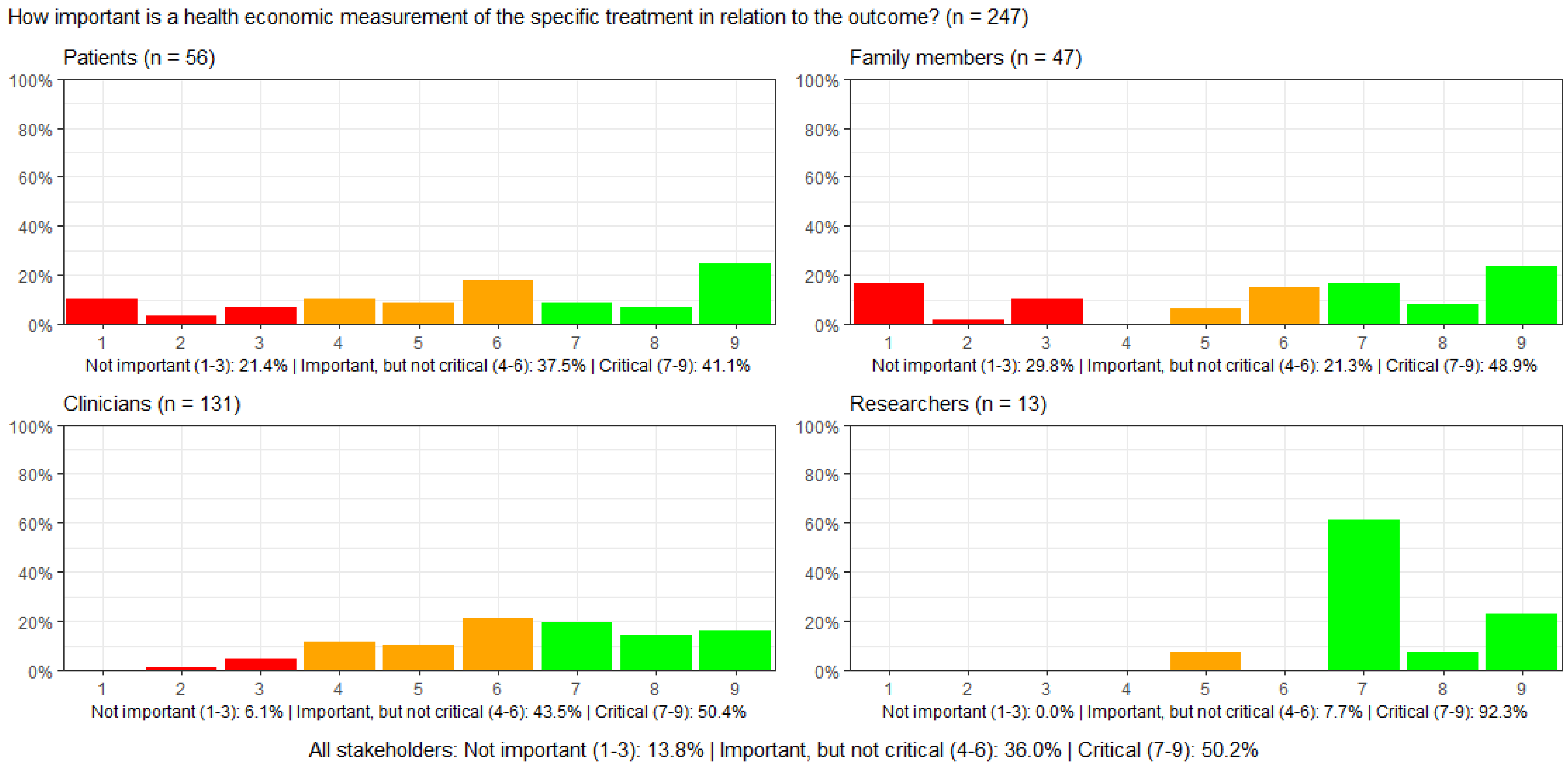

#### Figure S8: How important is the environmental consequences of a particular treatment in ICU?

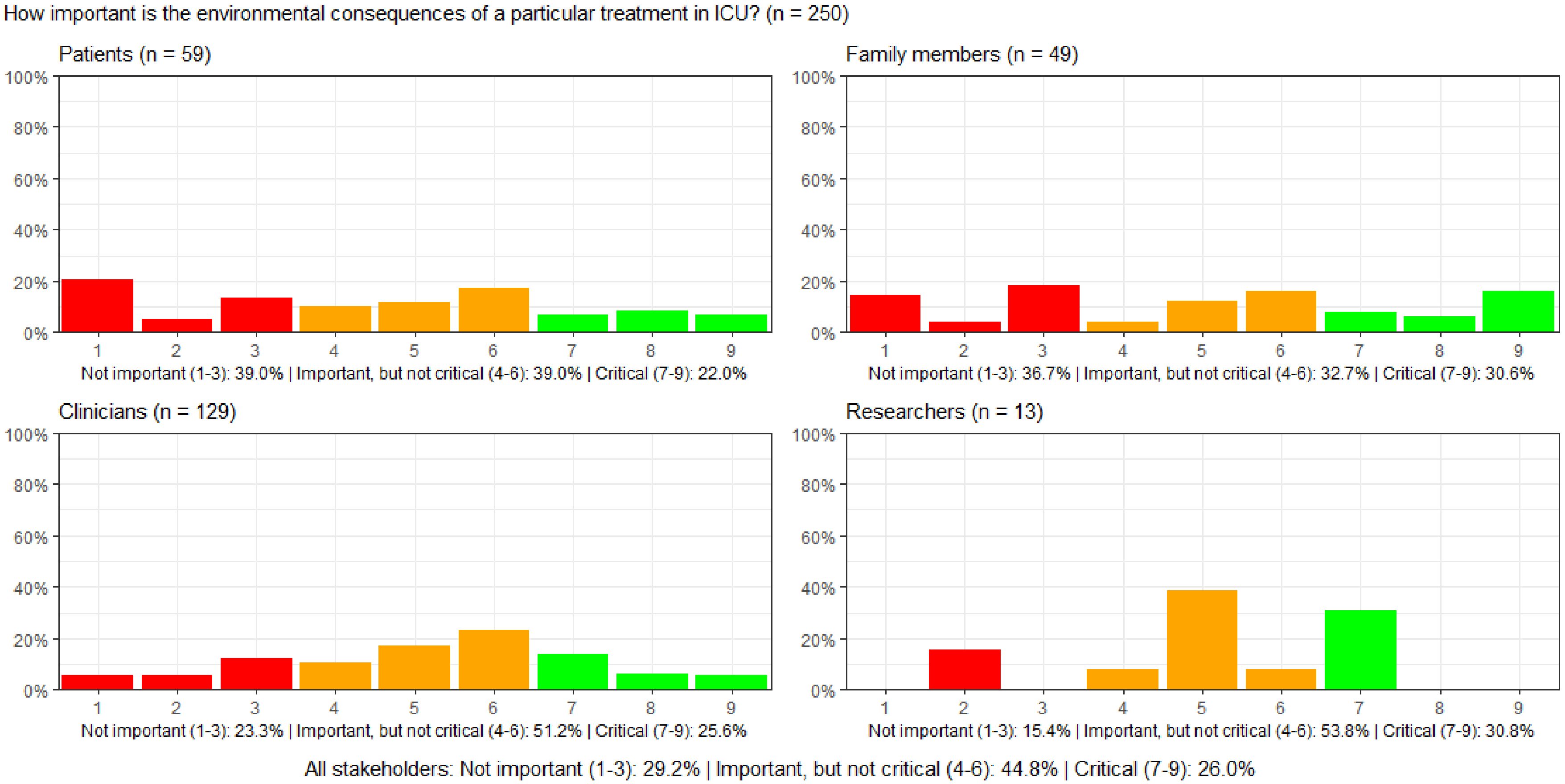

#### Figure S9: How important is it for blood test values to be within the normal limits during the ICU stay?

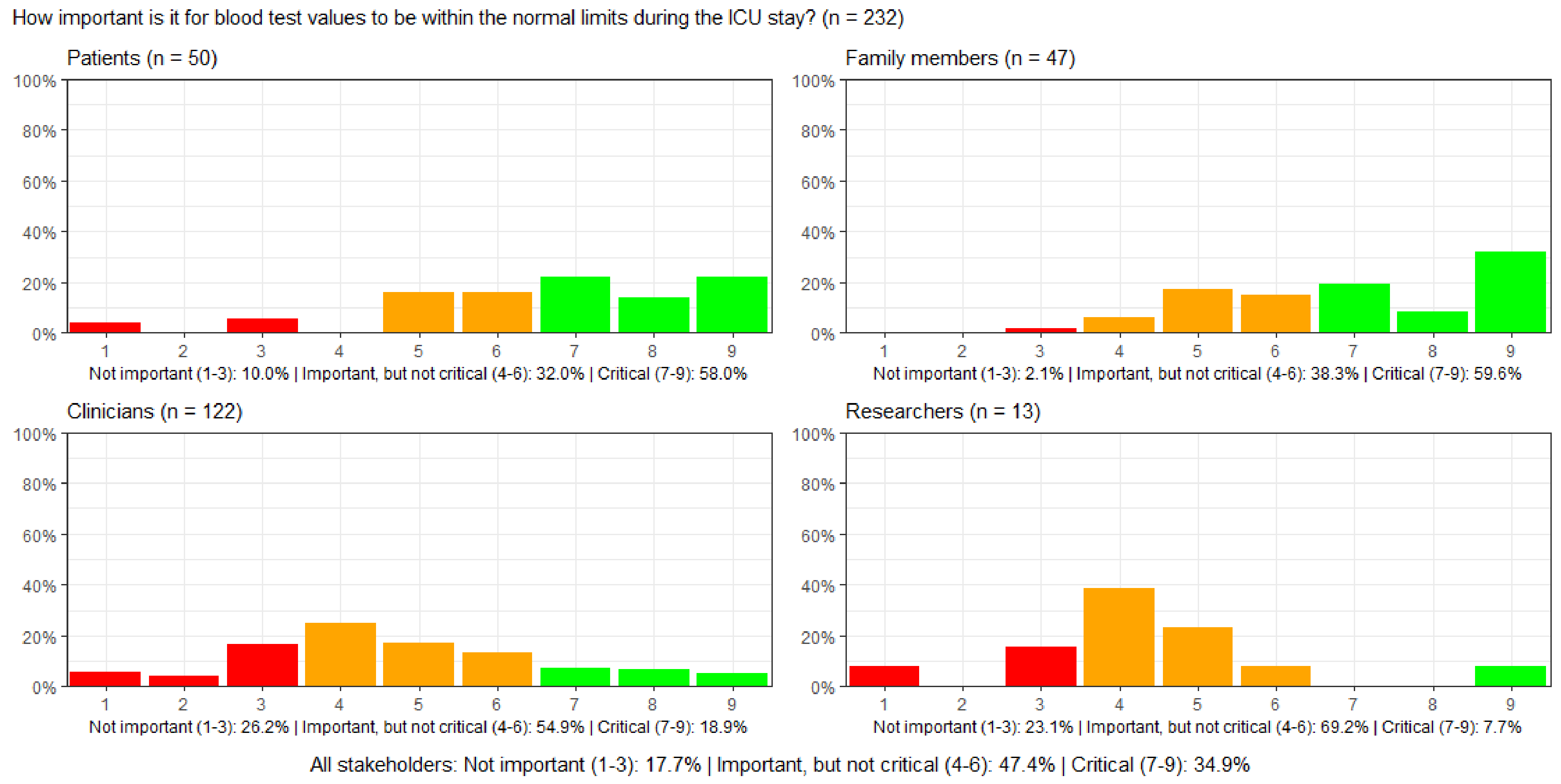

#### Figure S10: How important is it for vital signs measurements to be within the normal range during the ICU stay?

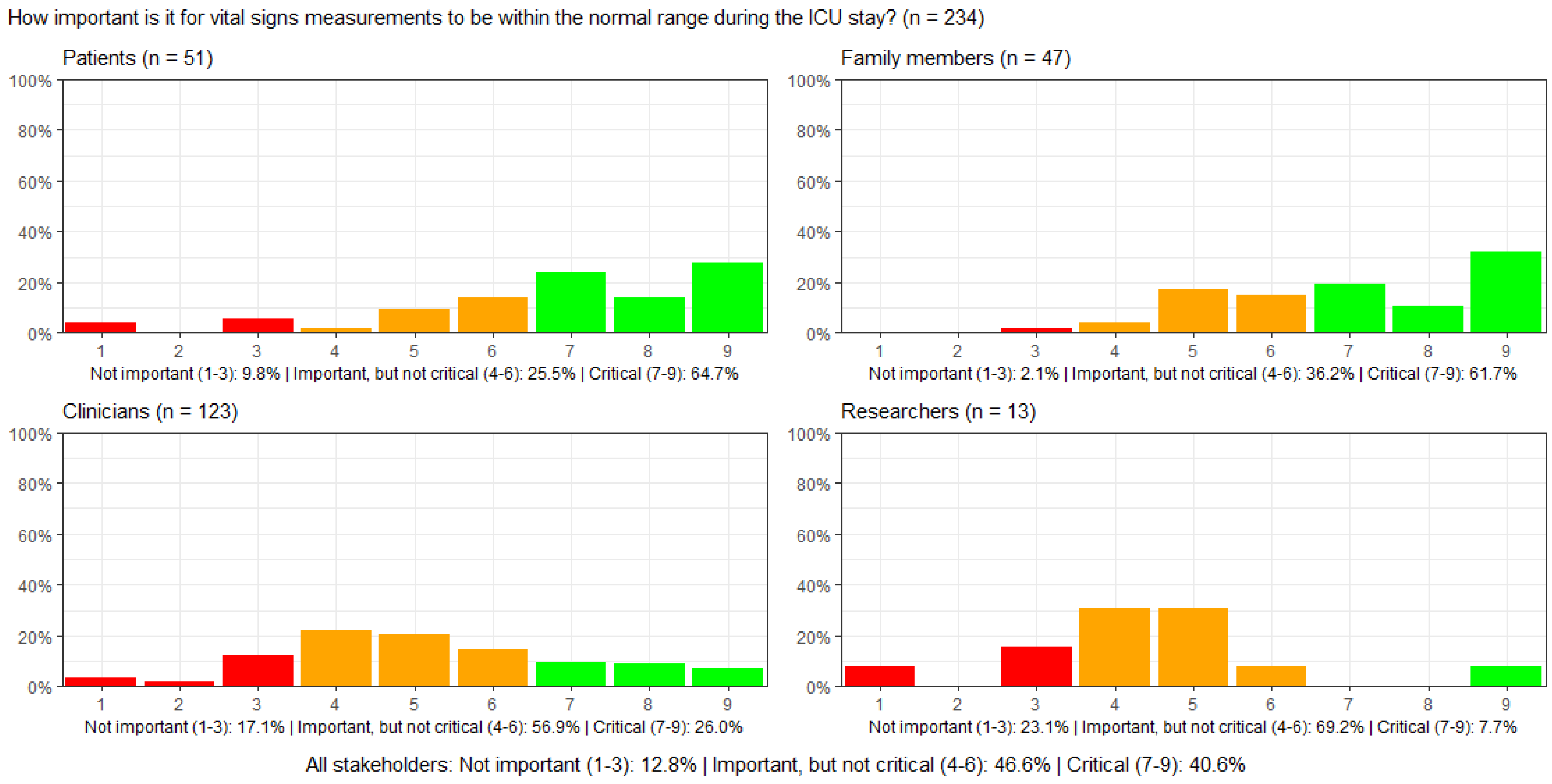

#### Figure S11: How important is the time spent on a mechanical ventilator during the ICU stay?

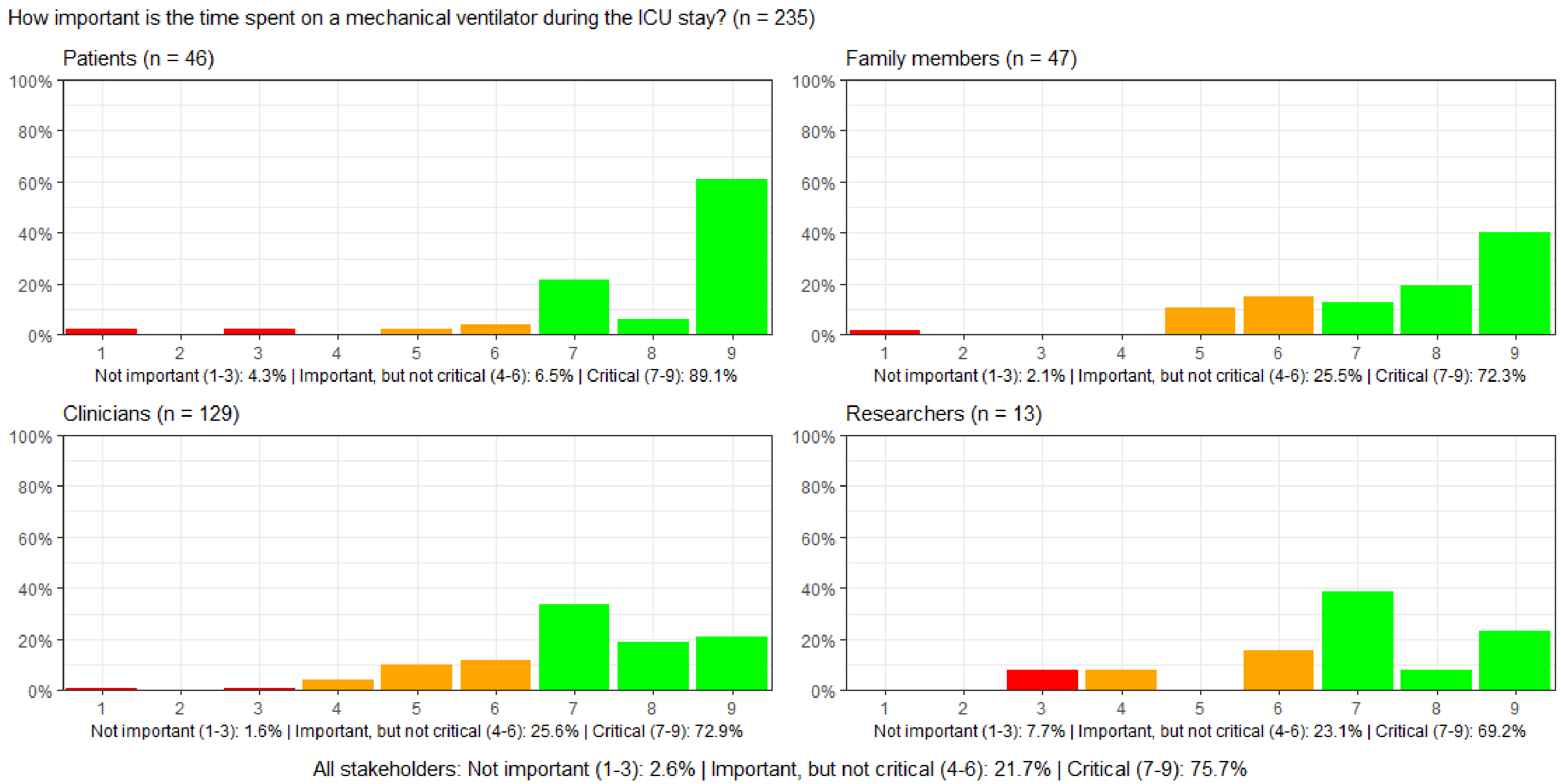

#### Figure S12: How important is the length of time one receives pharmacologic circulatory support during the ICU stay?

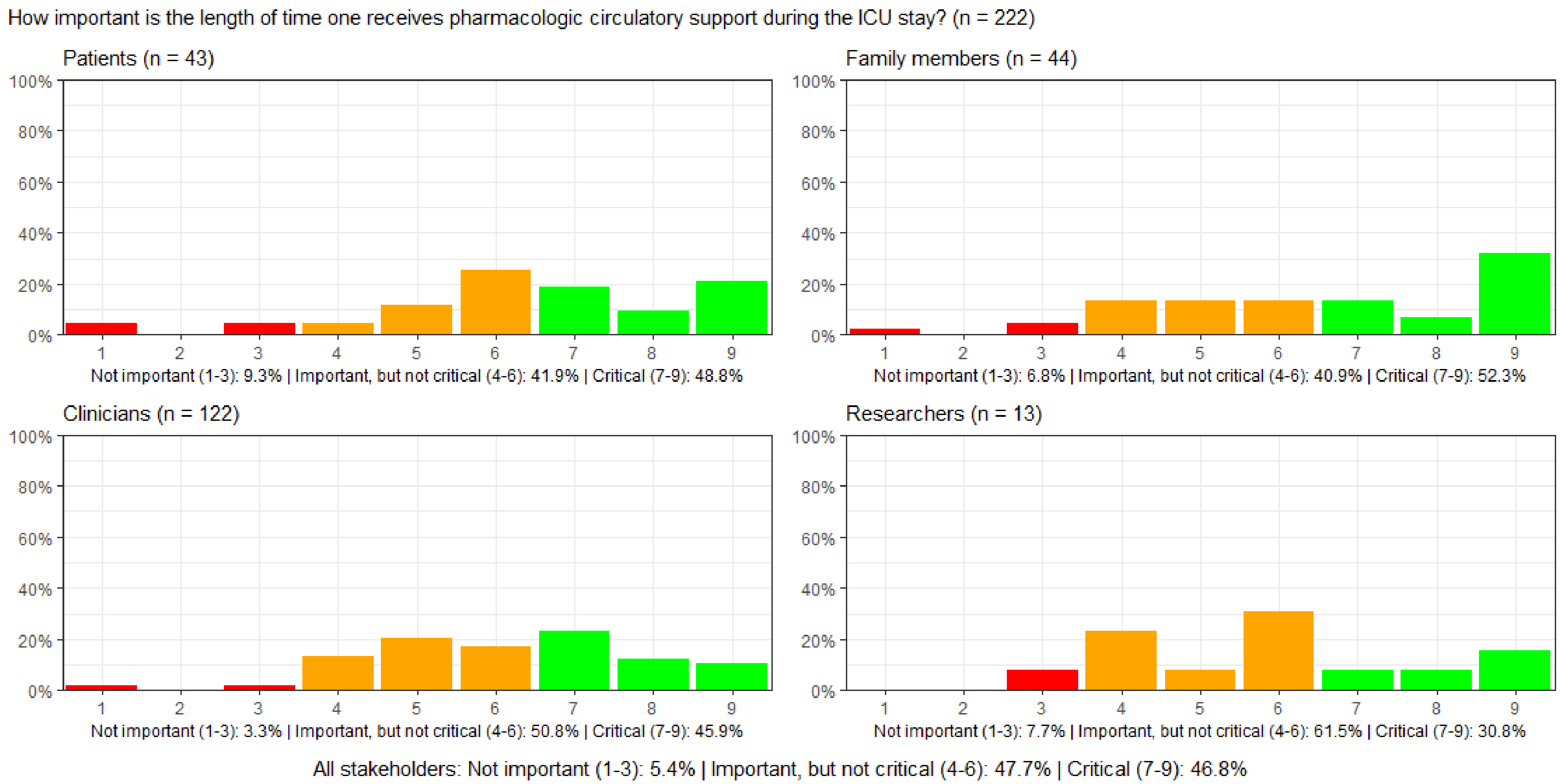

#### Figure S13: How important is the length of time one receives dialysis during the ICU stay?

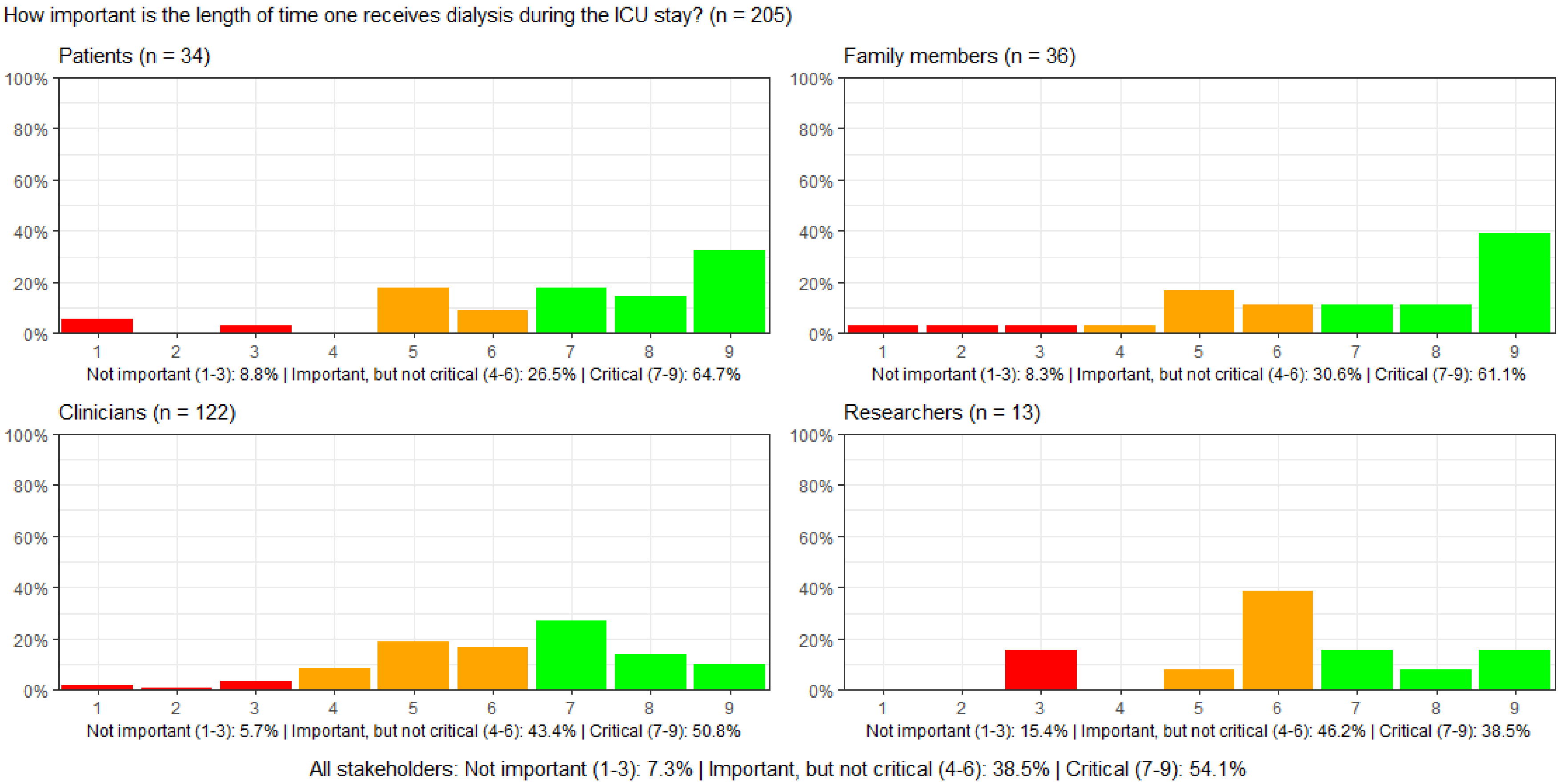

#### Figure S14: How important is the length of time one is in an artificial coma during the ICU stay?

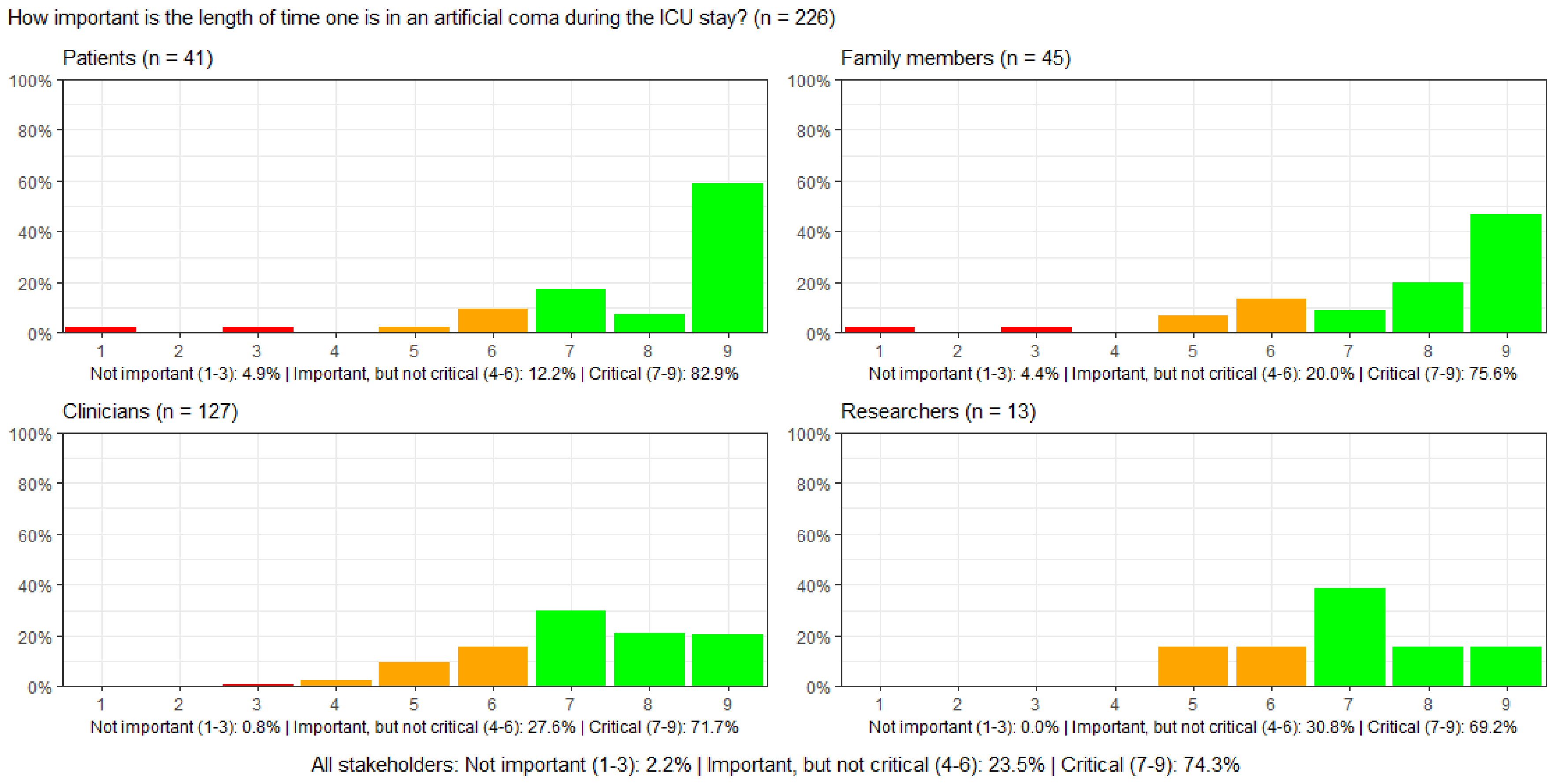

#### Figure S15: How important is it to spend the shortest possible time in delirium during the ICU stay?

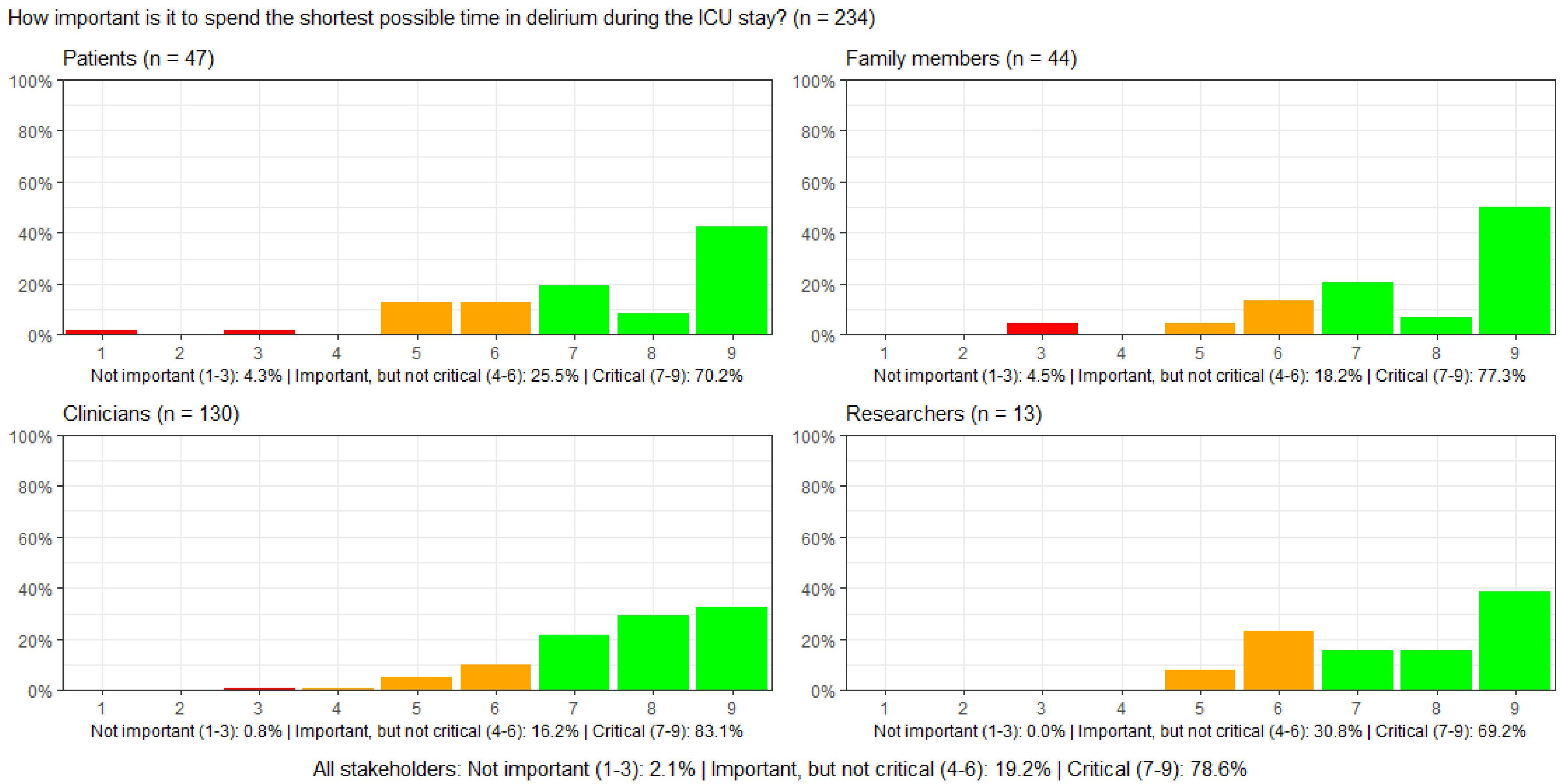

#### Figure S16: How important is the absence of nausea and vomiting during the ICU stay?

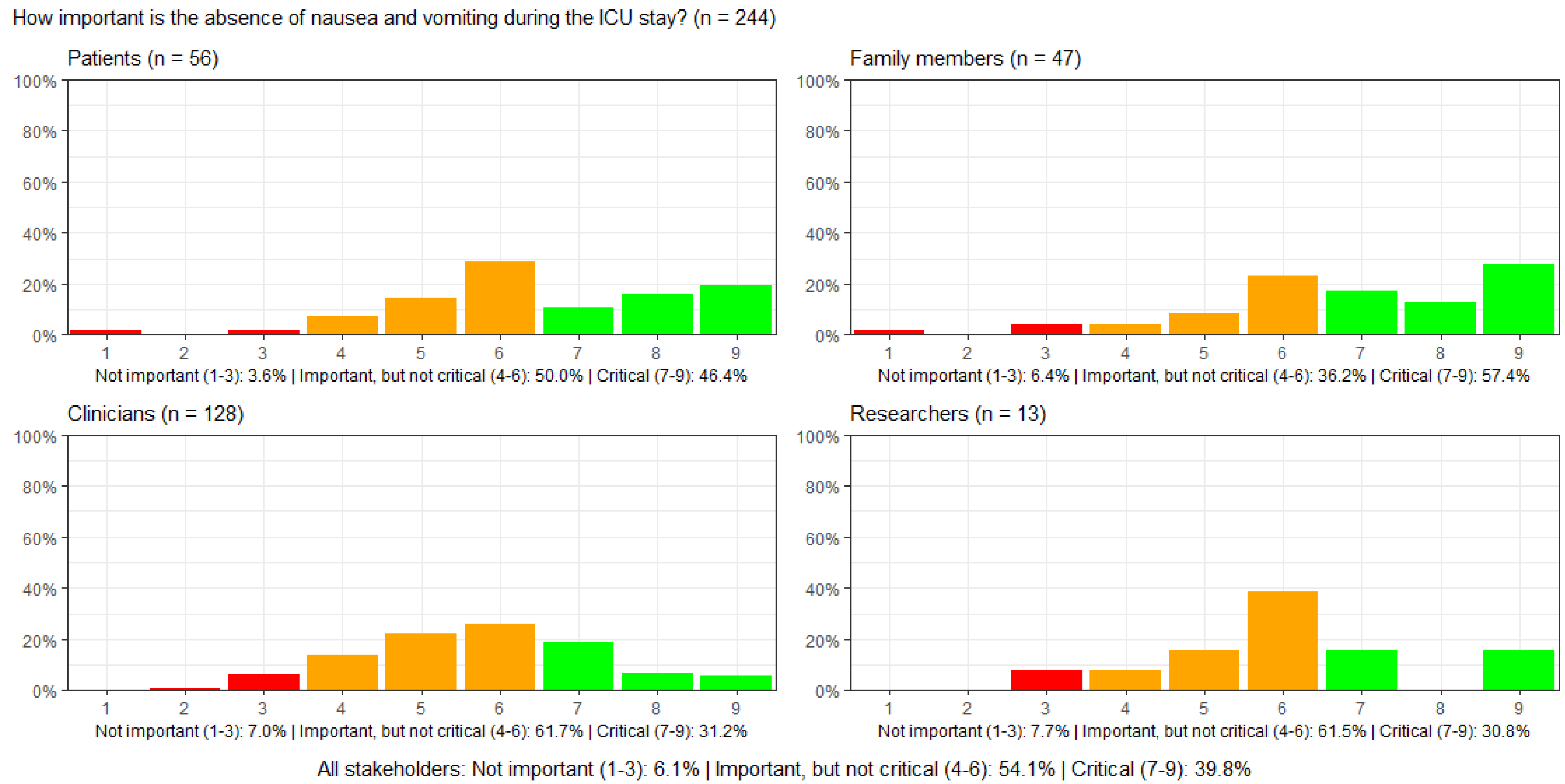

#### Figure S17: How important is the absence of thirst during the ICU stay?

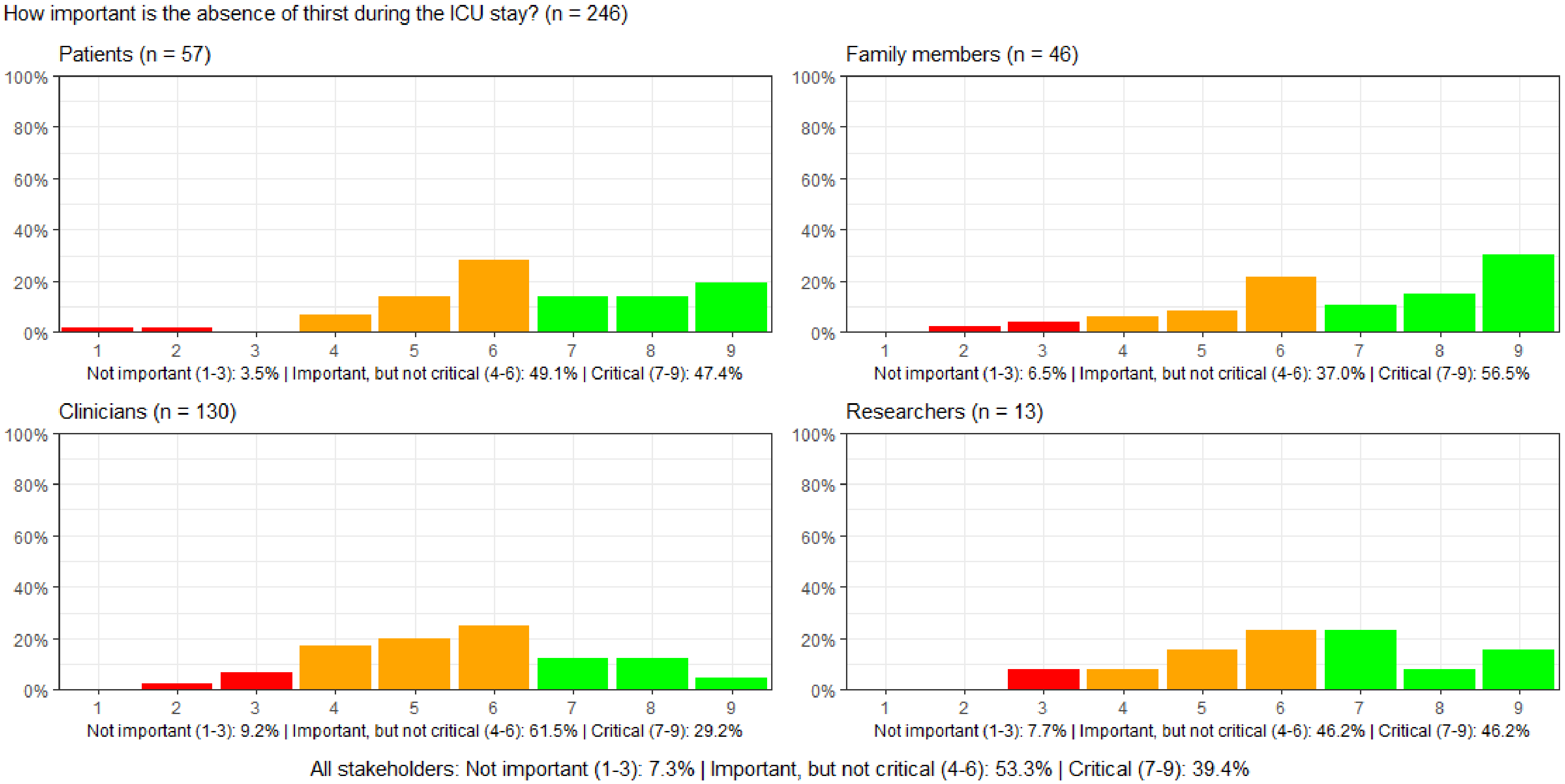

#### Figure S18: How important is the absence of shortness of breath during the ICU stay?

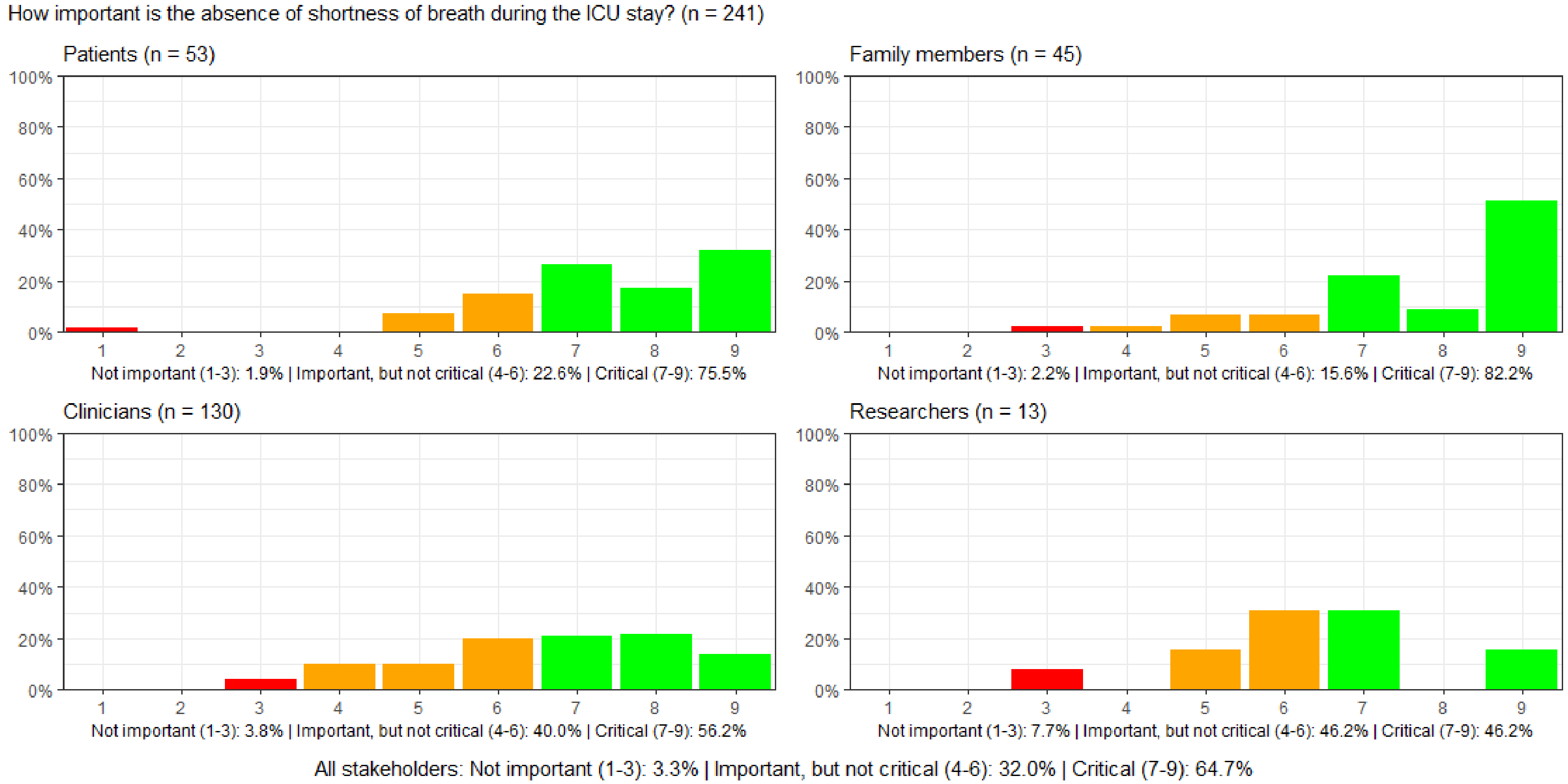

#### Figure S19: How important is the absence of pain during the ICU stay?

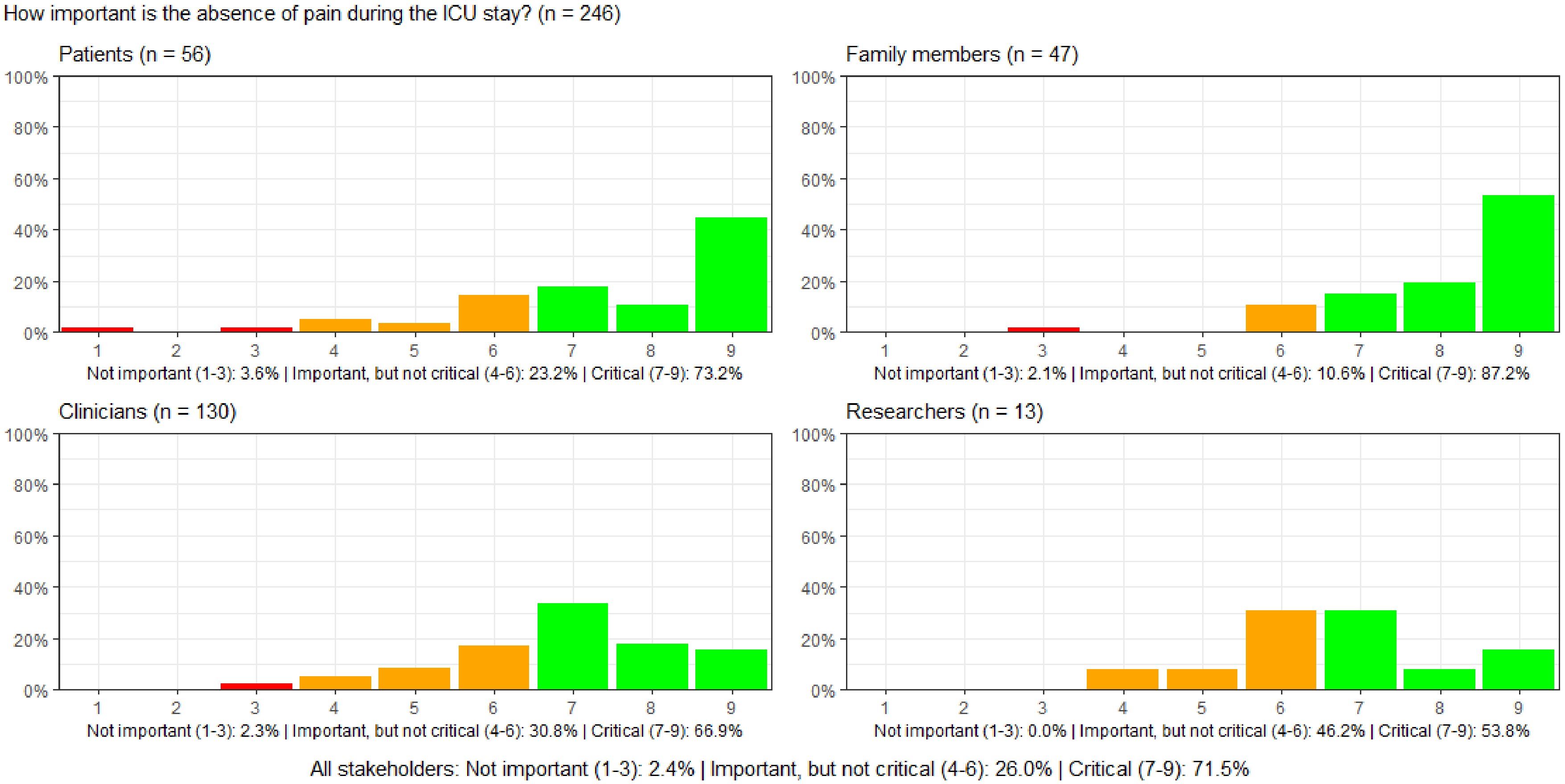

#### Figure S20: How important is the absence of anxiety during the ICU stay?

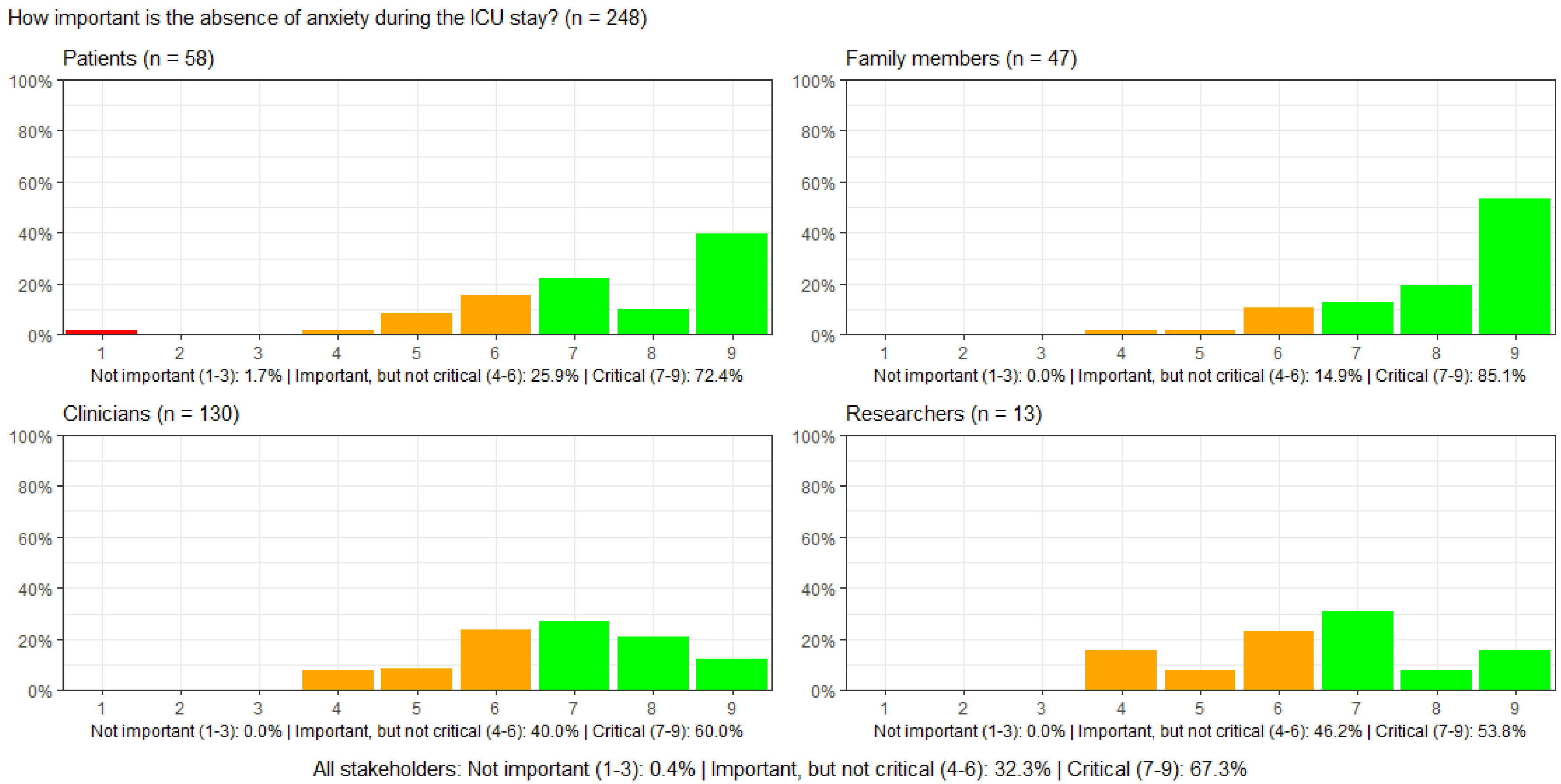

#### Figure S21: How important is sleep and maintaining circadian rhythm during the ICU stay?

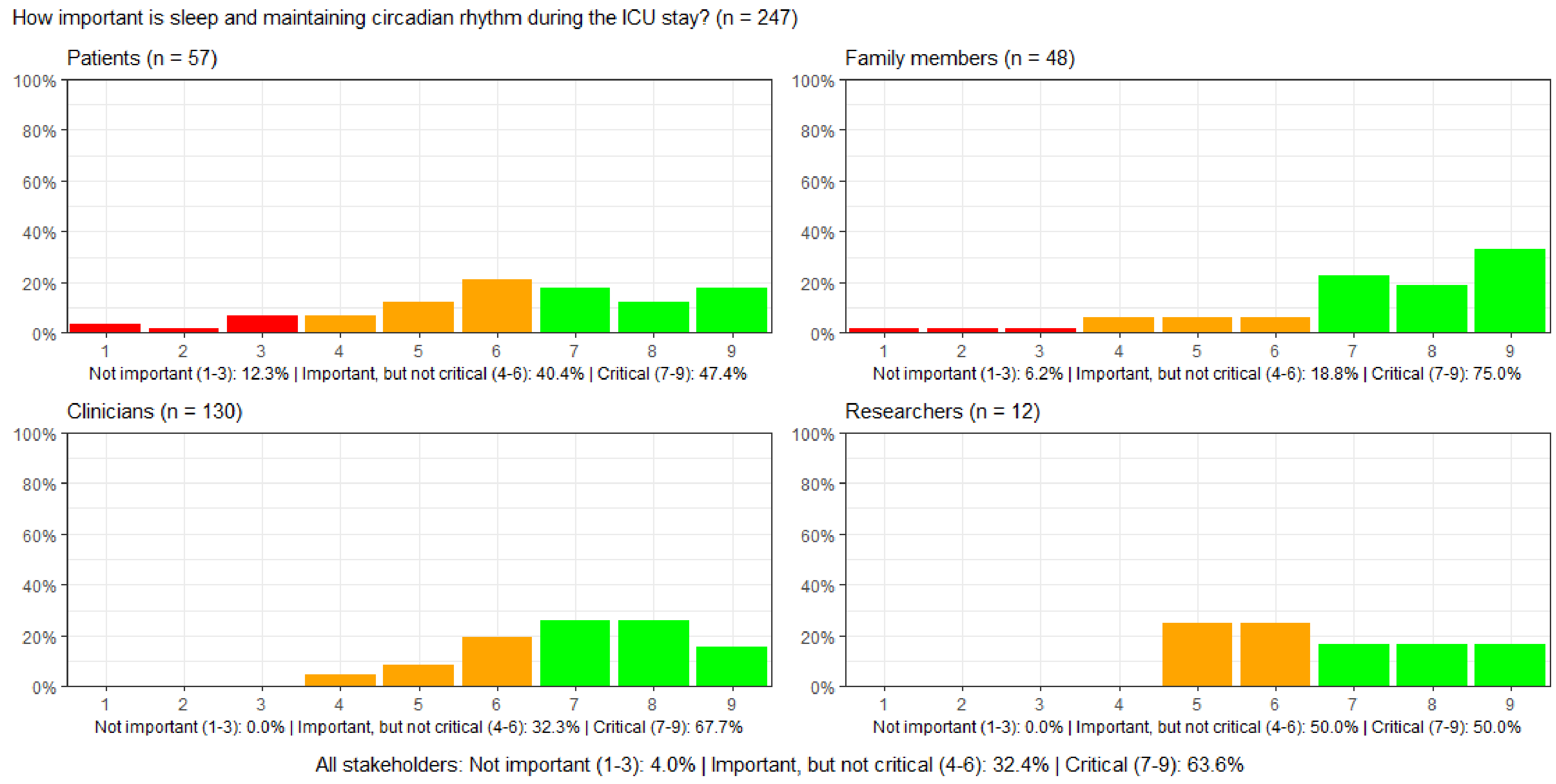

#### Figure S22: How important is it to be able to communicate during the ICU stay?

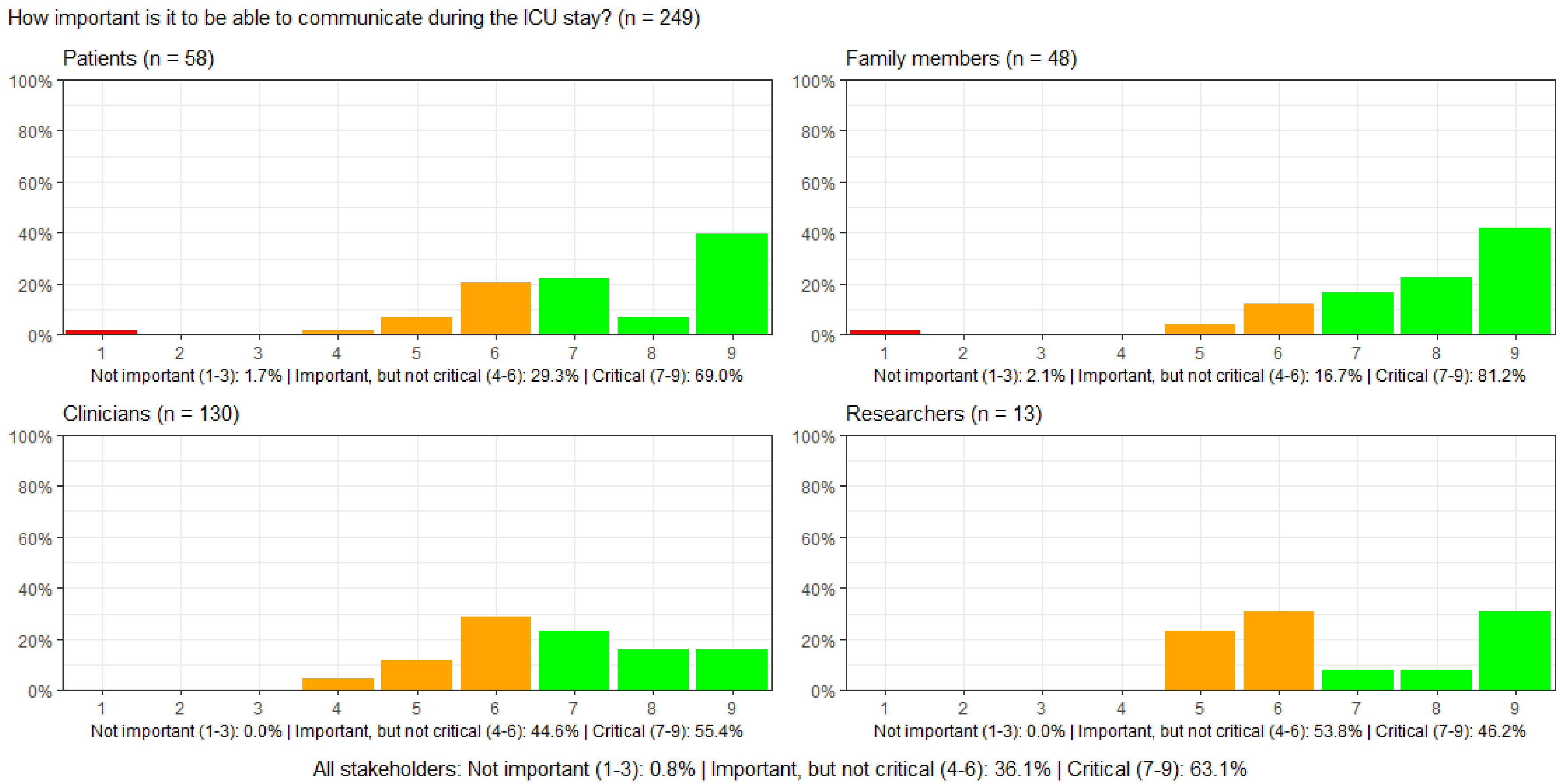

#### Figure S23: How important is the place one is discharged to after the ICU stay?

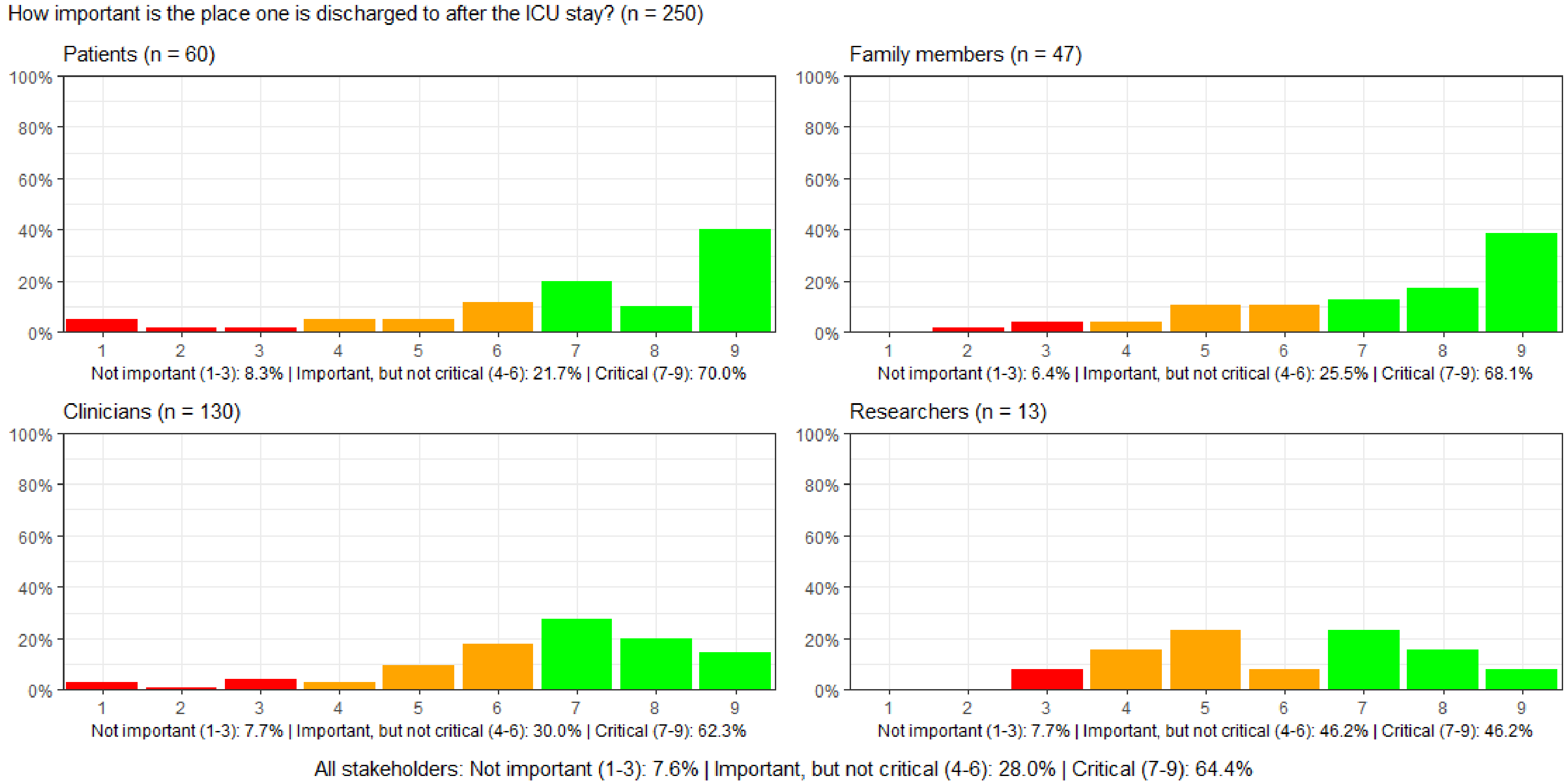

#### Figure S24: How important is the number of outpatient appointments or general practitioner visits in the follow-up after ICU discharge?

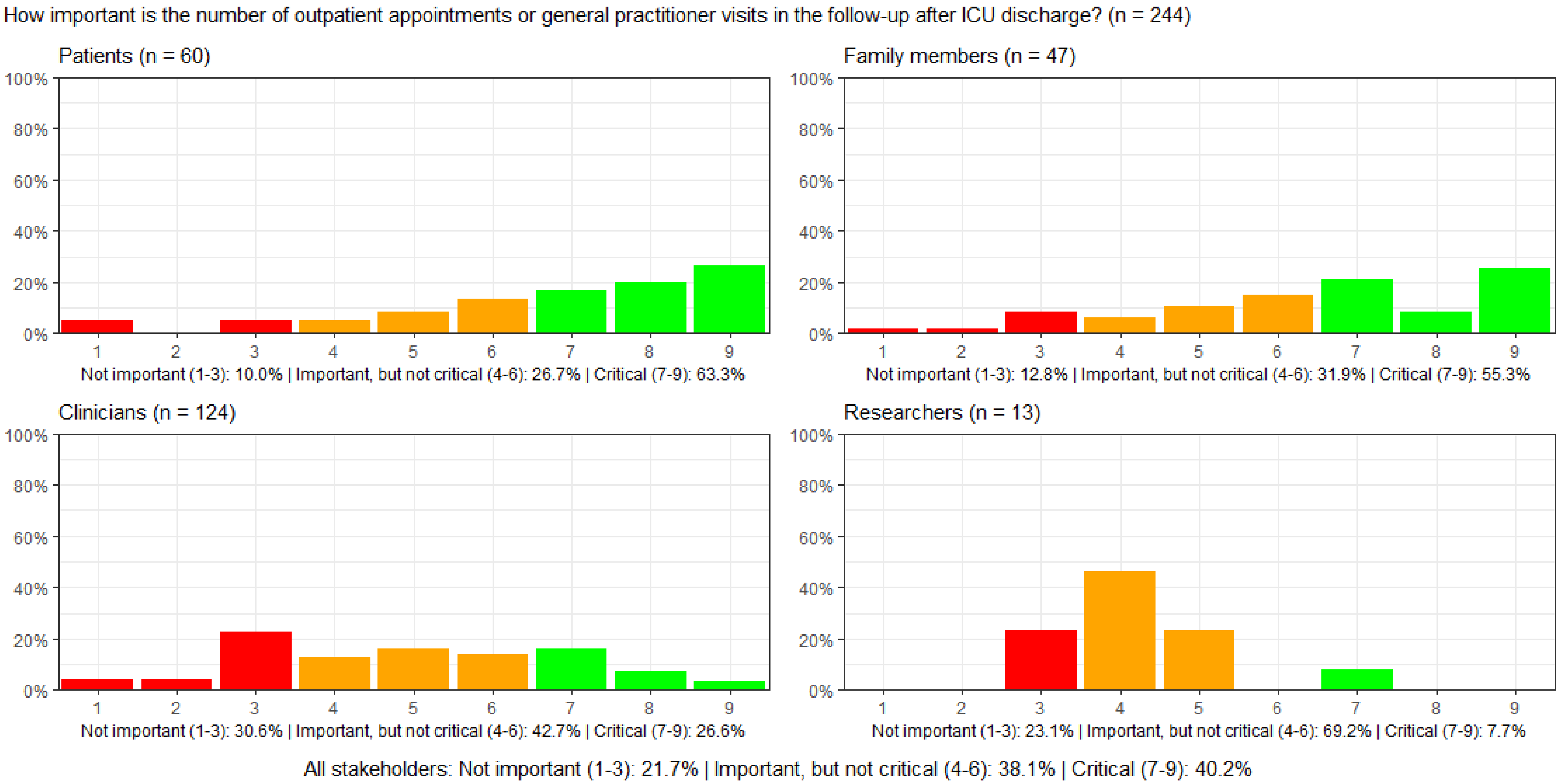

#### Figure S25: How important is it to avoid needing new medication after ICU discharge?

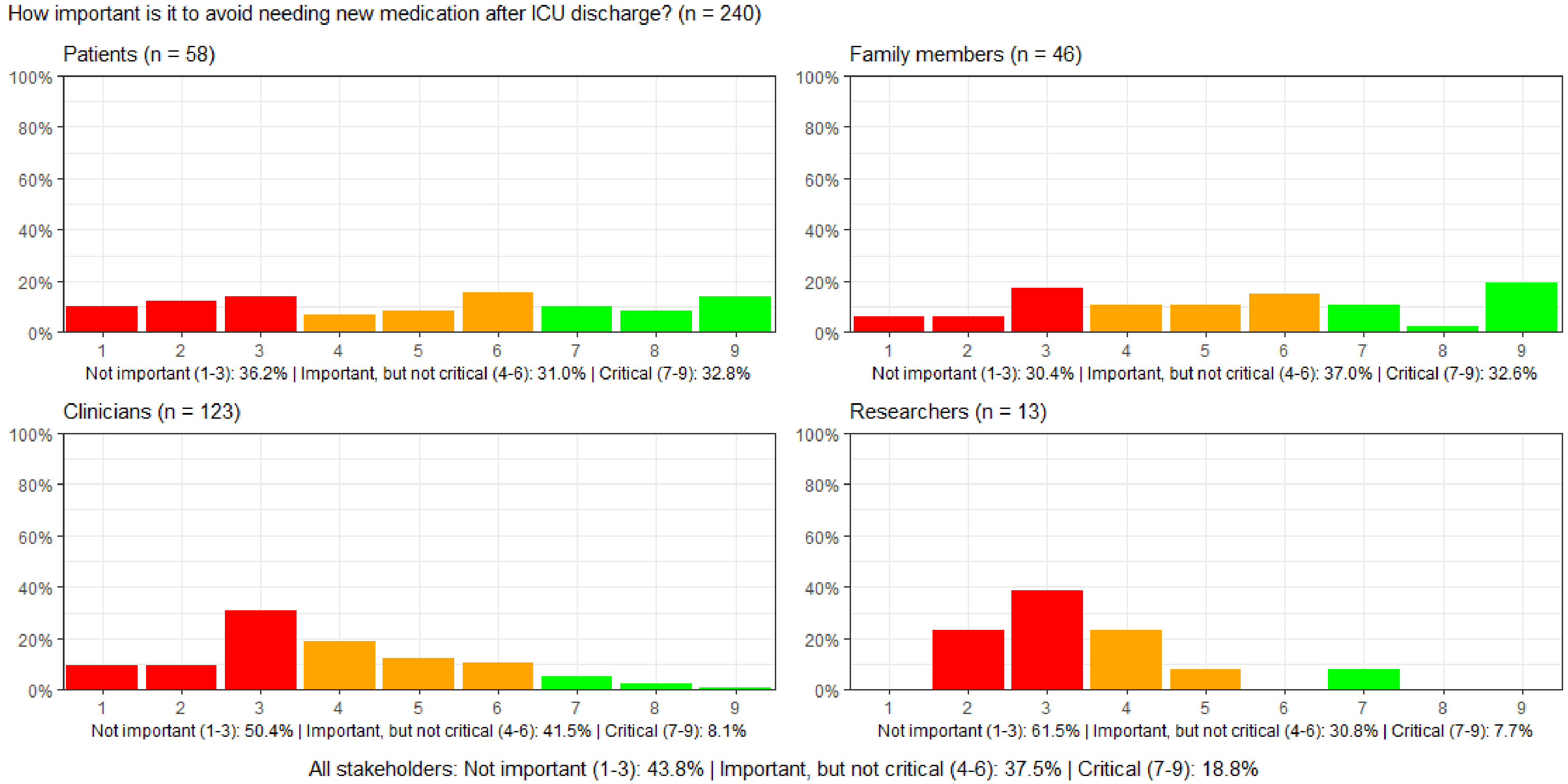

#### Figure S26: How important is health-related quality of life after ICU discharge?

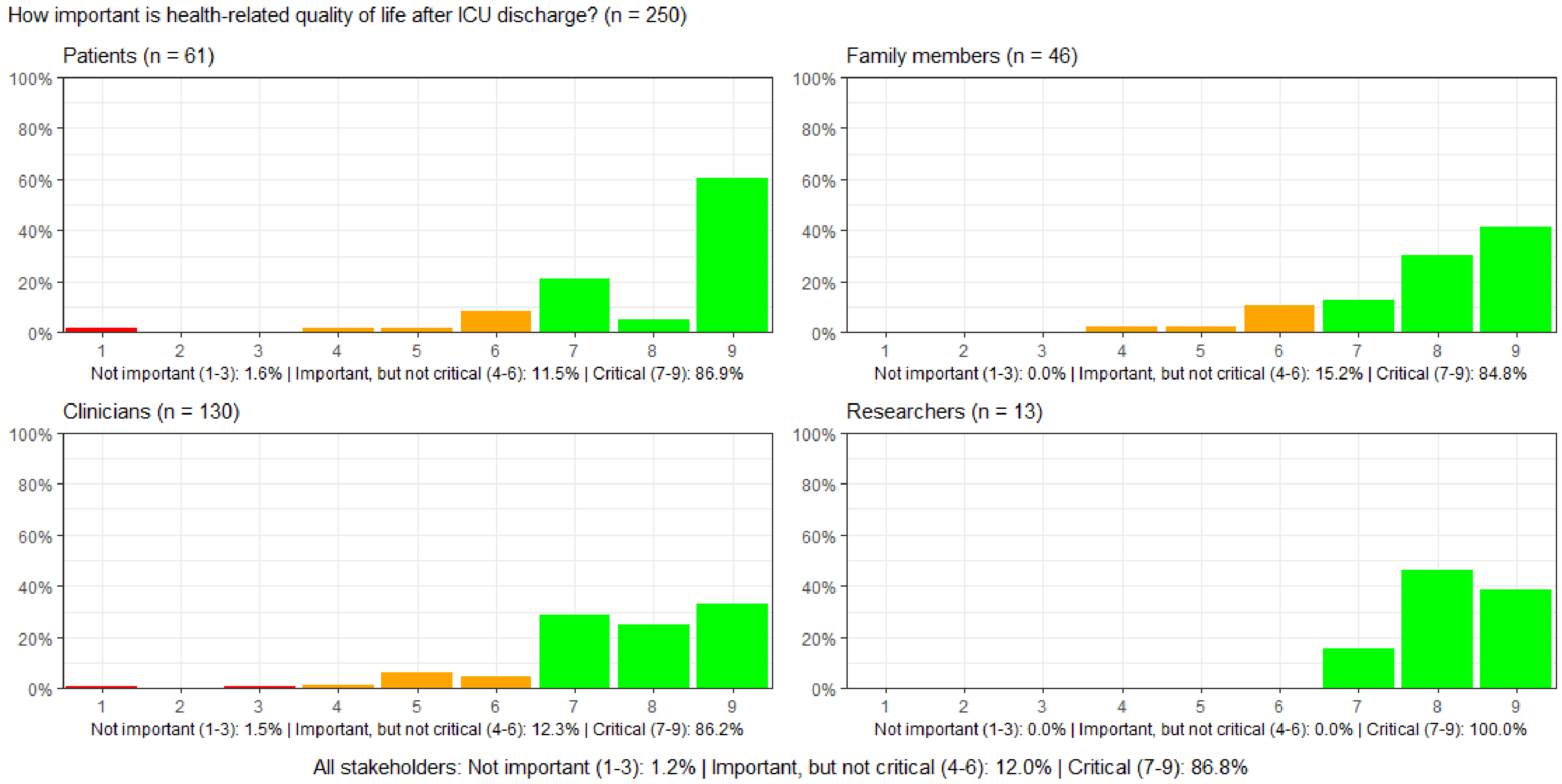

#### Figure S27: How important is it to not experience fatigue that cannot be slept away after ICU discharge?

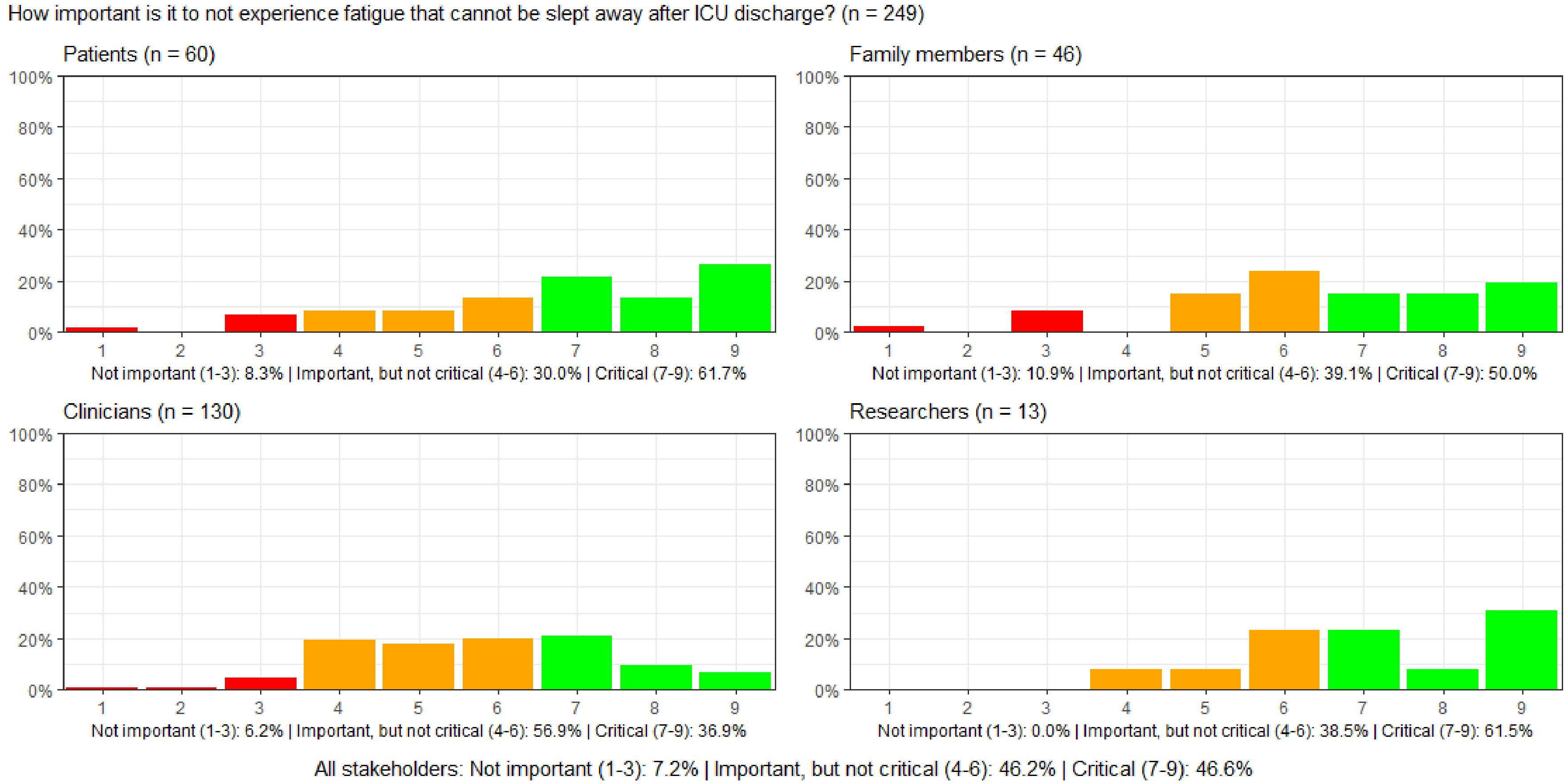

#### Figure S28: How important is it to not have pain after ICU discharge?

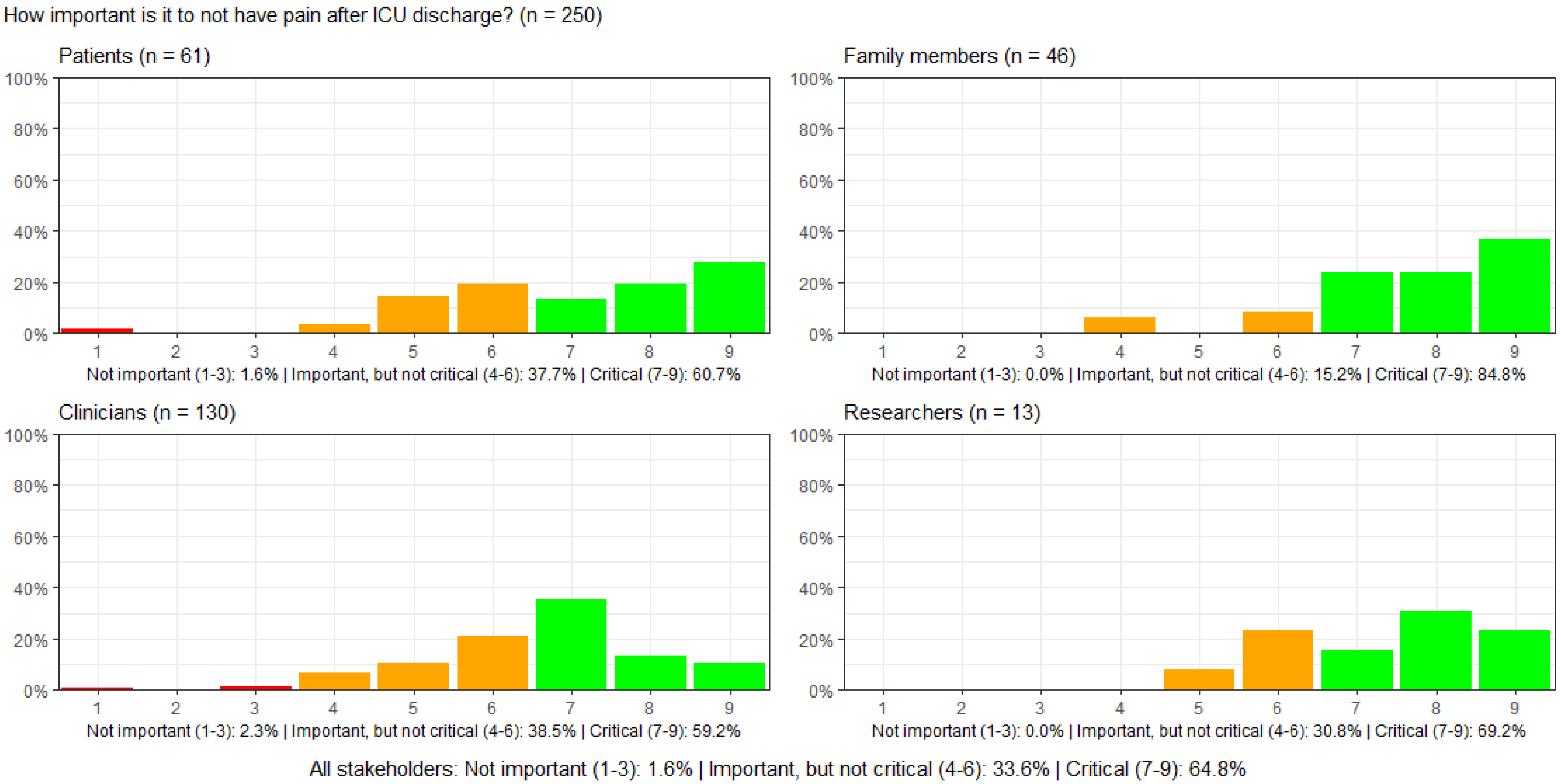

#### Figure S29: How important is it to resume one's usual role in daily life after ICU discharge?

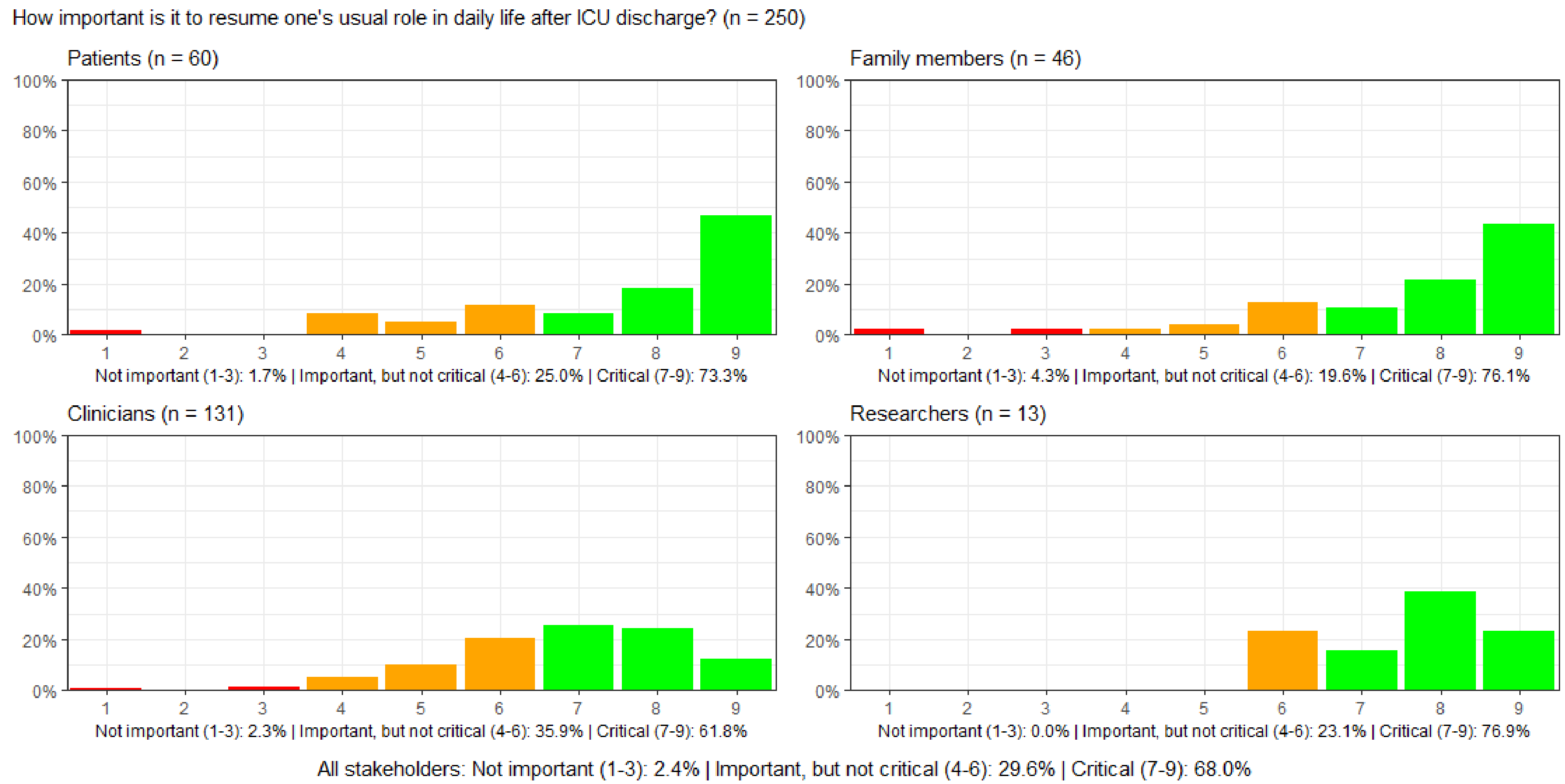

#### Figure S30: How important is family caregivers' experienced quality of life after ICU discharge?

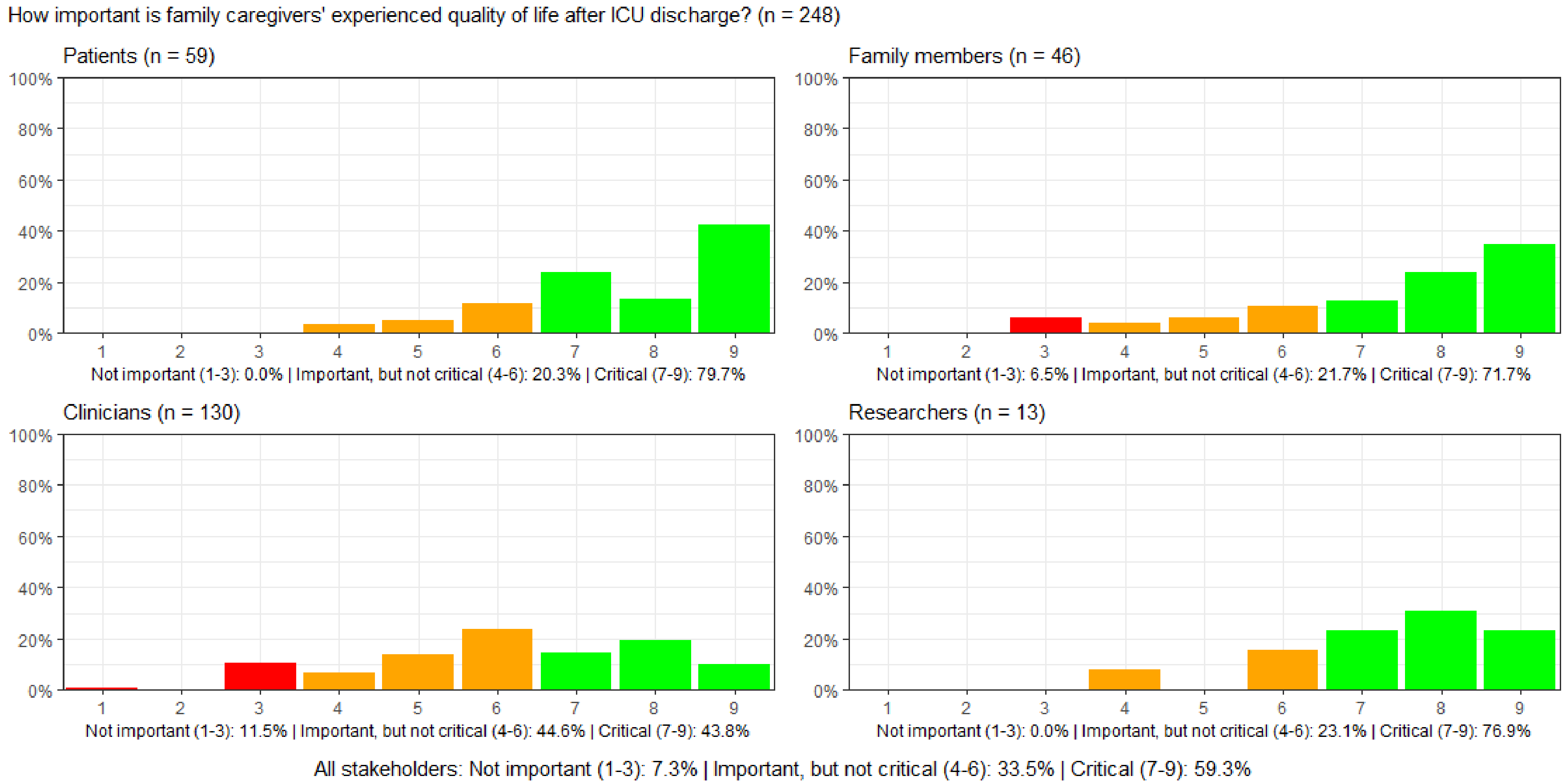

##### Figure S31: How important is it to be financially independent after ICU discharge?

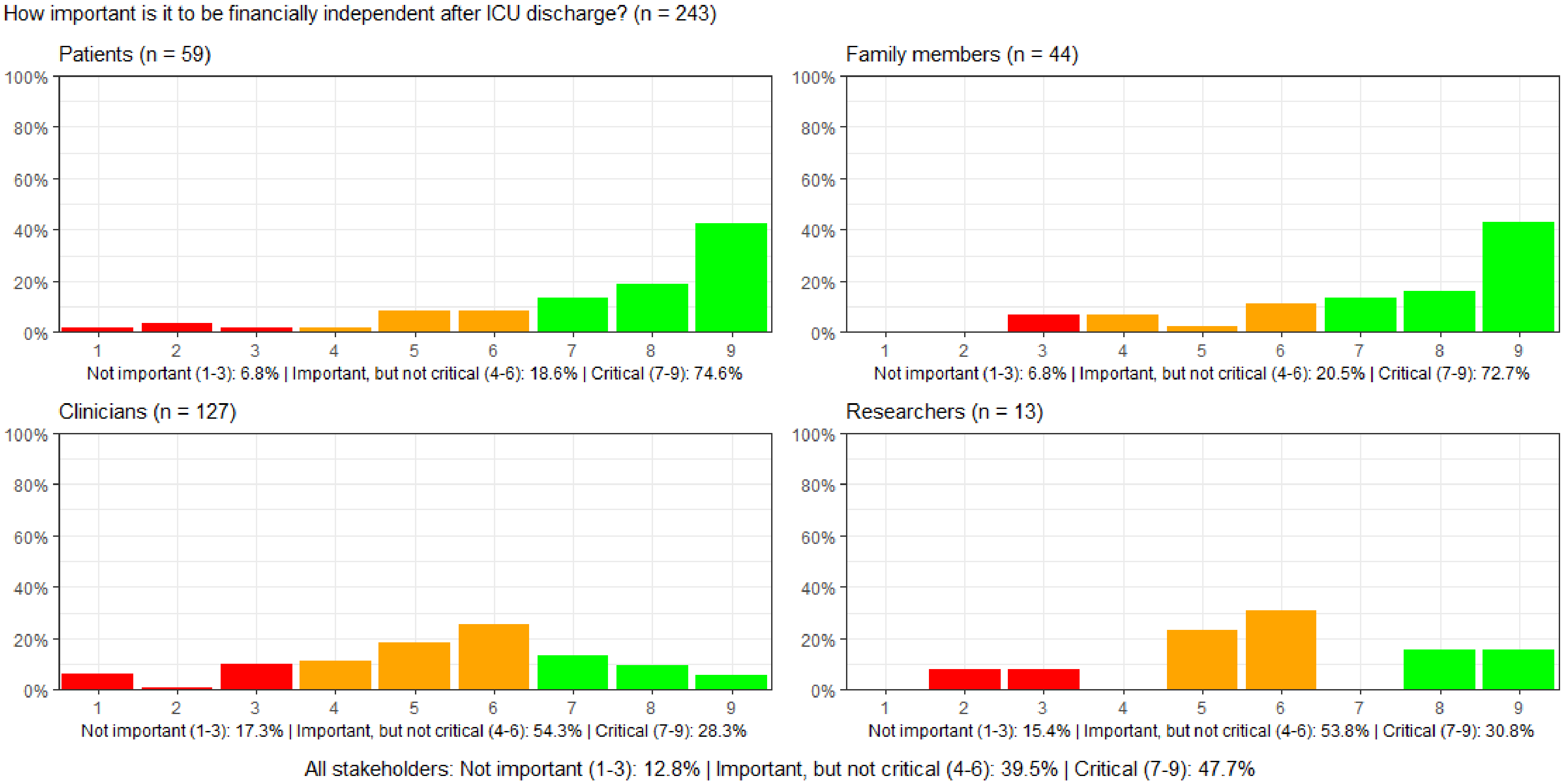

#### Figure S32: How important is it to return to work after ICU discharge?

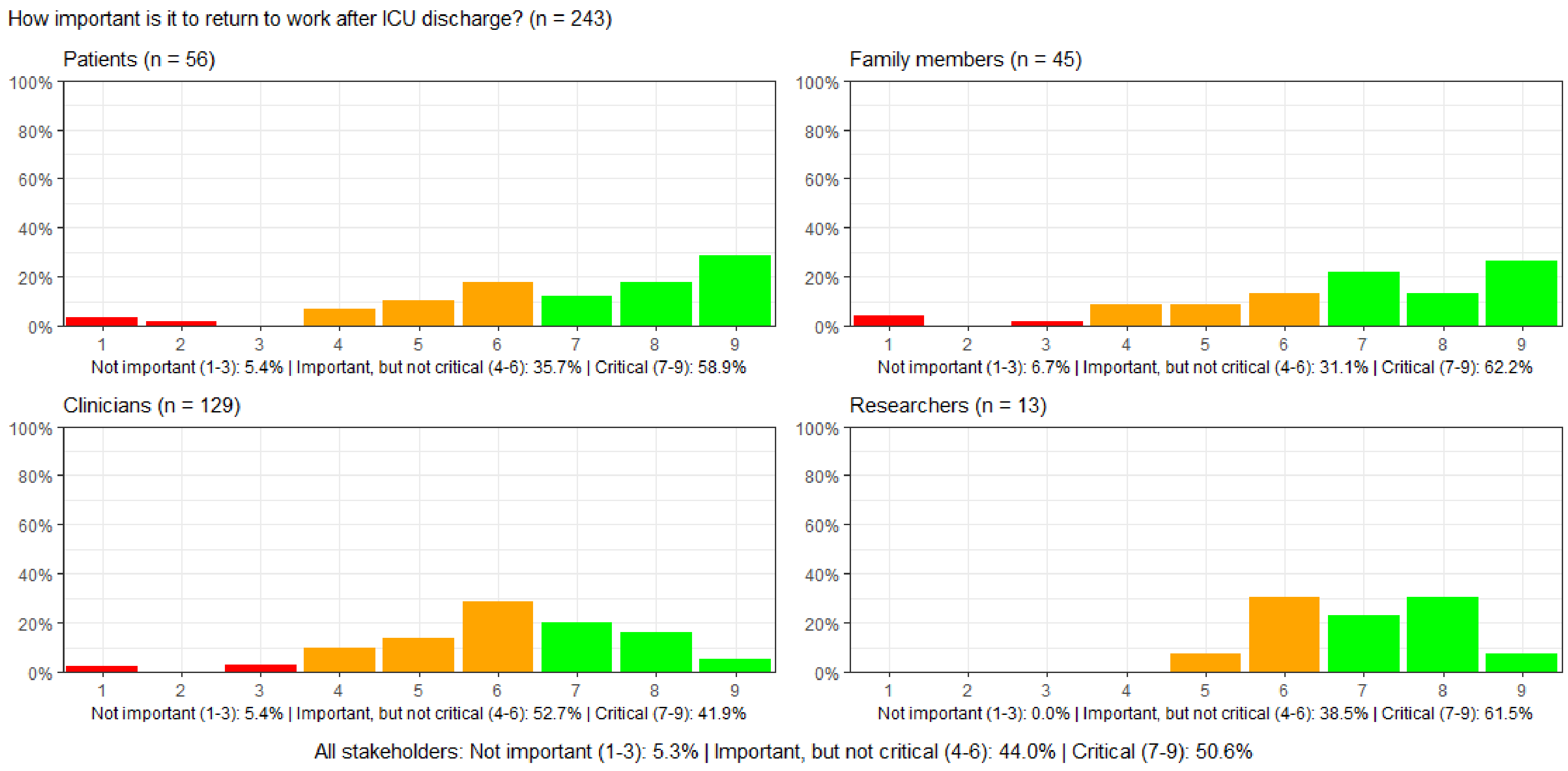

#### Figure S33: How important is it to avoid physical impediment after ICU discharge?

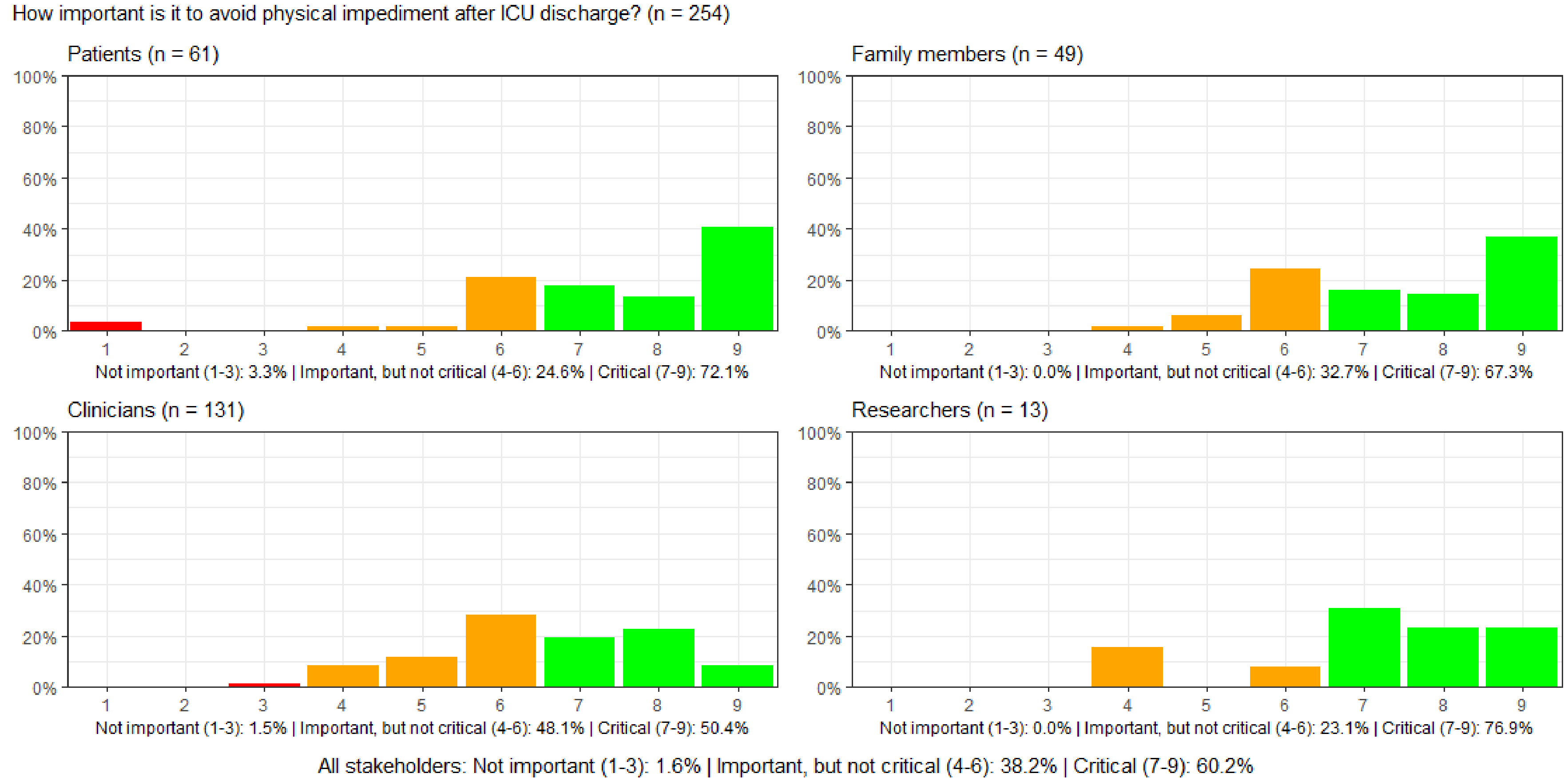

#### Figure S34: How important is it to be able to perform activities of daily living independently after ICU discharge?

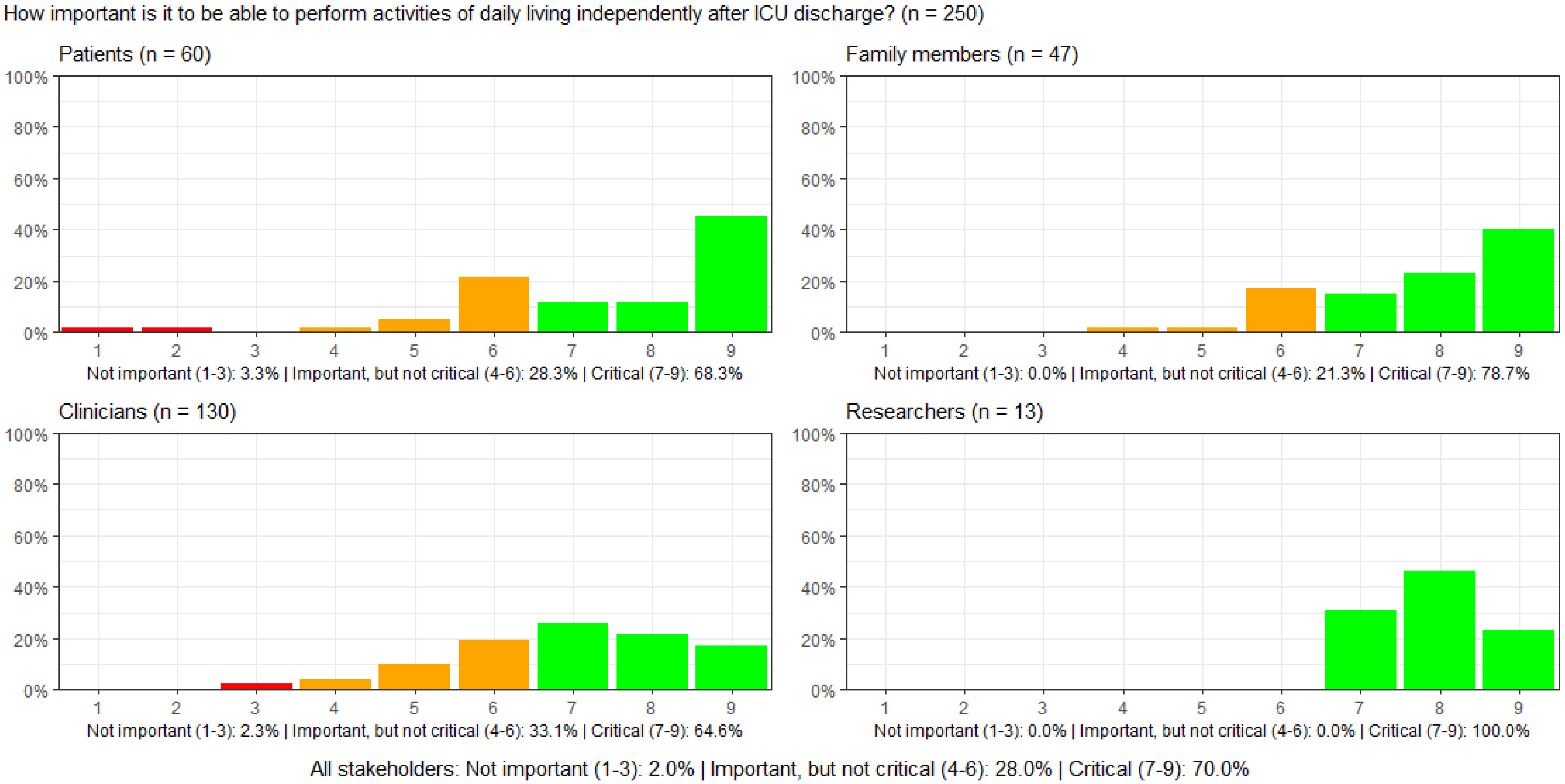

#### Figure S35: How important is it to be able to dress oneself after ICU discharge?

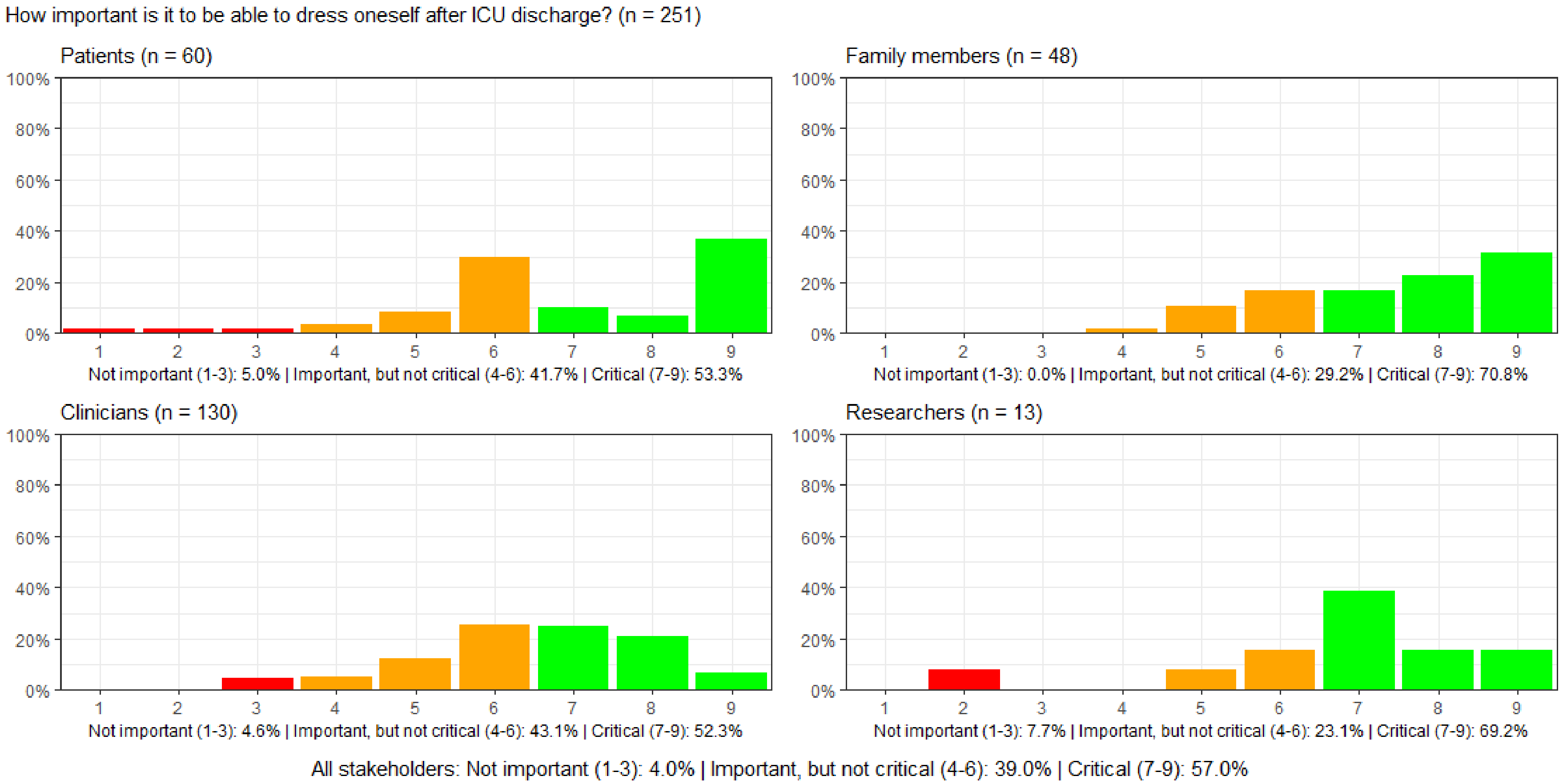

#### Figure S36: How important is it to be mobile after ICU discharge?

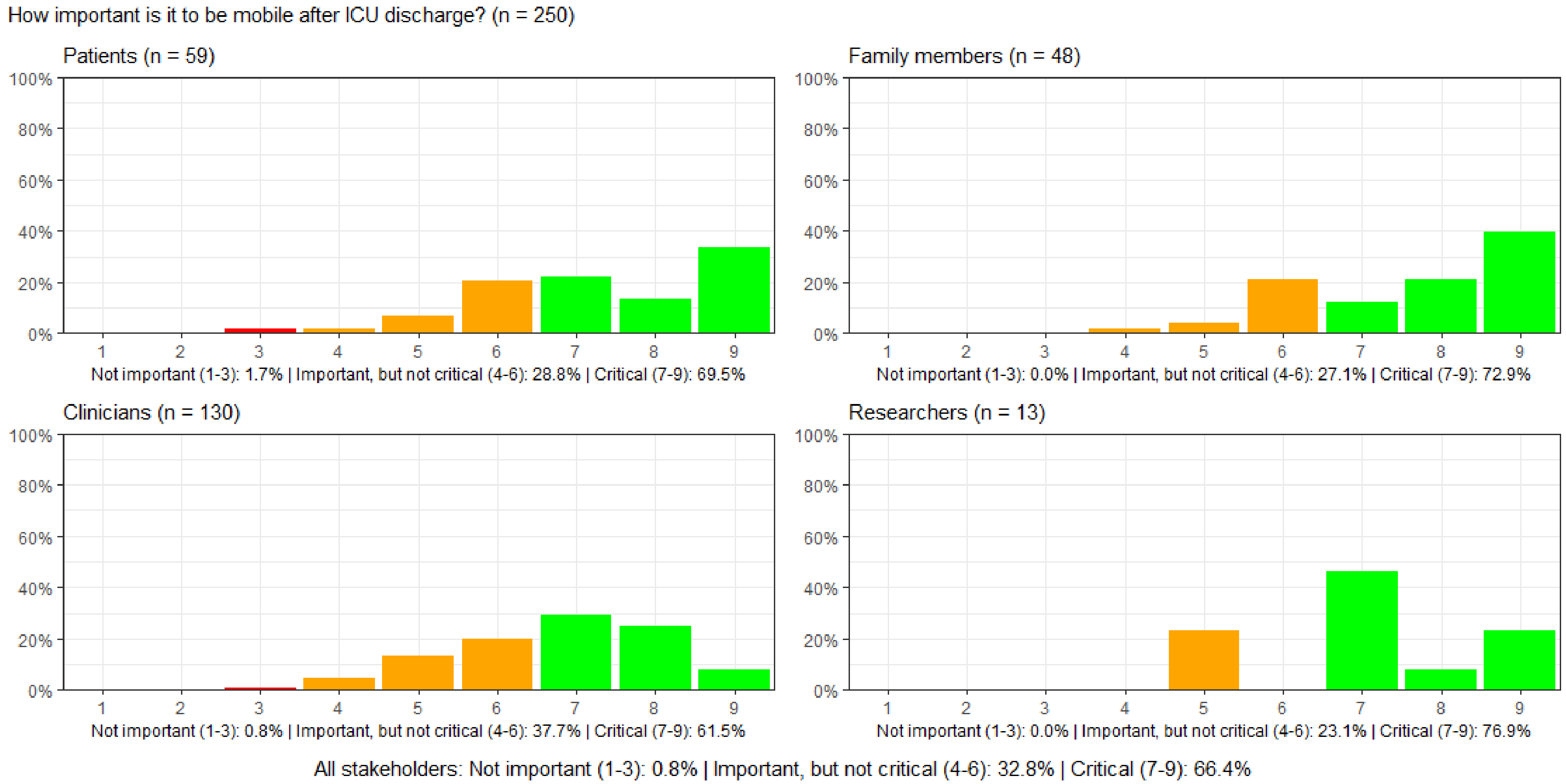

#### Figure S37: How important is it to be free from shortness of breath after ICU discharge?

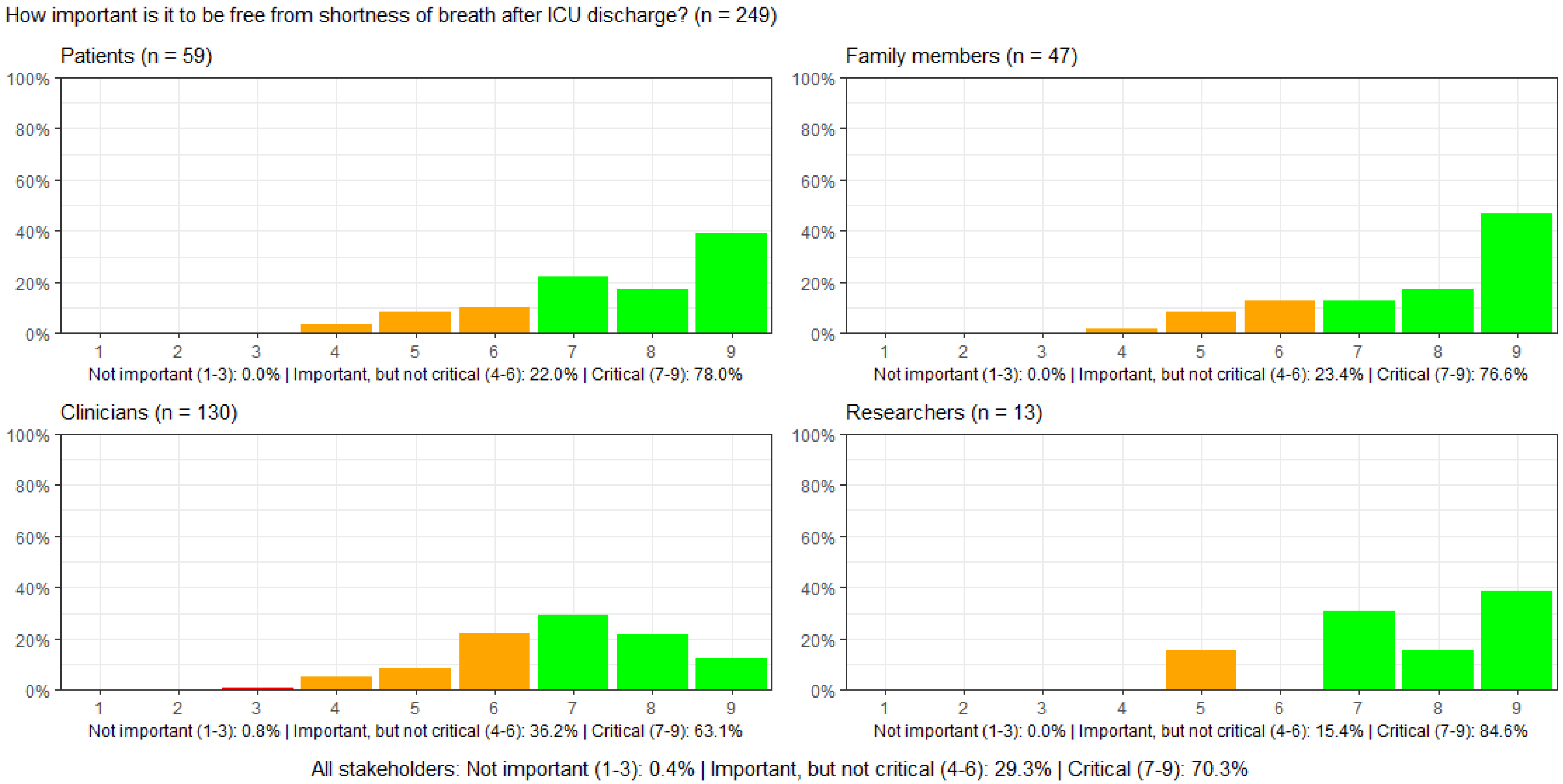

#### Figure S38: How important is it to be free from voice problems after ICU discharge?

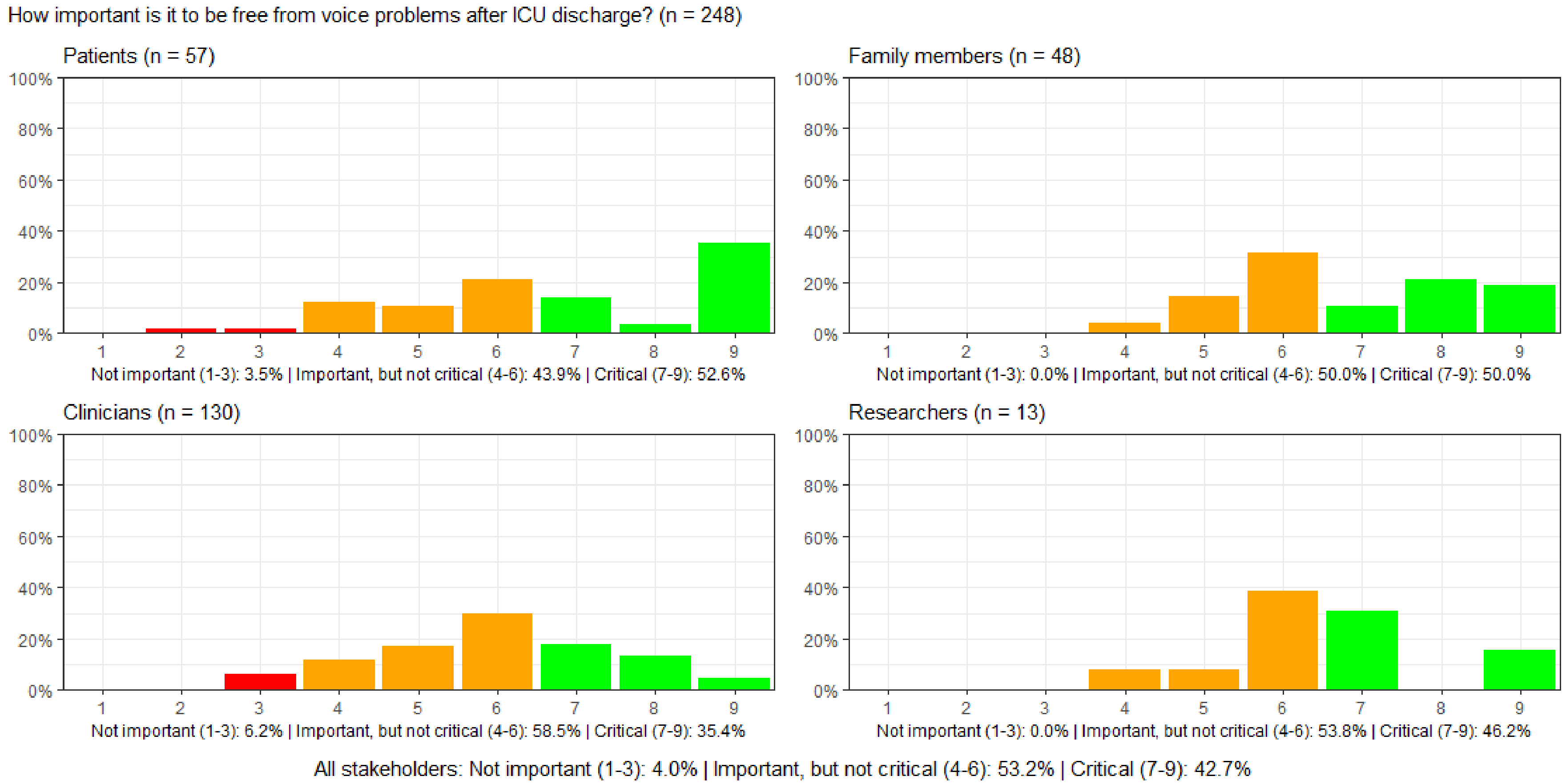

#### Figure S39: How important is it to be free from swallowing difficulties after ICU discharge?

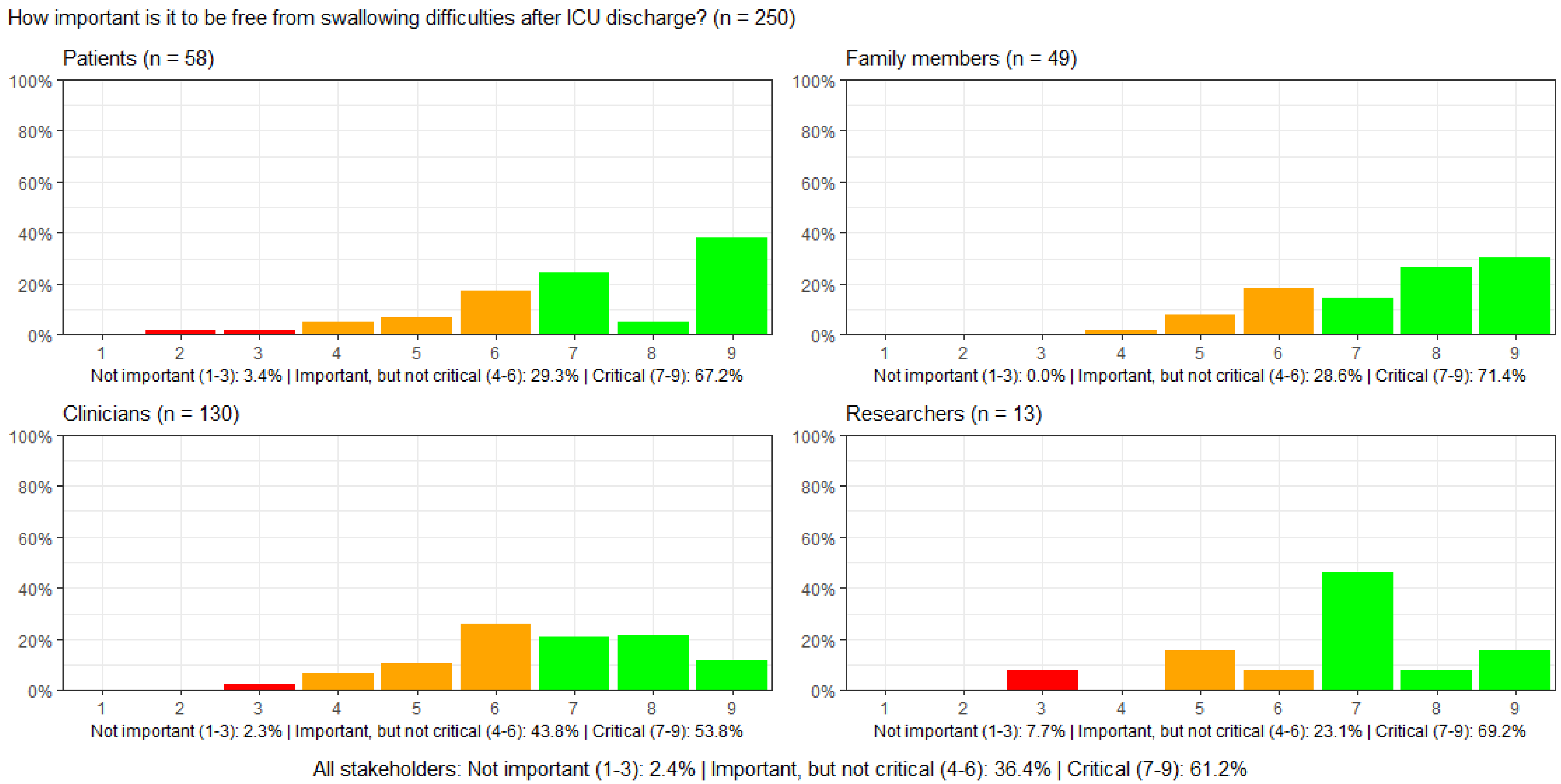

#### Figure S40: How important is it to maintain bowel function after ICU discharge?

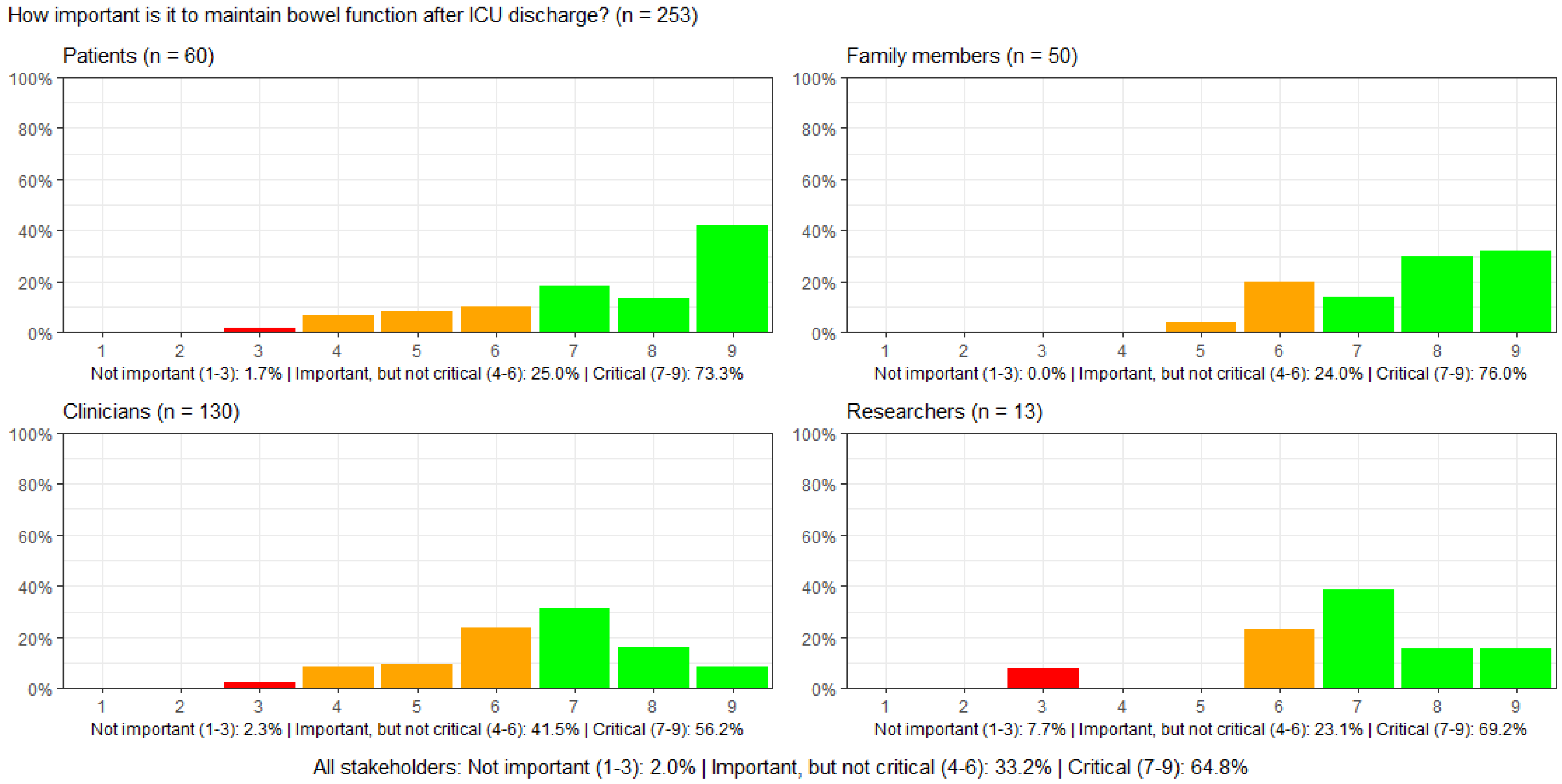

#### Figure S41: How important is it to maintain urinary function after ICU discharge?

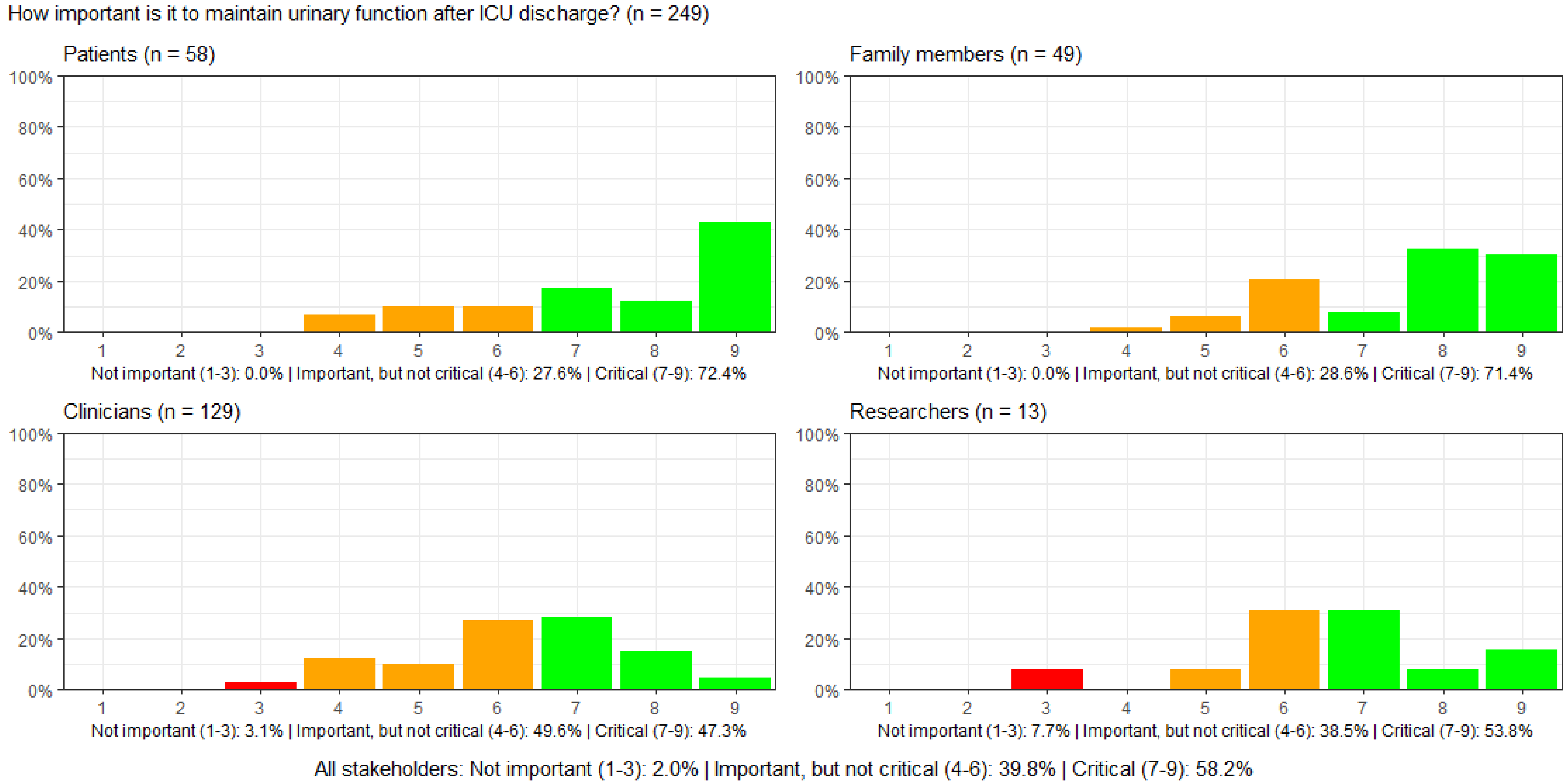

#### Figure S42: How important is it to have intact sexual function after ICU discharge?

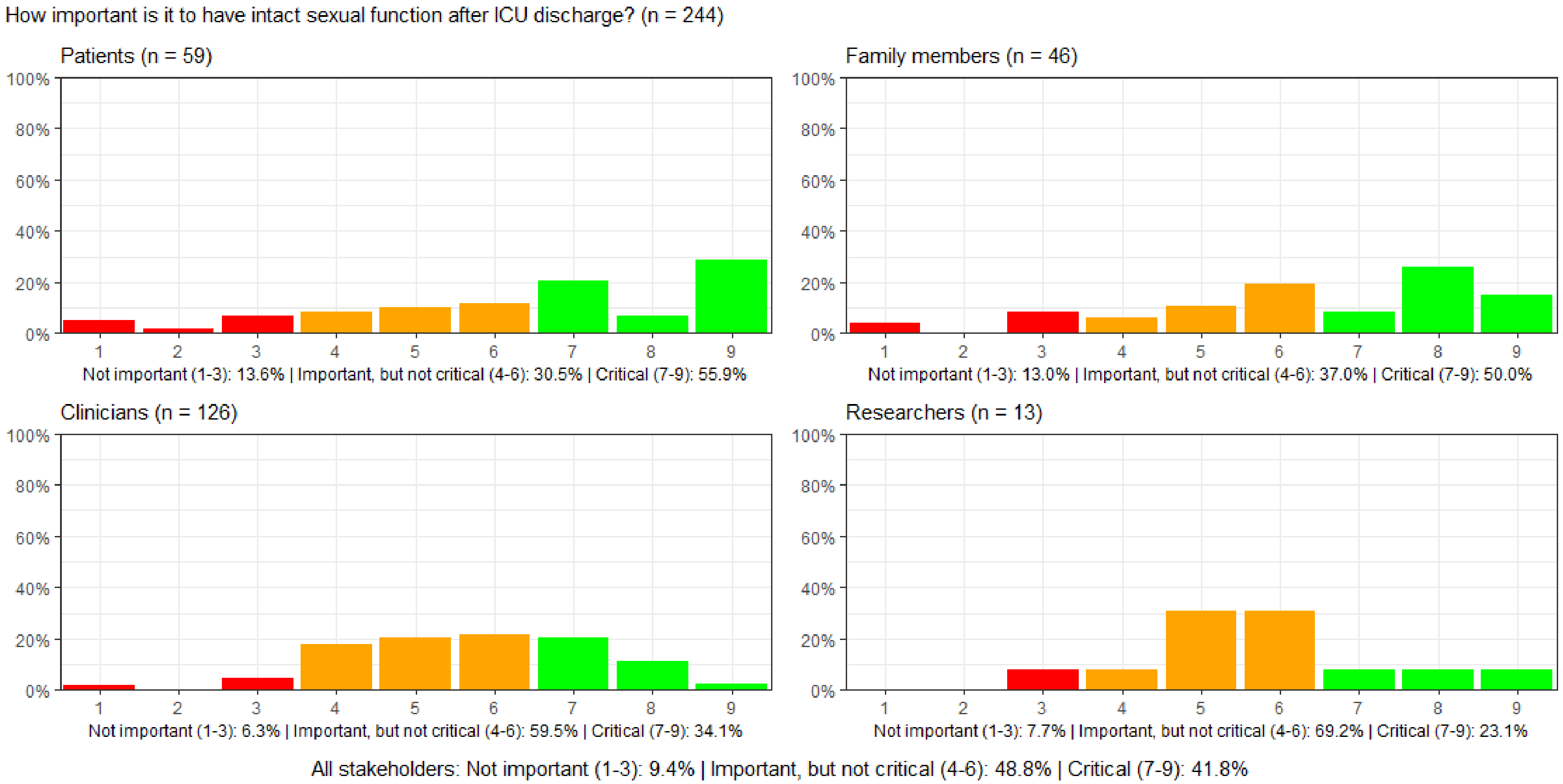

#### Figure S43: How important is it to have preserved senses after ICU discharge?

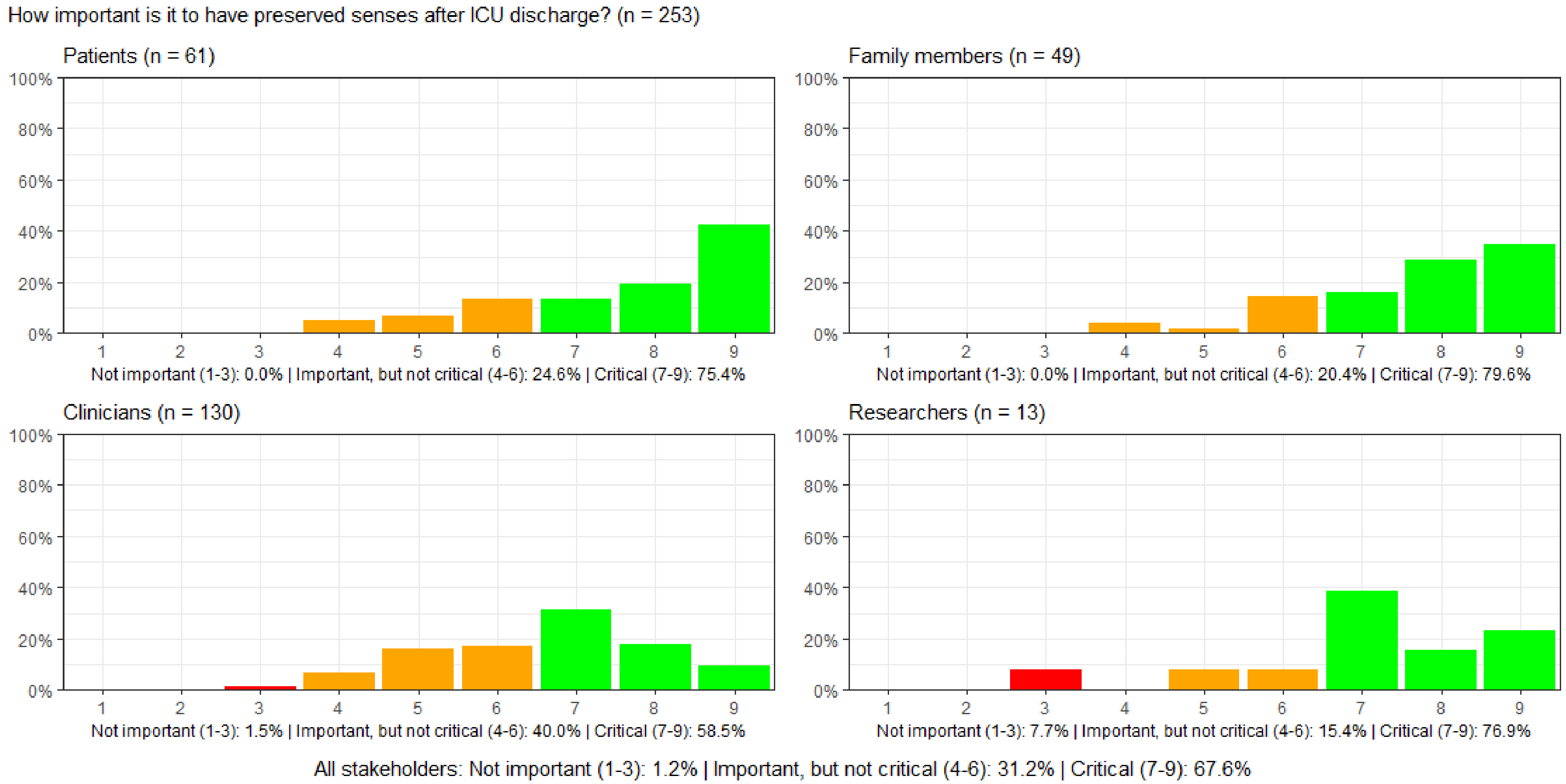

#### Figure S44: How important is it to have physical strength to perform acitvities of daily living after discharge from the hospital?

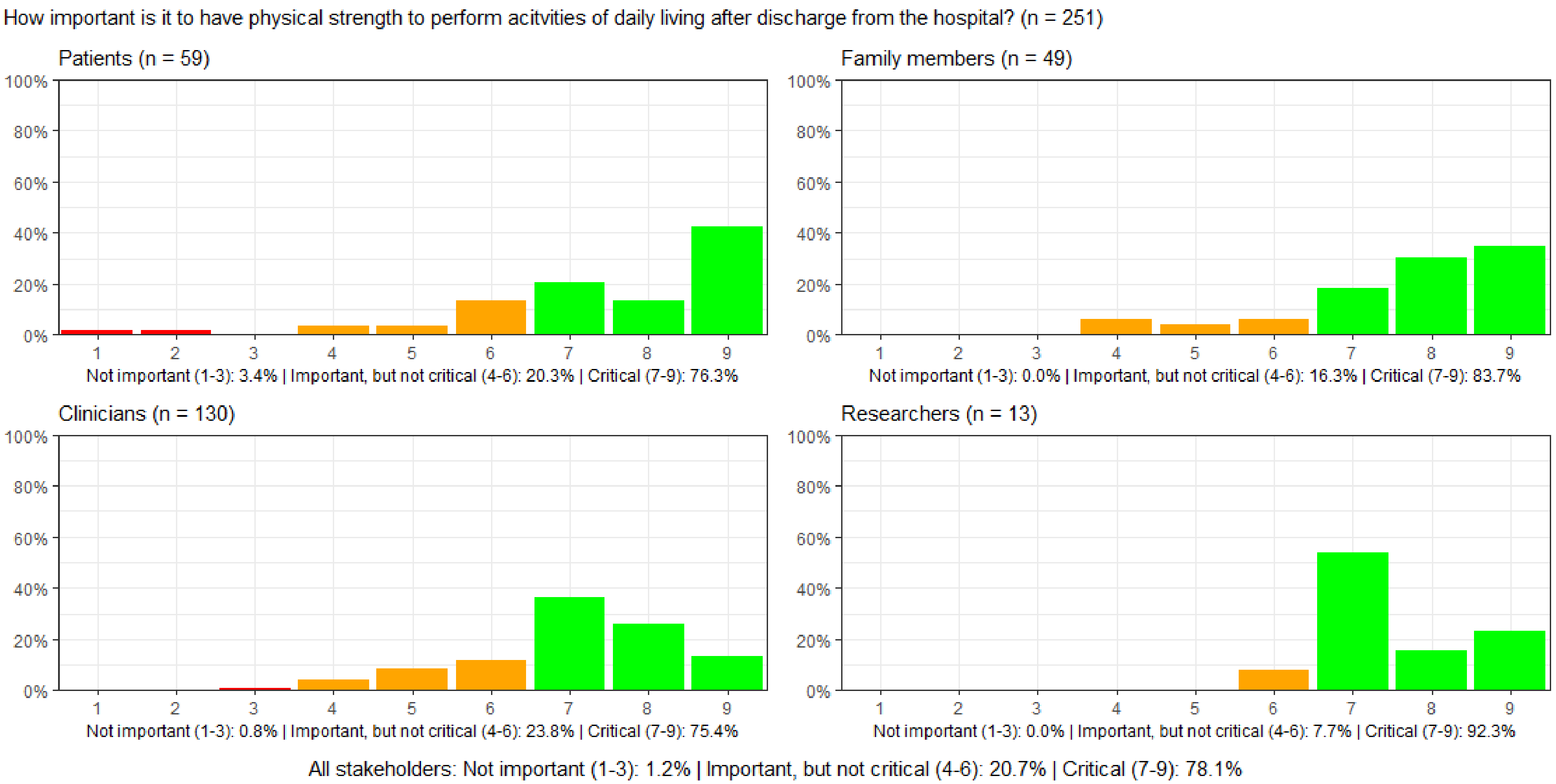

#### Figure S45: How important is it to maintain mental function after ICU discharge?

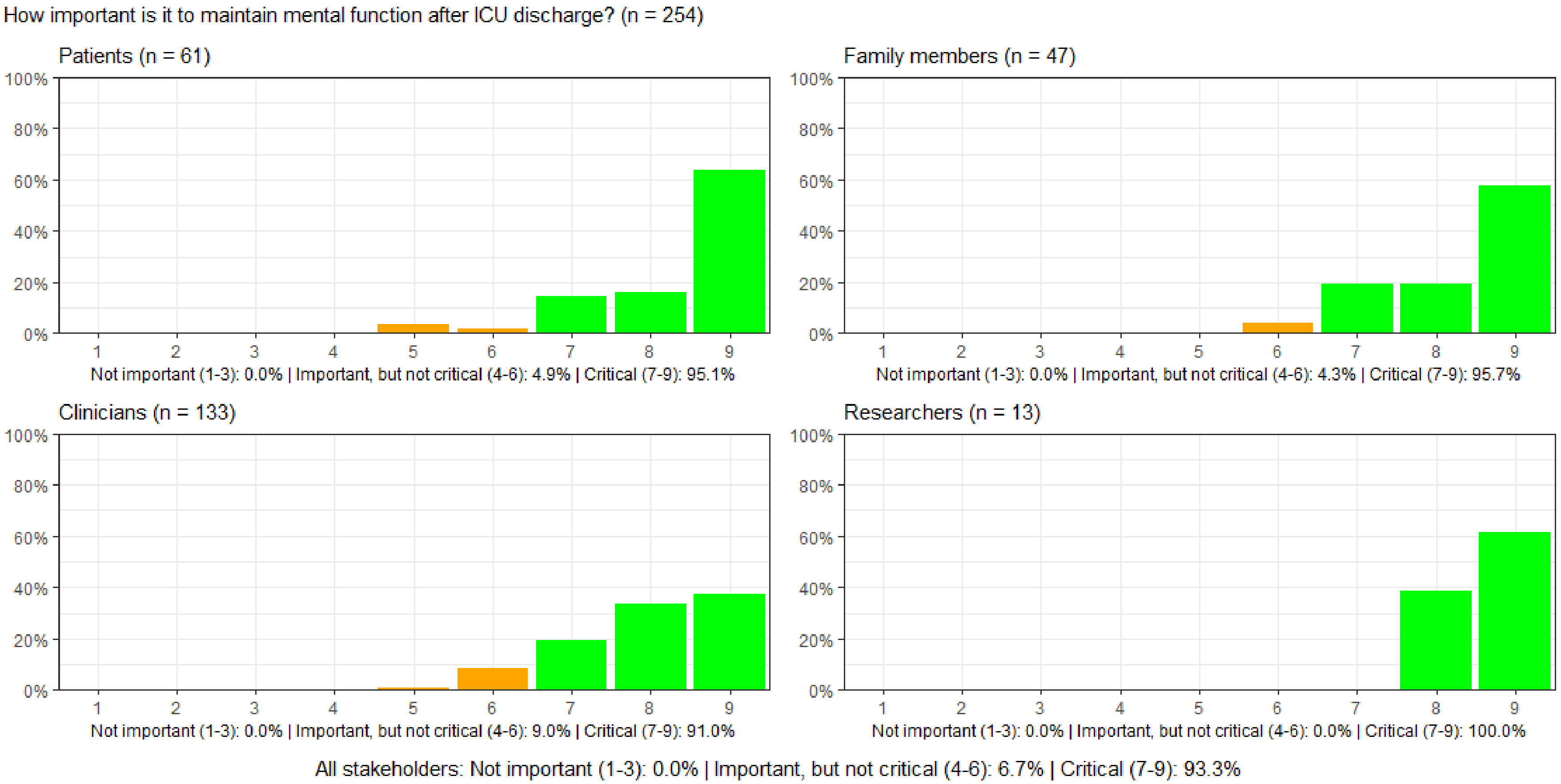

#### Figure S46: How important is it to record whether patients experience loneliness after ICU discharge?

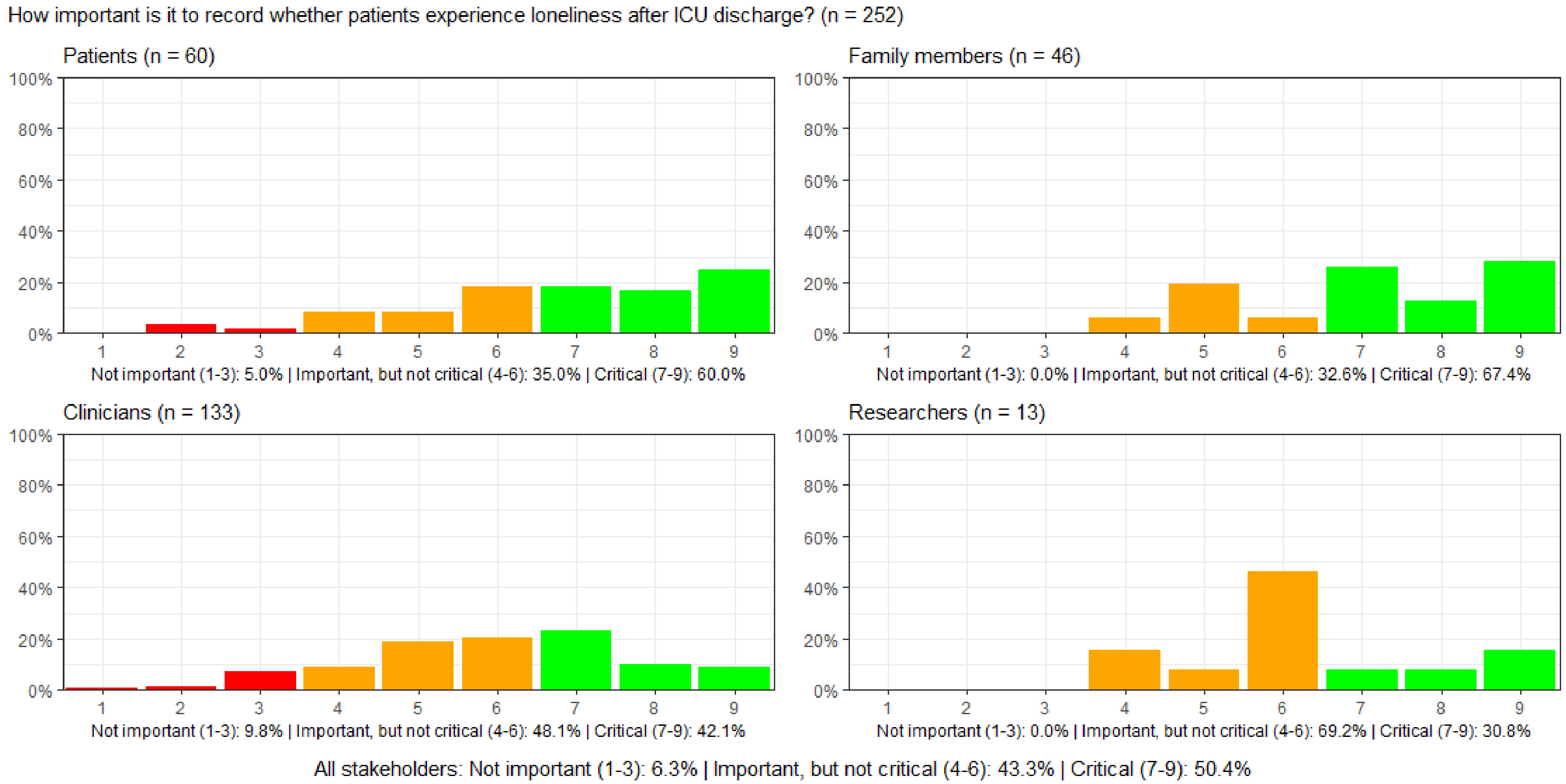

#### Figure S47: How important is it to record whether patients experience a post-traumatic stress reaction after ICU discharge?

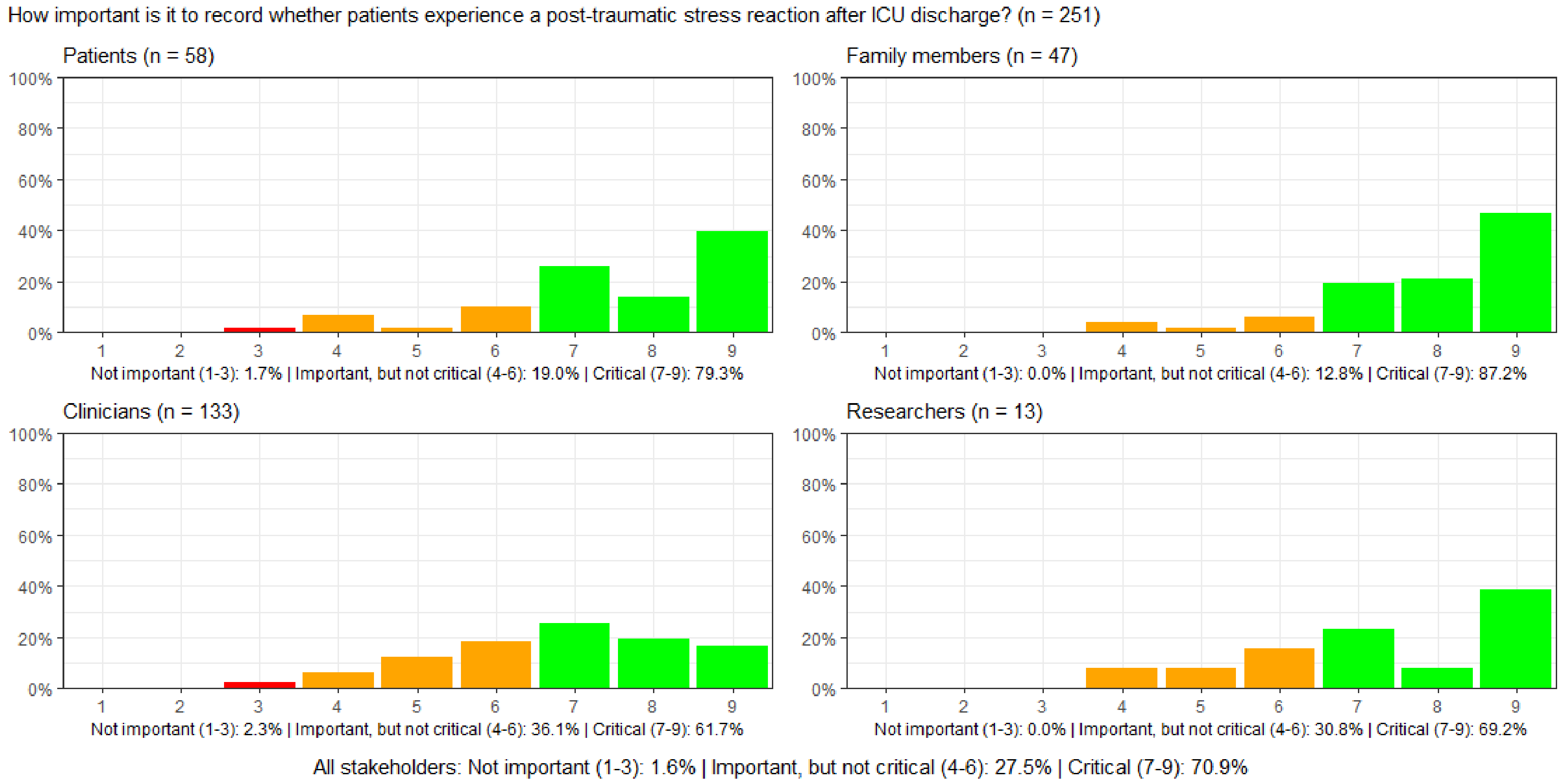

#### Figure S48: How important is it to record whether patients experience anxiety after ICU discharge?

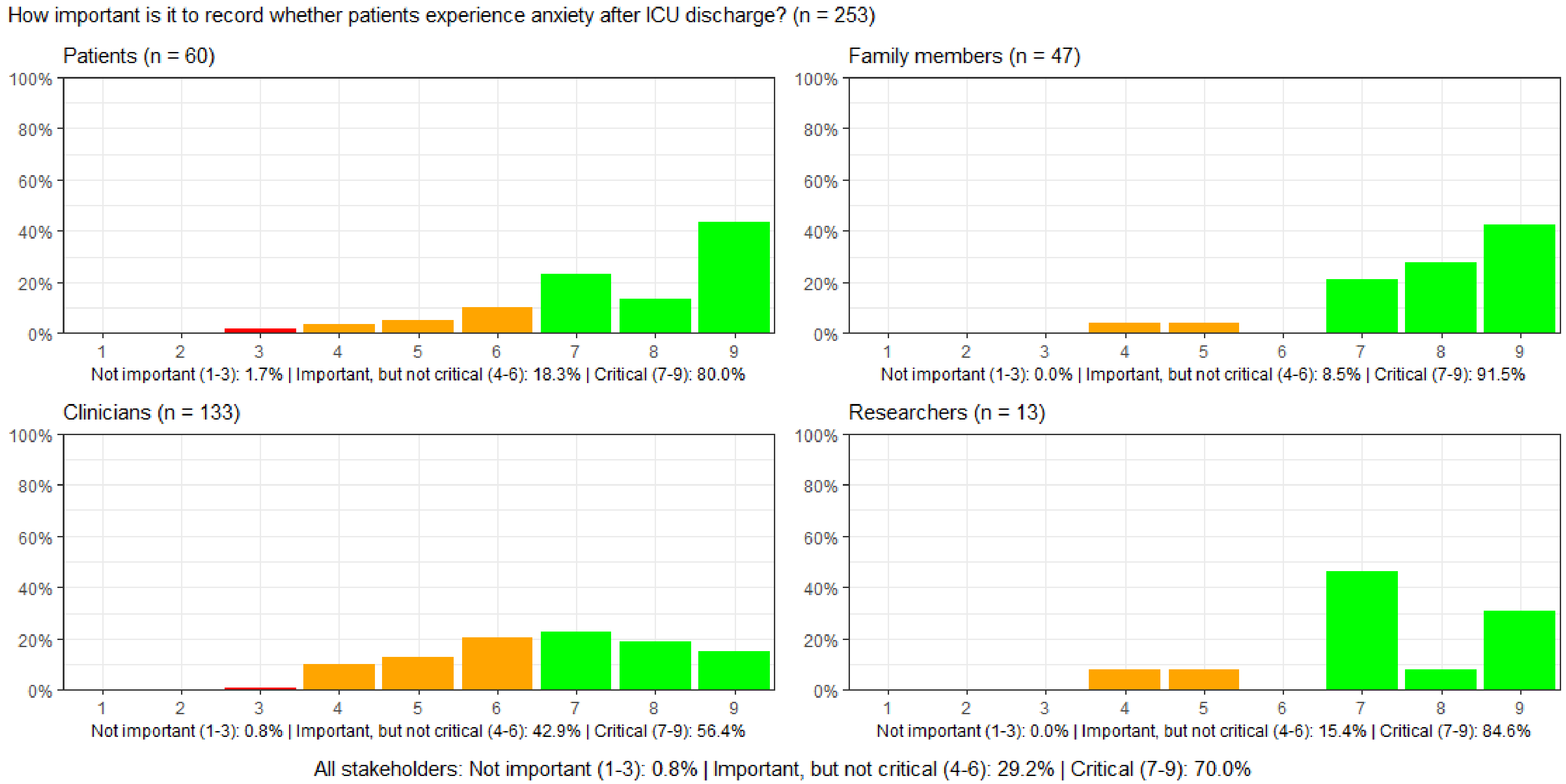

#### Figure S49: How important is it to record whether patients experience depression after ICU discharge?

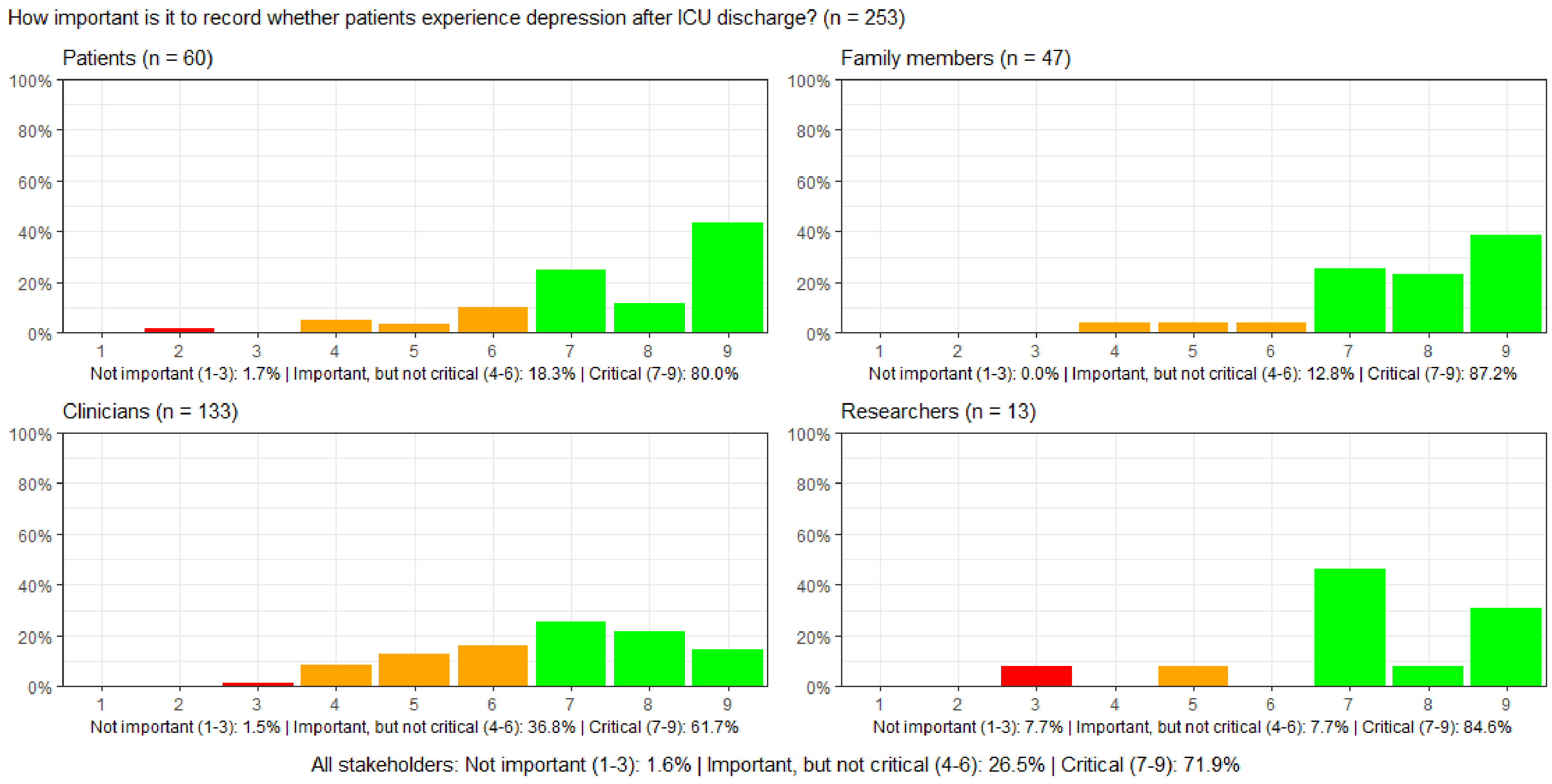

**Figure S50: How important is it to record whether patients experience nightmares after ICU discharge?**

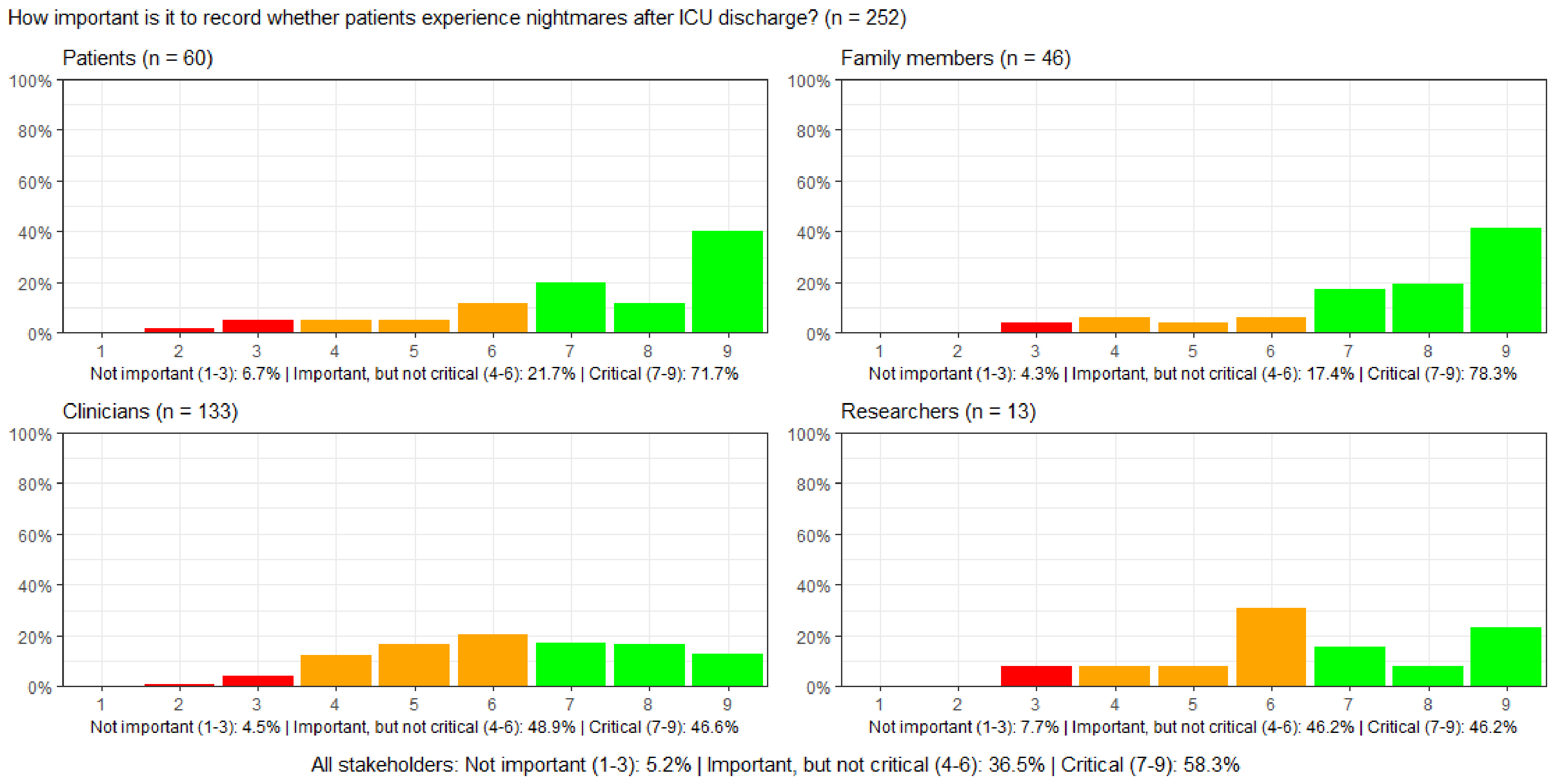

### Figure S51-S100. Graphs for the second survey round

#### Figure S51: How important is it to record ICU survival?

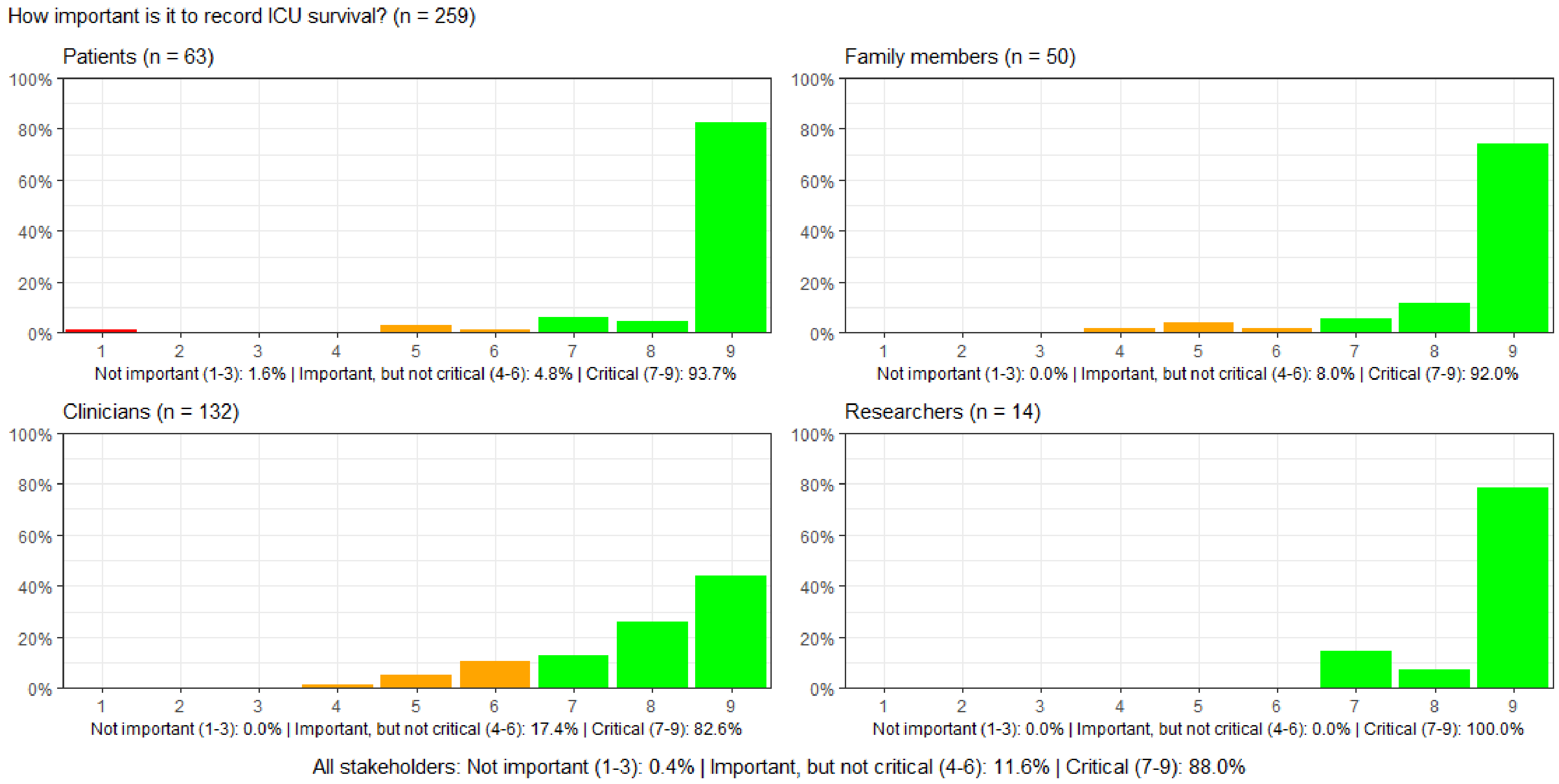

#### Figure S52: How important is it to record the ICU length of stay?

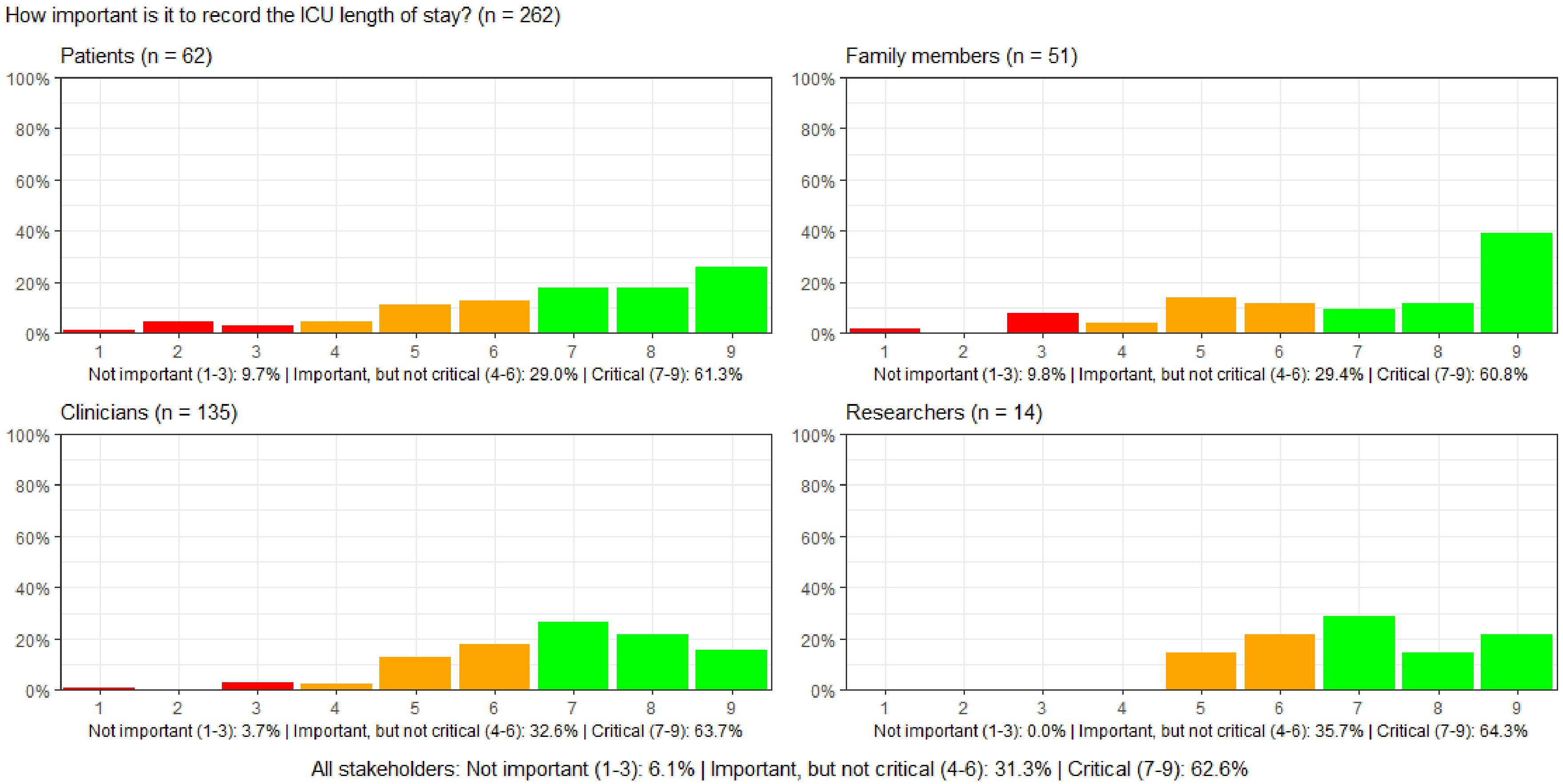

#### Figure S53: How important is it to record the hospital length of stay?

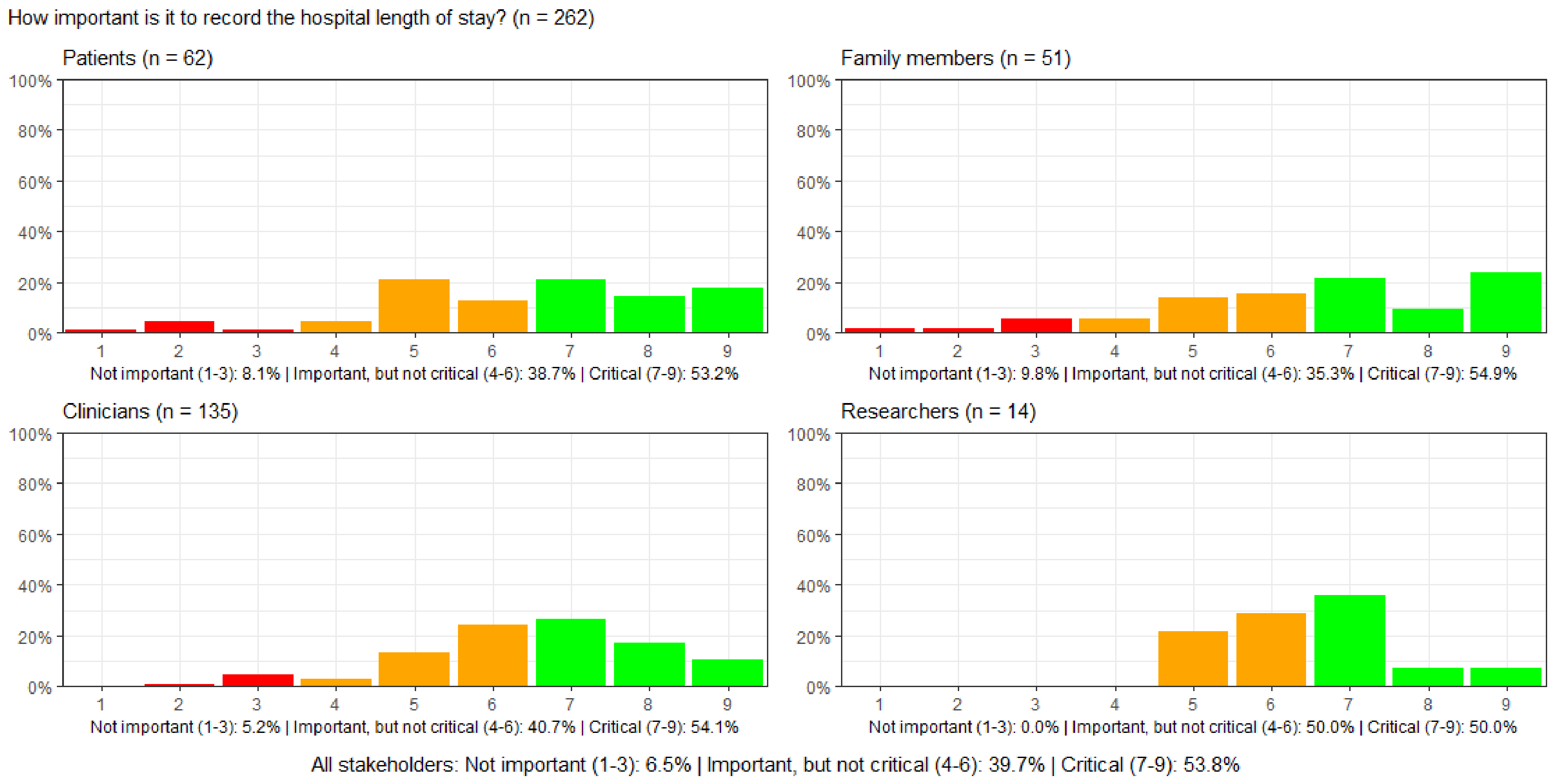

#### Figure S54: How important is it to record the ICU readmission rate during a hospital stay?

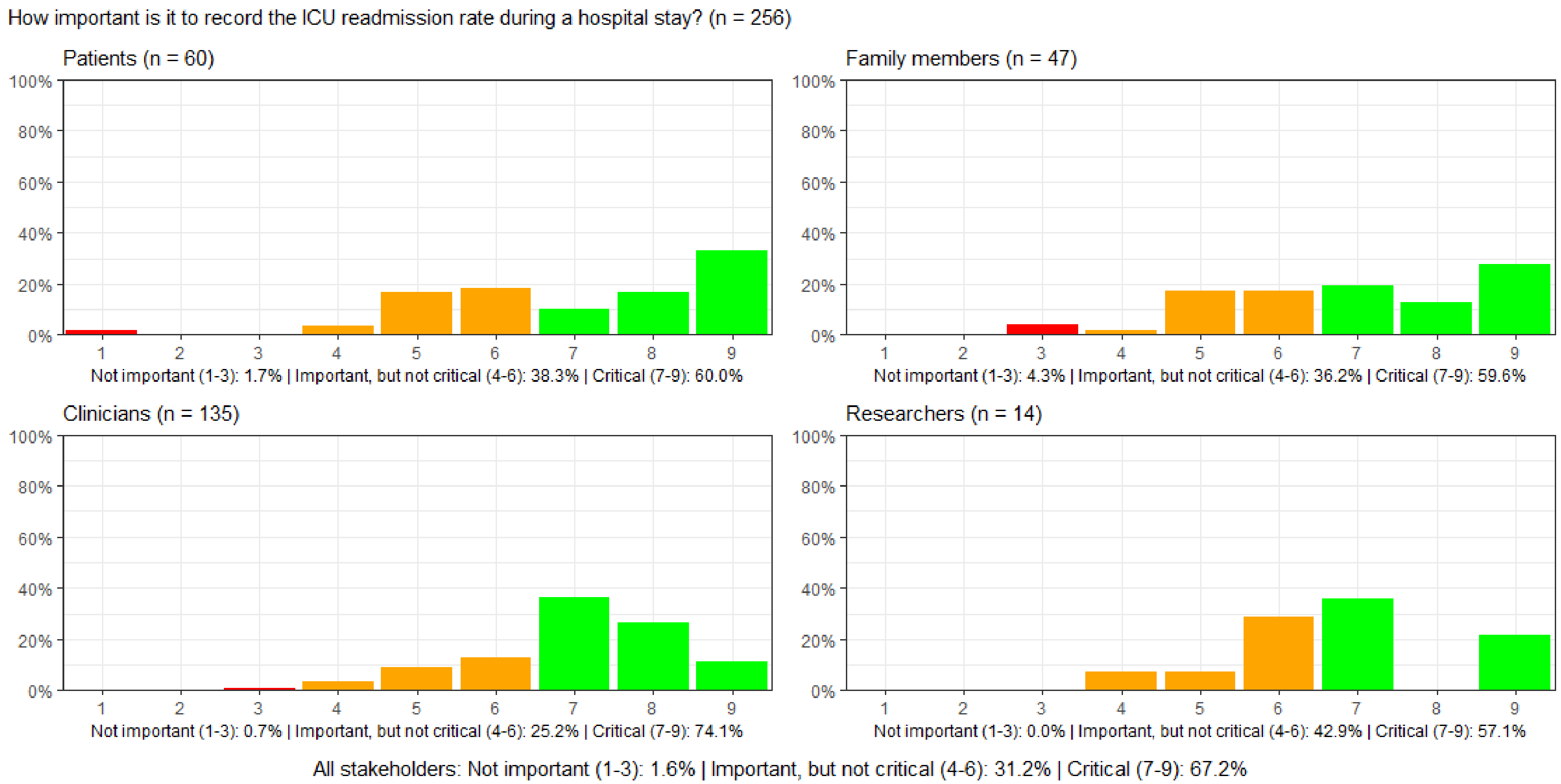

#### Figure S55: How important is it to record hospital readmission?

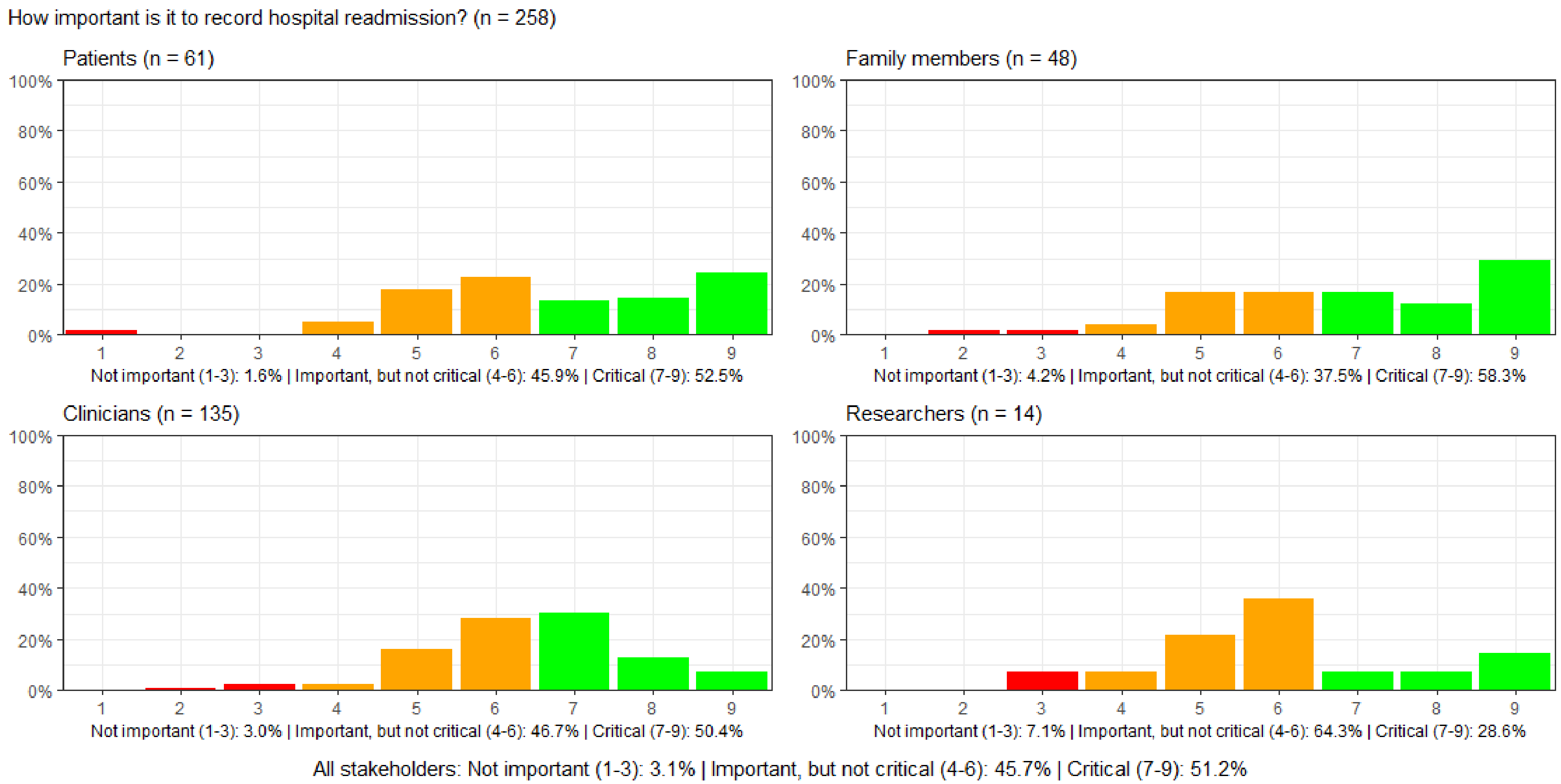

#### Figure S56: How important is it to record patients' overall well-being?

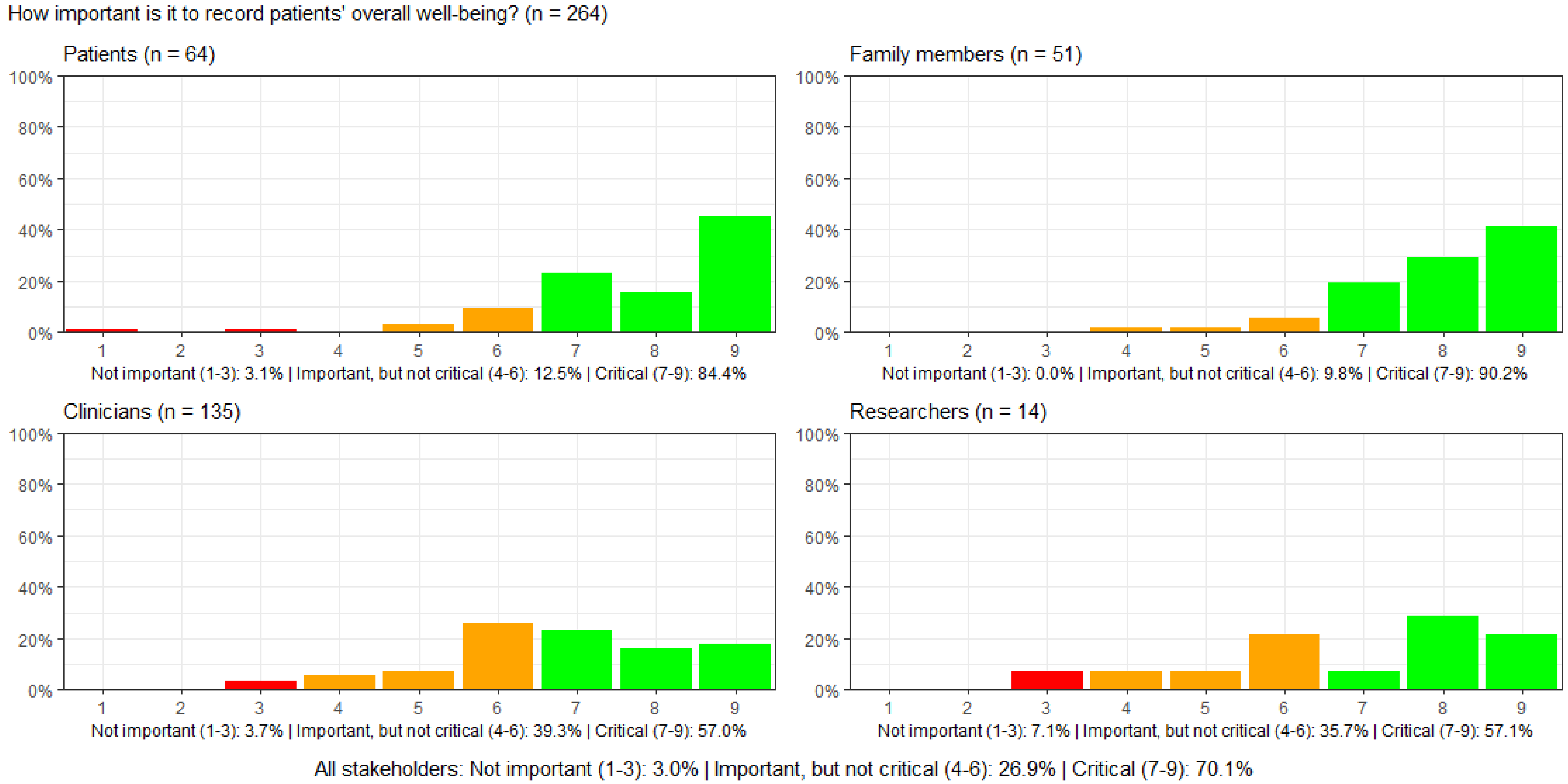

#### Figure S57: How important is it to measure the health economic consequences of a particular treatment in ICU?

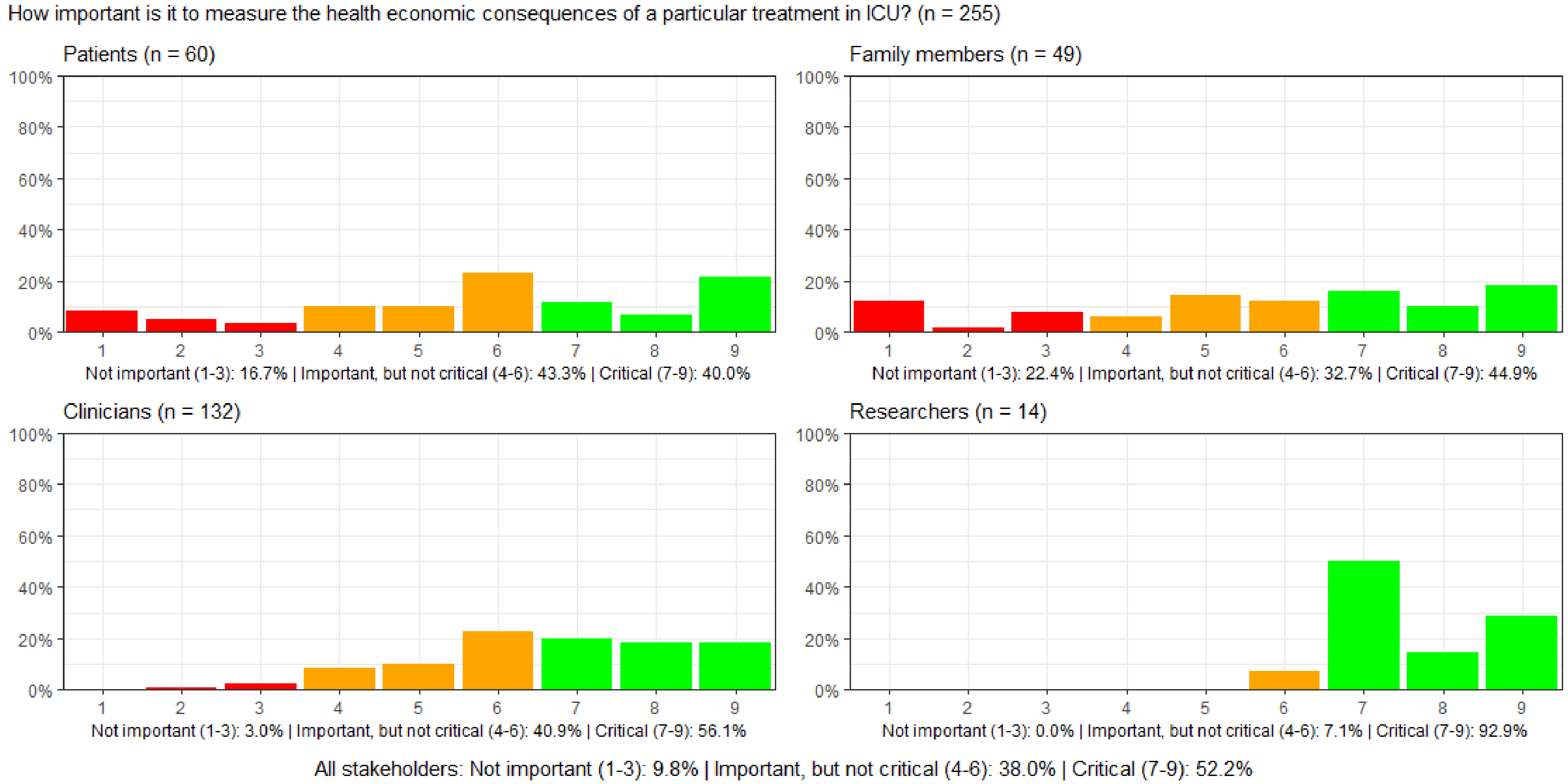

#### Figure S58: How important is it to measure the environmental consequences of a particular treatment in ICU?

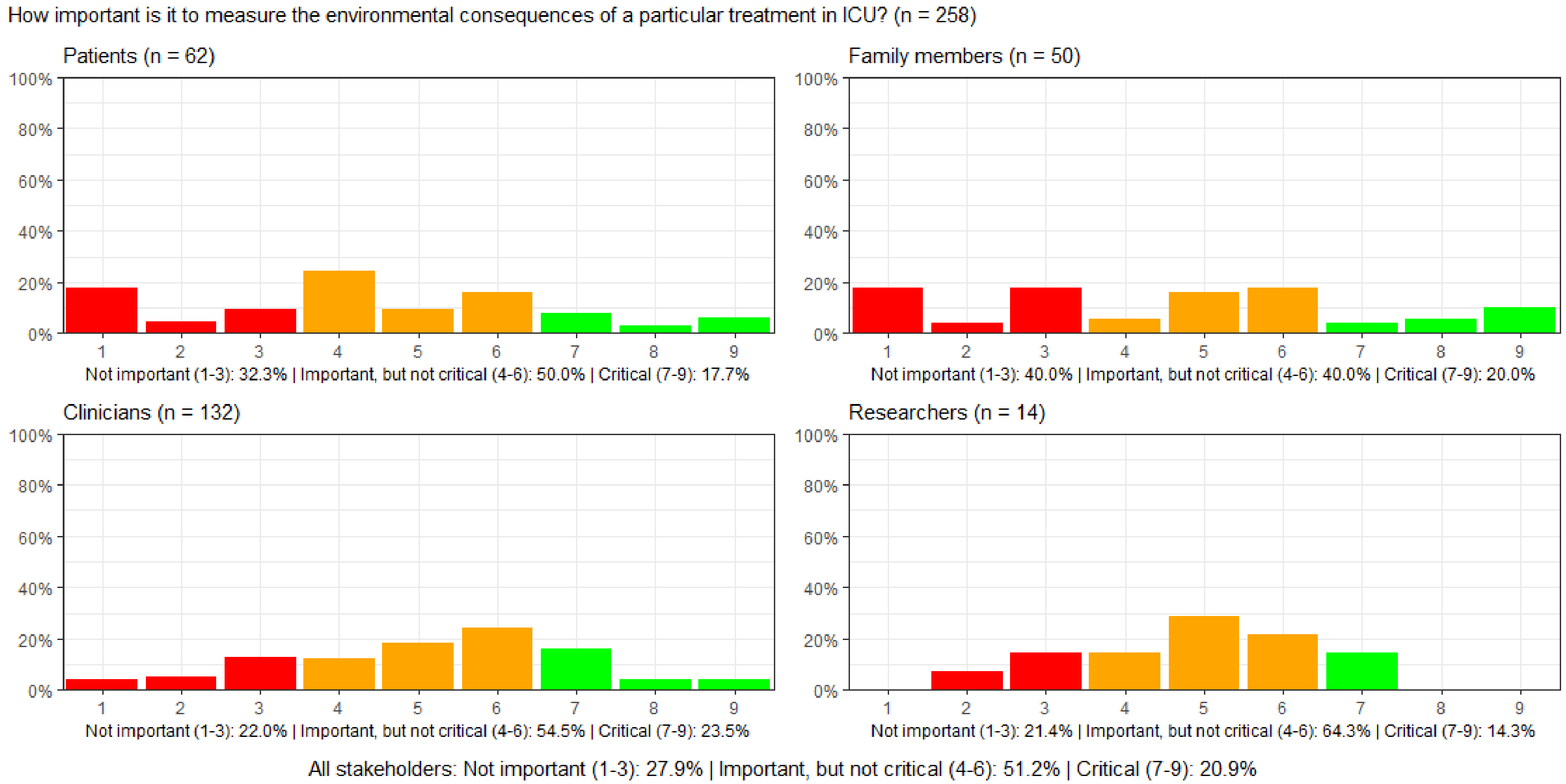

#### Figure S59: How important is it to measure whether blood values are within normal limits while in ICU?

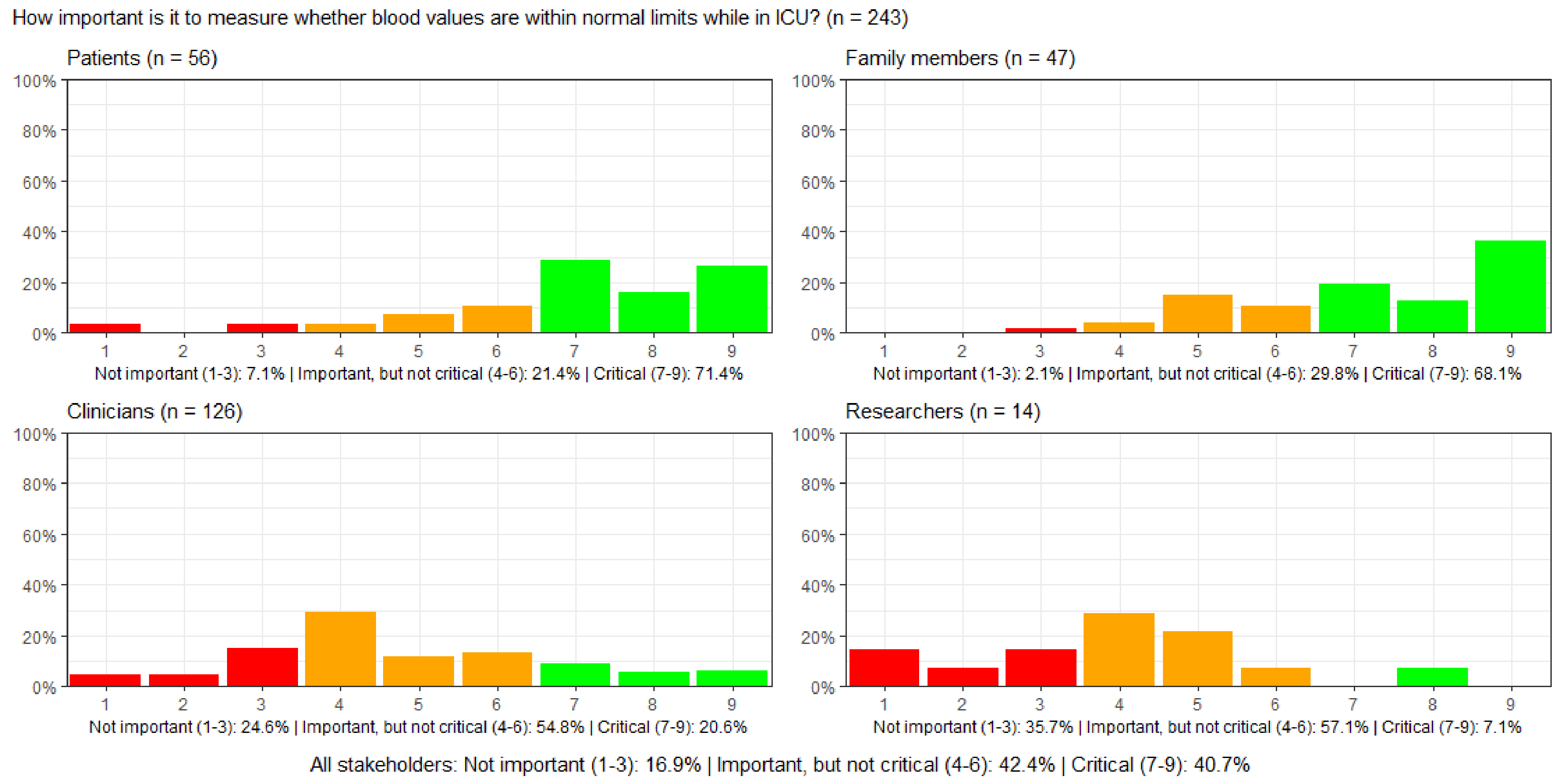

#### Figure S60: How important is it to record whether vital signs are within normal limits while in ICU?

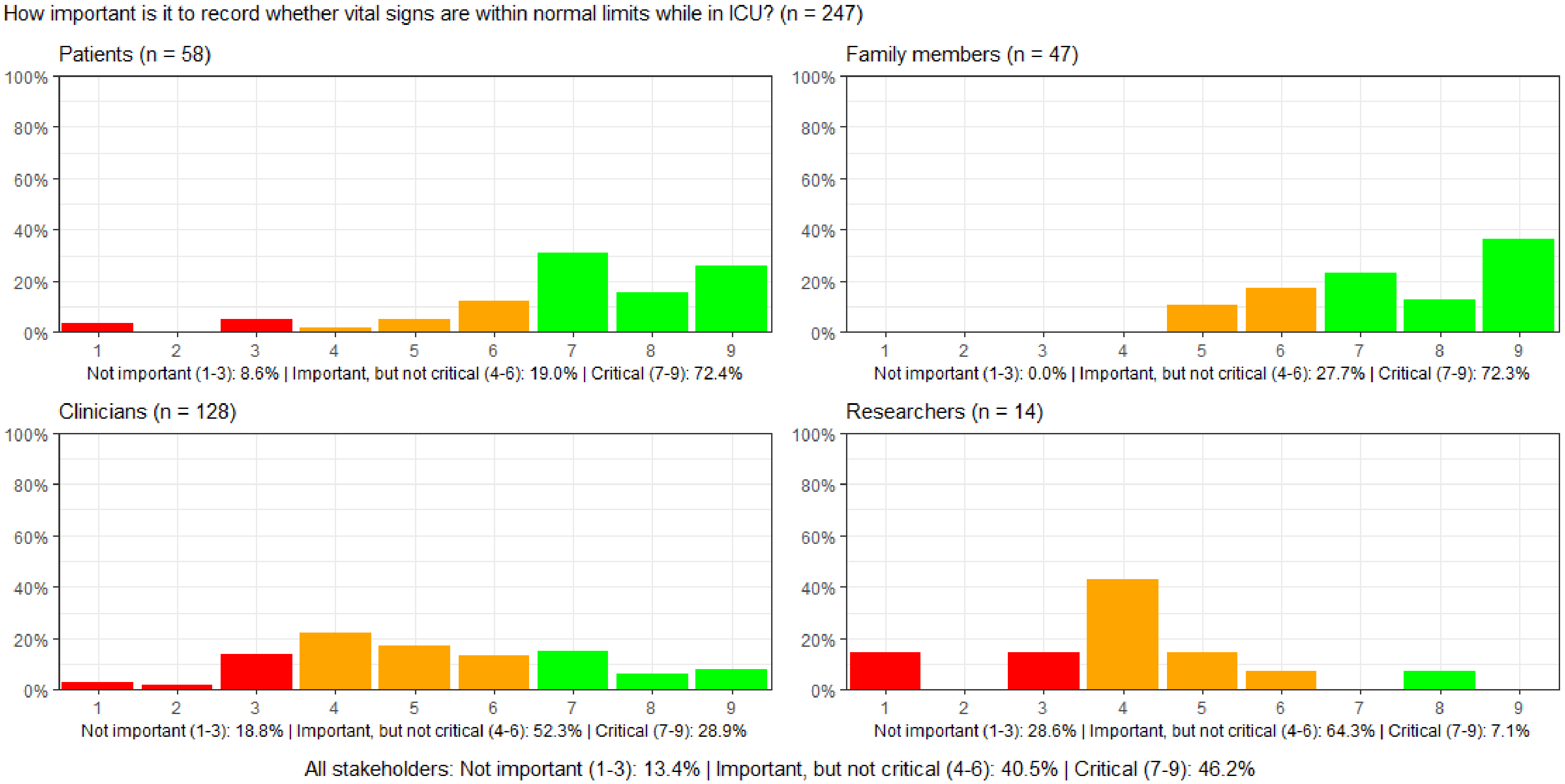

#### Figure S61: How important is it to record the duration of mechanical ventilation while in ICU?

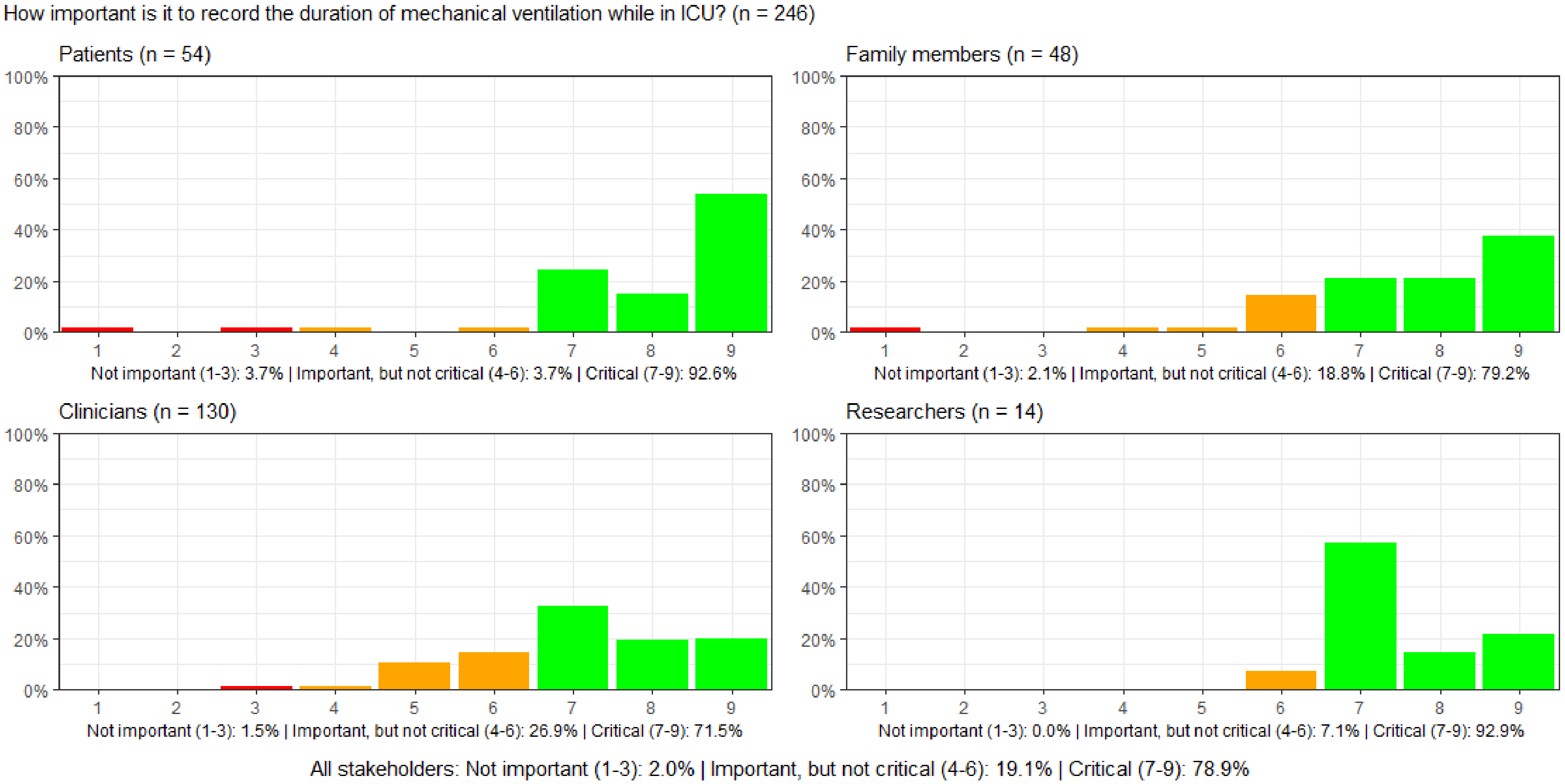

#### Figure S62: How important is it to record the duration of pharmacologic circulatory support while in ICU?

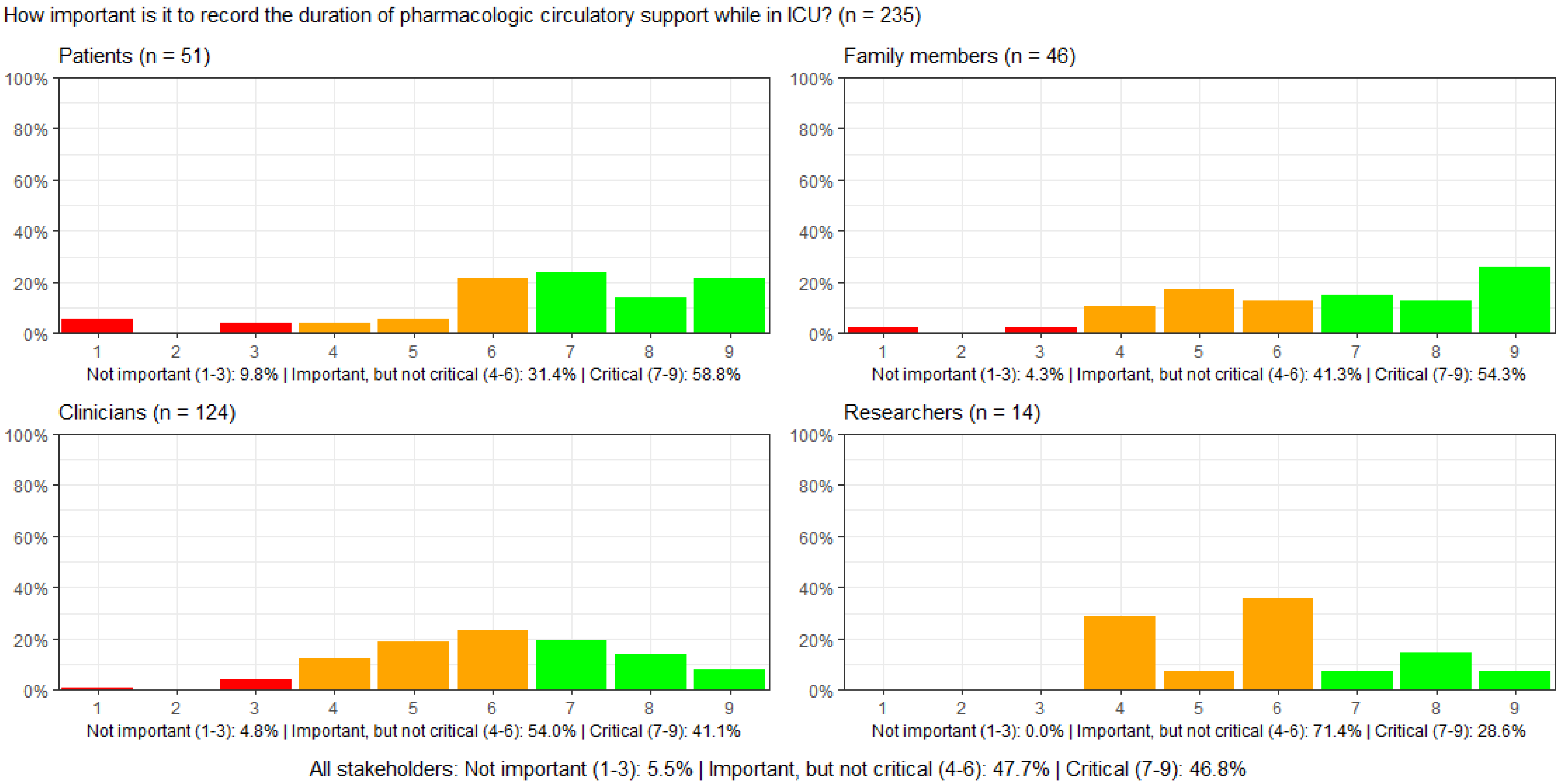

#### Figure S63: How important is it to record the duration of dialysis while in ICU?

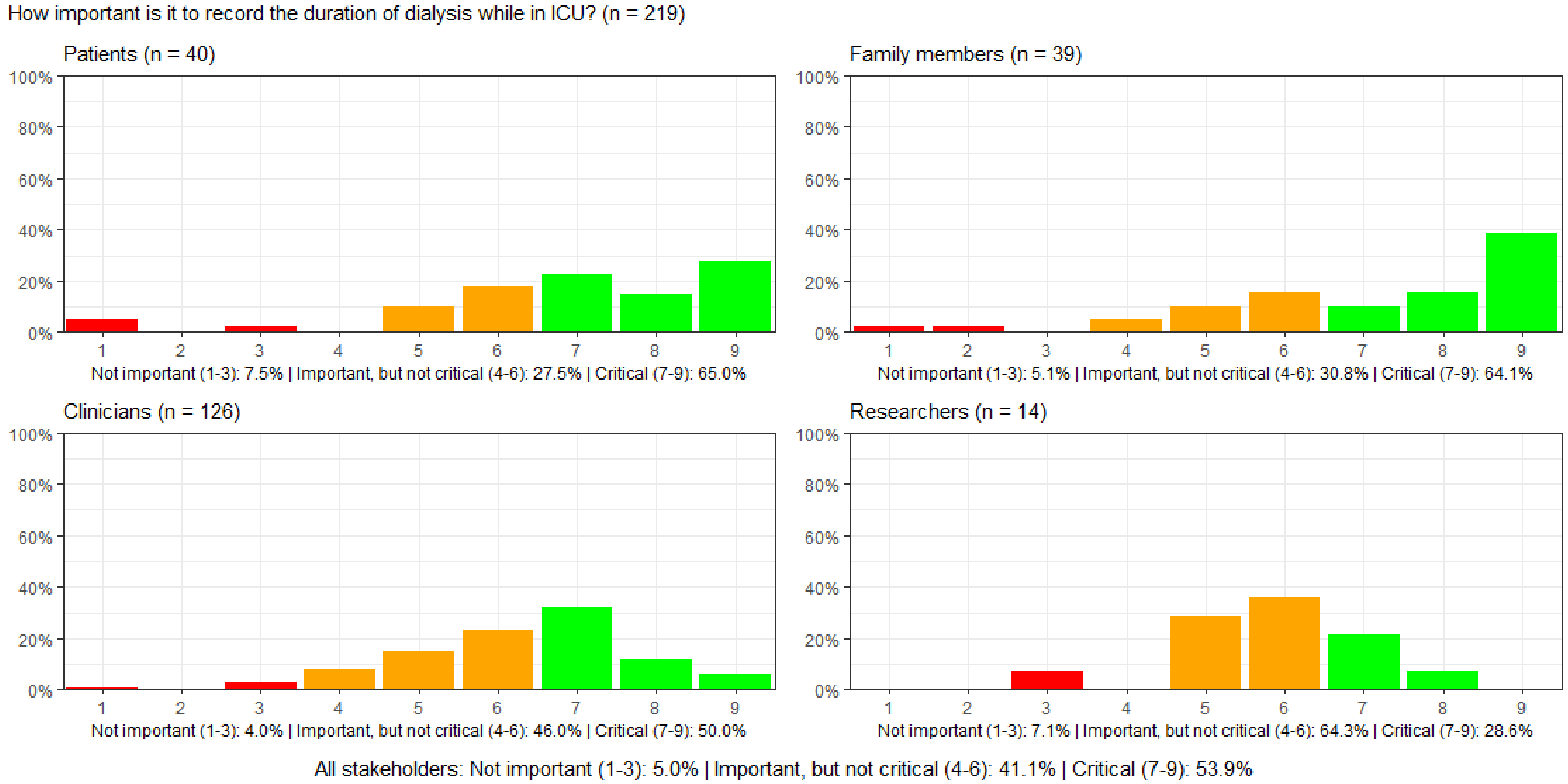

#### Figure S64: How important is it to record the duration of sedation while in ICU?

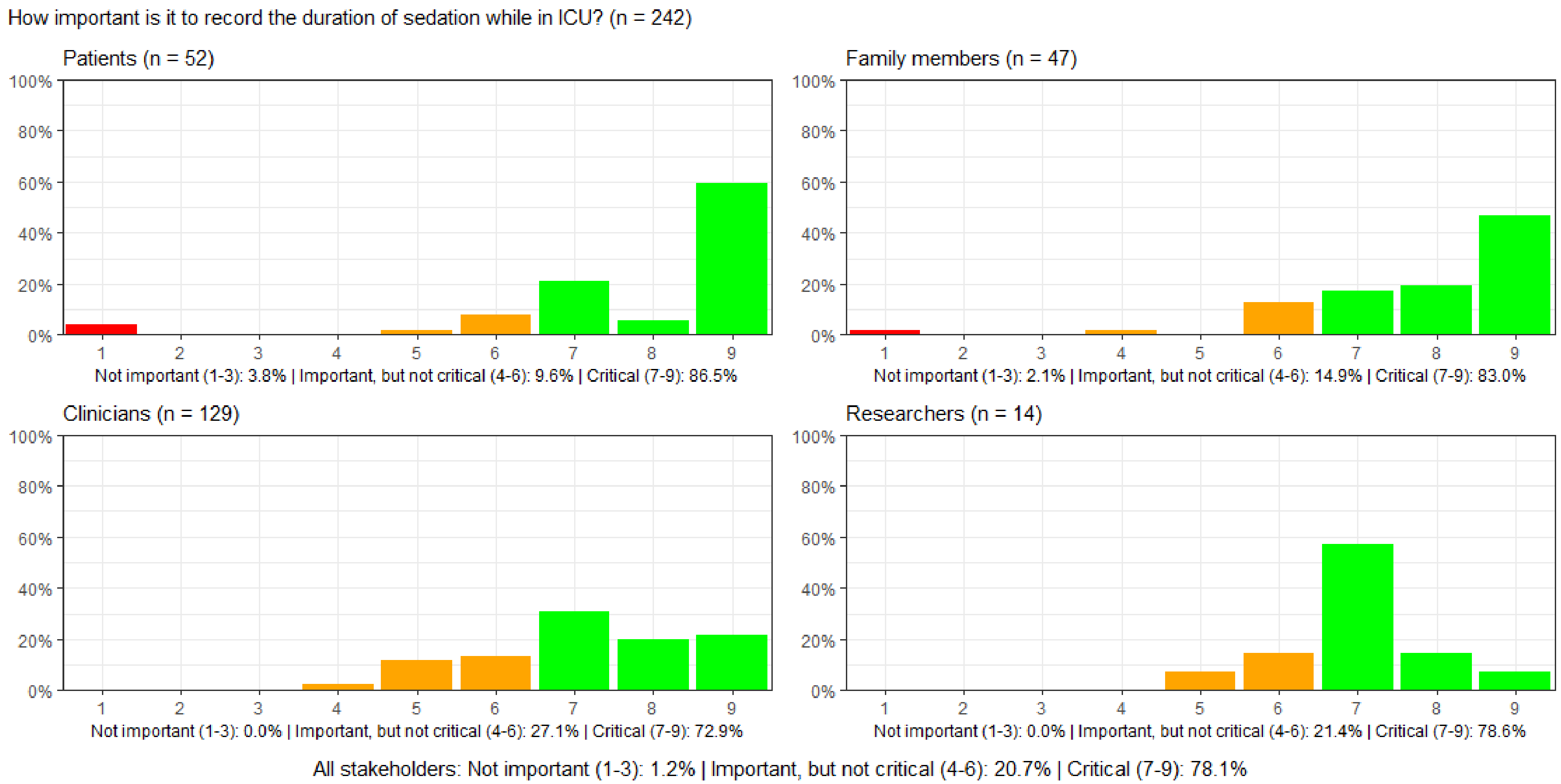

#### Figure S65: How important is it to record the duration of delirium while in ICU?

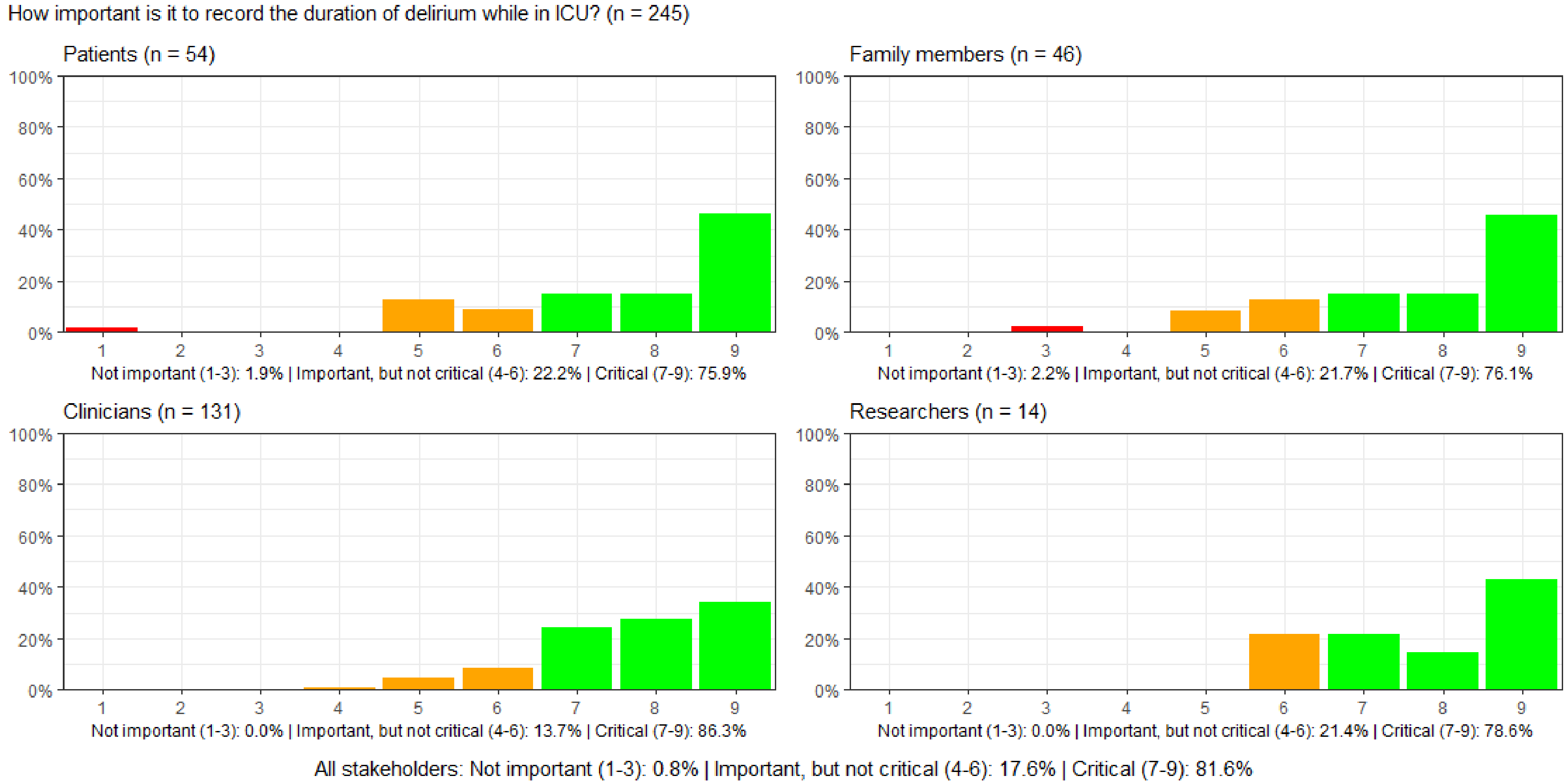

#### Figure S66: How important is it to record whether patients experience nausea and vomiting while in ICU?

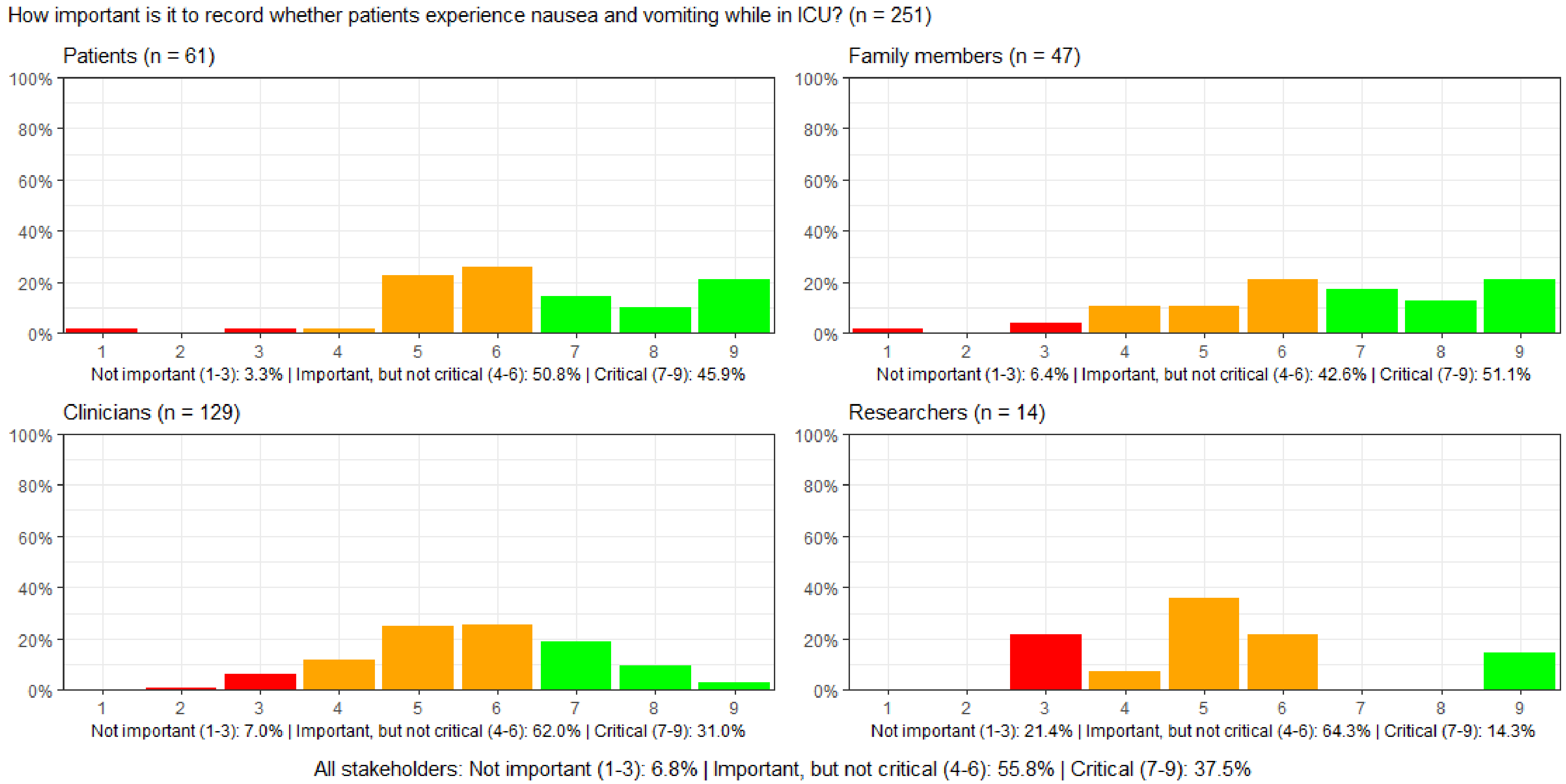

#### Figure S67: How important is it to record whether patients experience thirst while in ICU?

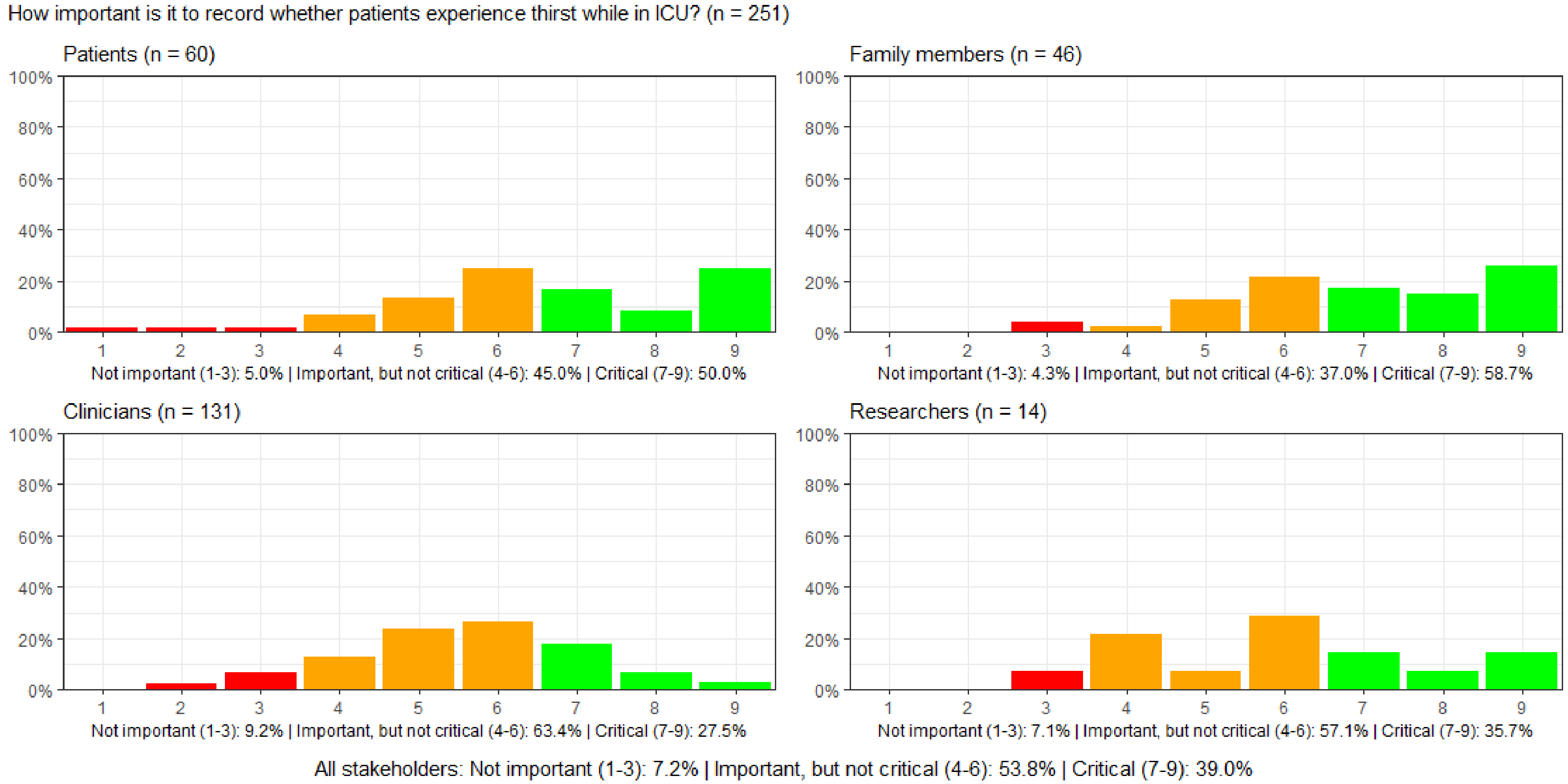

#### Figure S68: How important is it to record whether patients are short of breath while in ICU.

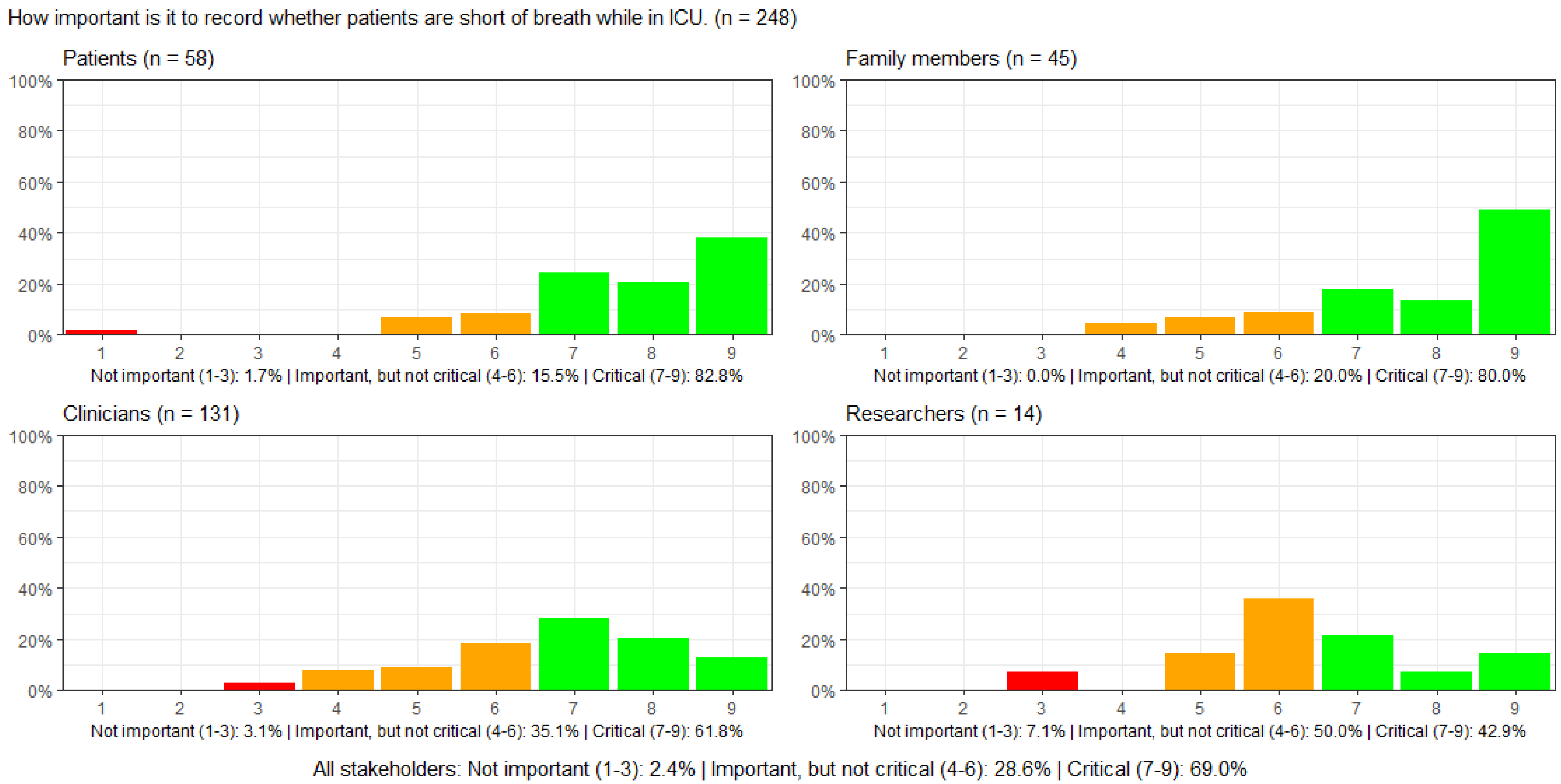

#### Figure S69: How important is it to record whether patients experience pain while in ICU?

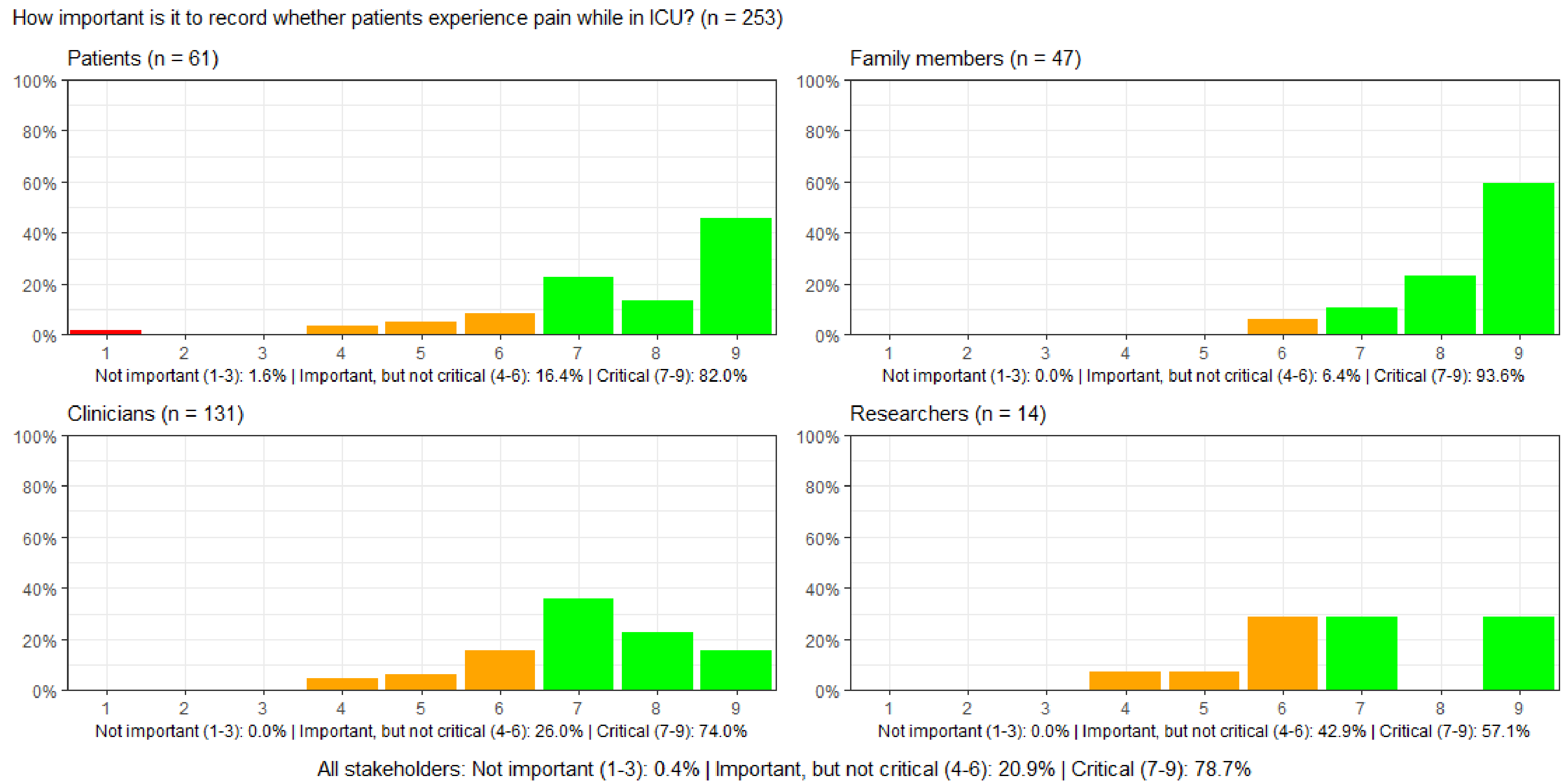

#### Figure S70: How important is it to record whether patients experience anxiety while in ICU?

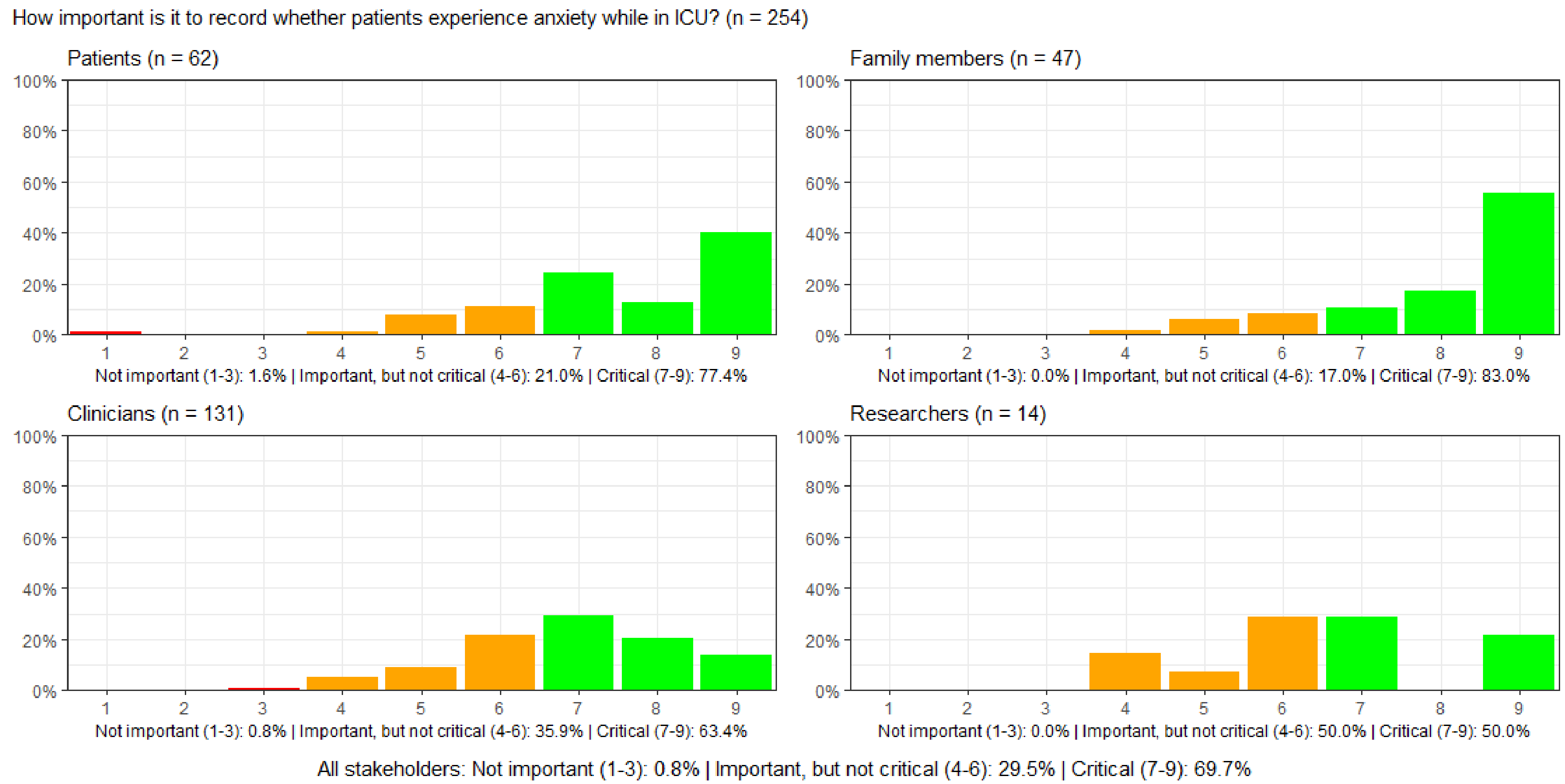

#### Figure S71: How important is it to record patients' sleep while in ICU?

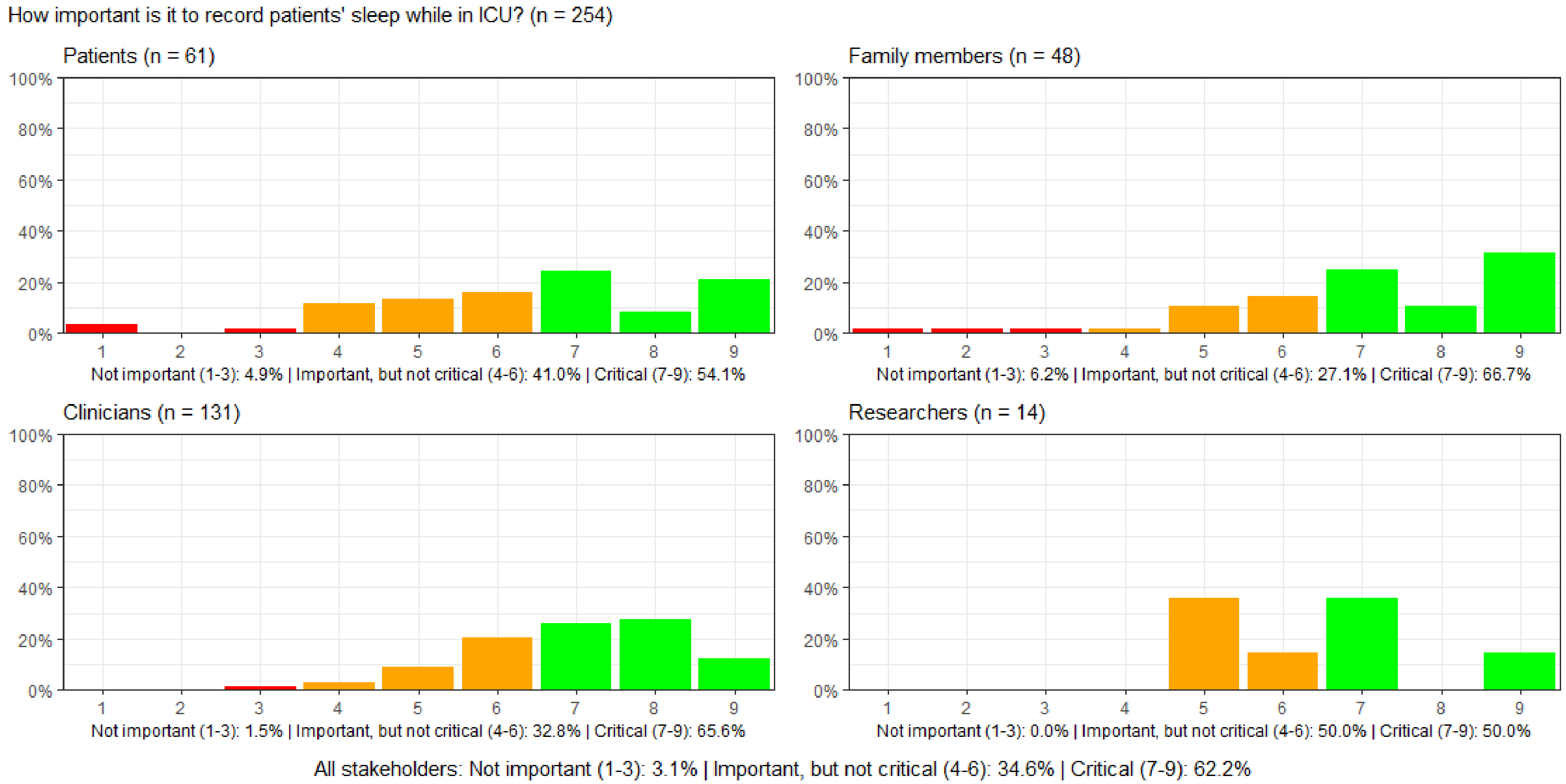

#### Figure S72: How important is it to record patients' ability to communicate while in ICU?

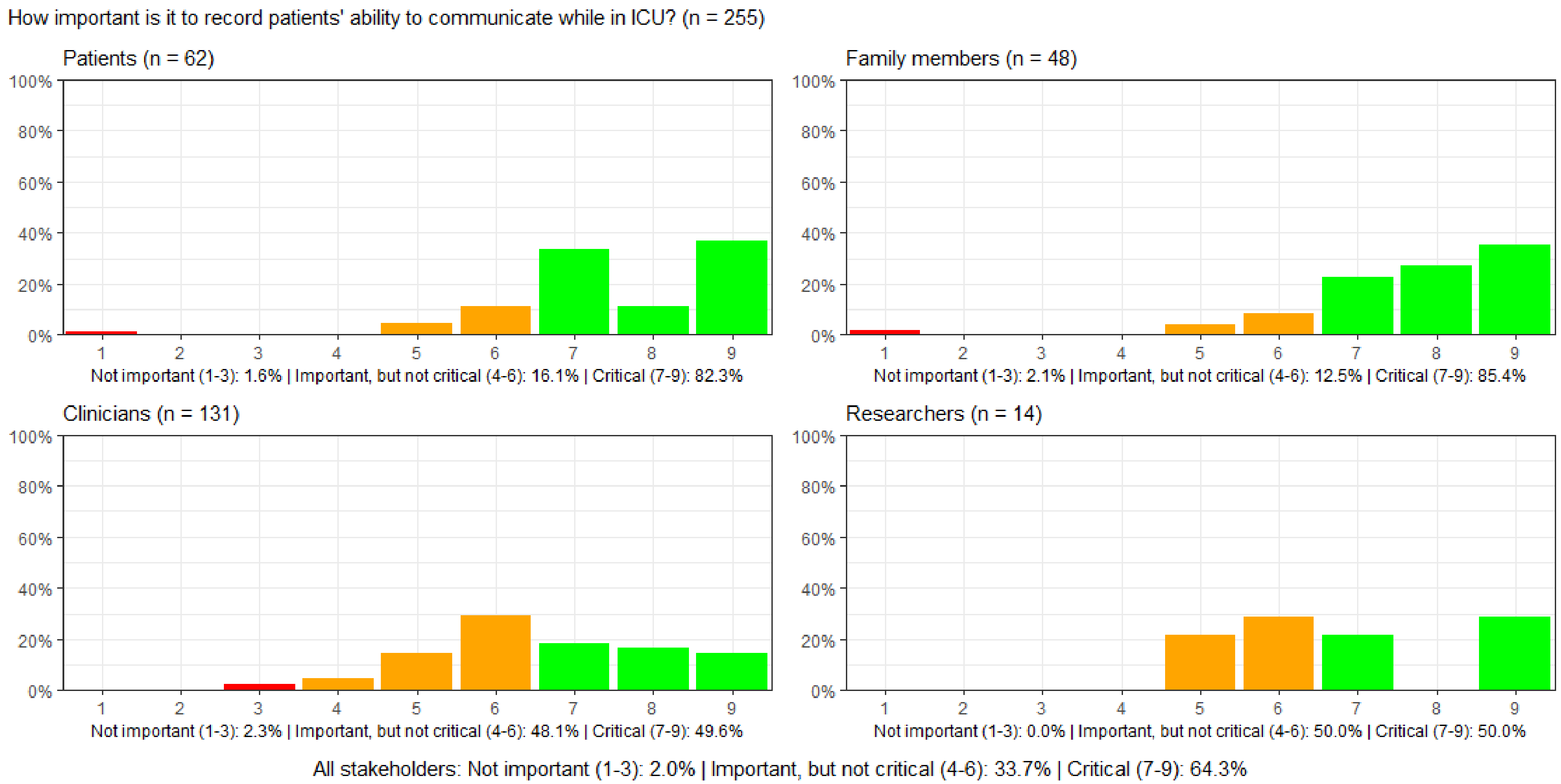

#### Figure S73: How important is it to record patients' discharge location after hospitalization?

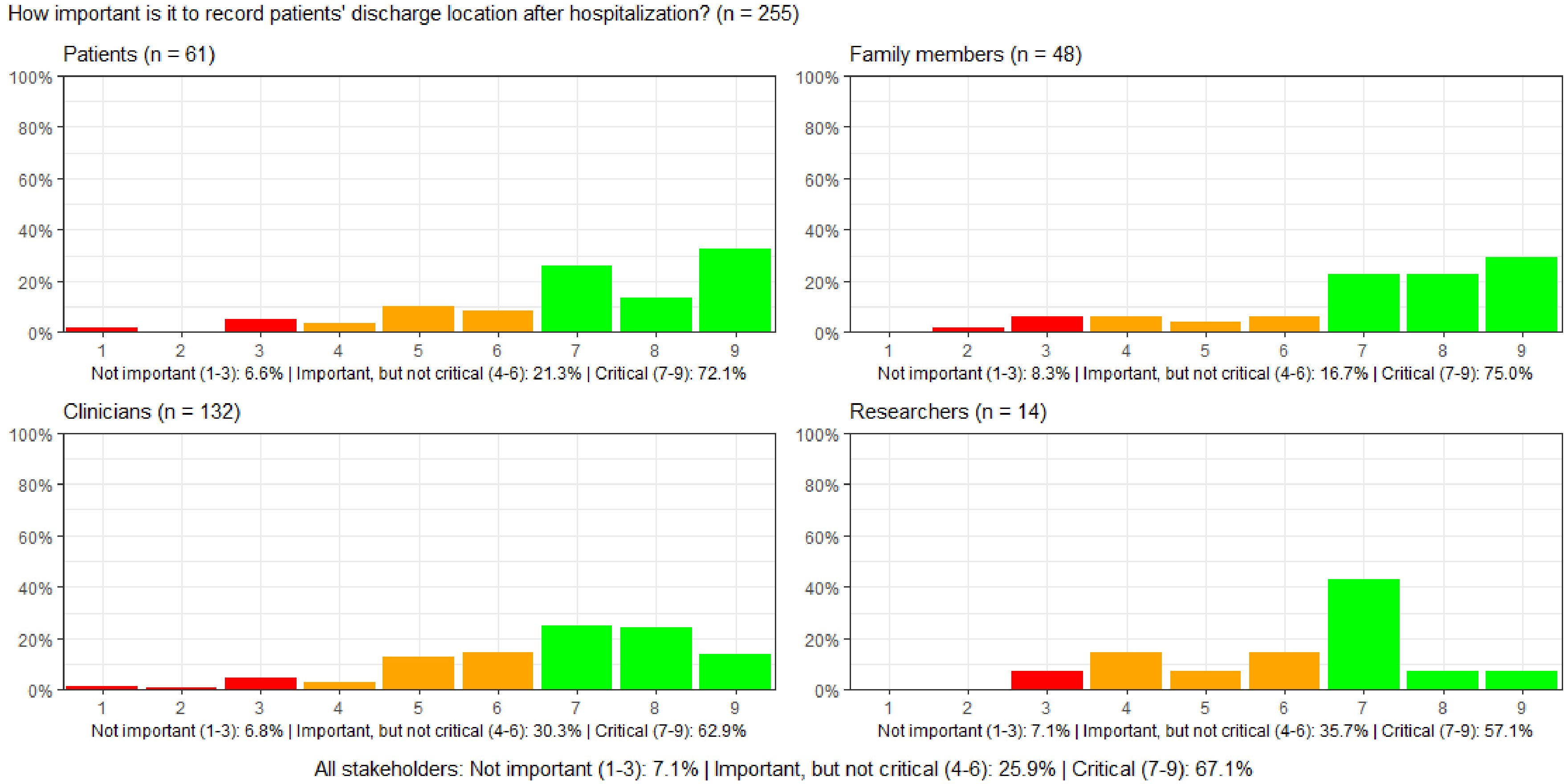

**Figure S74: How important is it to record patients' out-patient or general practitioner visits related to the long-term effects of intensive care?**

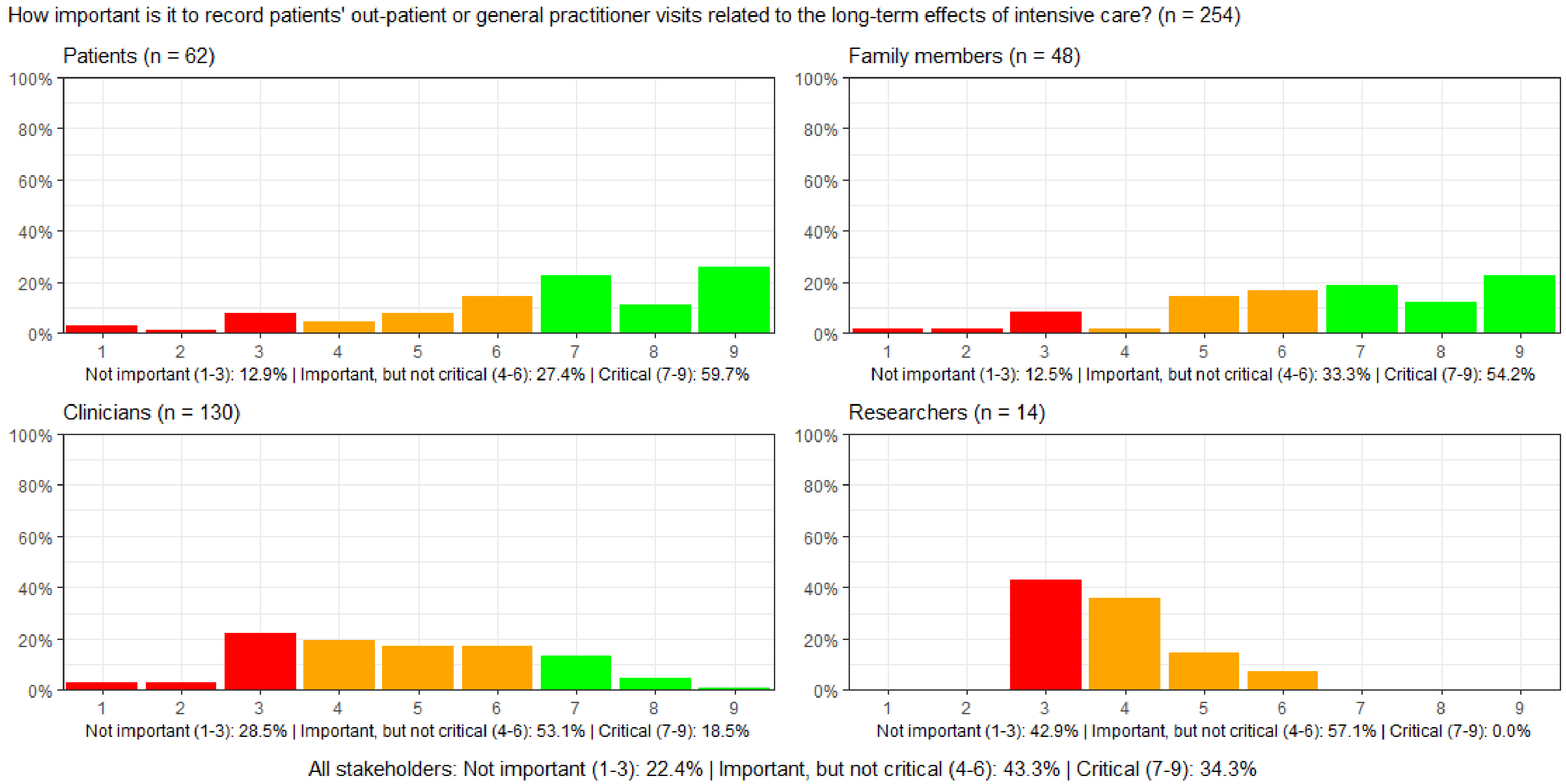

#### Figure S75: How important is it to record whether new medications are prescribed for the patient after ICU discharge?

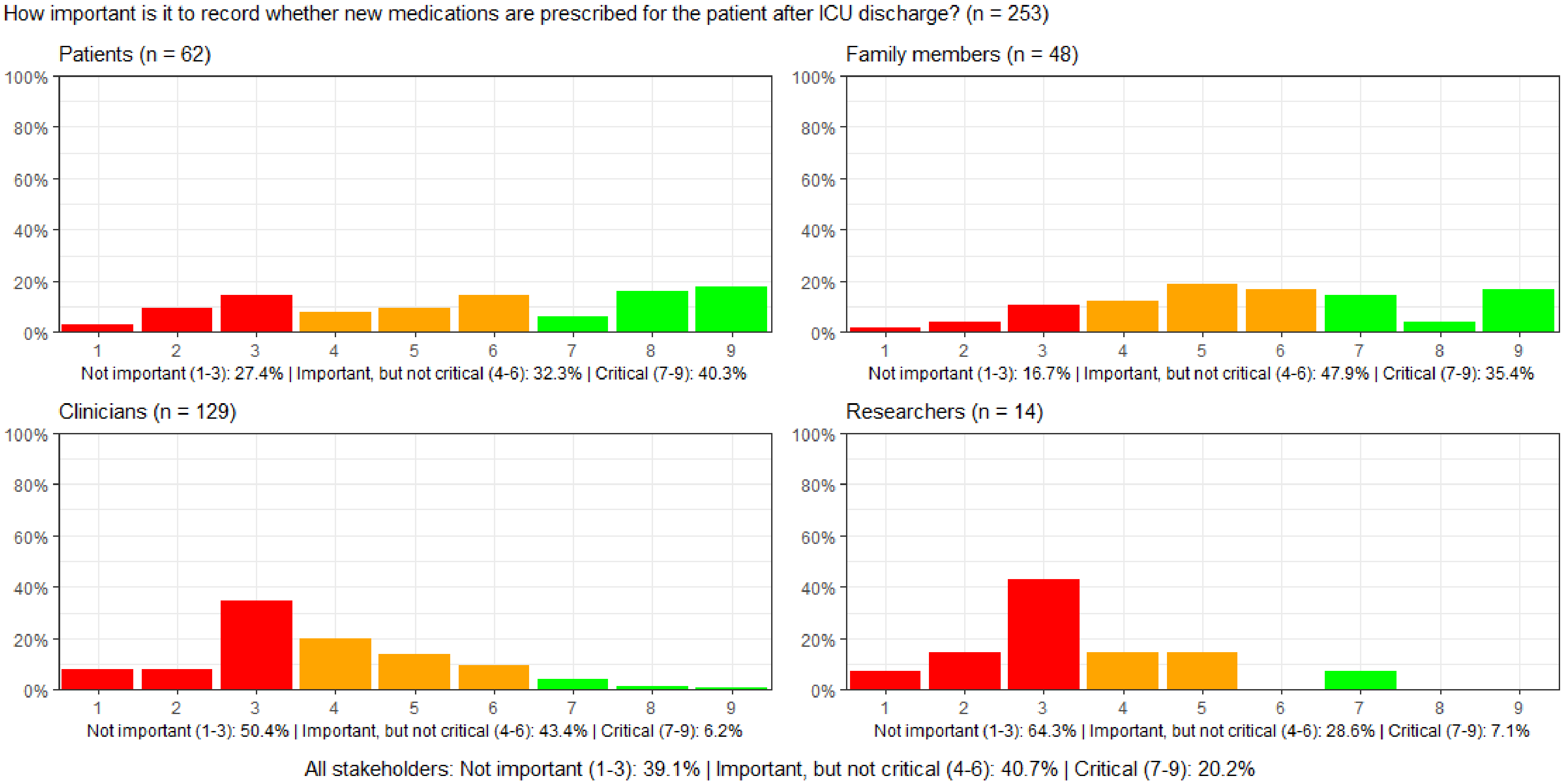

##### Figure S76: How important is it to record patients' health-related quality of life after ICU discharge?

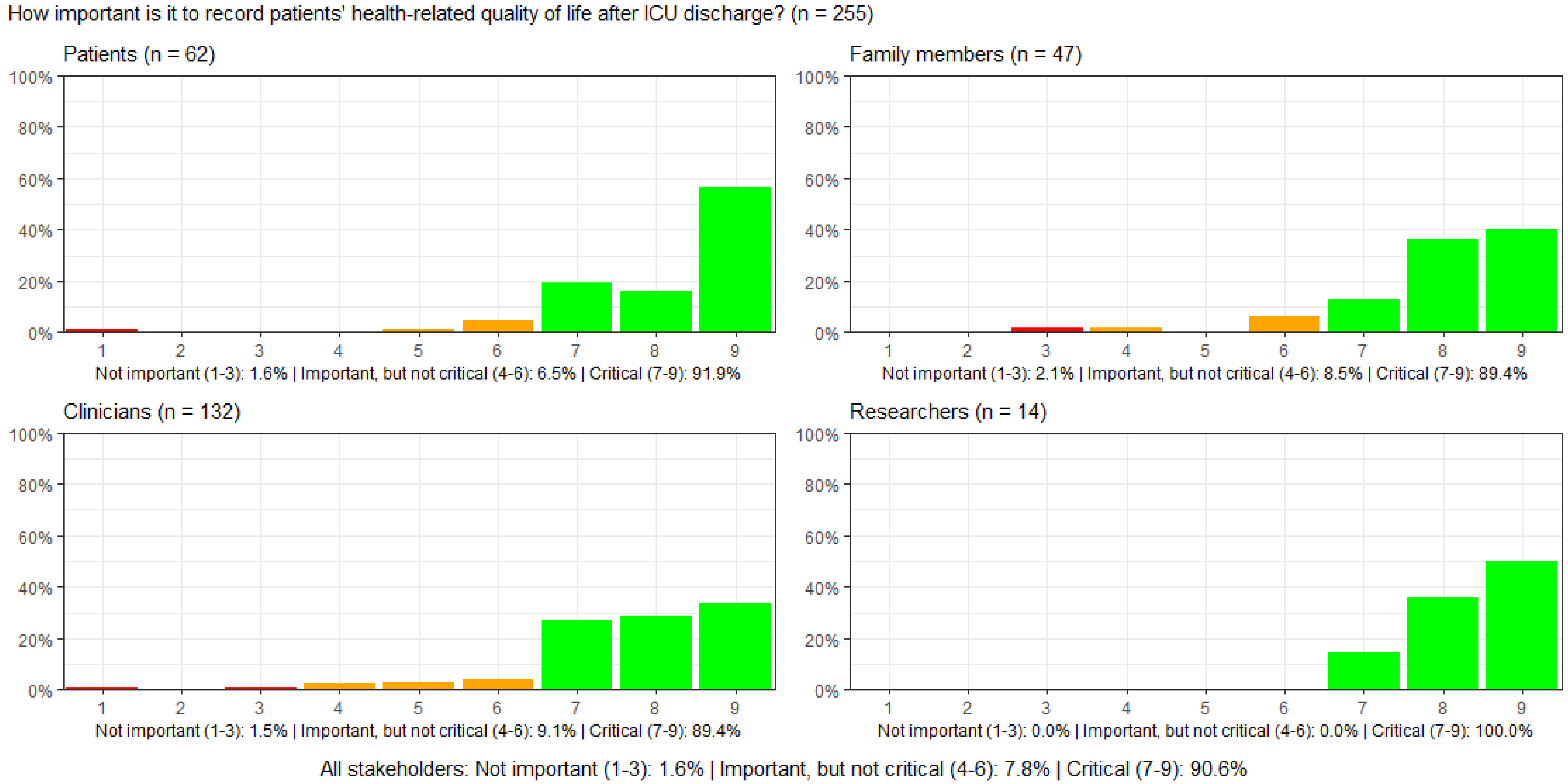

**Figure S77: How important is it to record whether patients experience newly onset tiredness or fatigue, unrelieved by sleep or rest, after ICU disch arge?**

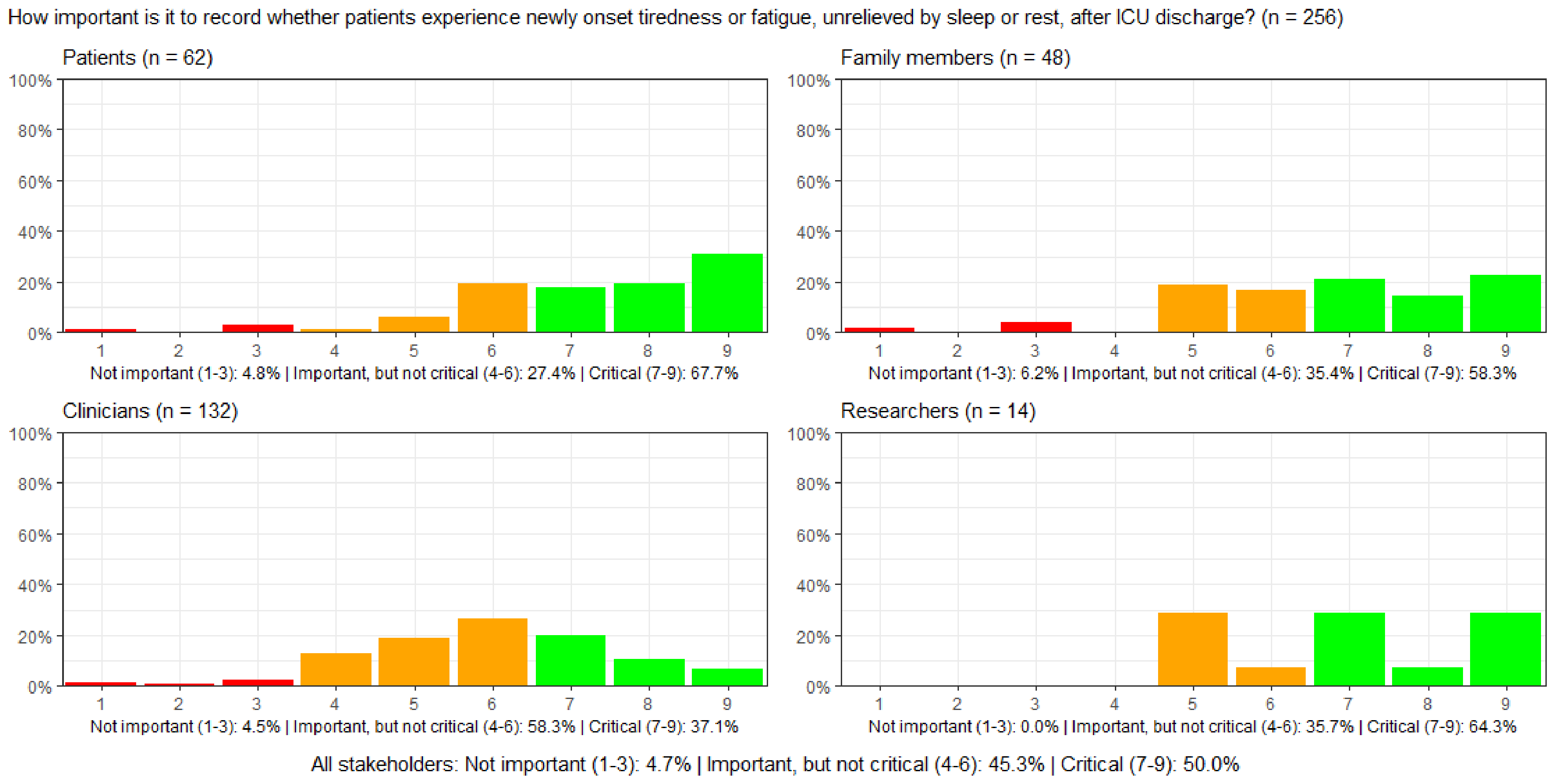

##### Figure S78: How important is it to record whether patients experience newly onset pain after ICU discharge?

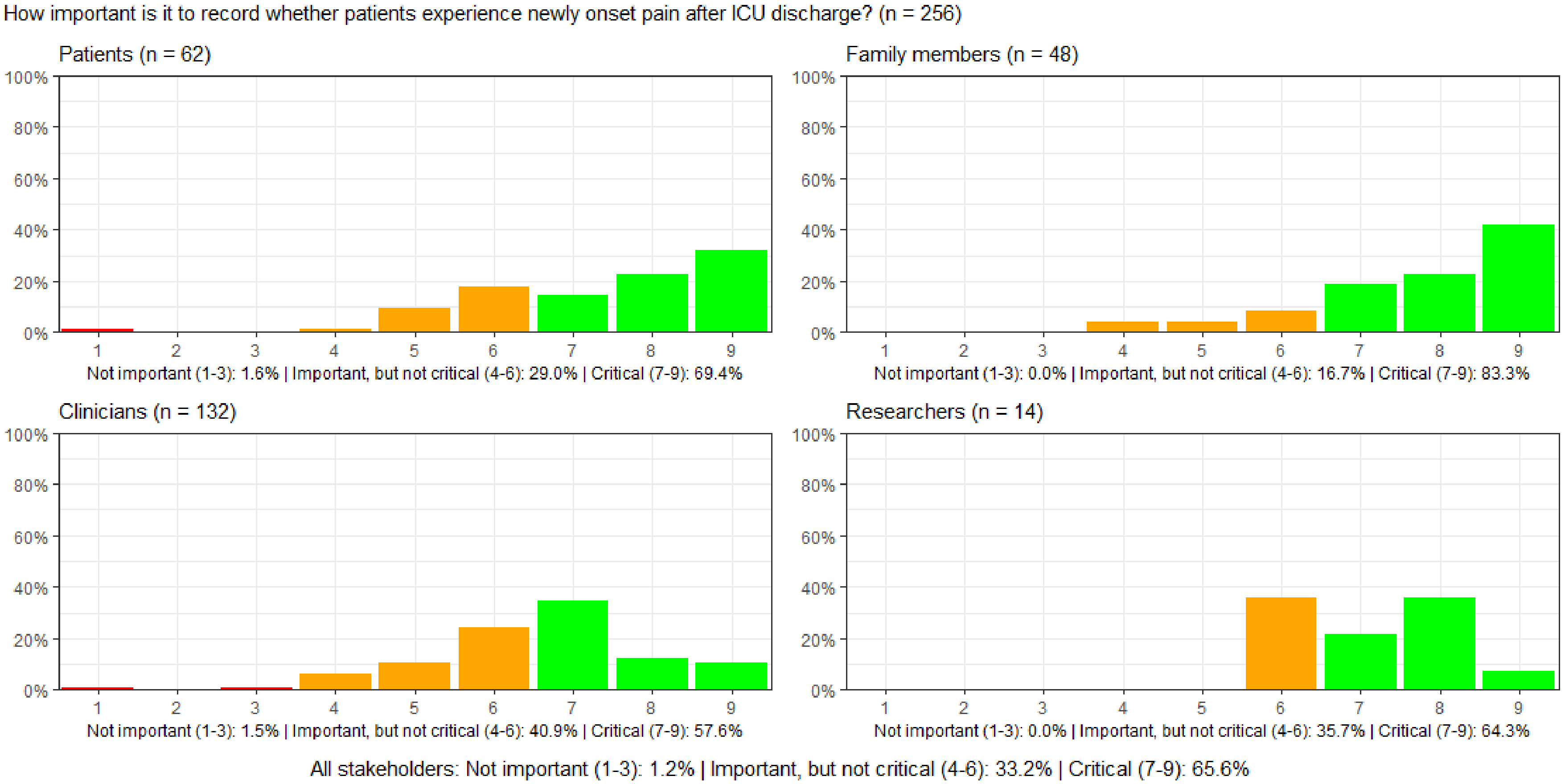

#### Figure S79: How important is it to record whether patients resume their usual roles in daily life after ICU discharge?

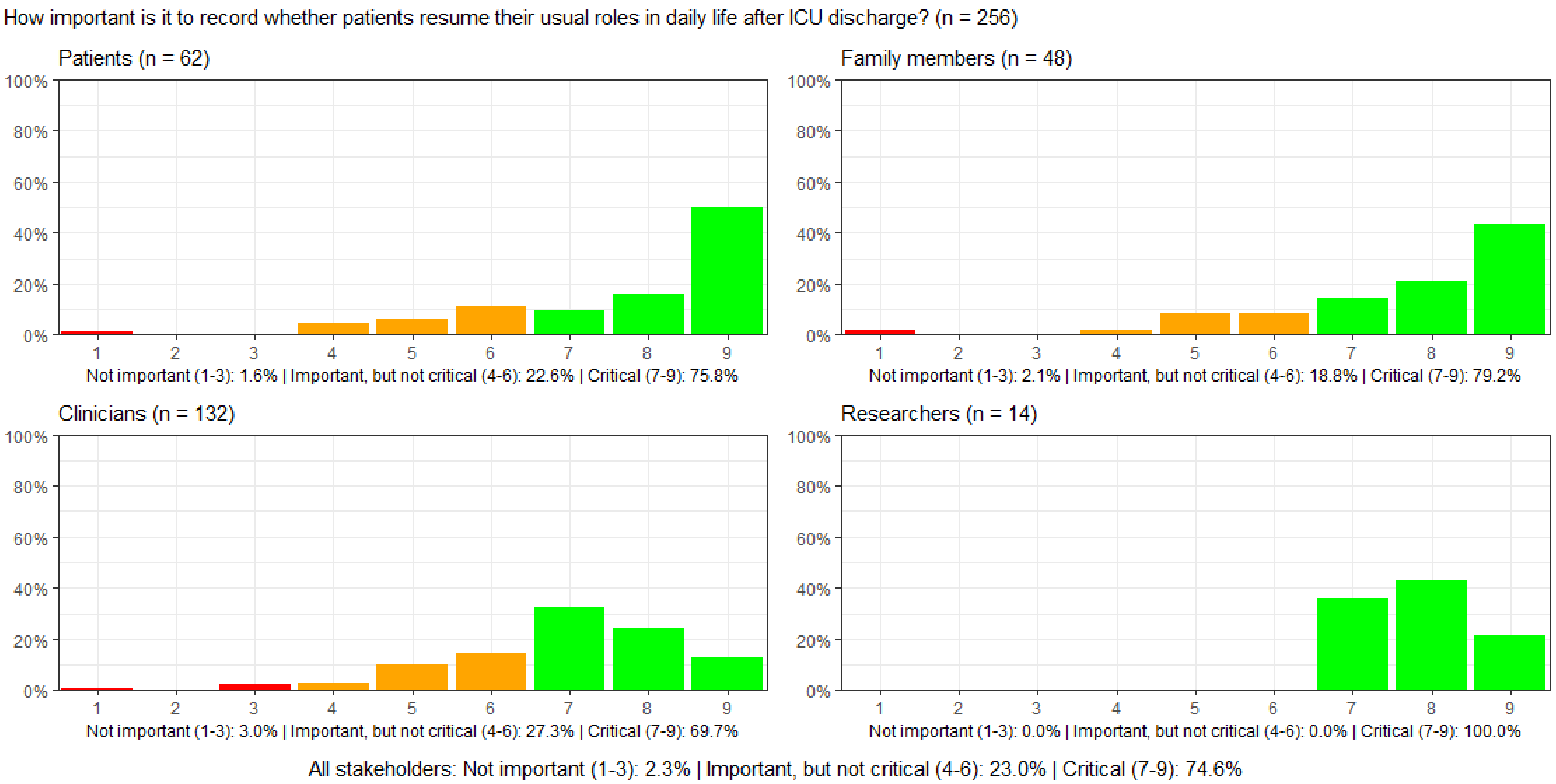

#### Figure S80: How important is it to record family caregivers' experienced quality of life after the ICU stay?

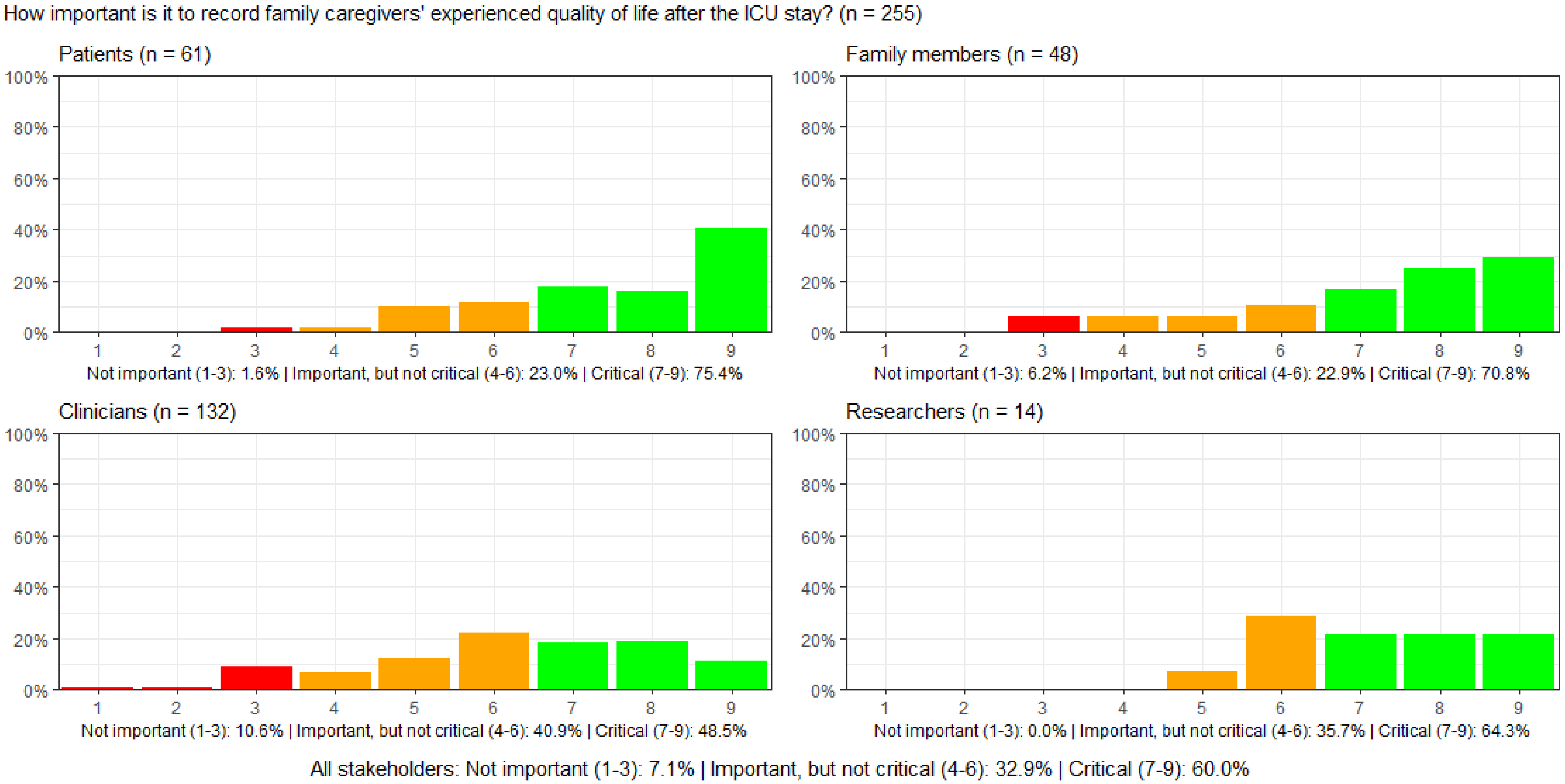

#### Figure S81: How important is it to record patients' financial capabilities after, compared to before, the ICU stay?

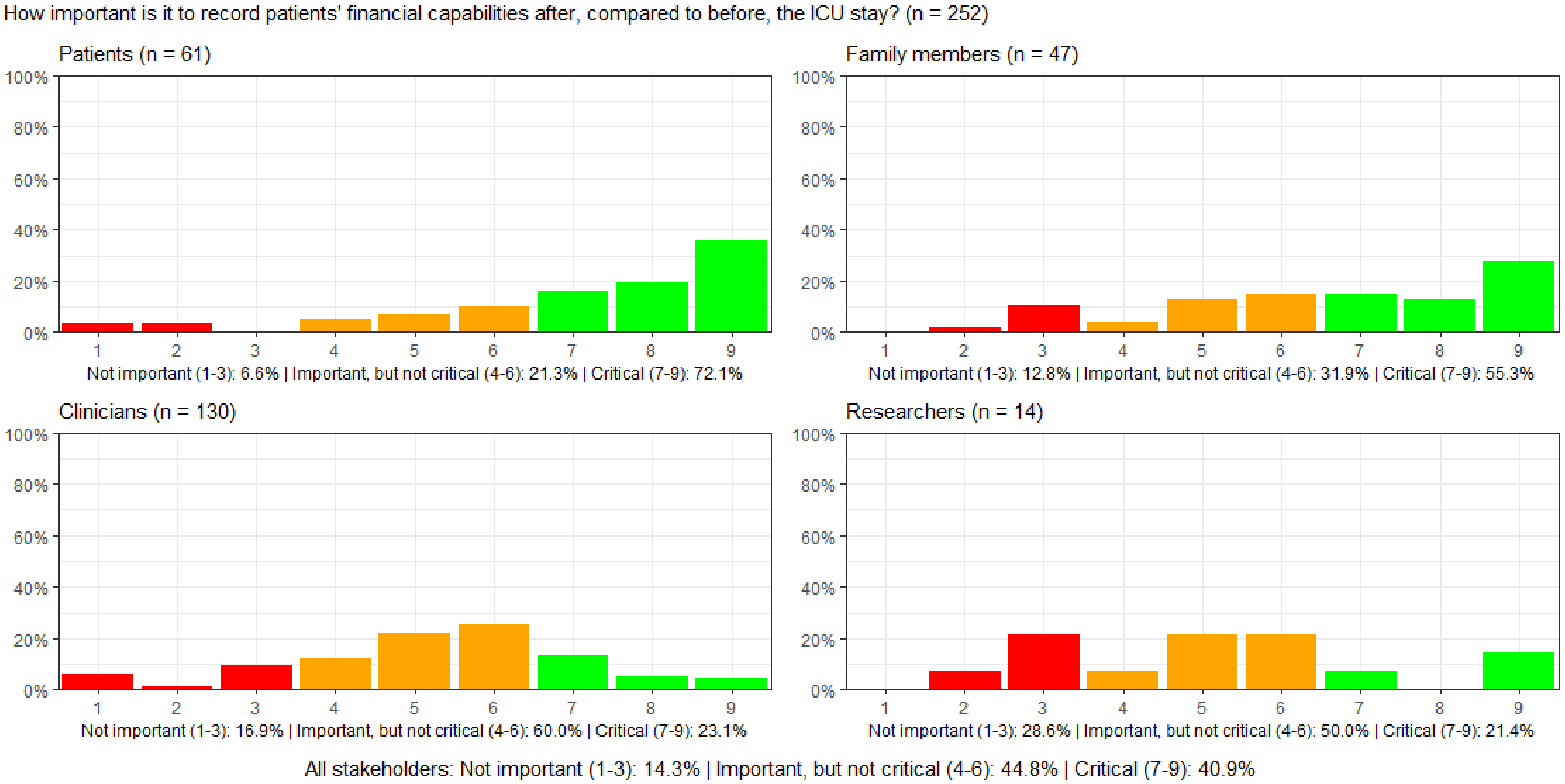

**Figure S82: How important is it to record whether patients after ICU discharge, compared to before, have the same labour market attachment.**

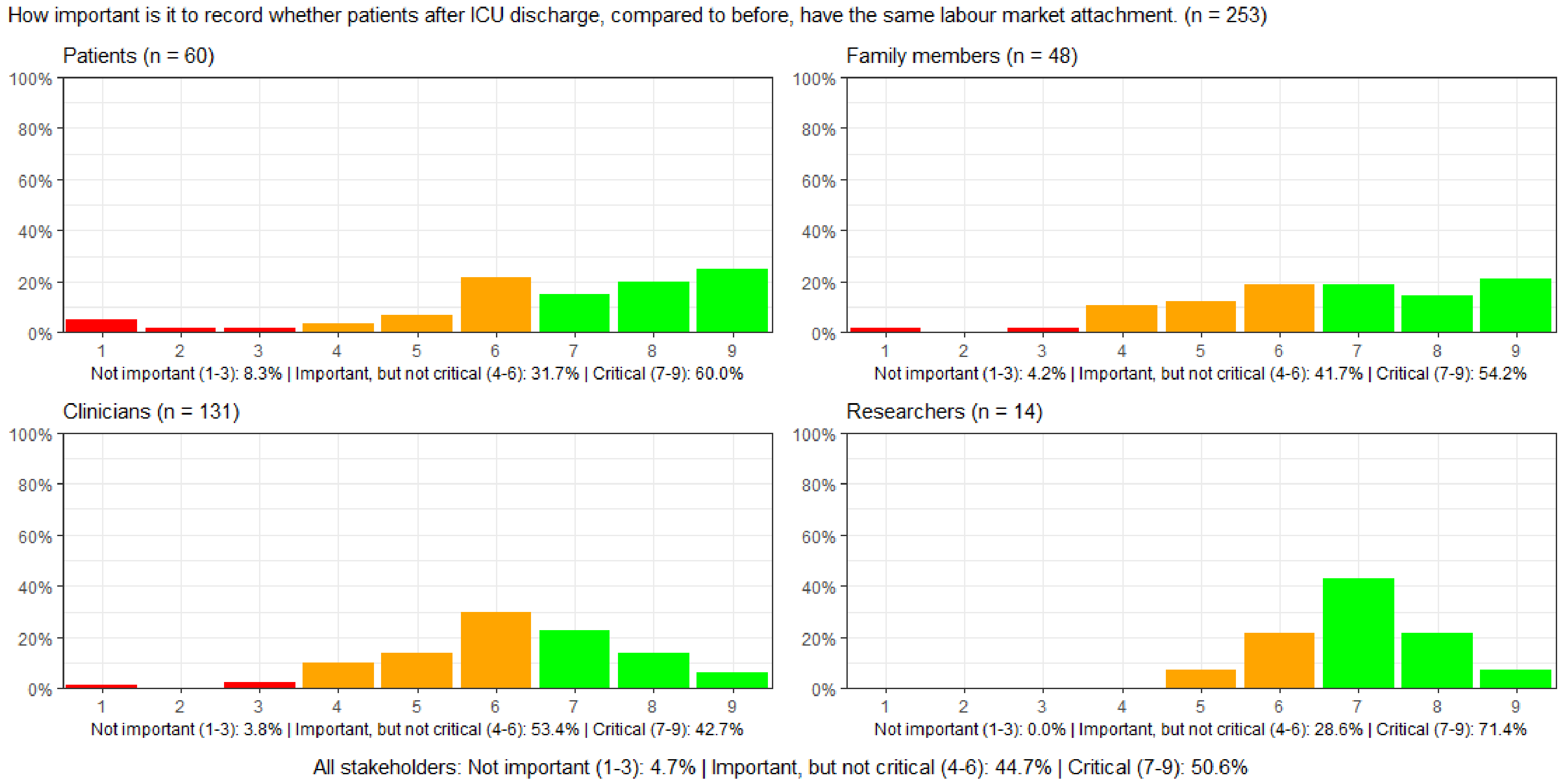

#### Figure S83: How important is it to record patients' physical impediments such as scars, wounds, or ostomy after ICU discharge?

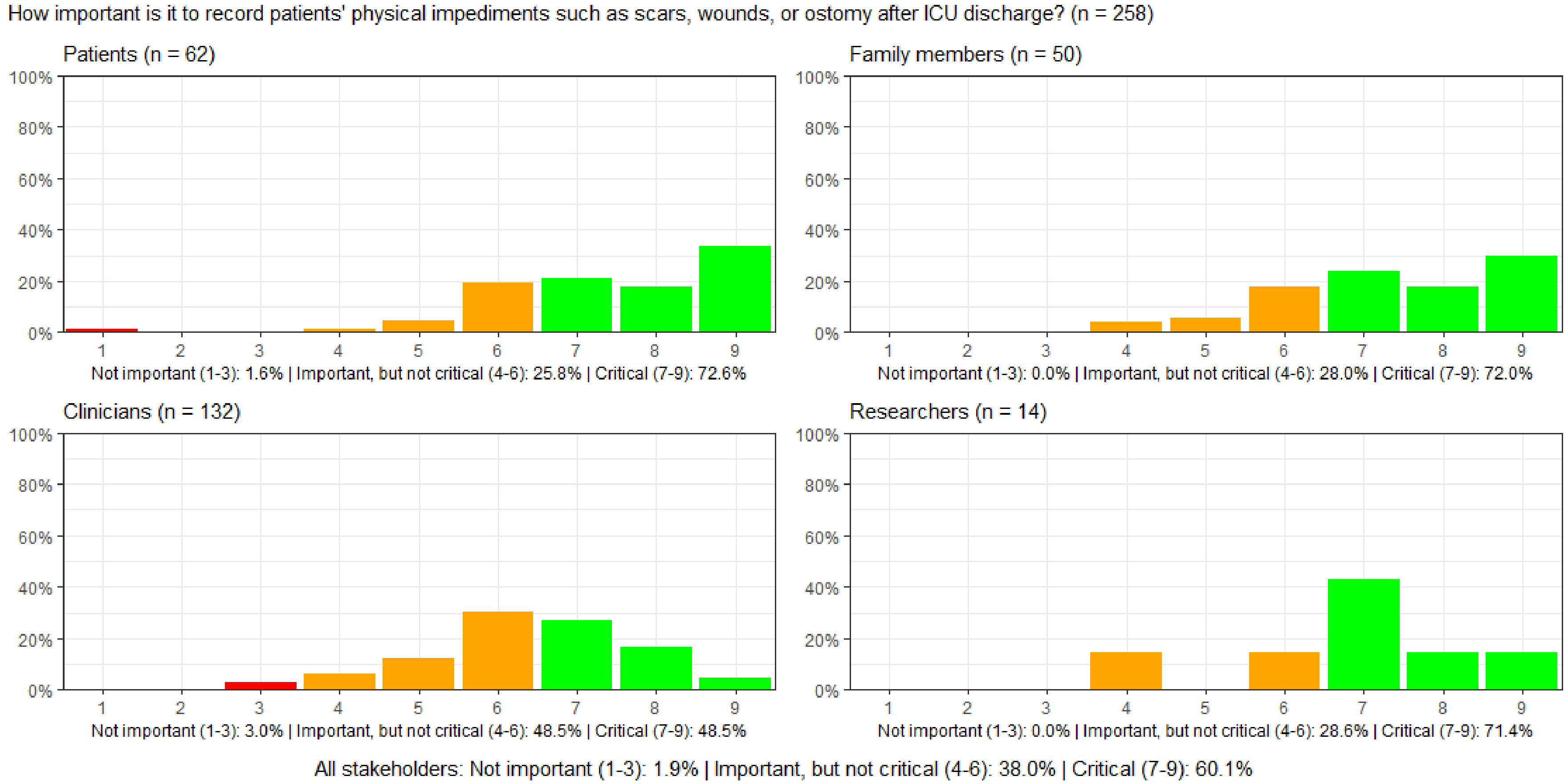

#### Figure S84: How important is it to record whether patients independently perform activities of daily living after ICU discharge?

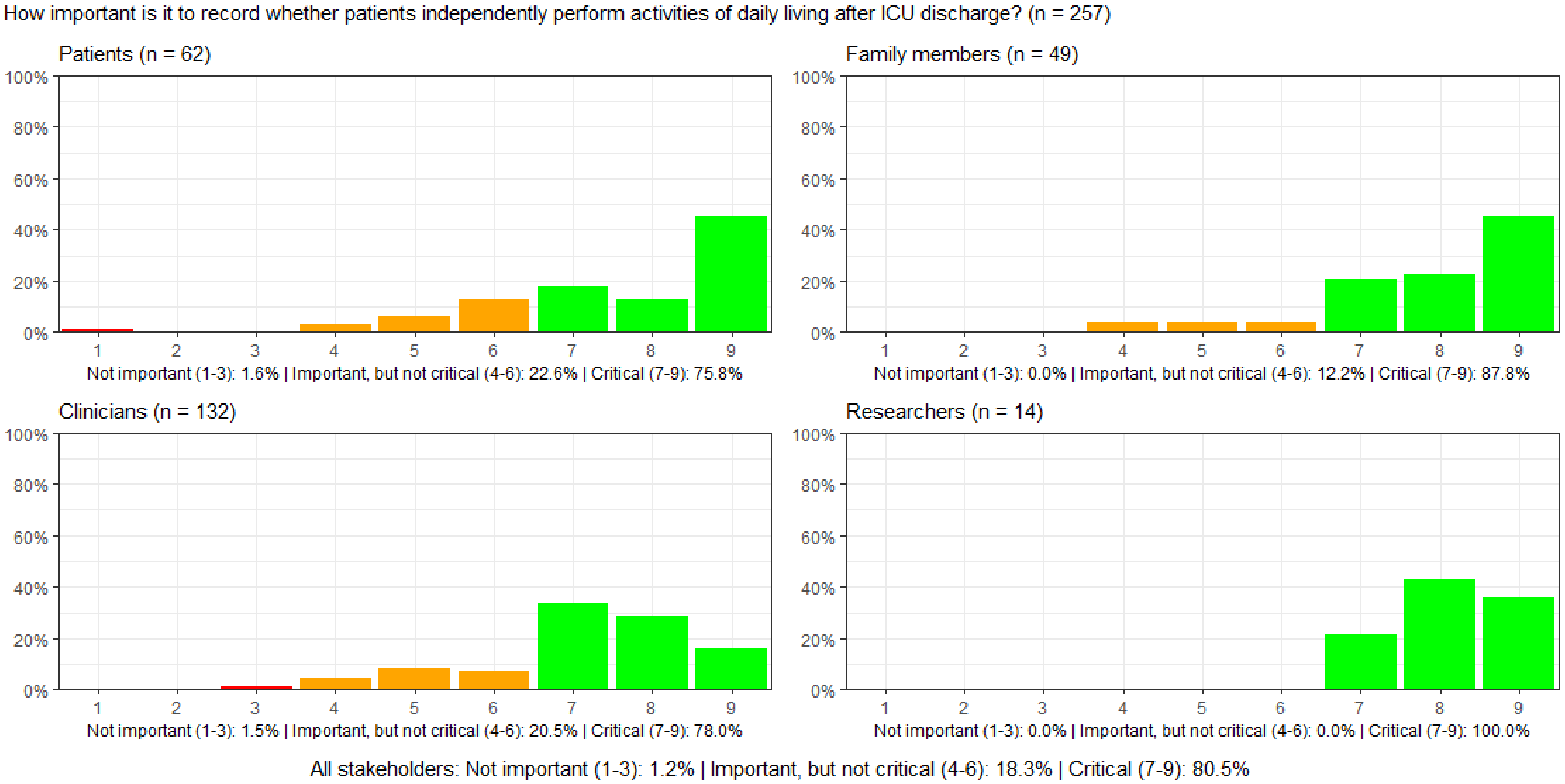

**Figure S85: How important is it to record whether patients are independently able to get dressed after ICU discharge?**

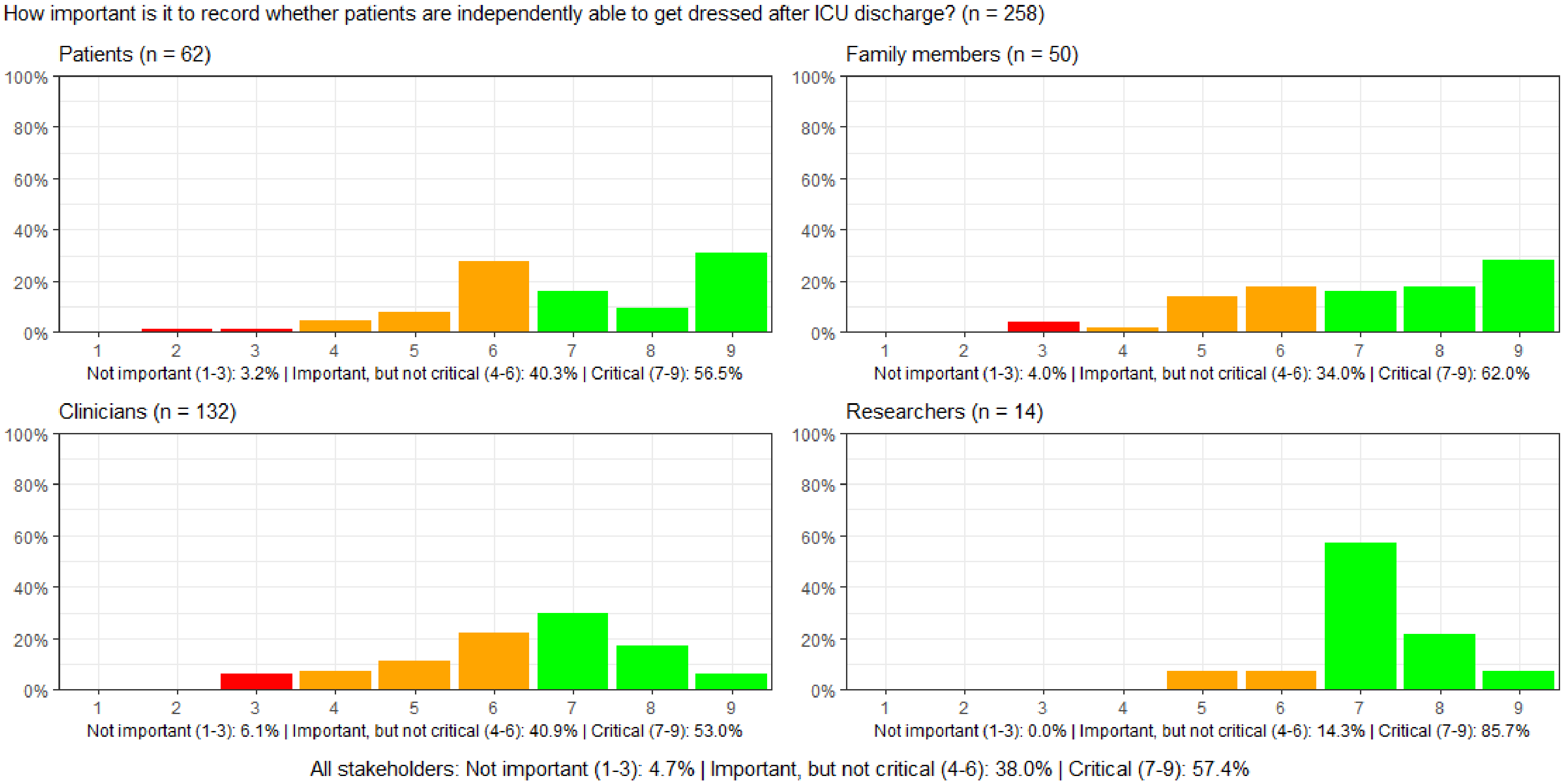

#### Figure S86: How important is it to record whether patients are mobile after ICU discharge?

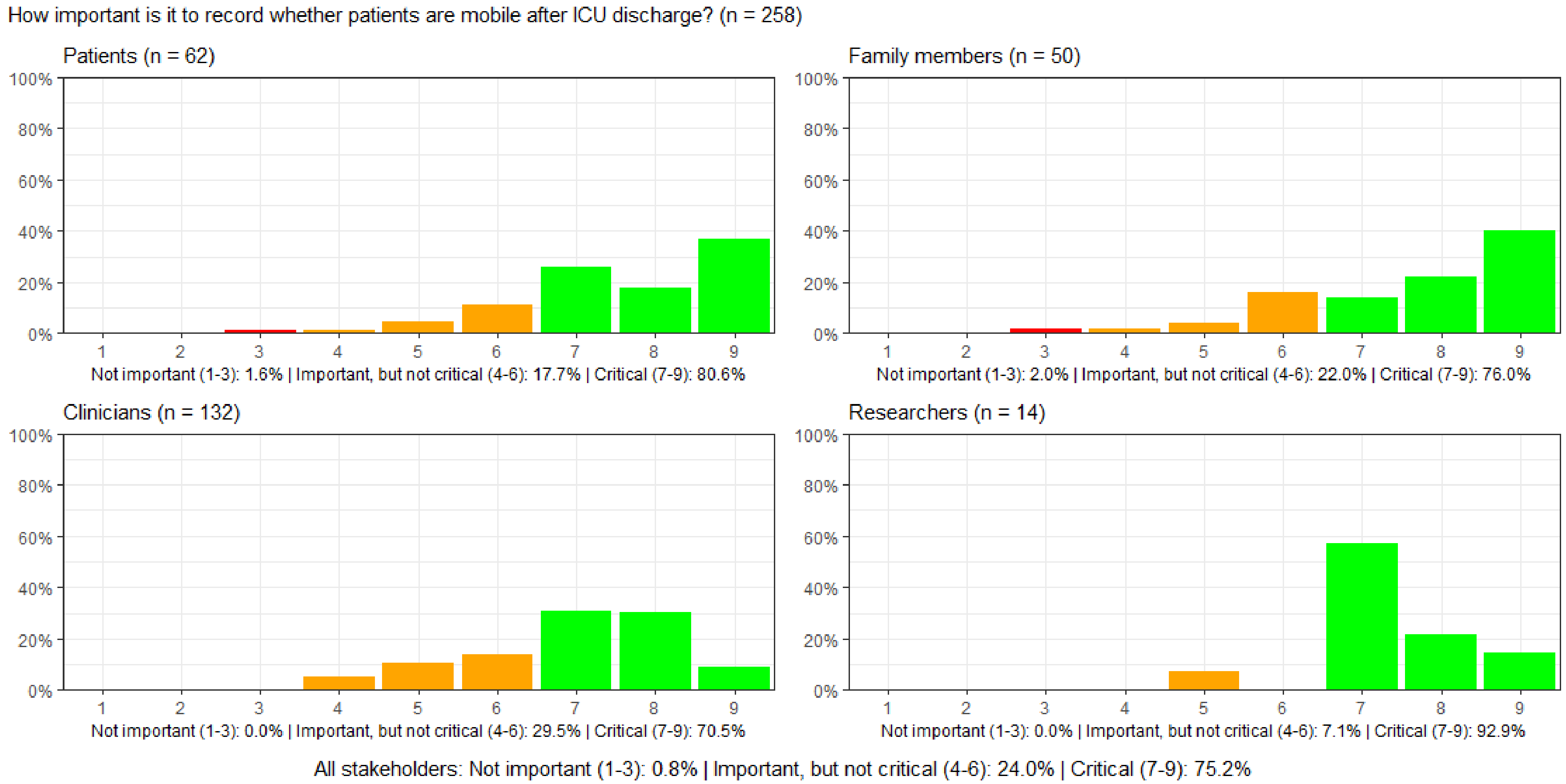

#### Figure S87: How important is it to record whether patients experience newly onset shortness of breath after ICU discharge?

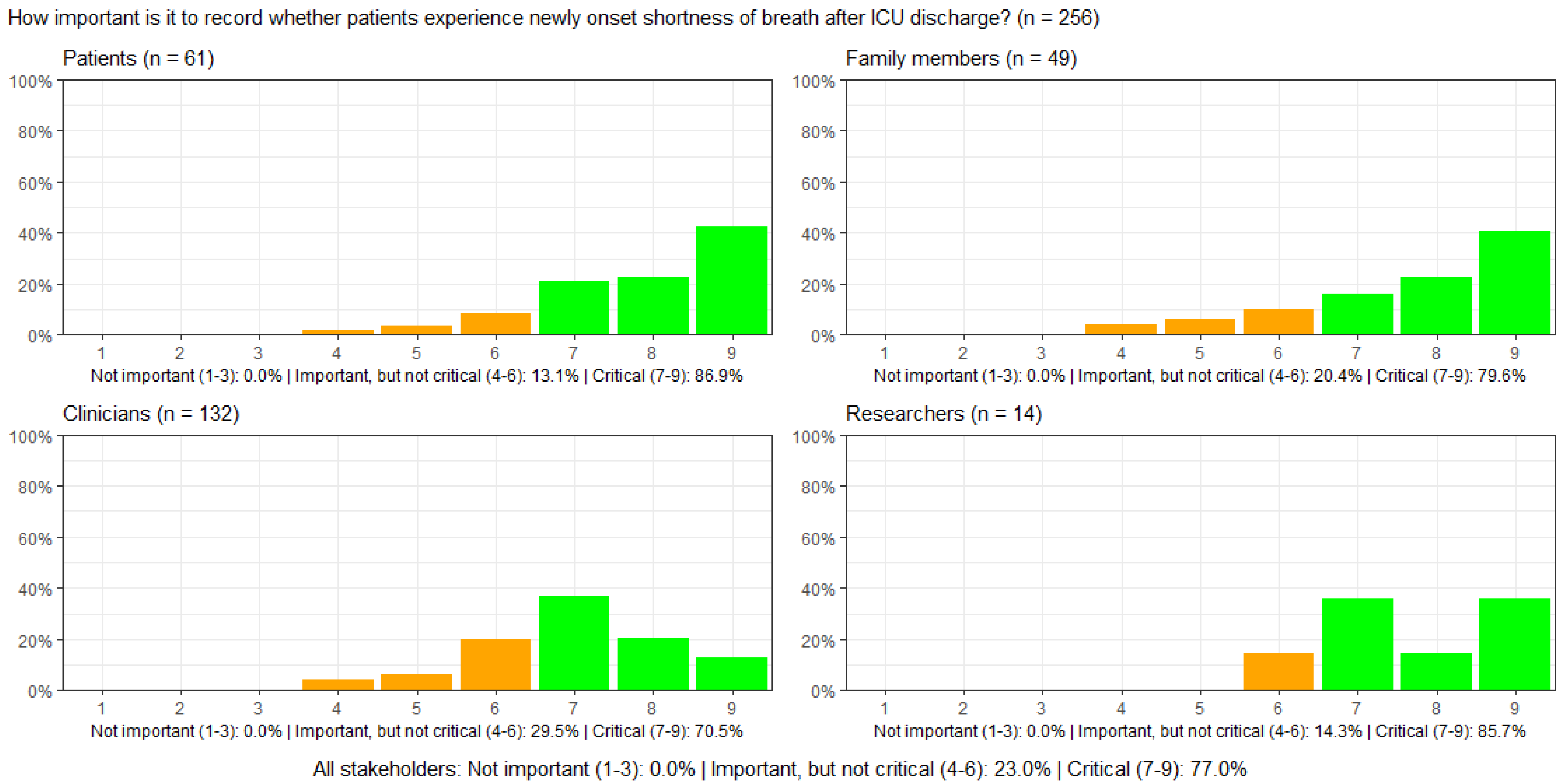

#### Figure S88: How important is it to record whether patients have newly onset trouble speaking after ICU discharge?

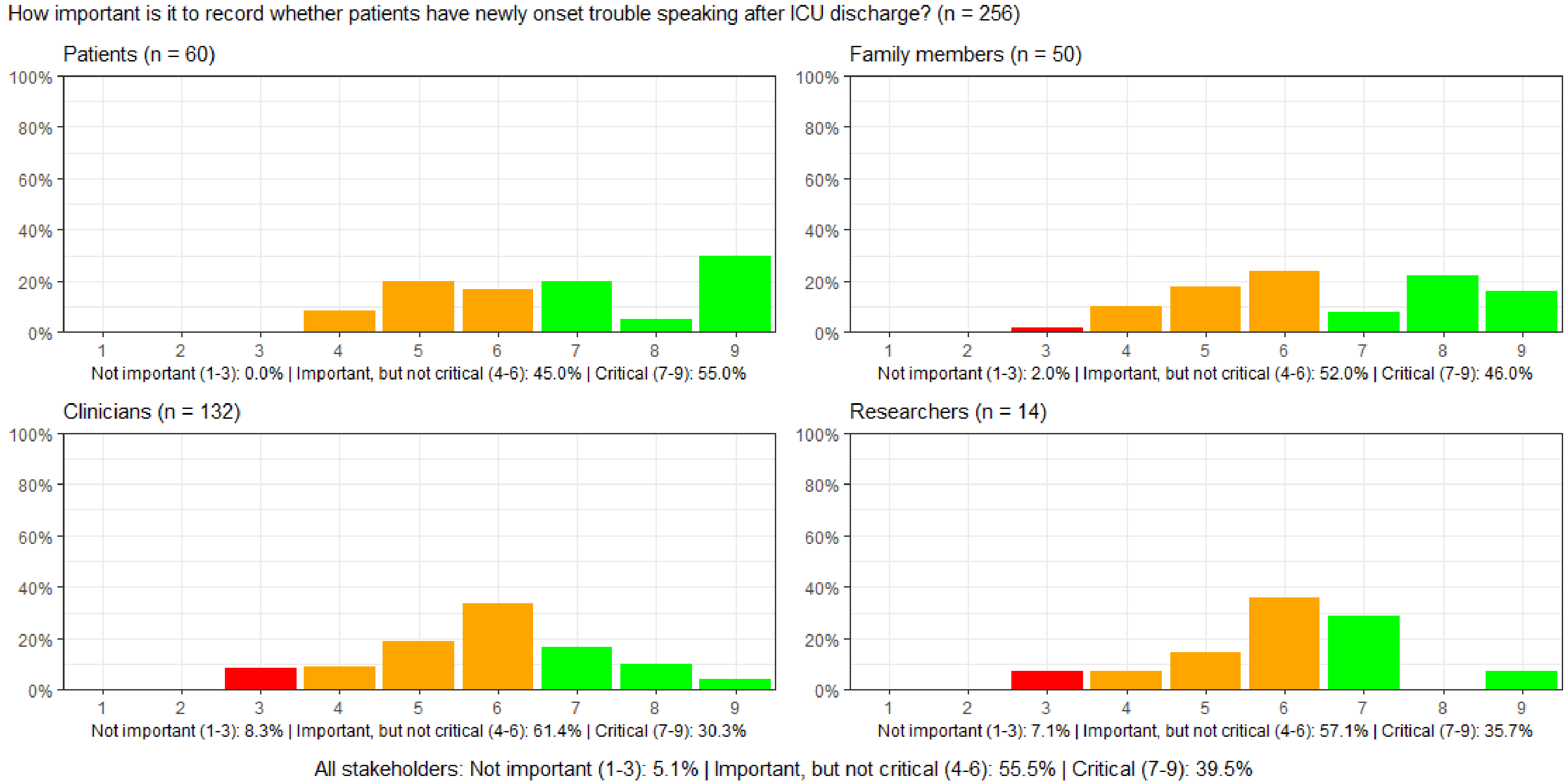

#### Figure S89: How important is it to record whether patients have newly onset difficulty swallowing after ICU discharge?

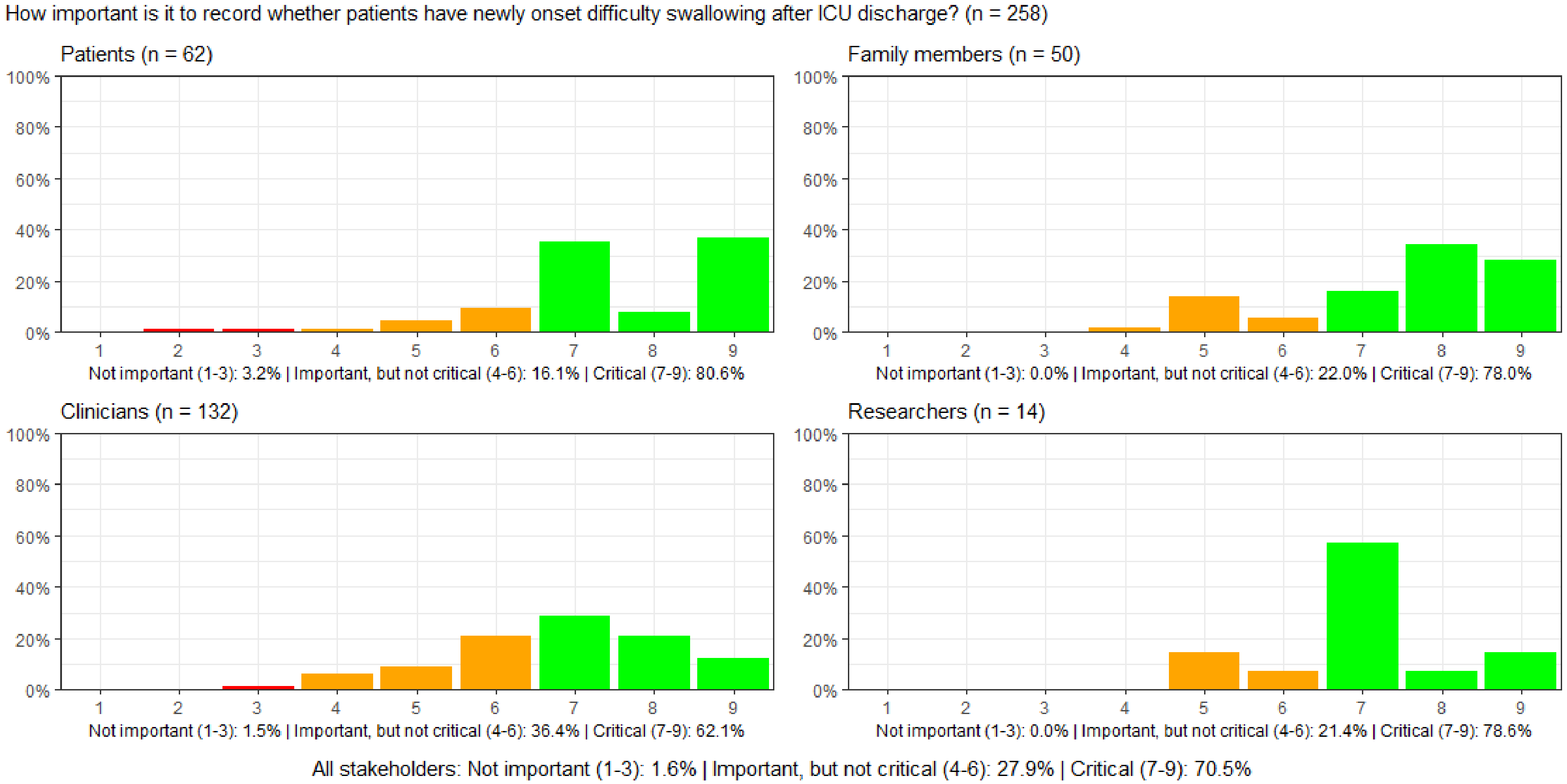

#### Figure S90: How important is it to record patients' bowel function after ICU discharge?

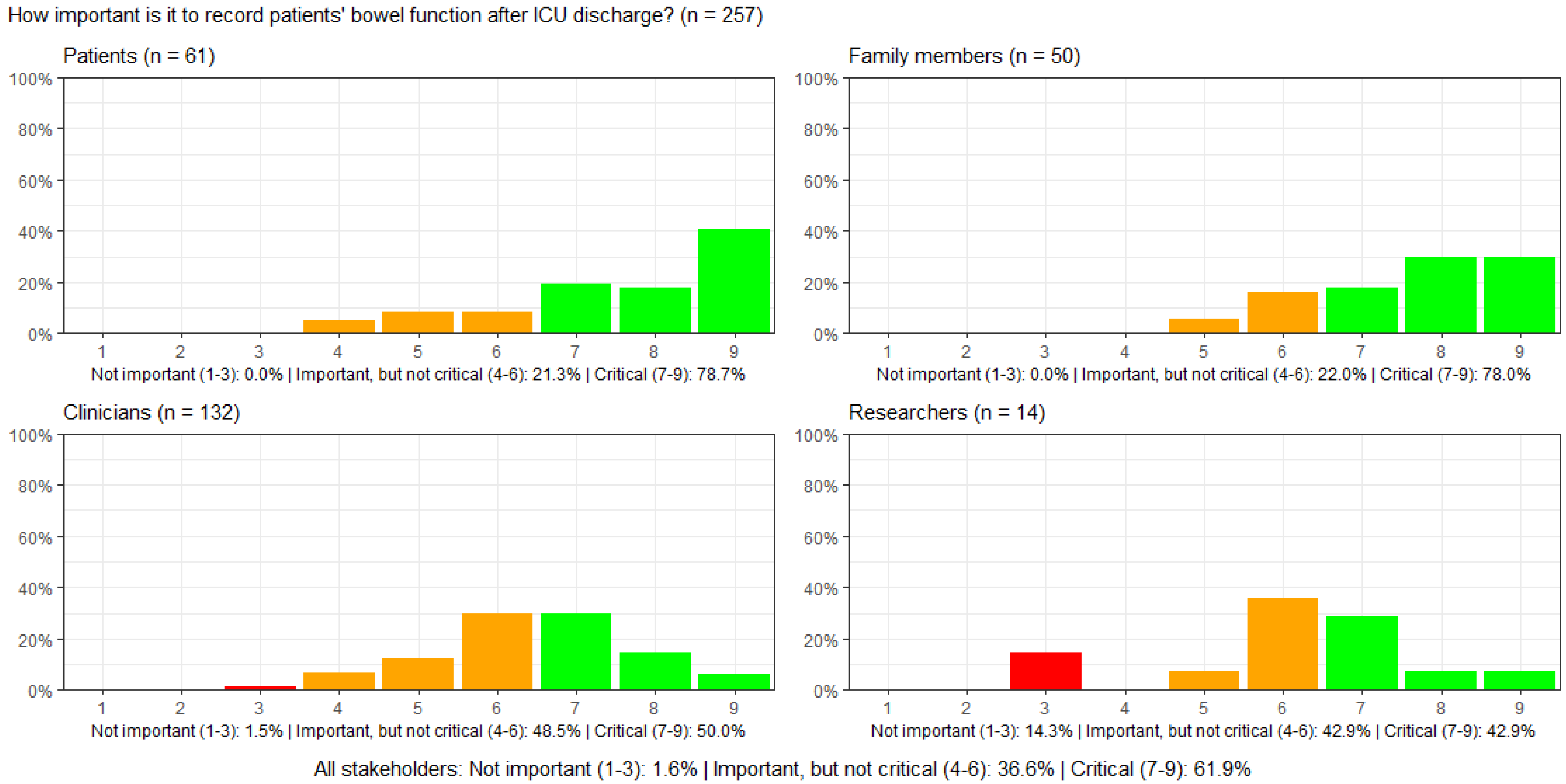

#### Figure S91: How important is it to record patients' urinary function after ICU discharge?

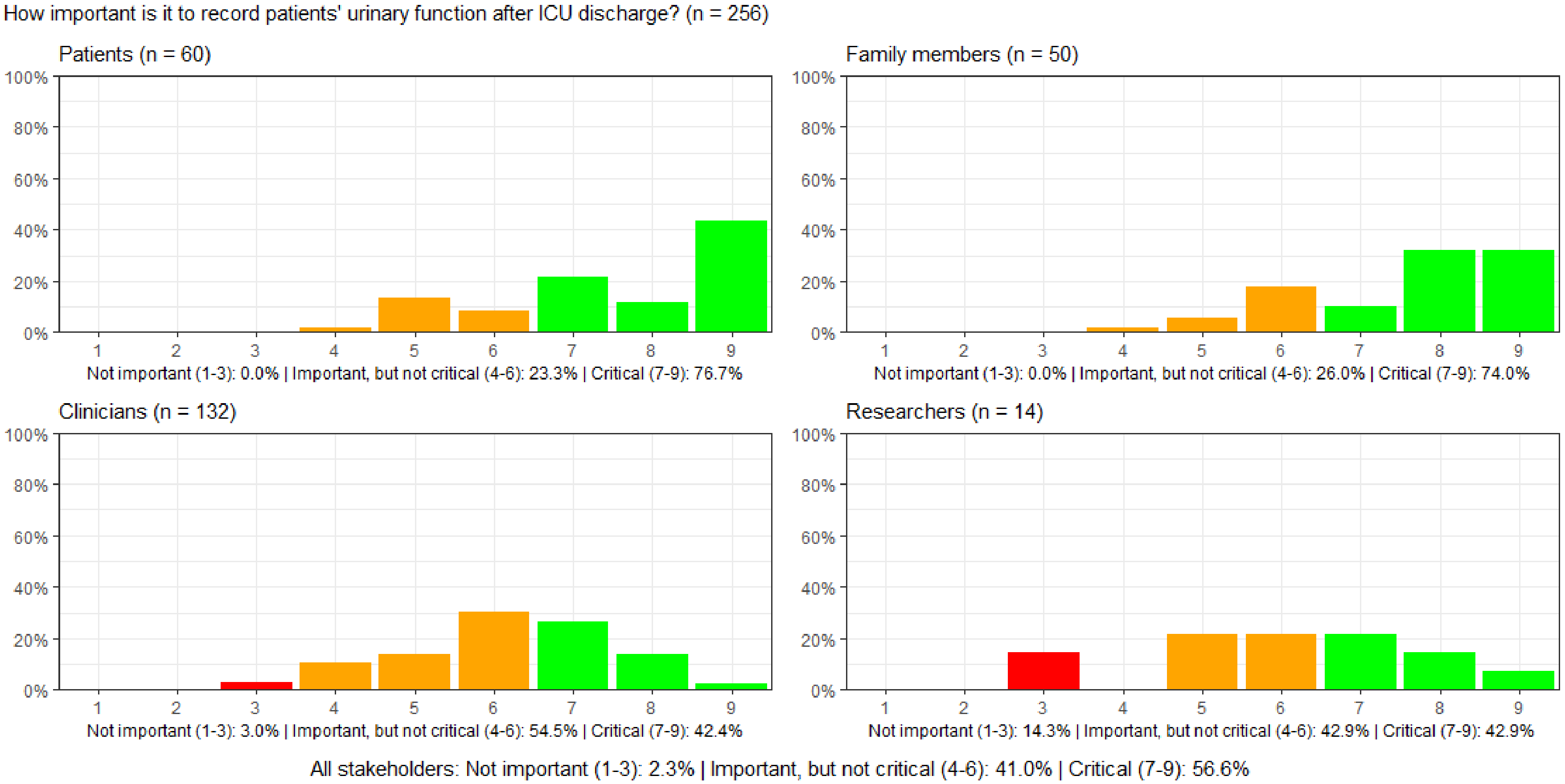

#### Figure S92: How important is it to record the patient's sexual function after ICU discharge?

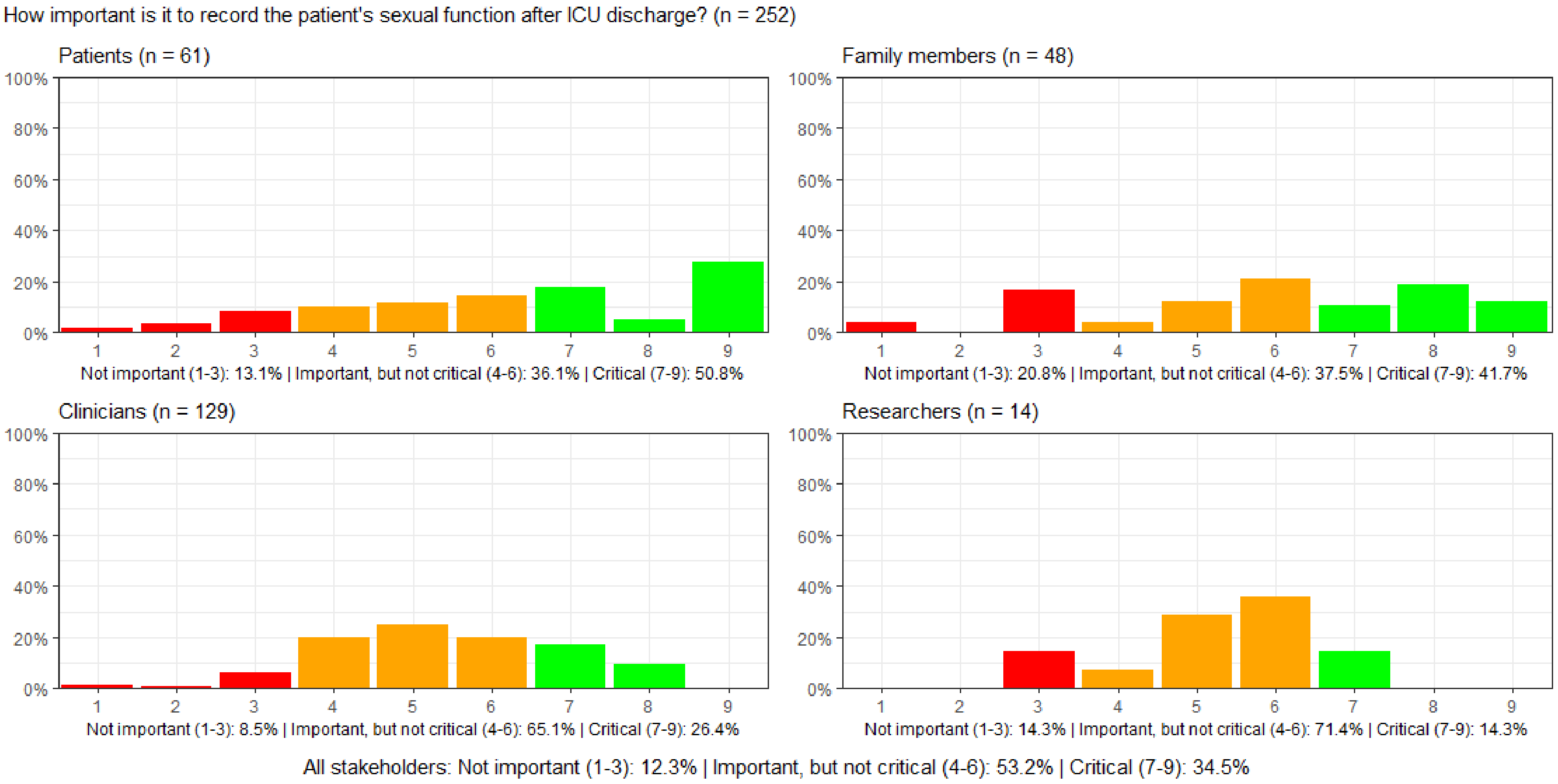

#### Figure S93: How important is it to record patients' senses: vision, hearing, touch, smell, and taste after ICU discharge?

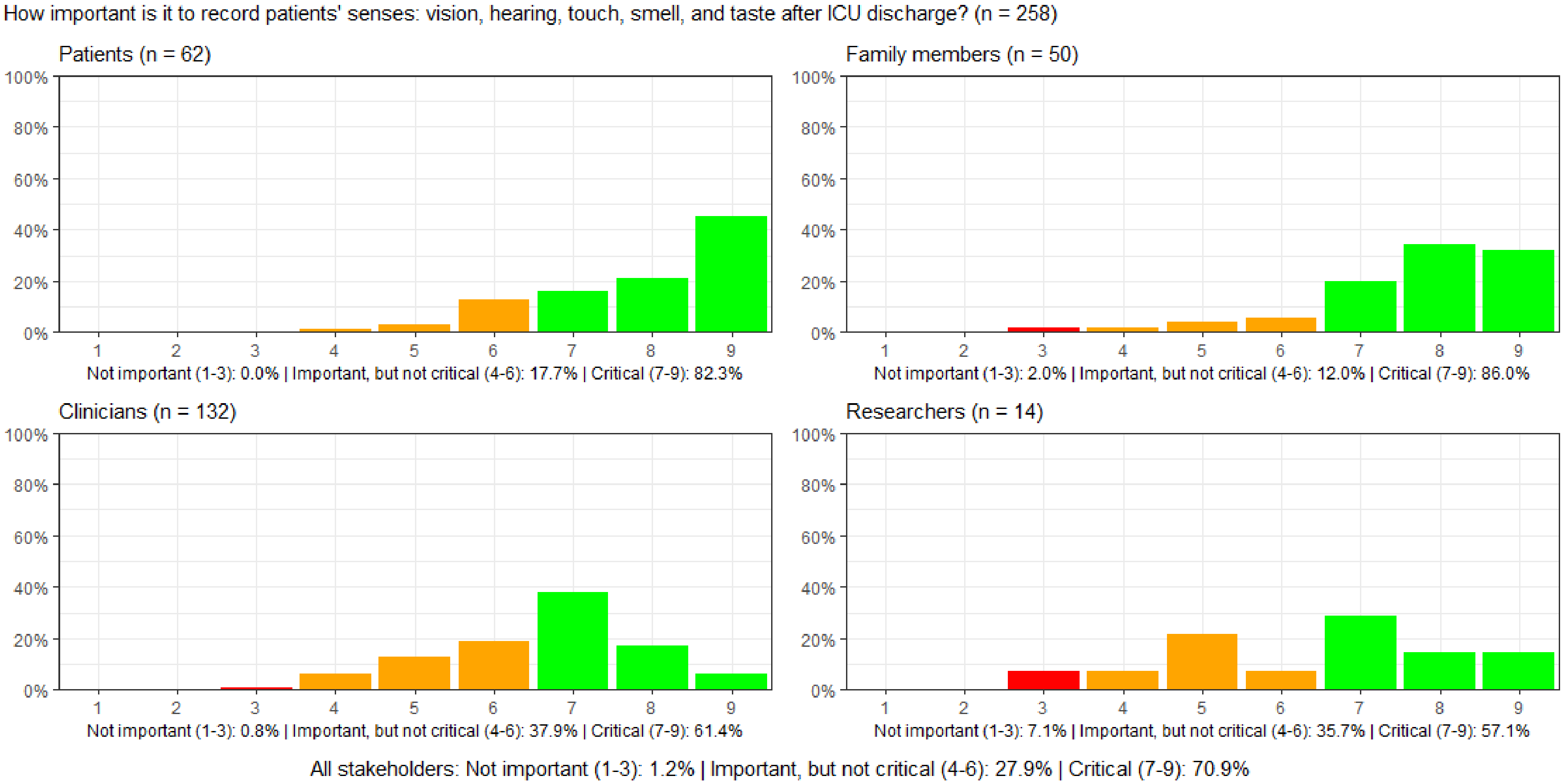

#### Figure S94: How important is it to record whether patients have the physical strength to perform activities of daily living after ICU discharge?

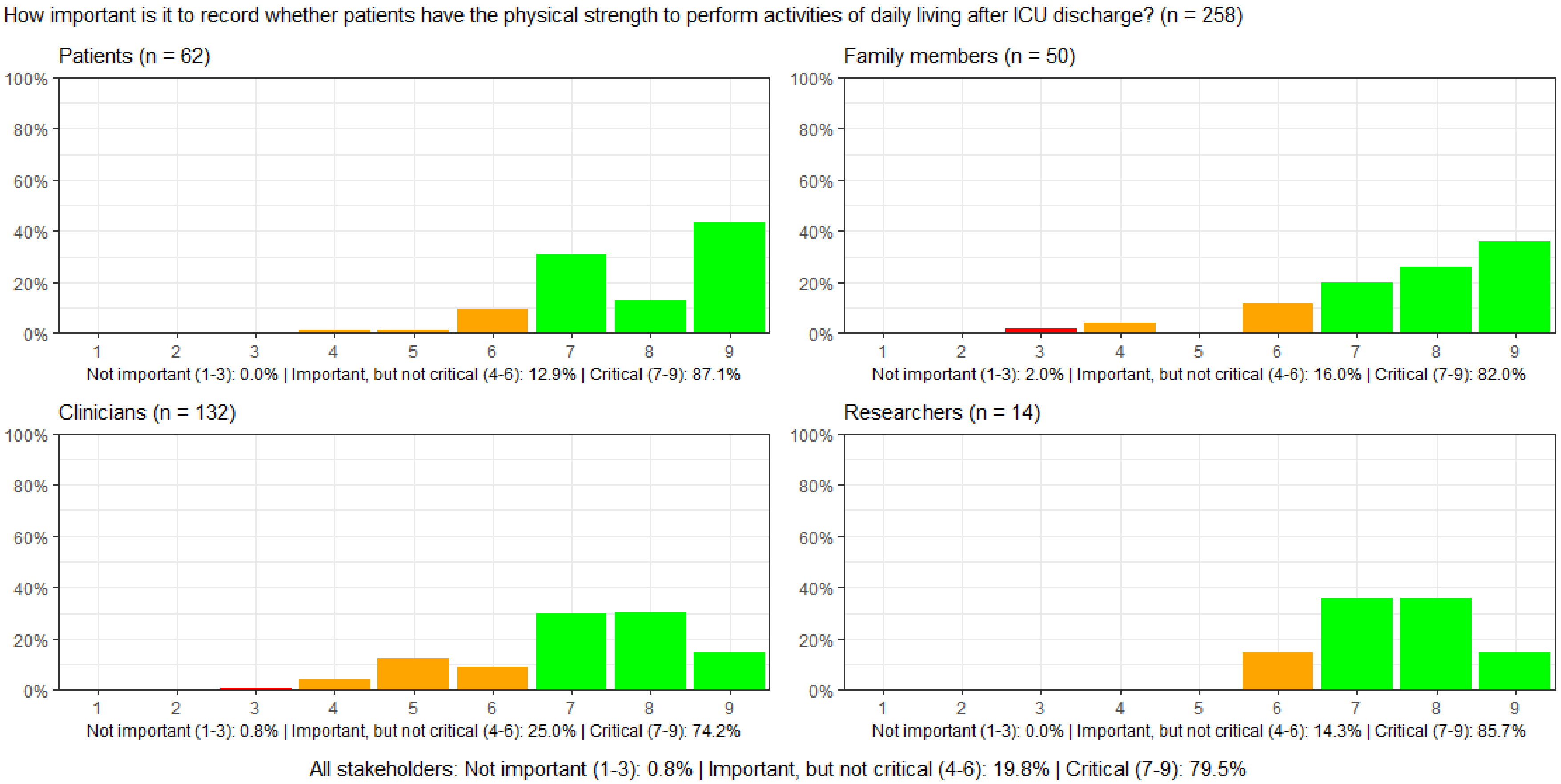

##### Figure S95: How important is it to record patients' mental function (cognitive function) after ICU discharge?

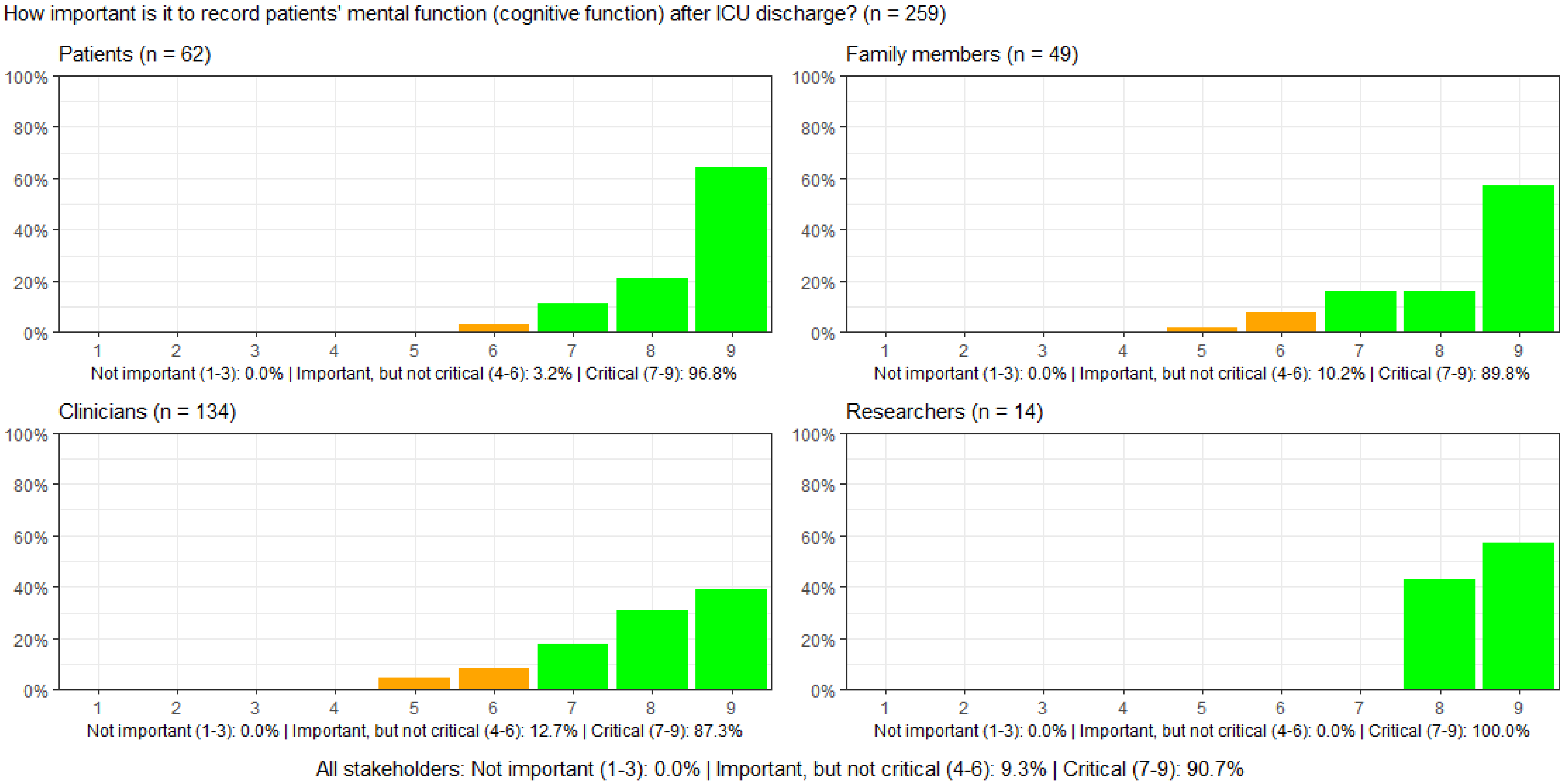

#### Figure S96: How important is it to record whether patients experience loneliness after ICU discharge?

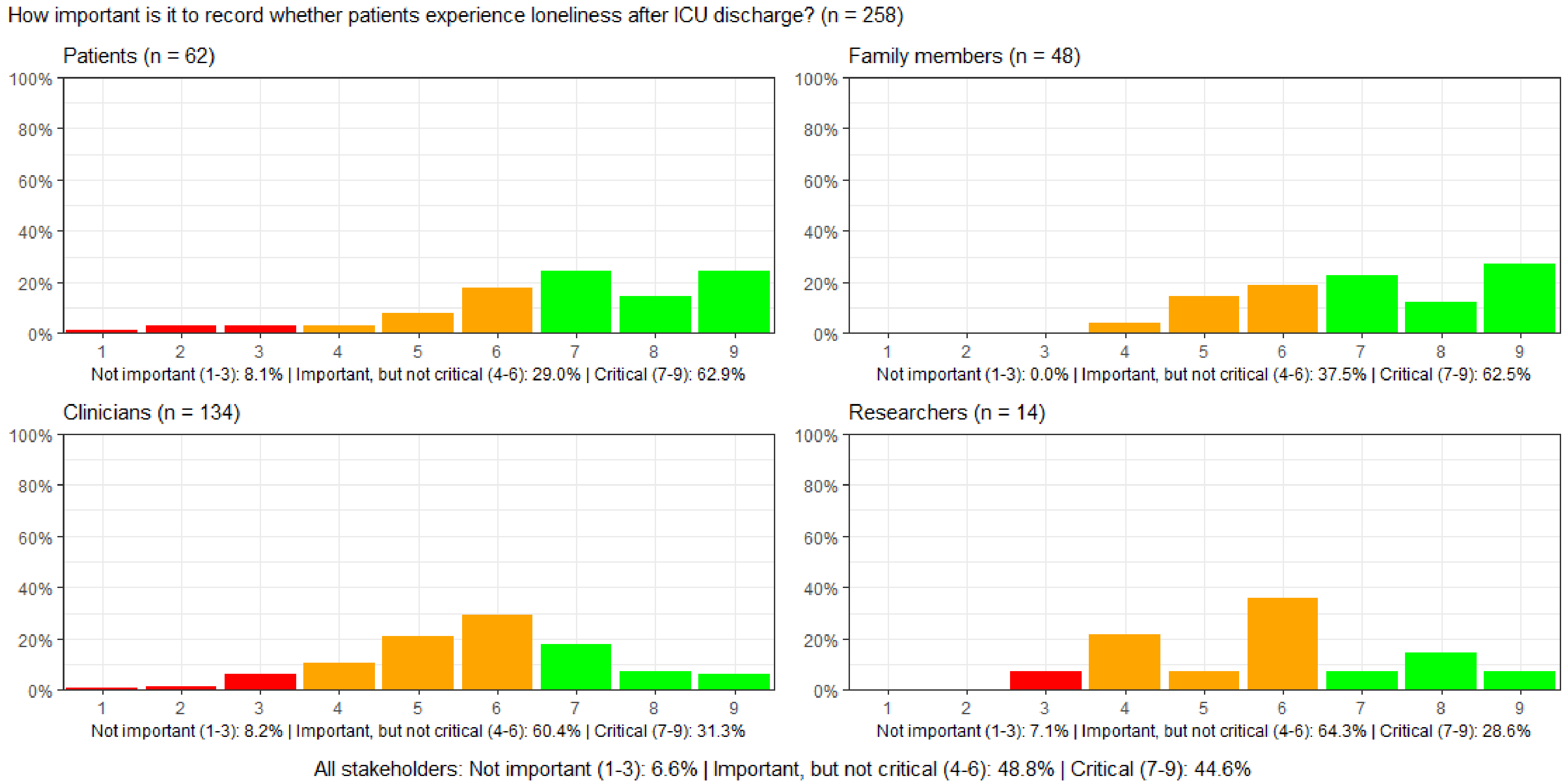

#### Figure S97: How important is it to record whether patients are diagnosed with posttraumatic stress syndrome (PTSD) after ICU discharge?

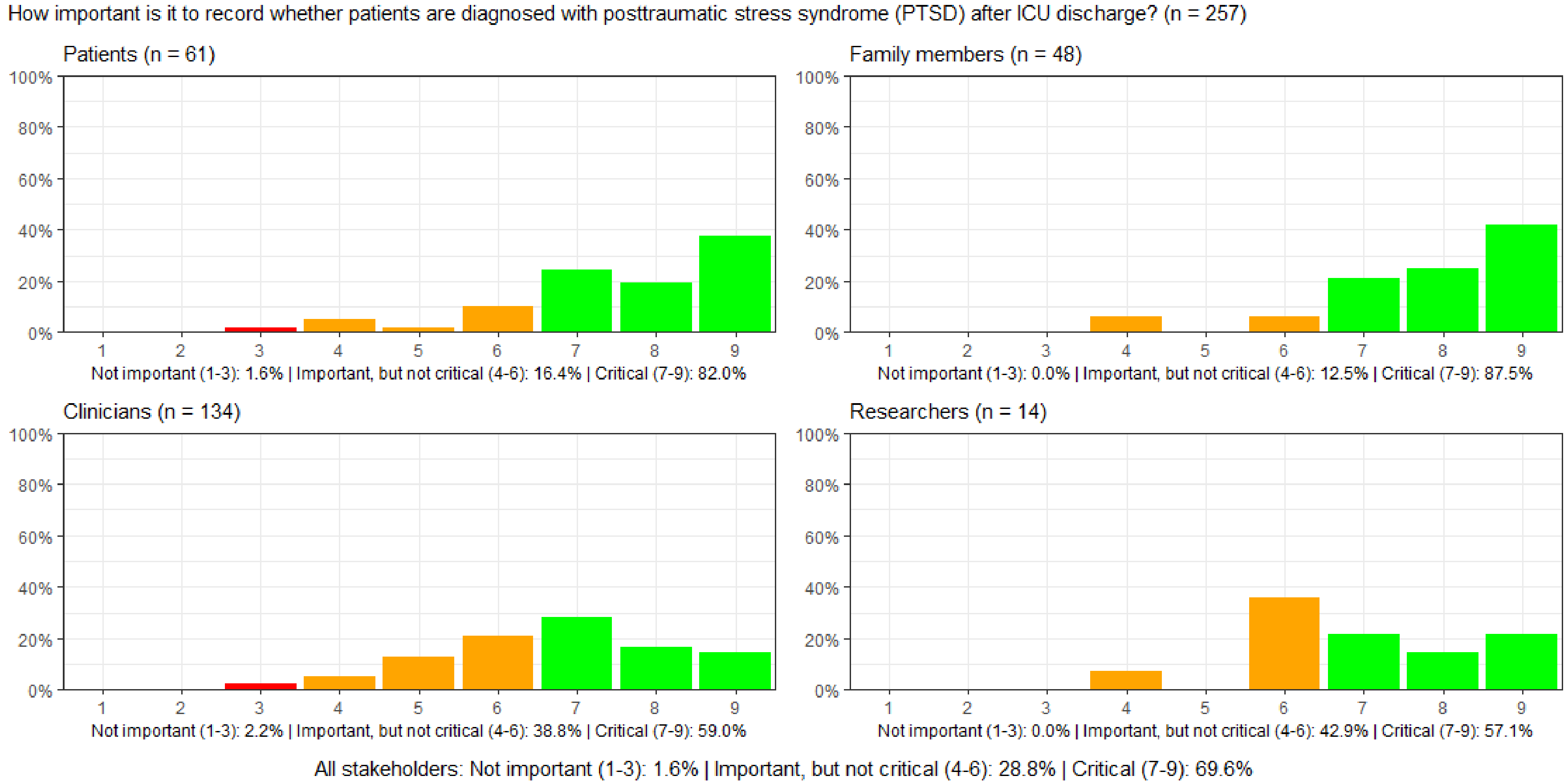

#### Figure S98: How important is it to record whether patients experience anxiety after ICU discharge?

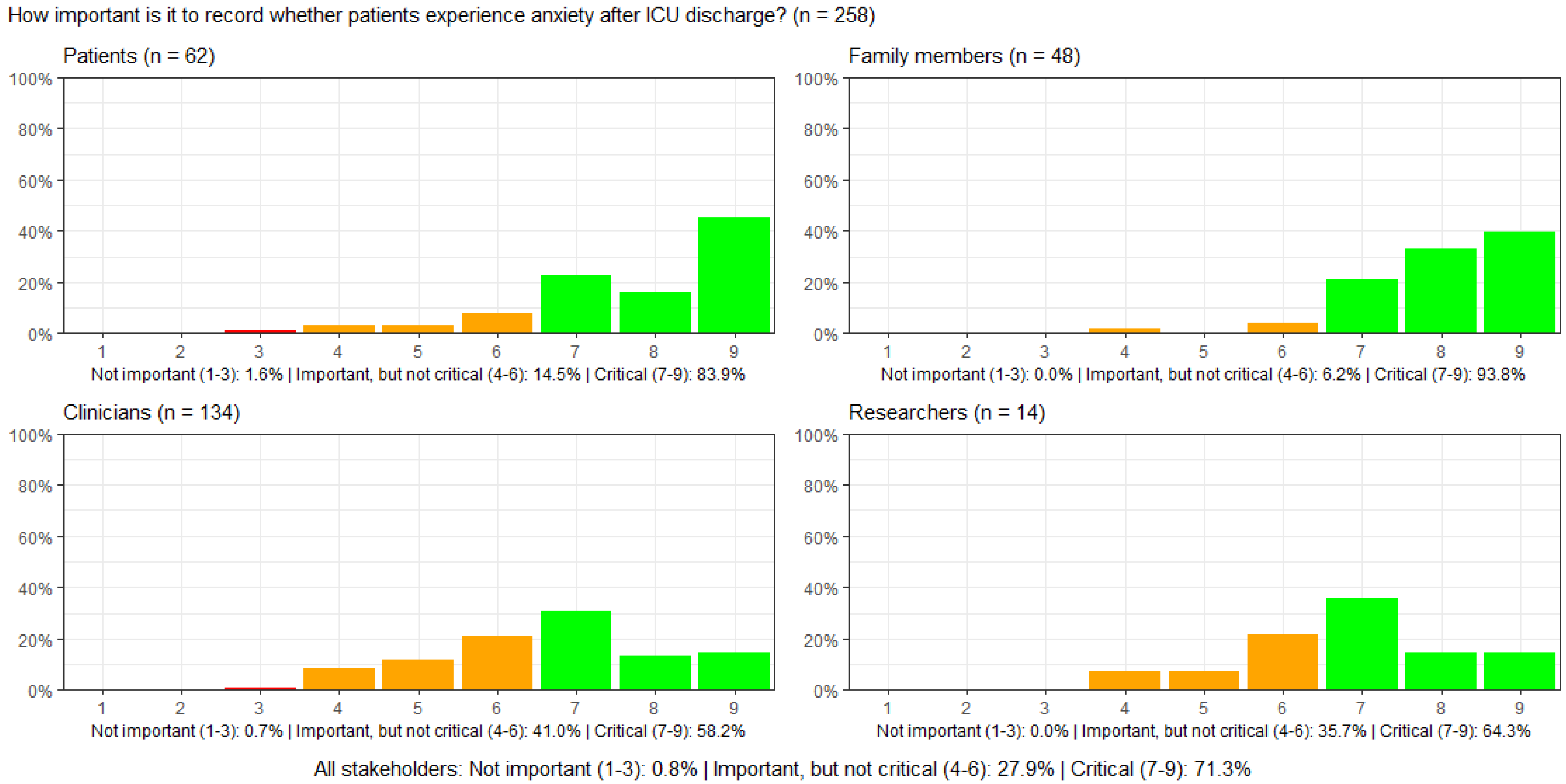

#### Figure S99: How important is it to record whether patients experience depression after ICU discharge?

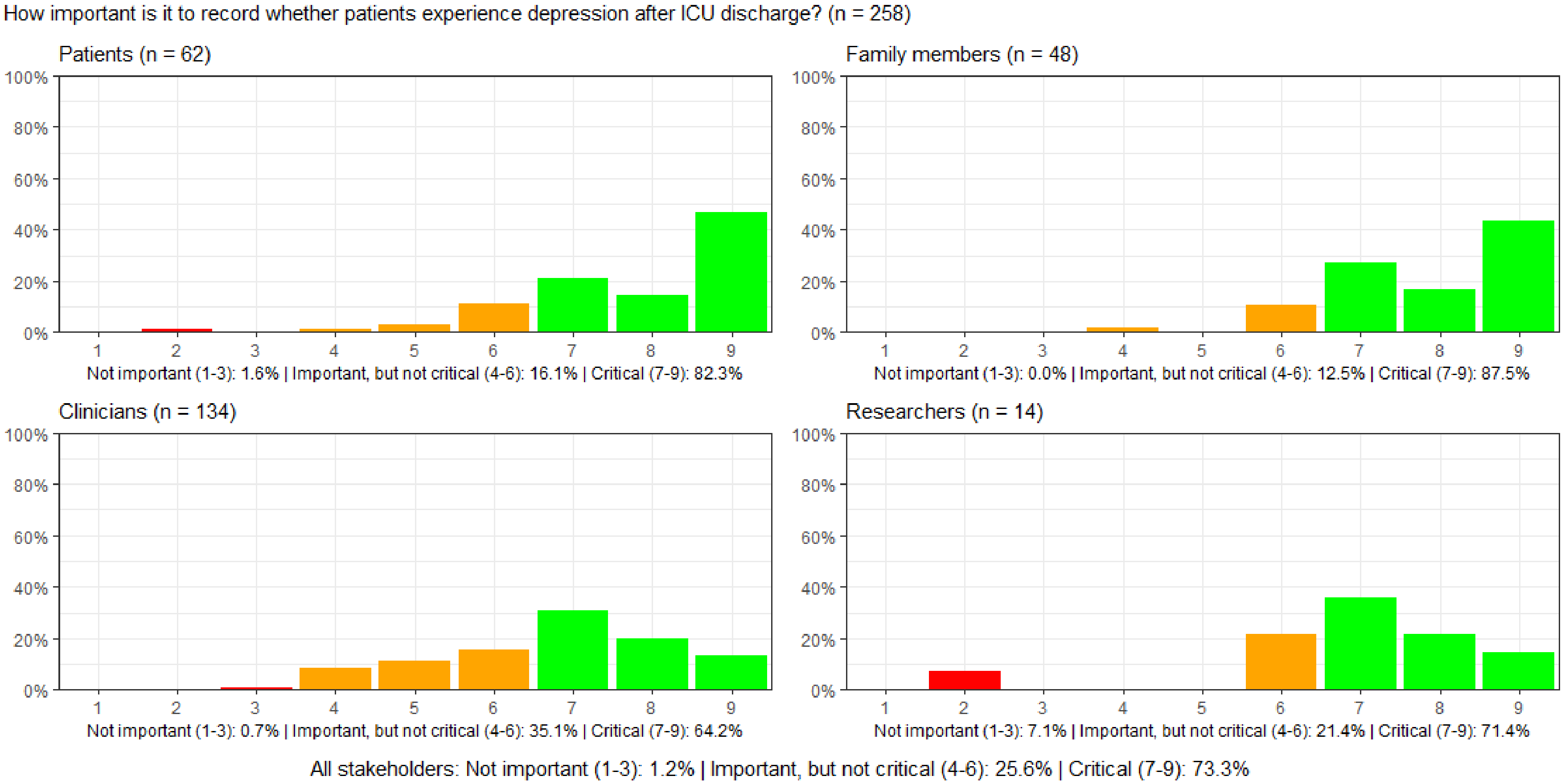

#### Figure S100: How important is it to record whether patients experience nightmares after ICU discharge?

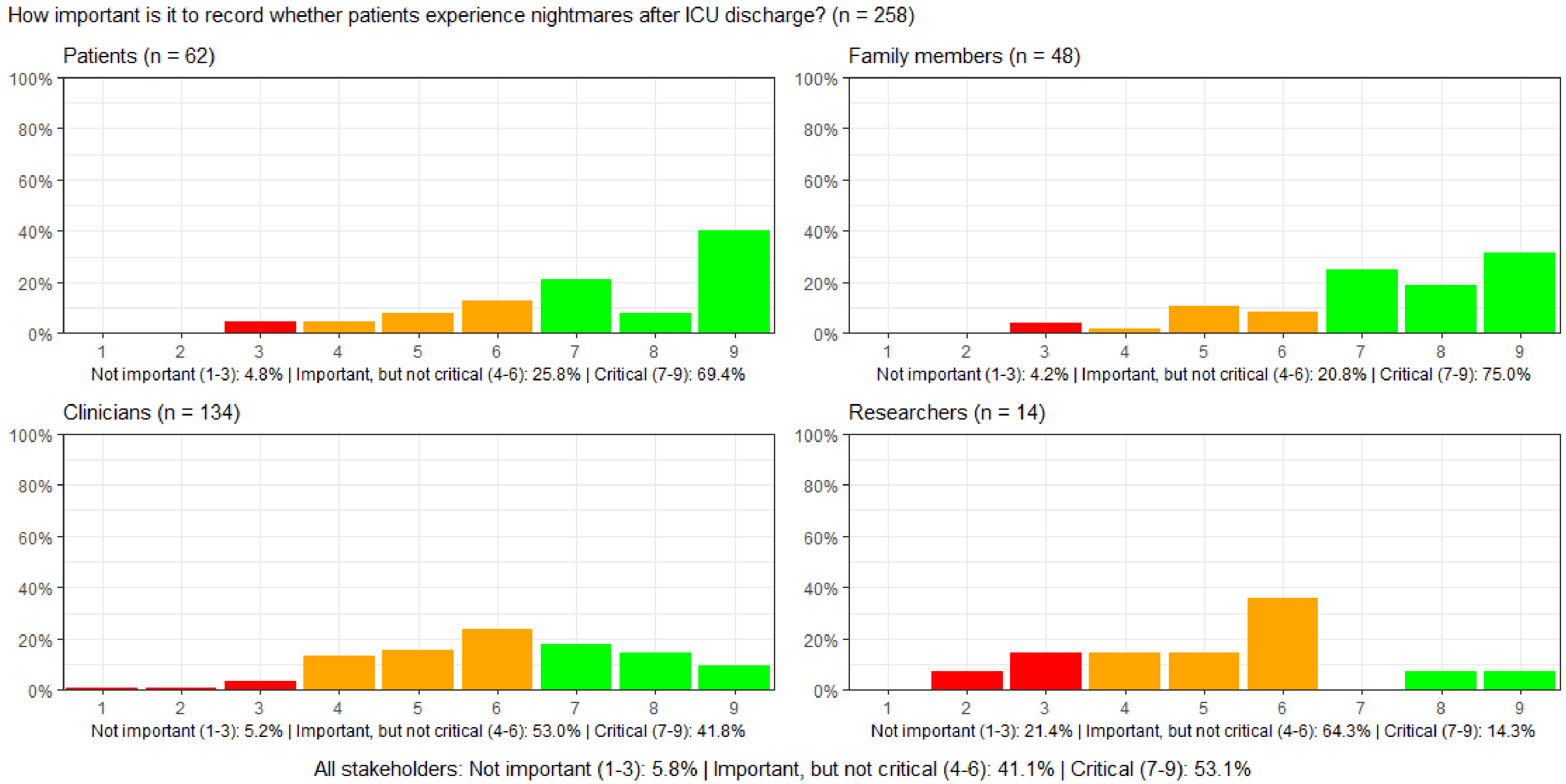

#### Figure S101-150. Graphs for the third survey round

##### Figure S101: How important is it to record ICU survival?

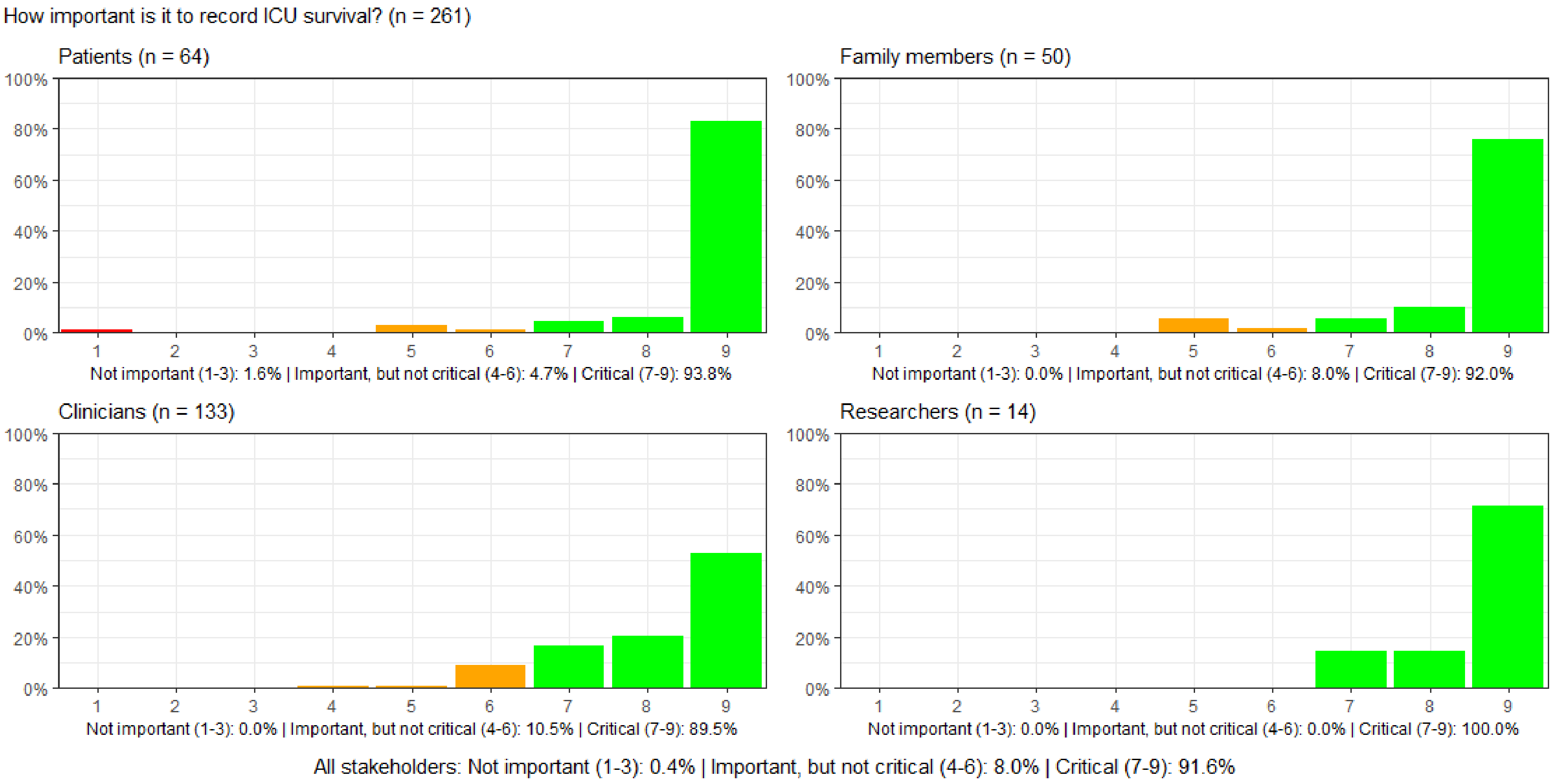

#### Figure S102: How important is it to record the ICU length of stay?

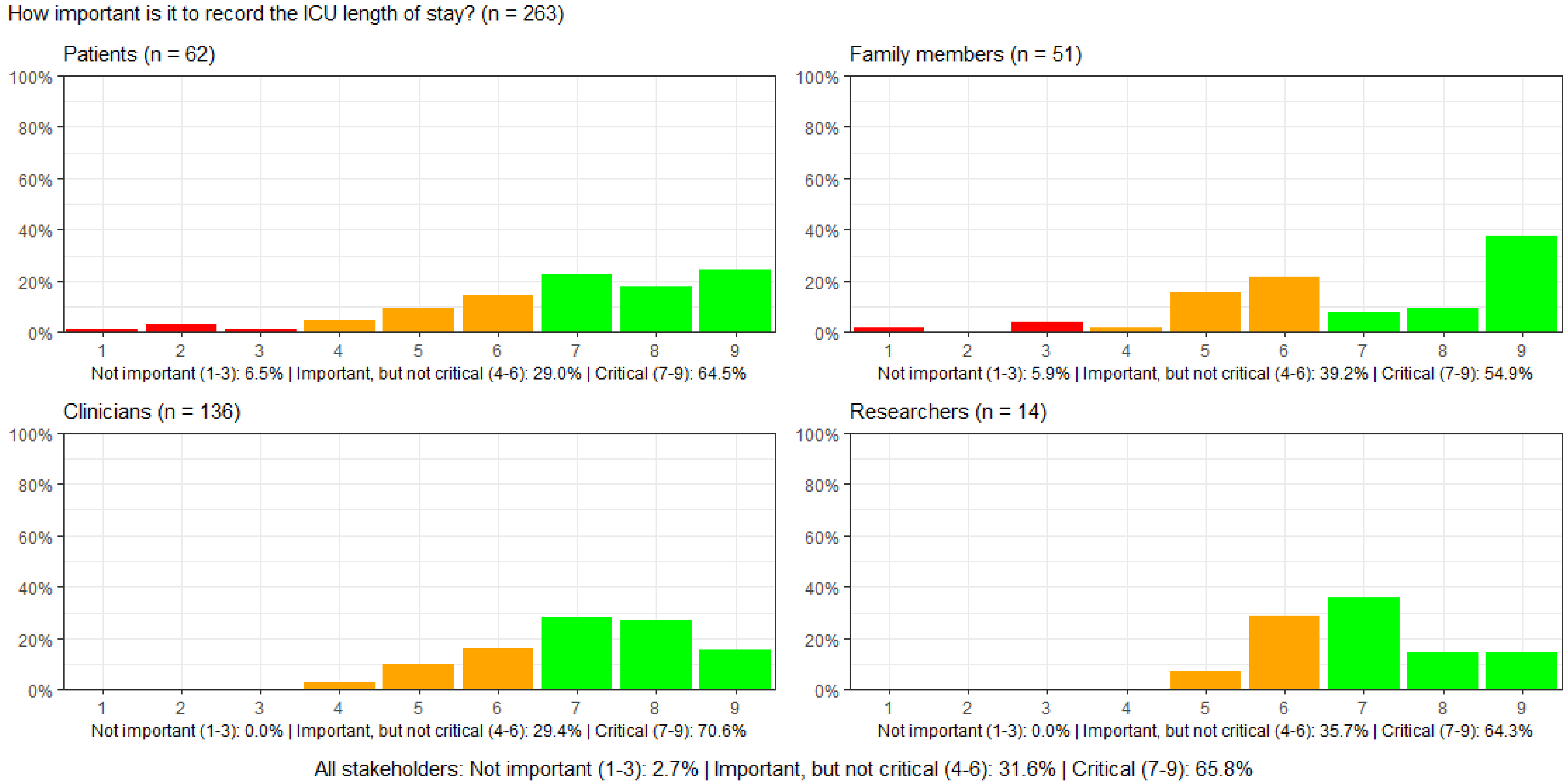

#### Figure S103: How important is it to record the hospital length of stay?

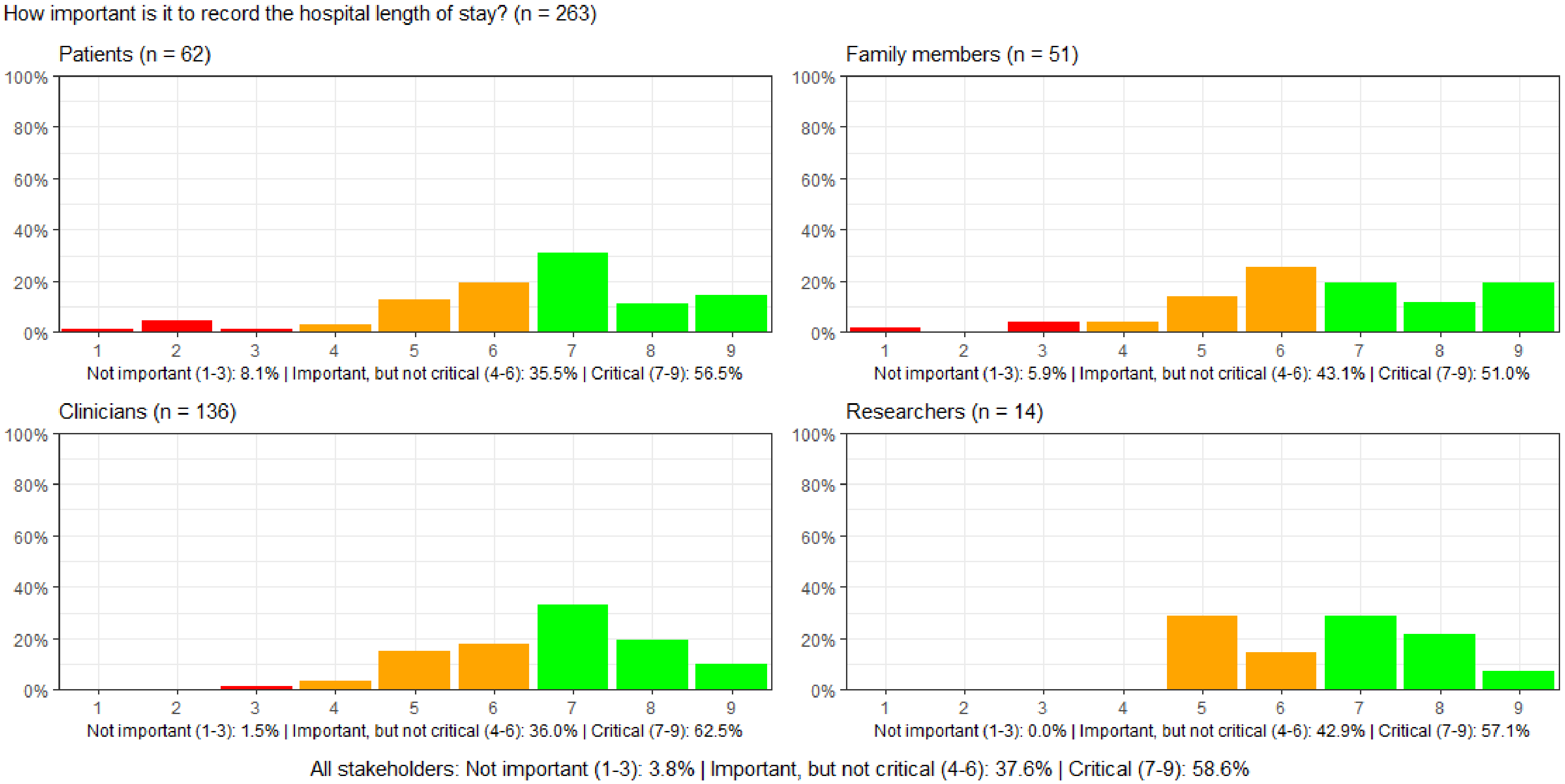

#### Figure S104: How important is it to record the ICU readmission rate during a hospital stay?

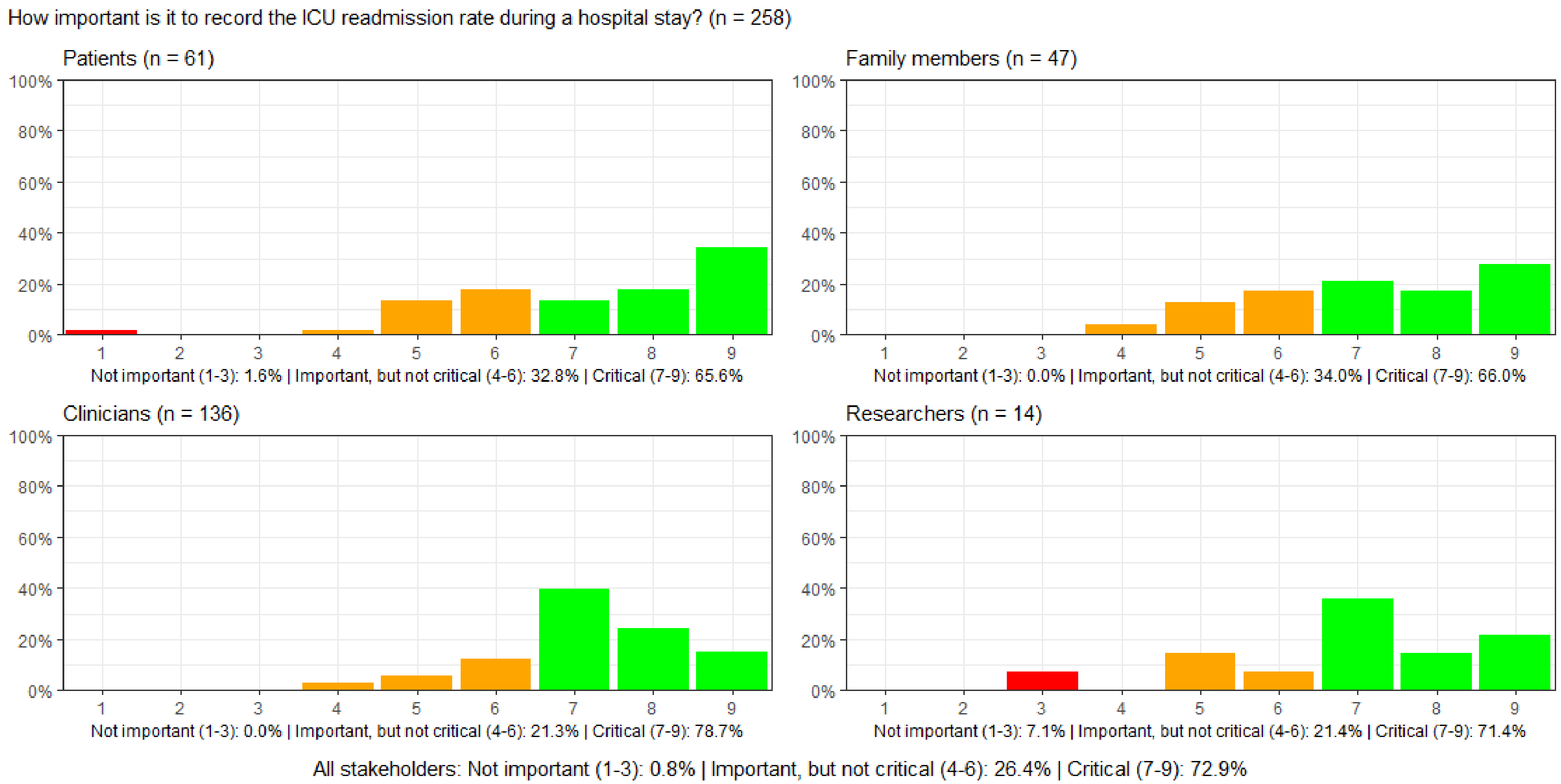

#### Figure S105: How important is it to record hospital readmission?

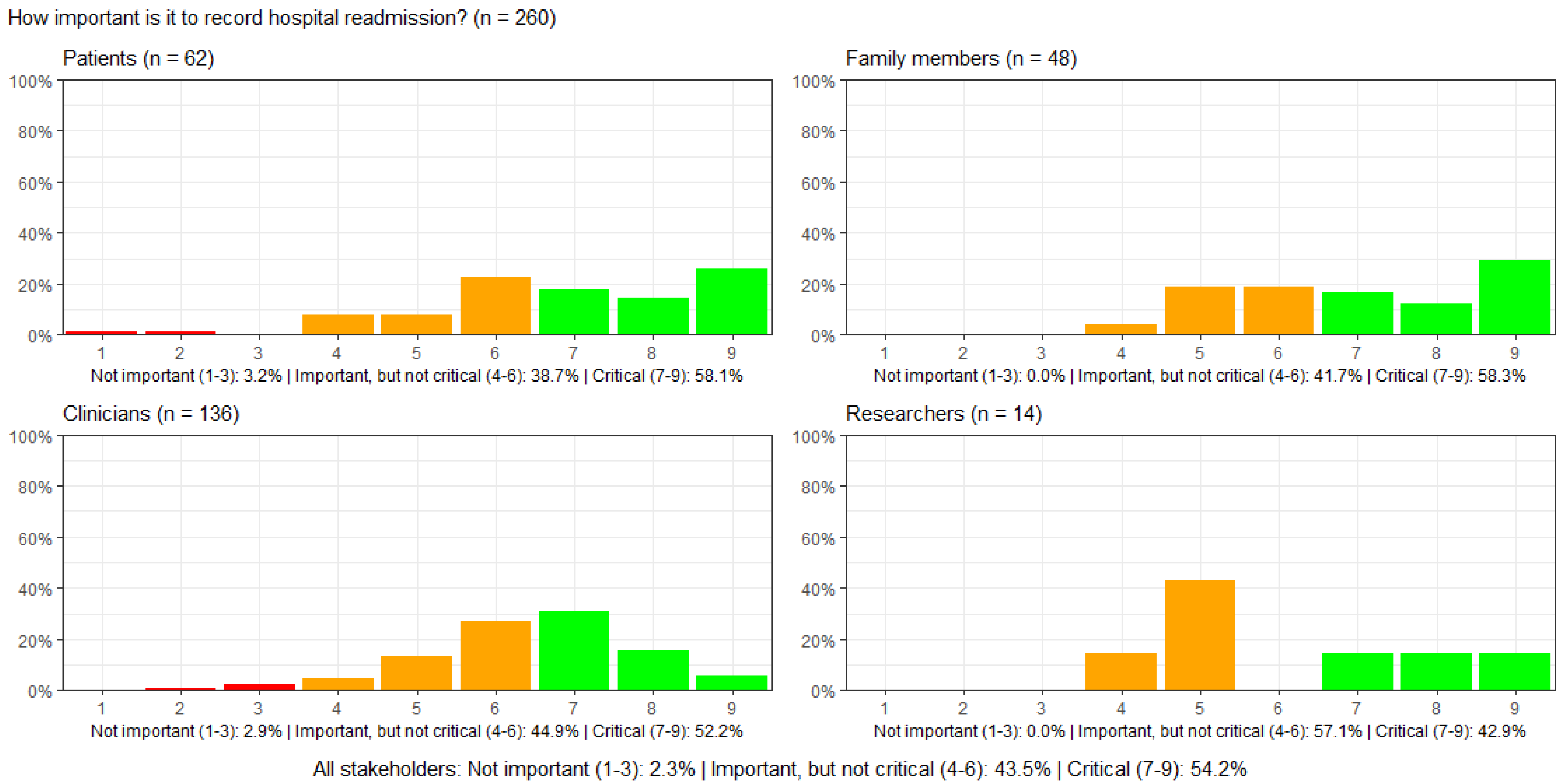

#### Figure S106: How important is it to record patients' overall well-being?

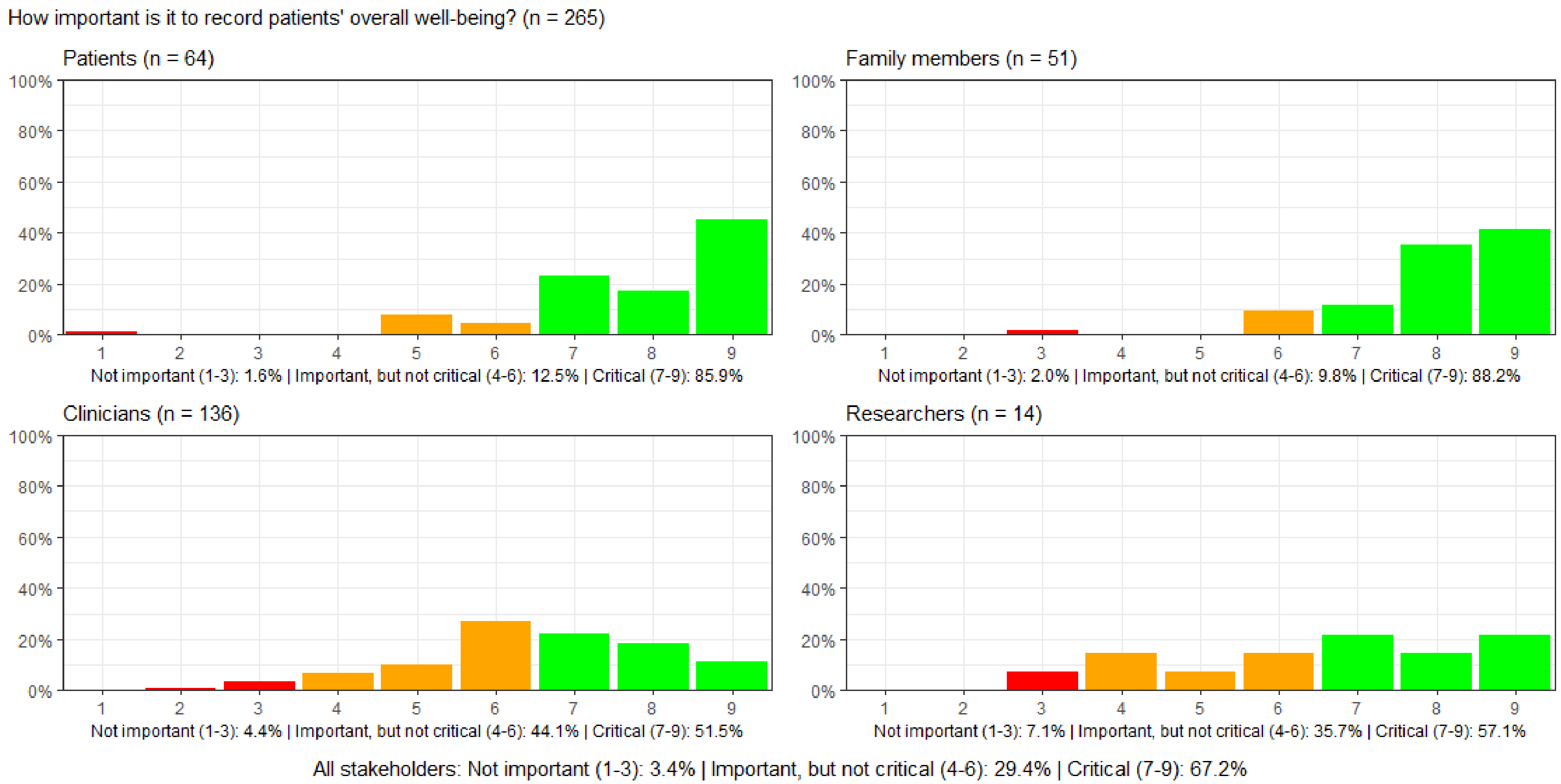

#### Figure S107: How important is it to measure the health economic consequences of a particular treatment in ICU?

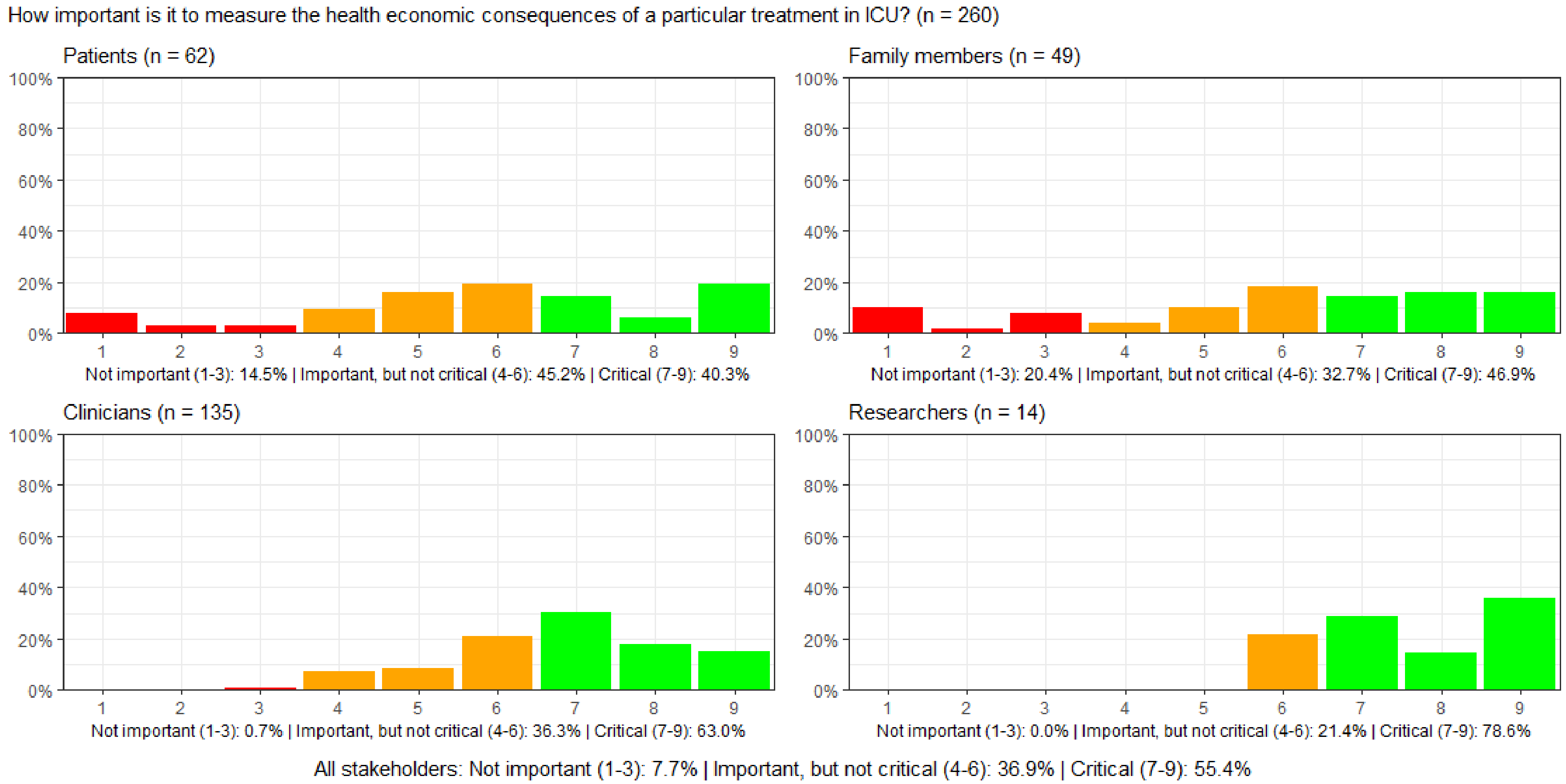

#### Figure S108: How important is it to measure the environmental consequences of a particular treatment in ICU?

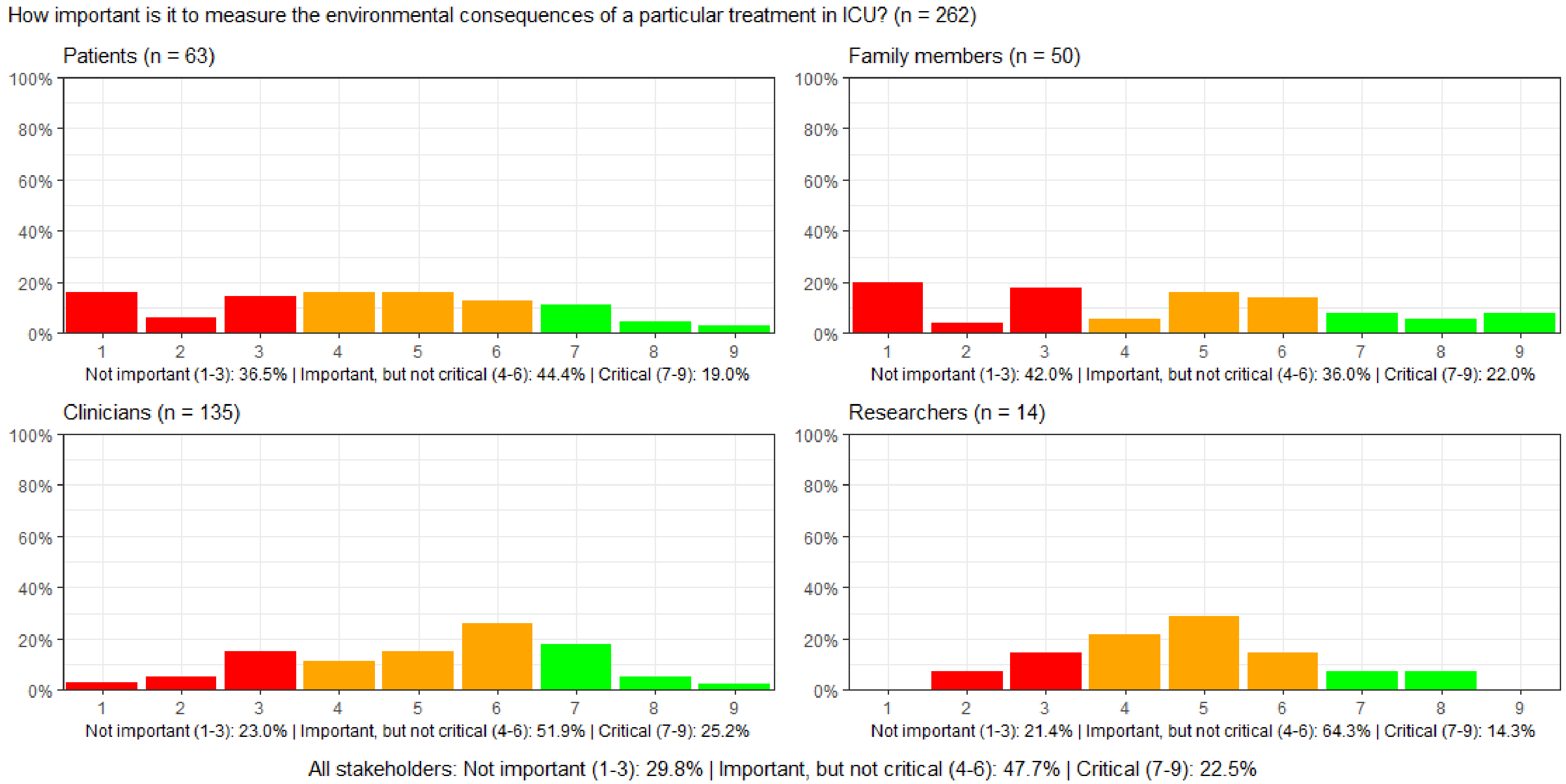

##### Figure S109: How important is it to measure whether blood values are within normal limits while in ICU?

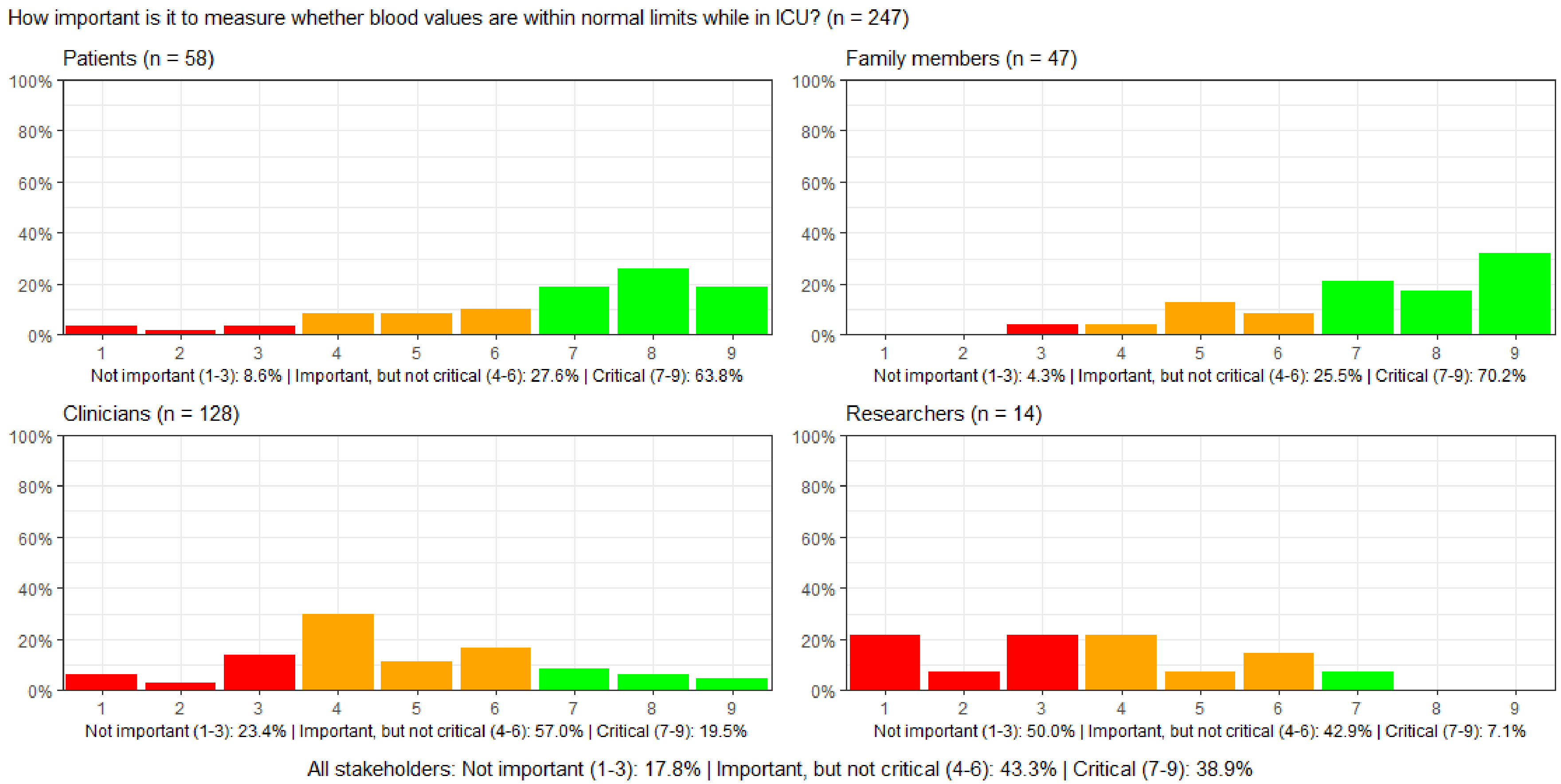

#### Figure S110: How important is it to record whether vital signs are within normal limits while in ICU?

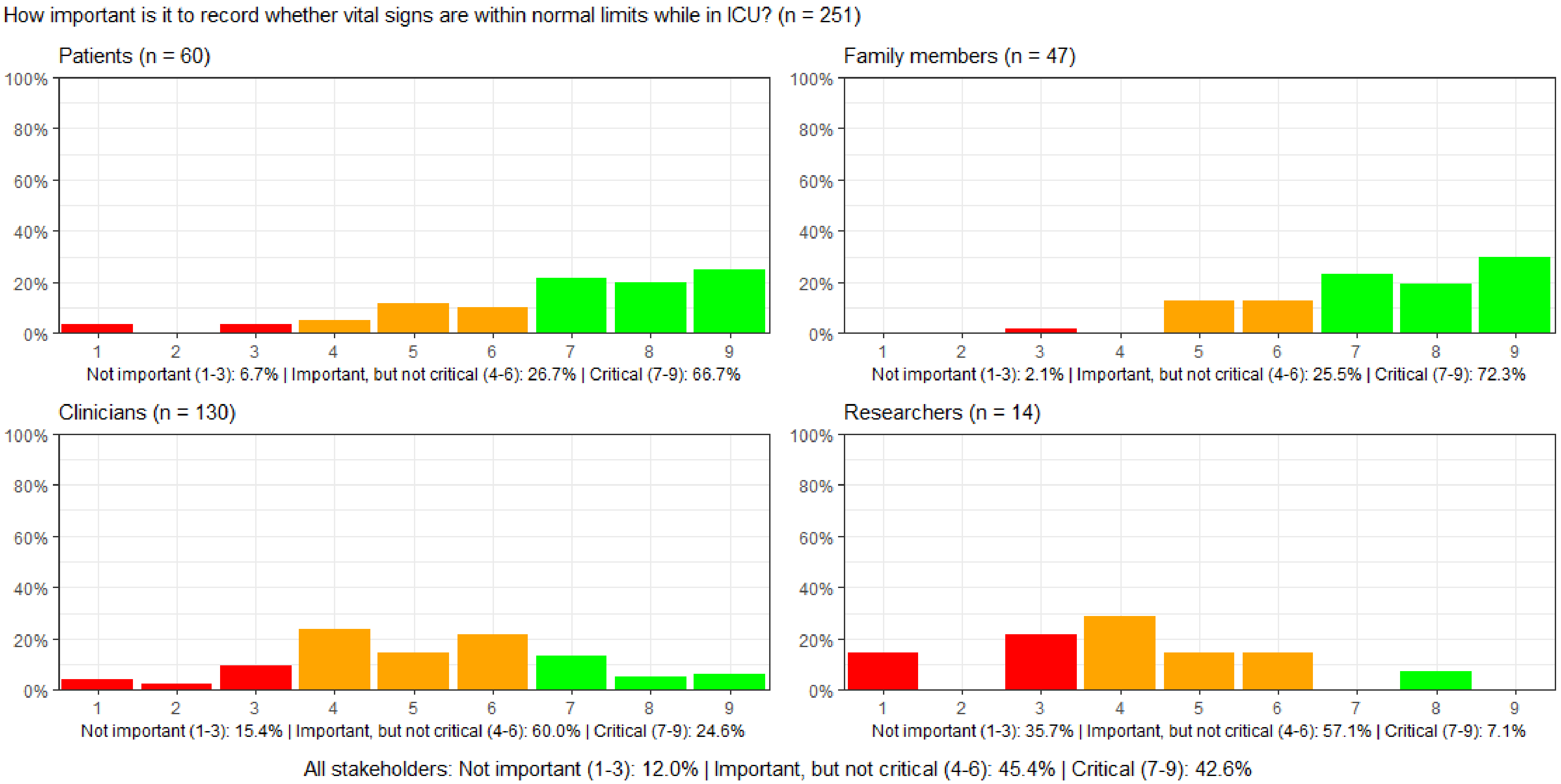

#### Figure S111: How important is it to record the duration of mechanical ventilation while in ICU?

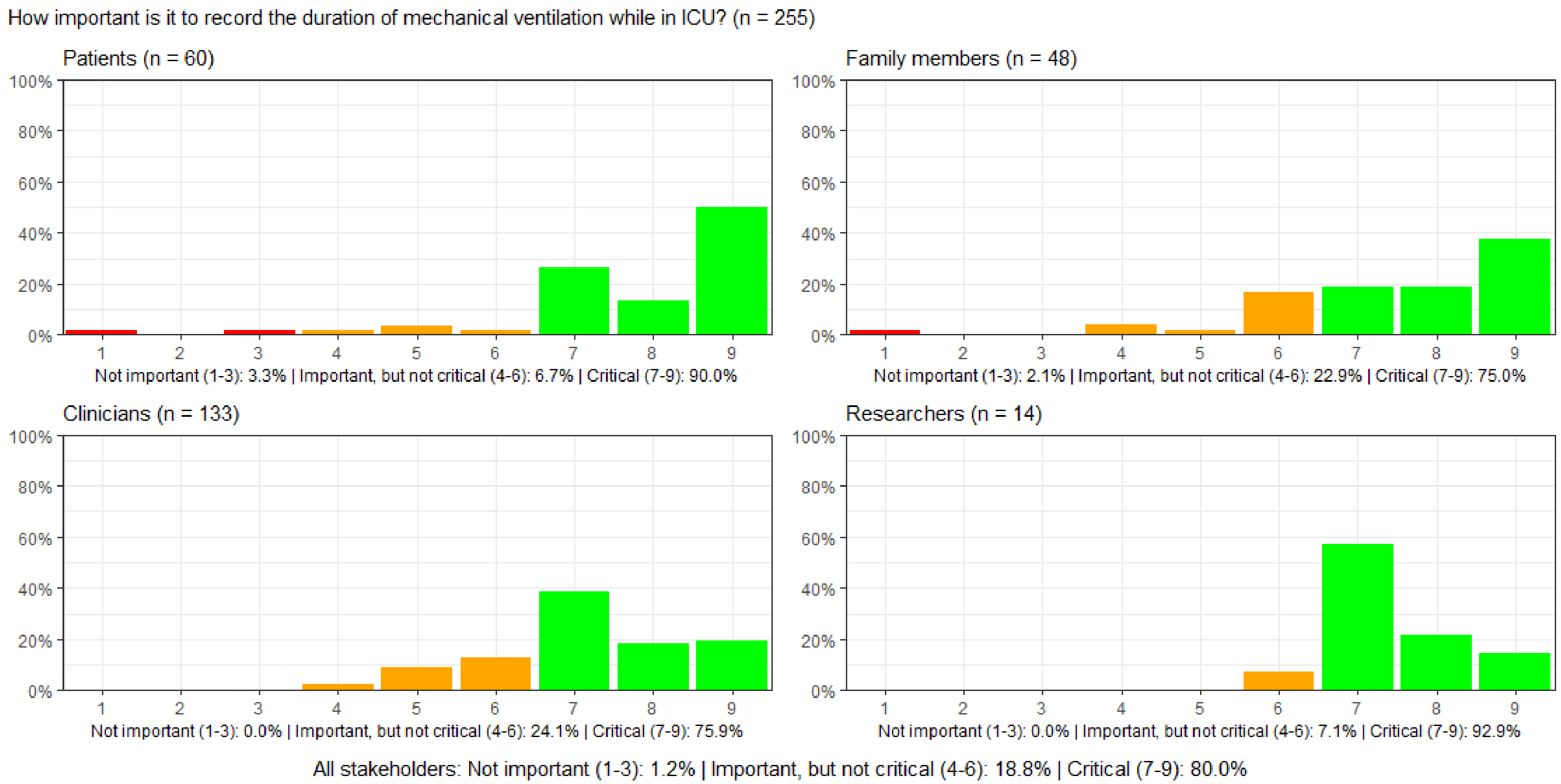

#### Figure S112: How important is it to record the duration of pharmacologic circulatory support while in ICU?

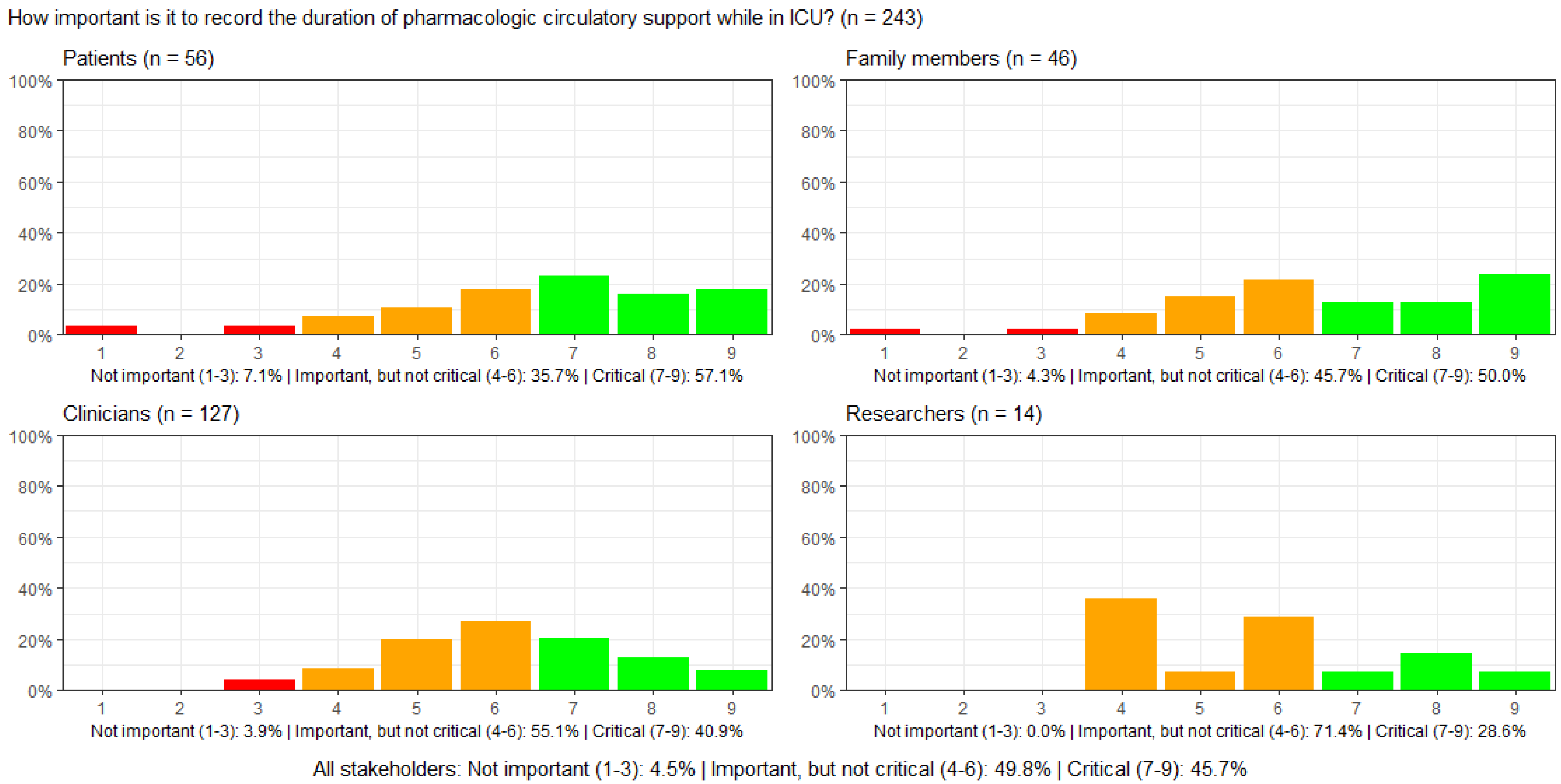

#### Figure S113: How important is it to record the duration of dialysis while in ICU?

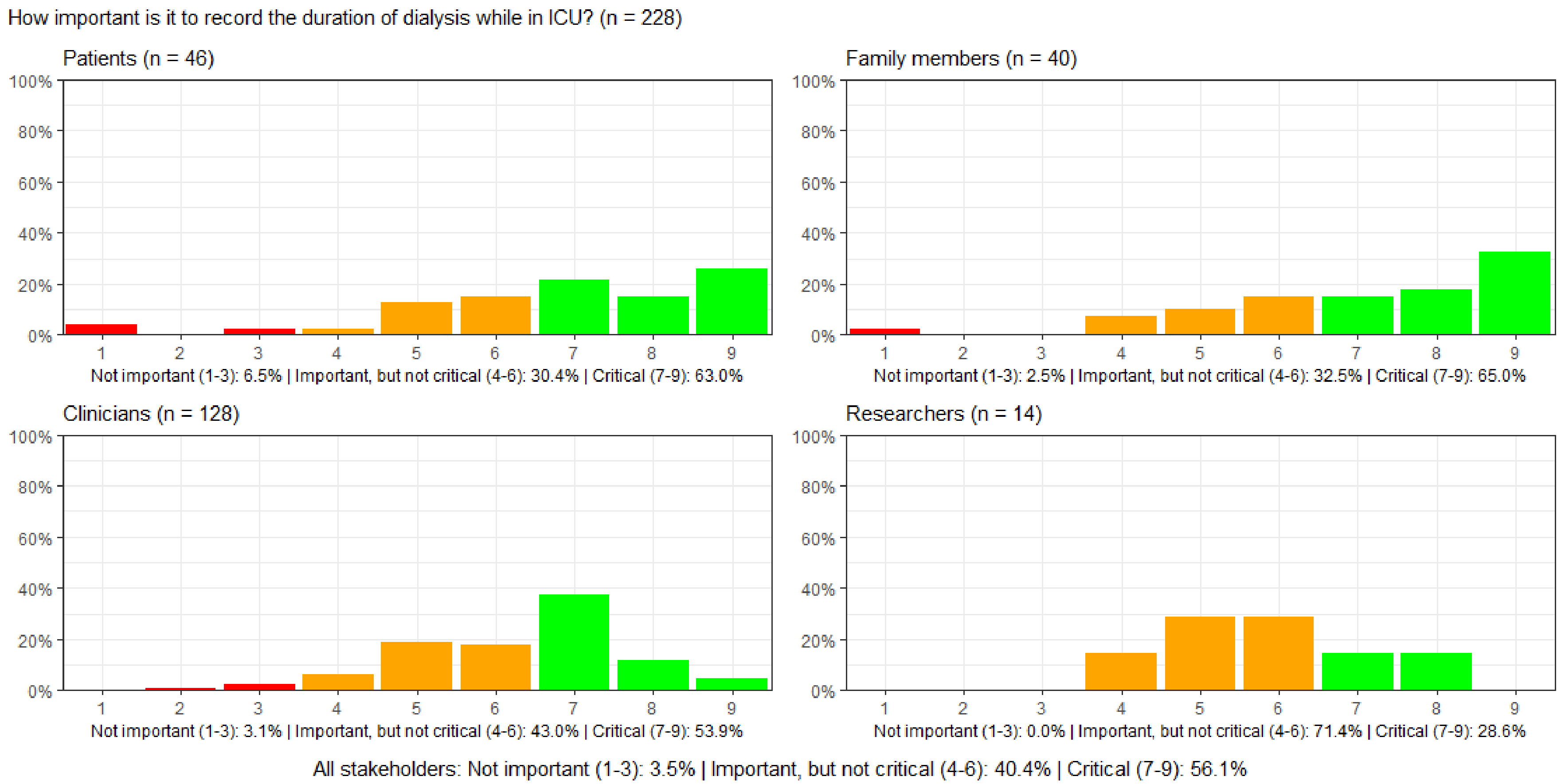

#### Figure S114: How important is it to record the duration of sedation while in ICU?

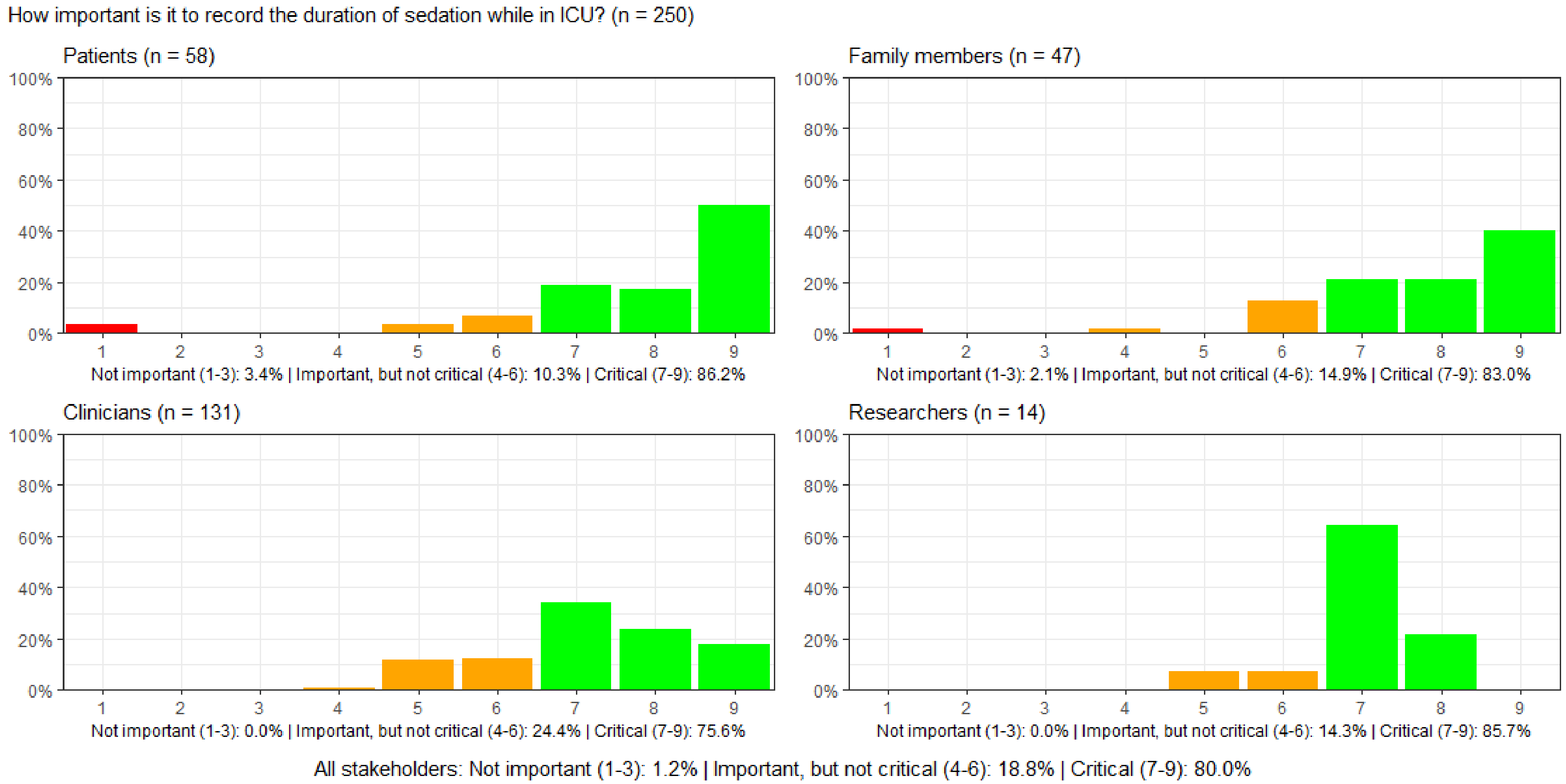

#### Figure S115: How important is it to record the duration of delirium while in ICU?

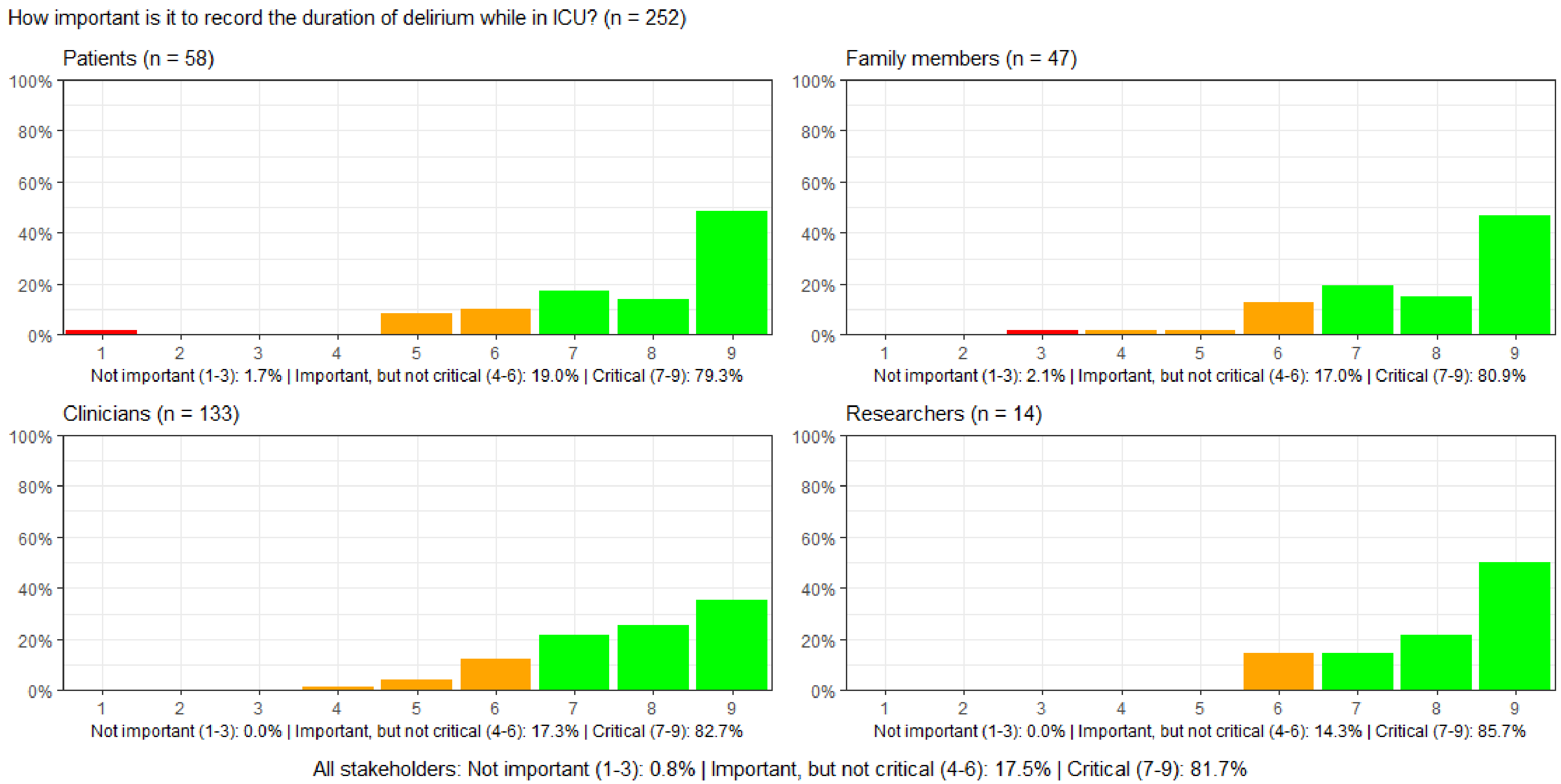

#### Figure S116: How important is it to record whether patients experience nausea and vomiting while in ICU?

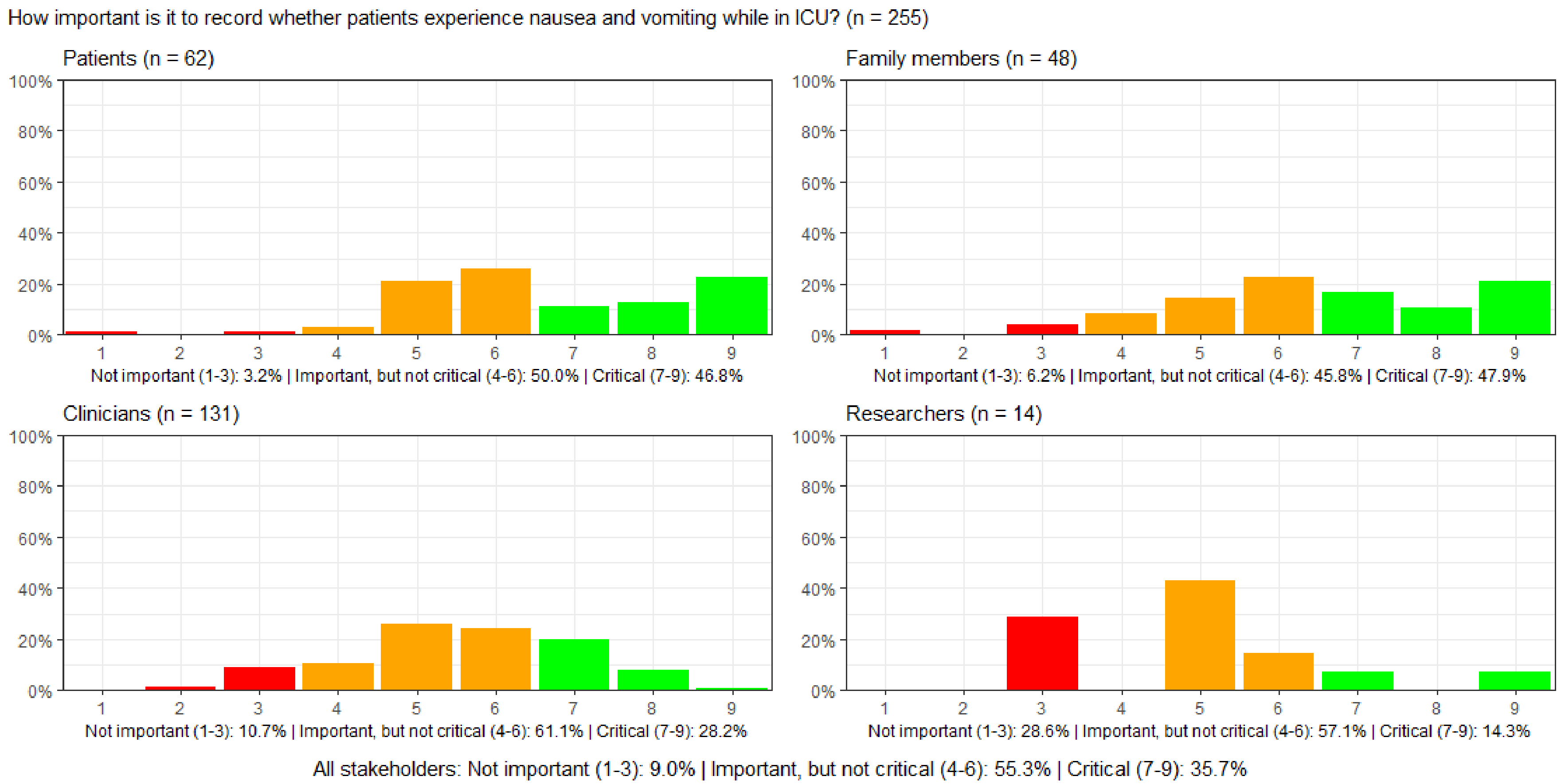

#### Figure S117: How important is it to record whether patients experience thirst while in ICU?

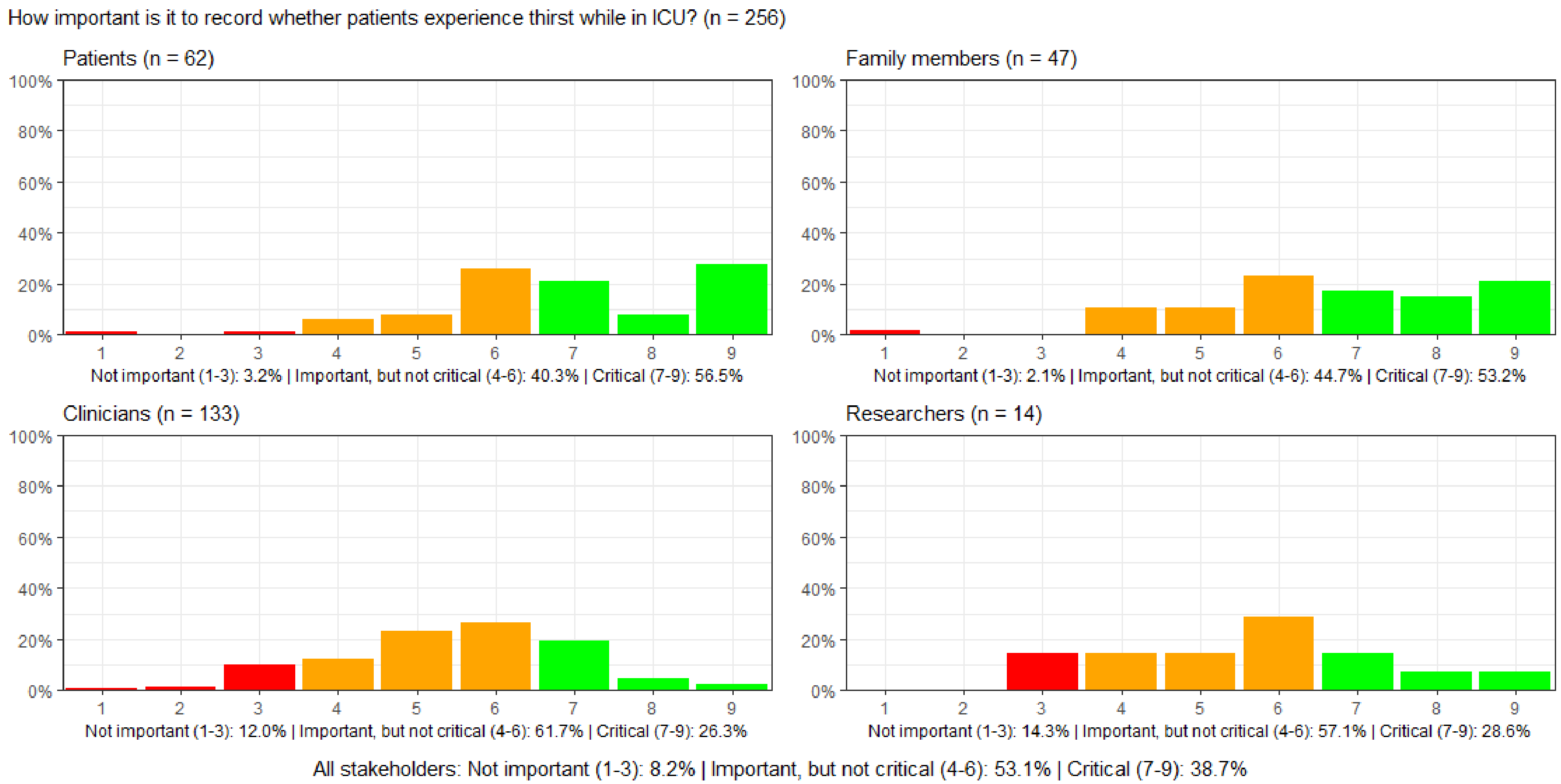

#### Figure S118: How important is it to record whether patients are short of breath while in ICU.

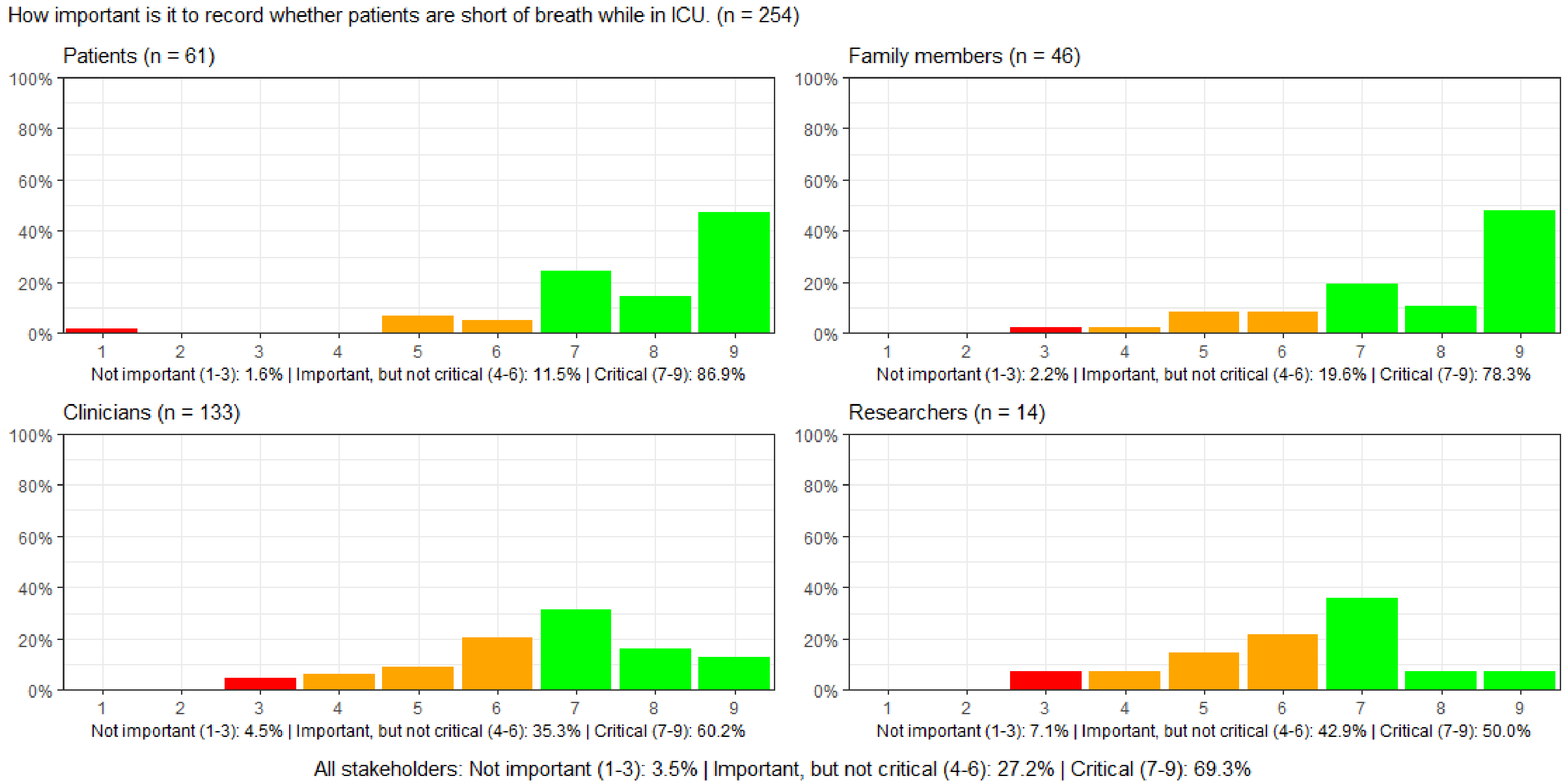

#### Figure S119: How important is it to record whether patients experience pain while in ICU?

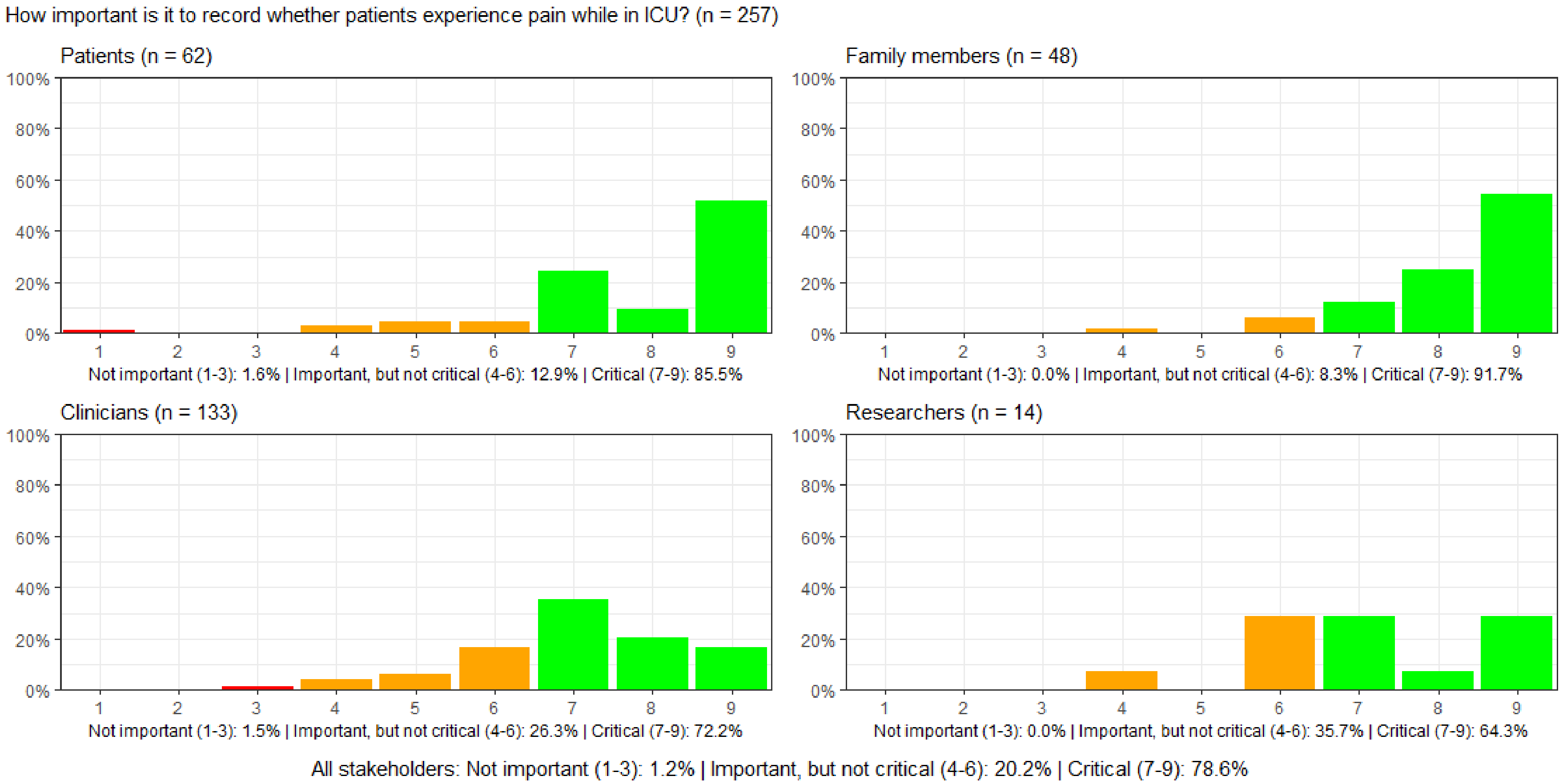

##### Figure S120: How important is it to record whether patients experience anxiety while in ICU?

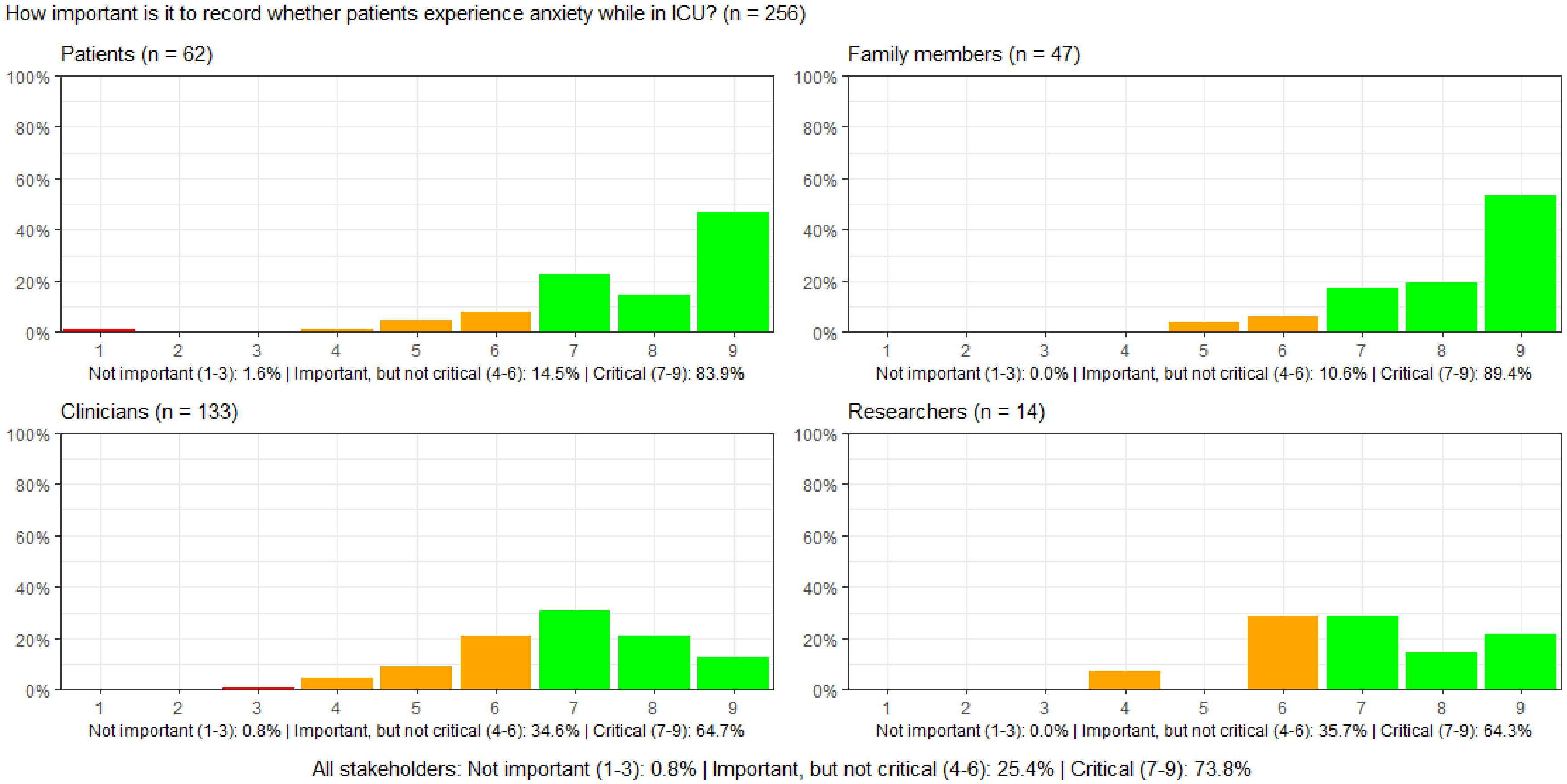

#### Figure S121: How important is it to record patients' sleep while in ICU?

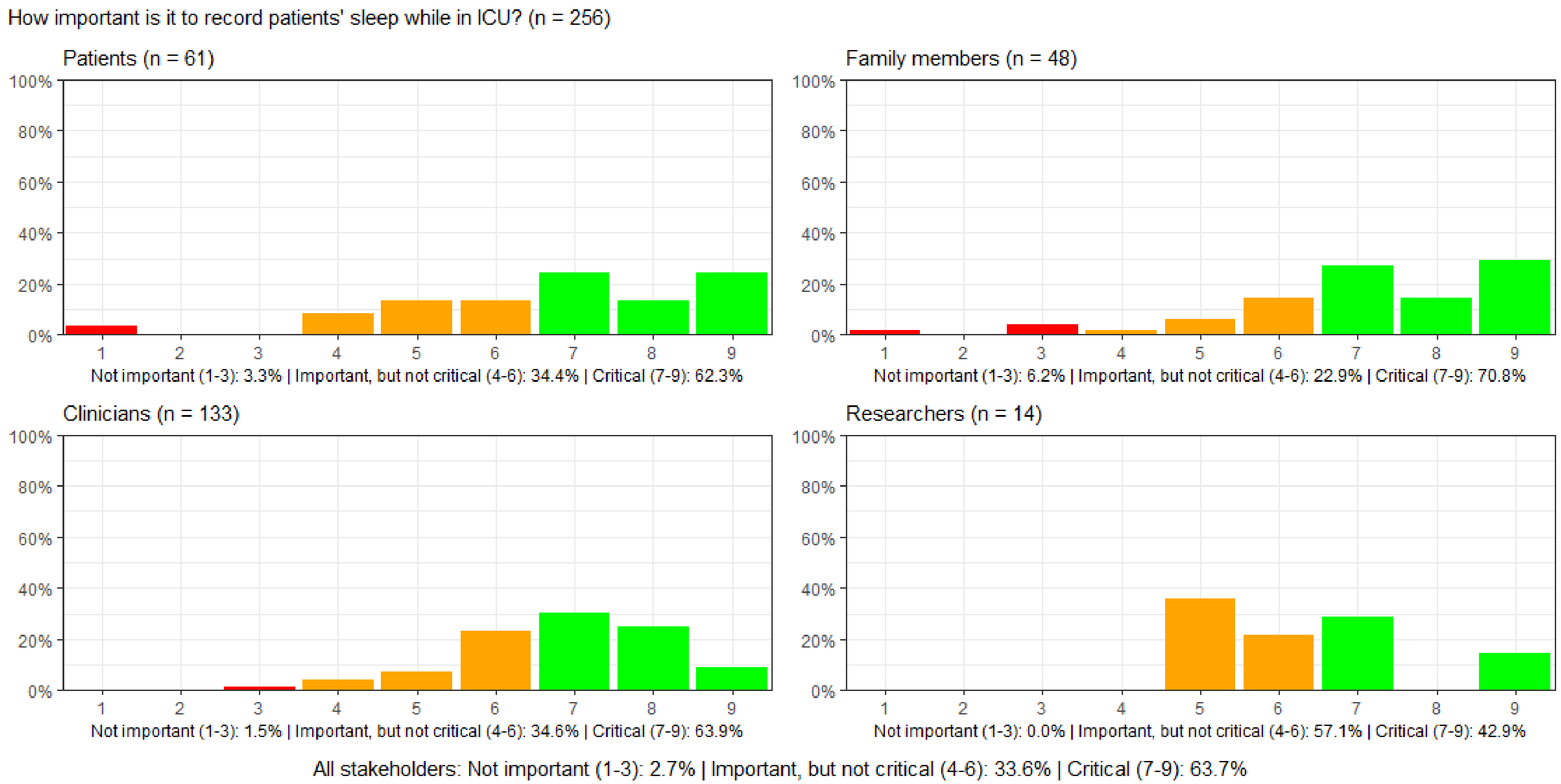

#### Figure S122: How important is it to record patients' ability to communicate while in ICU?

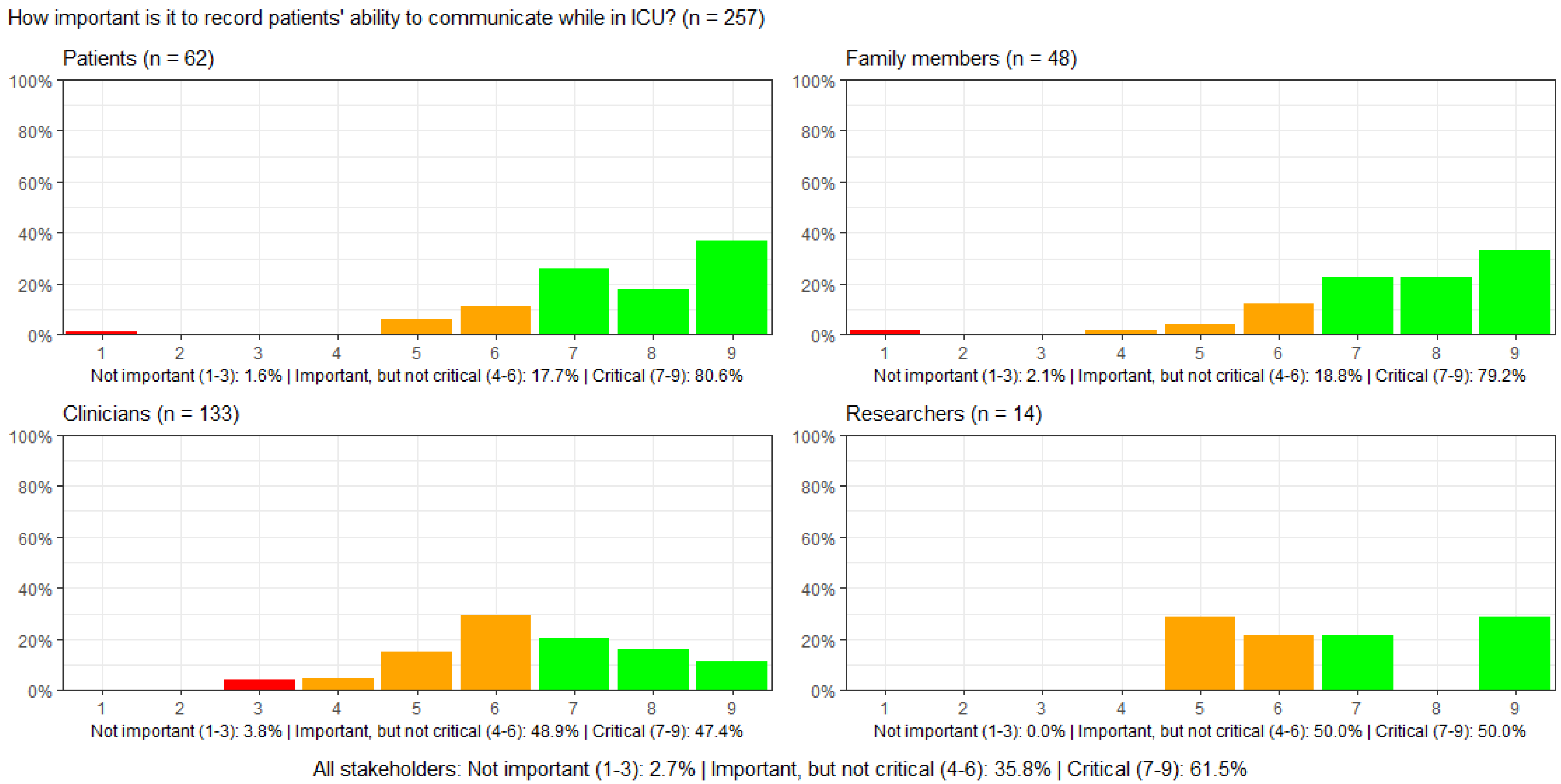

#### Figure S123: How important is it to record patients' discharge location after hospitalization?

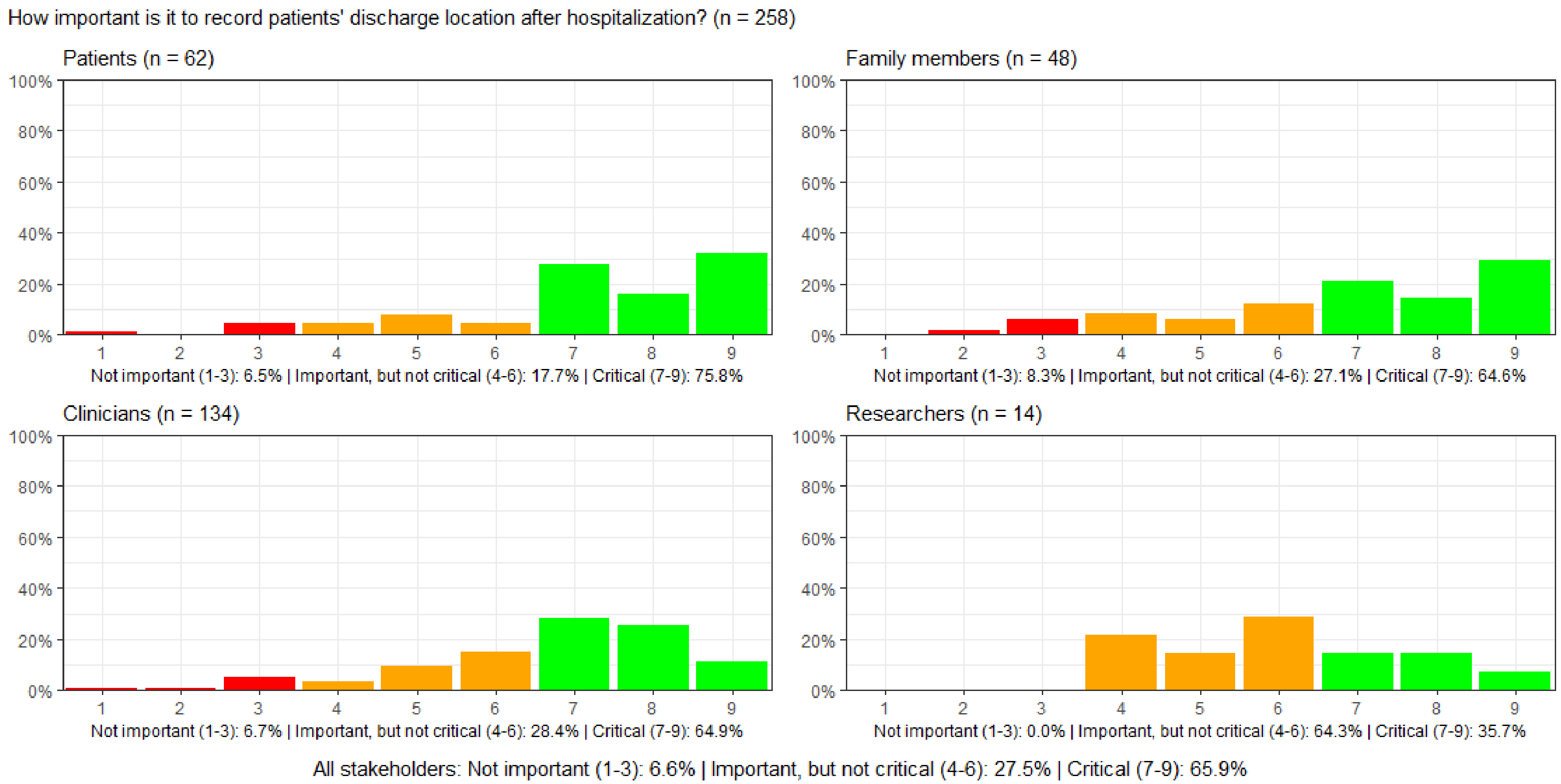

#### Figure S124: How important is it to record patients' outpatient or general practitioner visits following hospitalization?

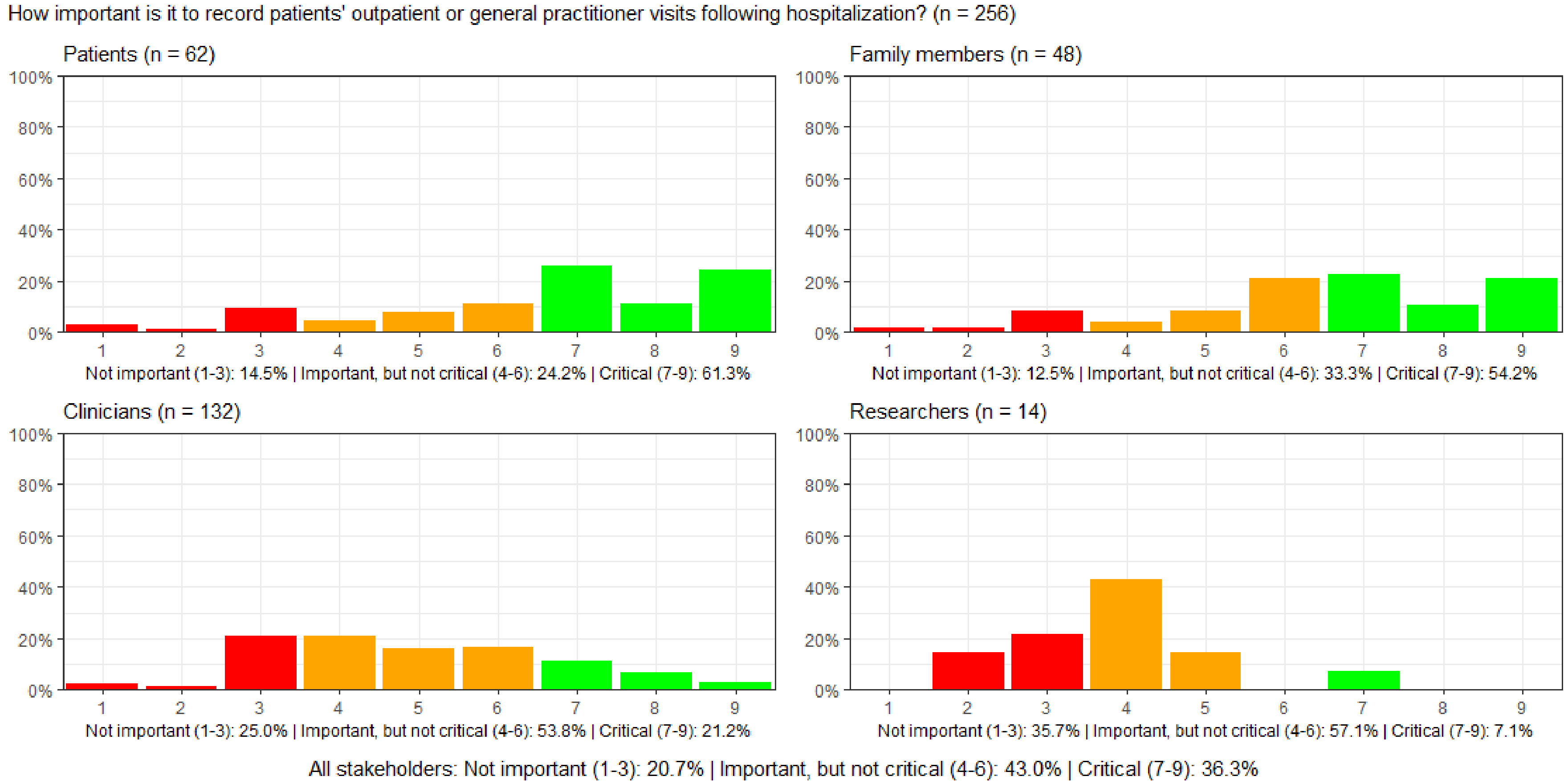

#### Figure S125: How important is it to record whether new medications are prescribed for the patient after ICU discharge?

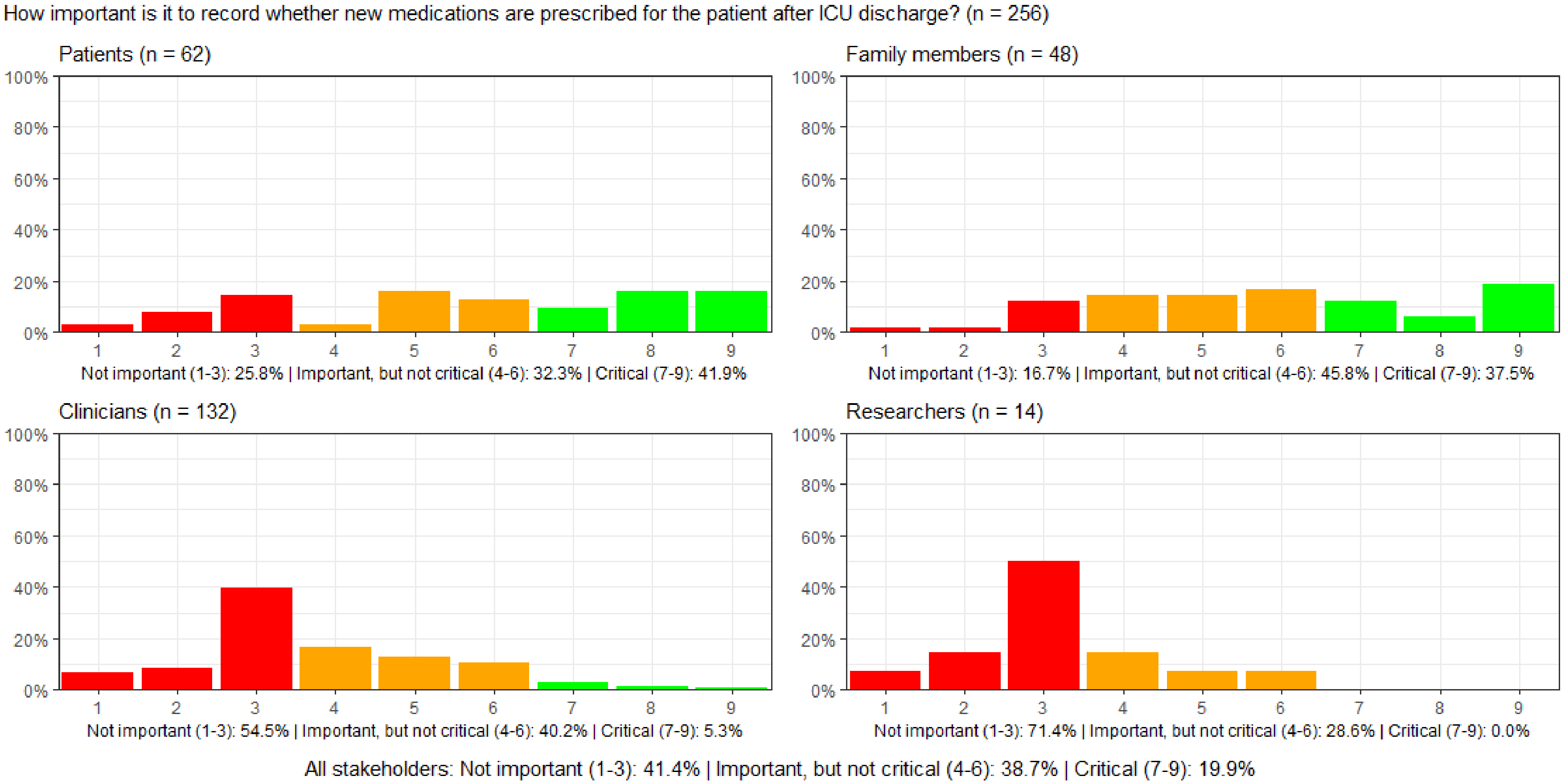

#### Figure S126: How important is it to record patients' health-related quality of life after ICU discharge?

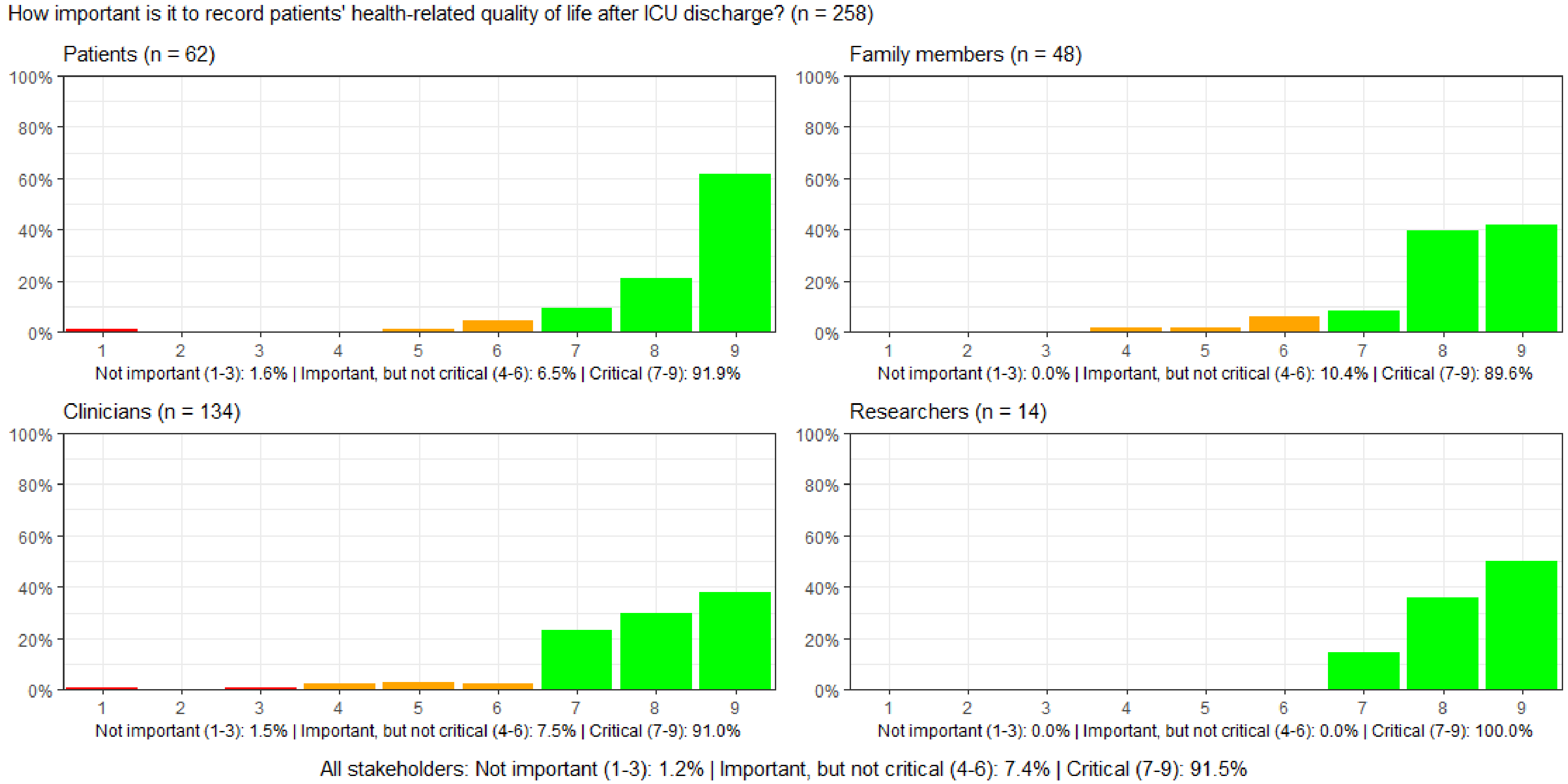

#### Figure S127: How important is it to record whether patients experience newly onset tiredness or fatigue, unrelieved by sleep or rest, after ICU discharge?

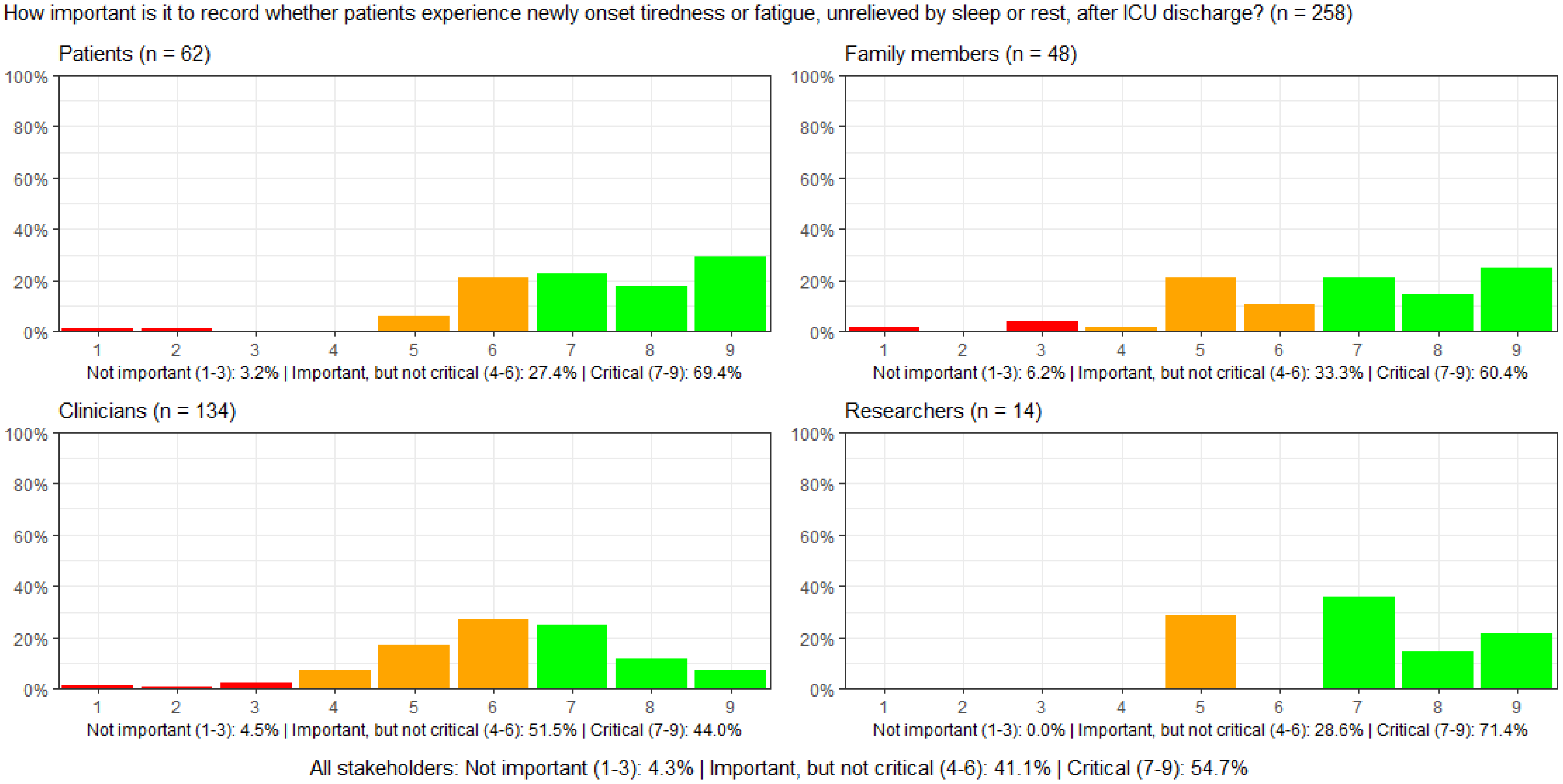

#### Figure S128: How important is it to record whether patients experience newly onset pain after ICU discharge?

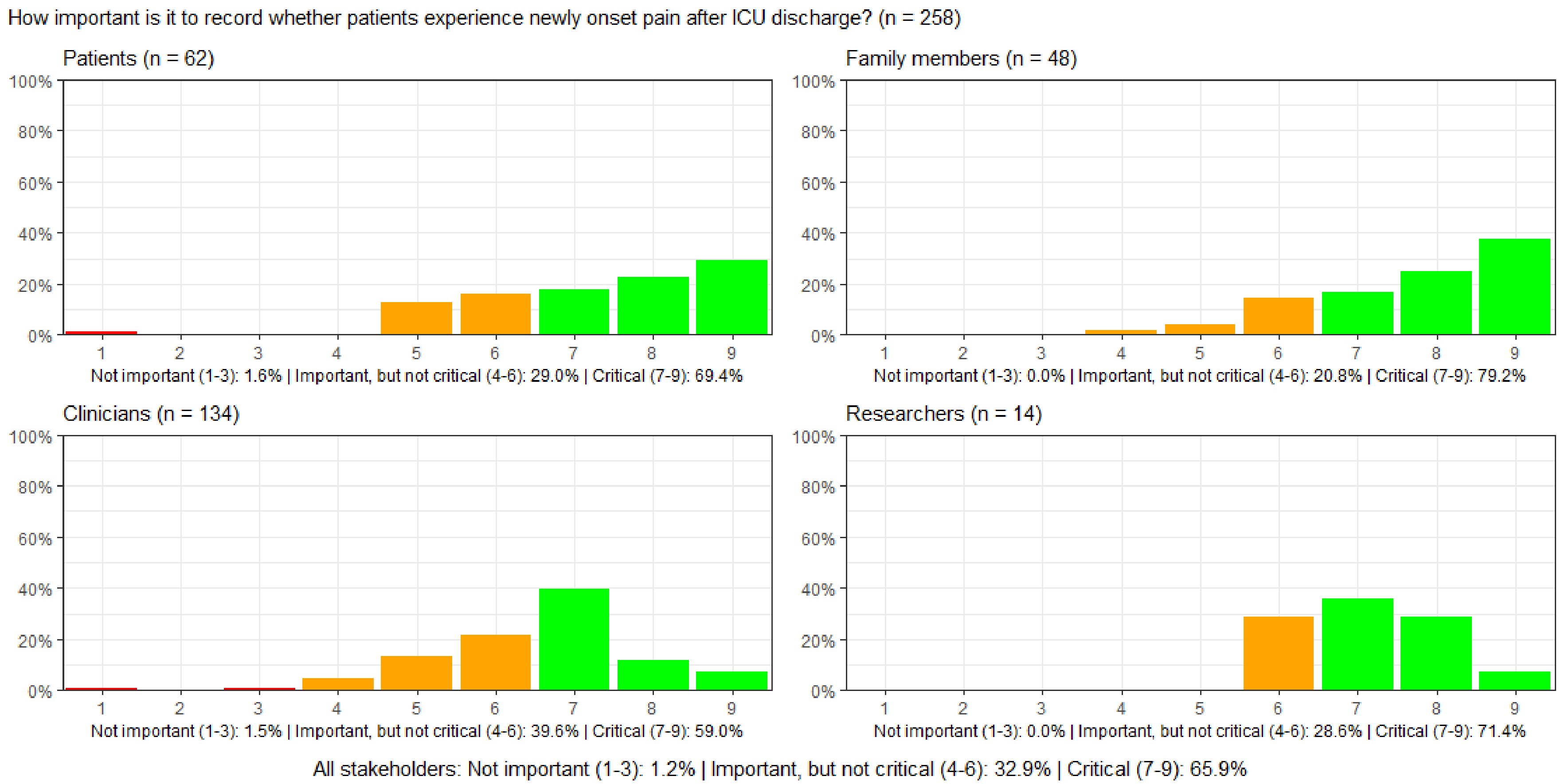

#### Figure S129: How important is it to record whether patients resume their usual roles in daily life after ICU discharge?

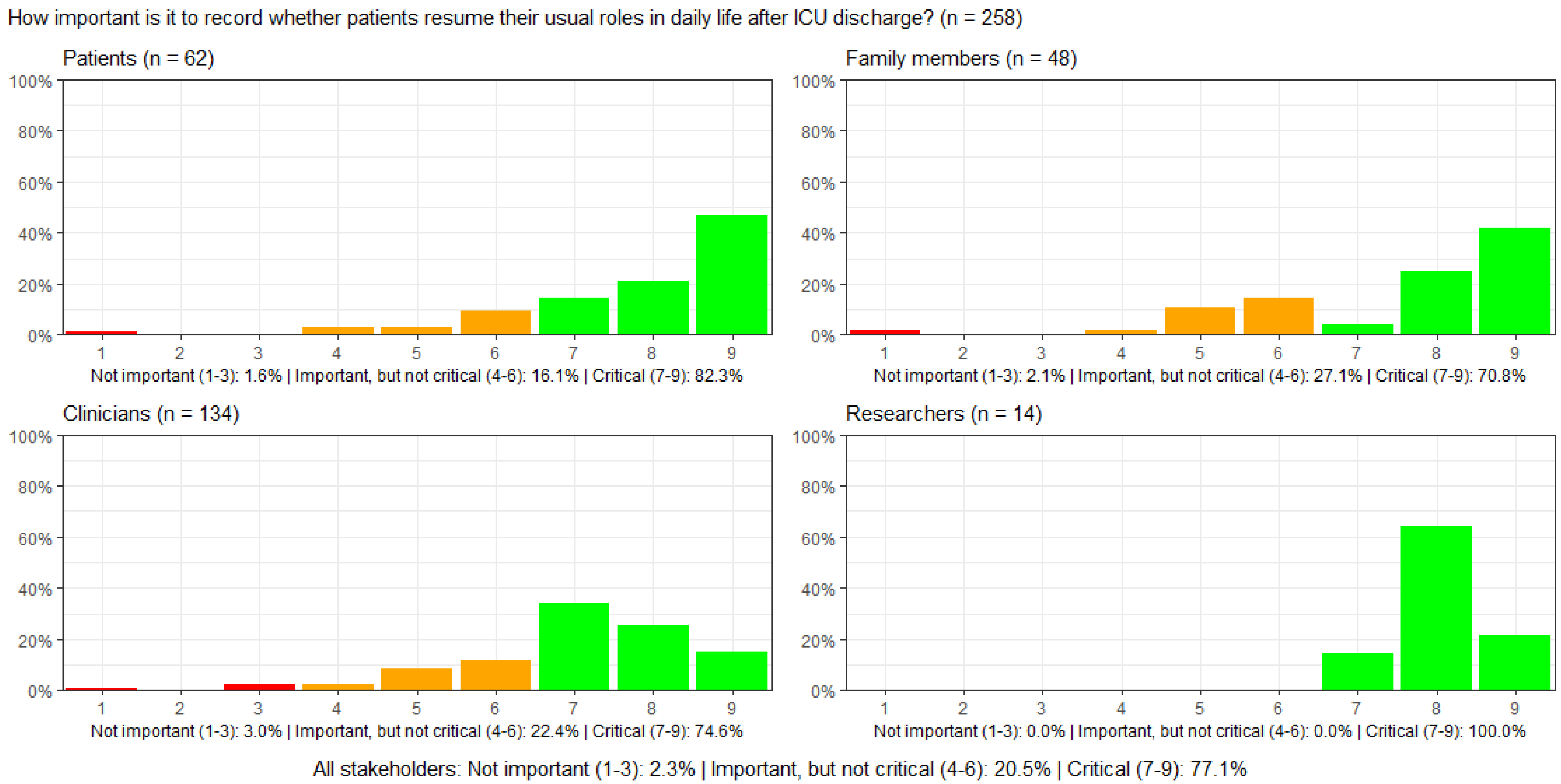

#### Figure S130: How important is it to record family caregivers' experienced quality of life after the ICU stay?

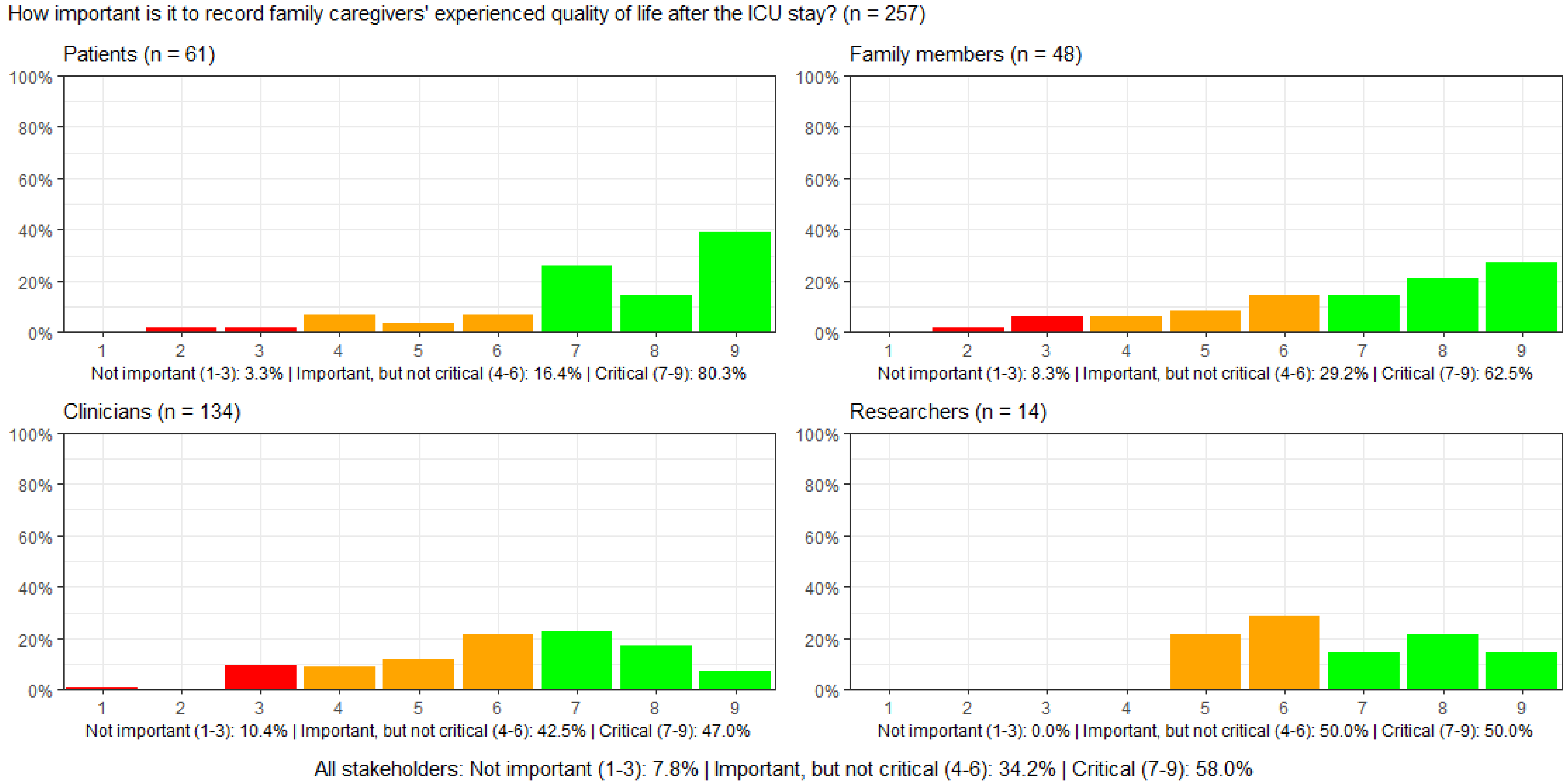

#### Figure S131: How important is it to record patients' financial capabilities after, compared to before, the ICU stay?

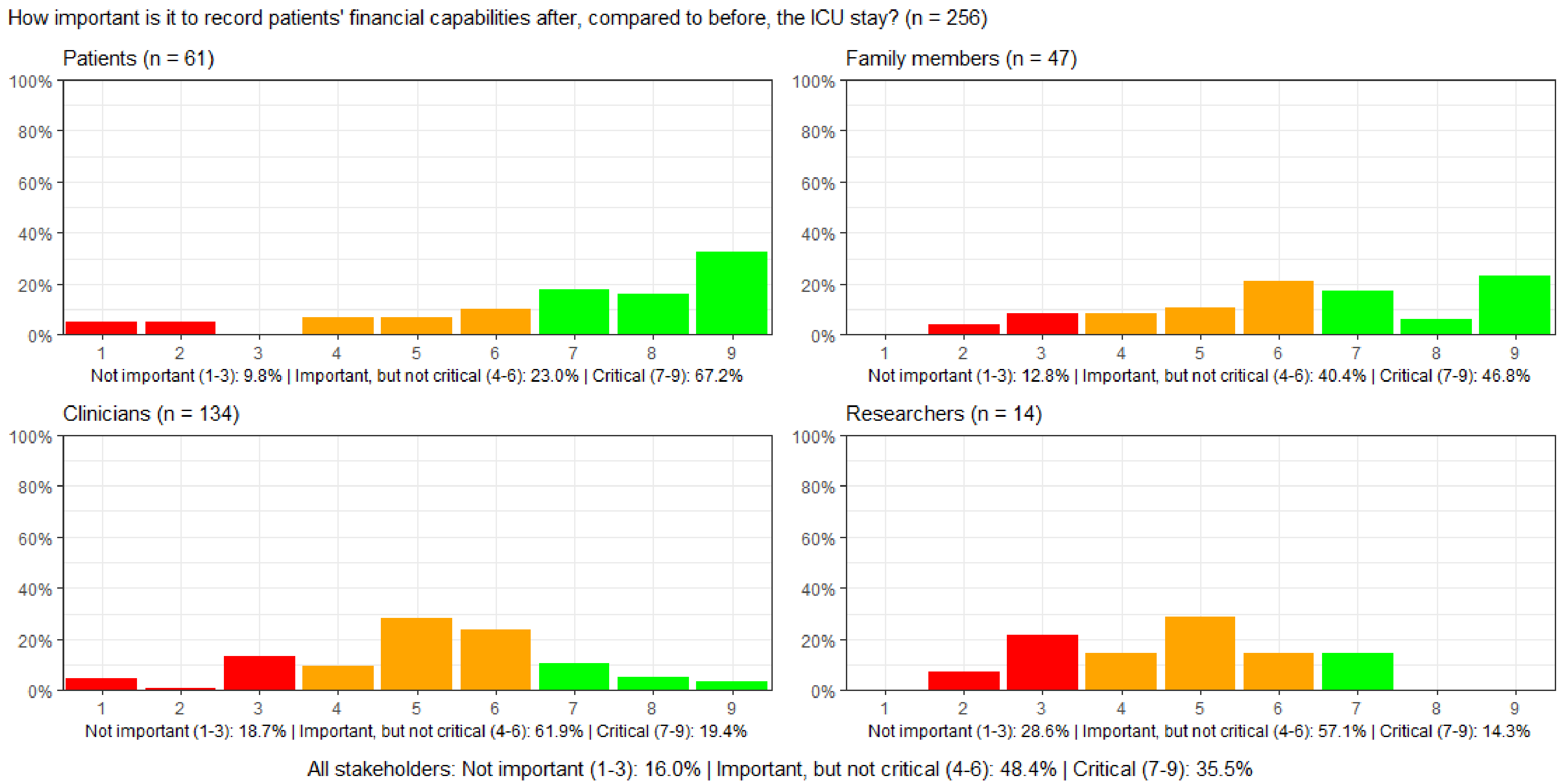

**Figure S132: How important is it to record whether patients after ICU discharge, compared to before, have the same labour market attachment.**

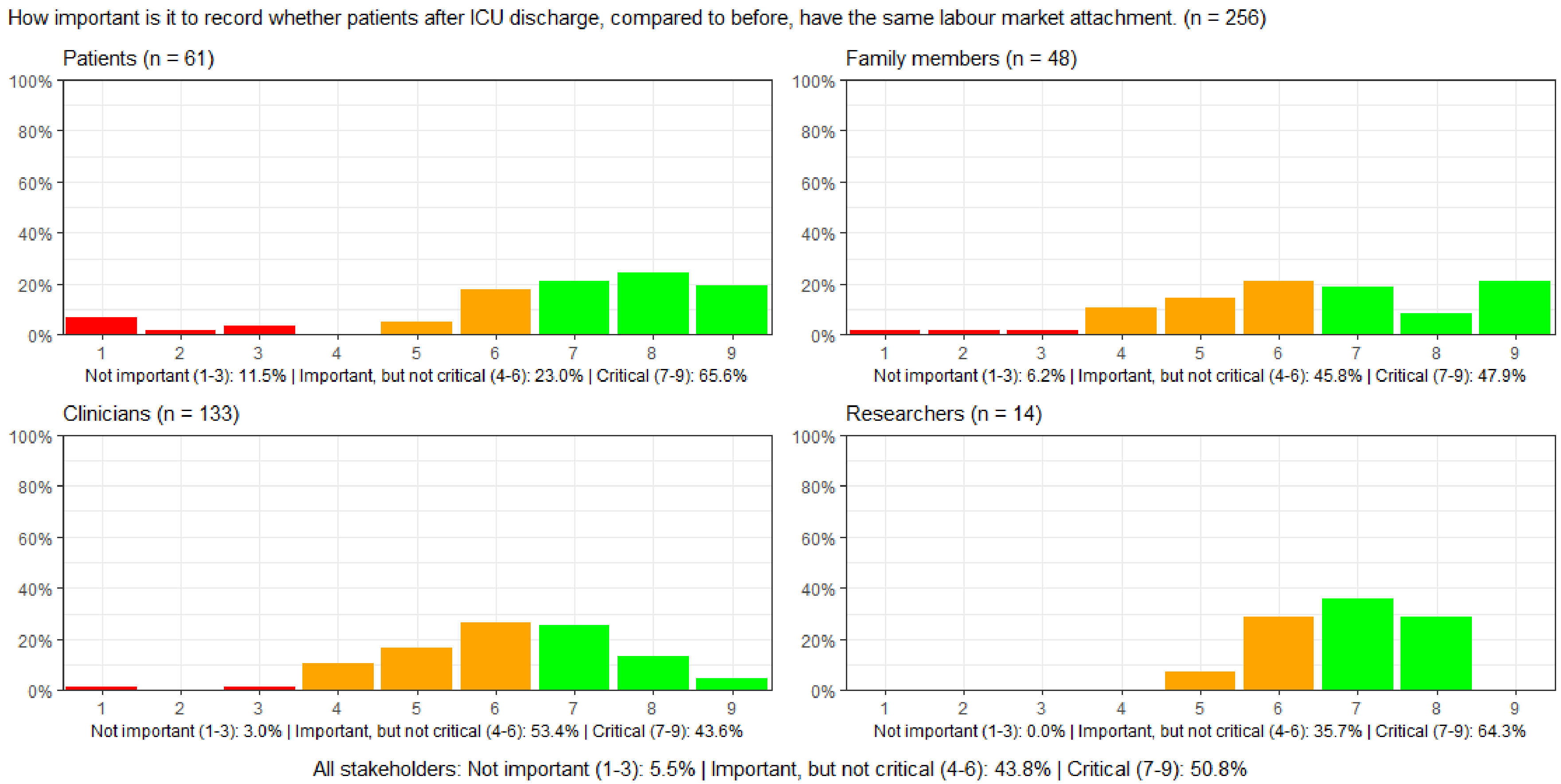

#### Figure S133: How important is it to record patients' physical impediments such as scars, wounds, or ostomy after ICU discharge?

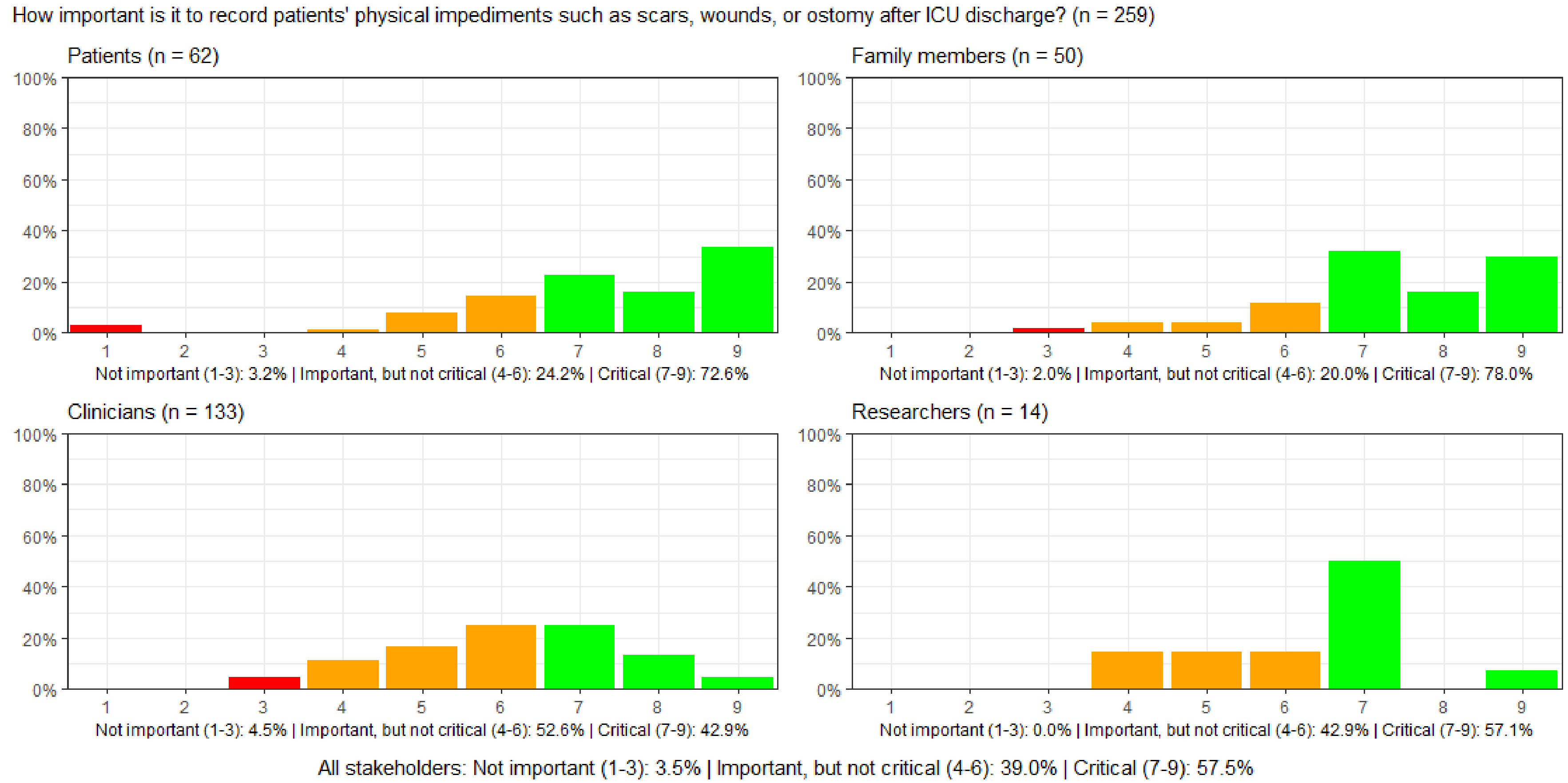

#### Figure S134: How important is it to record whether patients independently perform activities of daily living after ICU discharge?

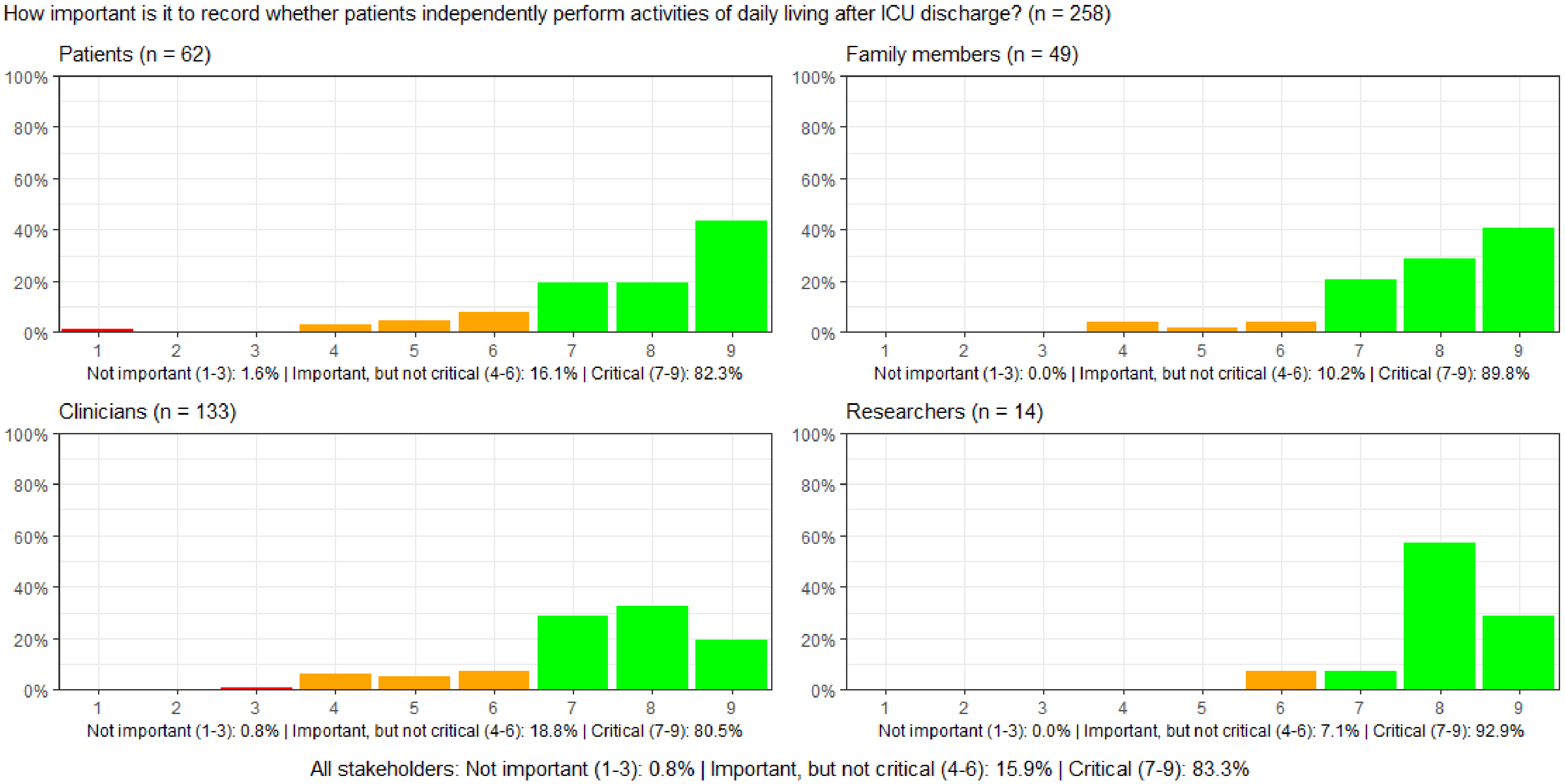

#### Figure S135: How important is it to record whether patients are independently able to get dressed after ICU discharge?

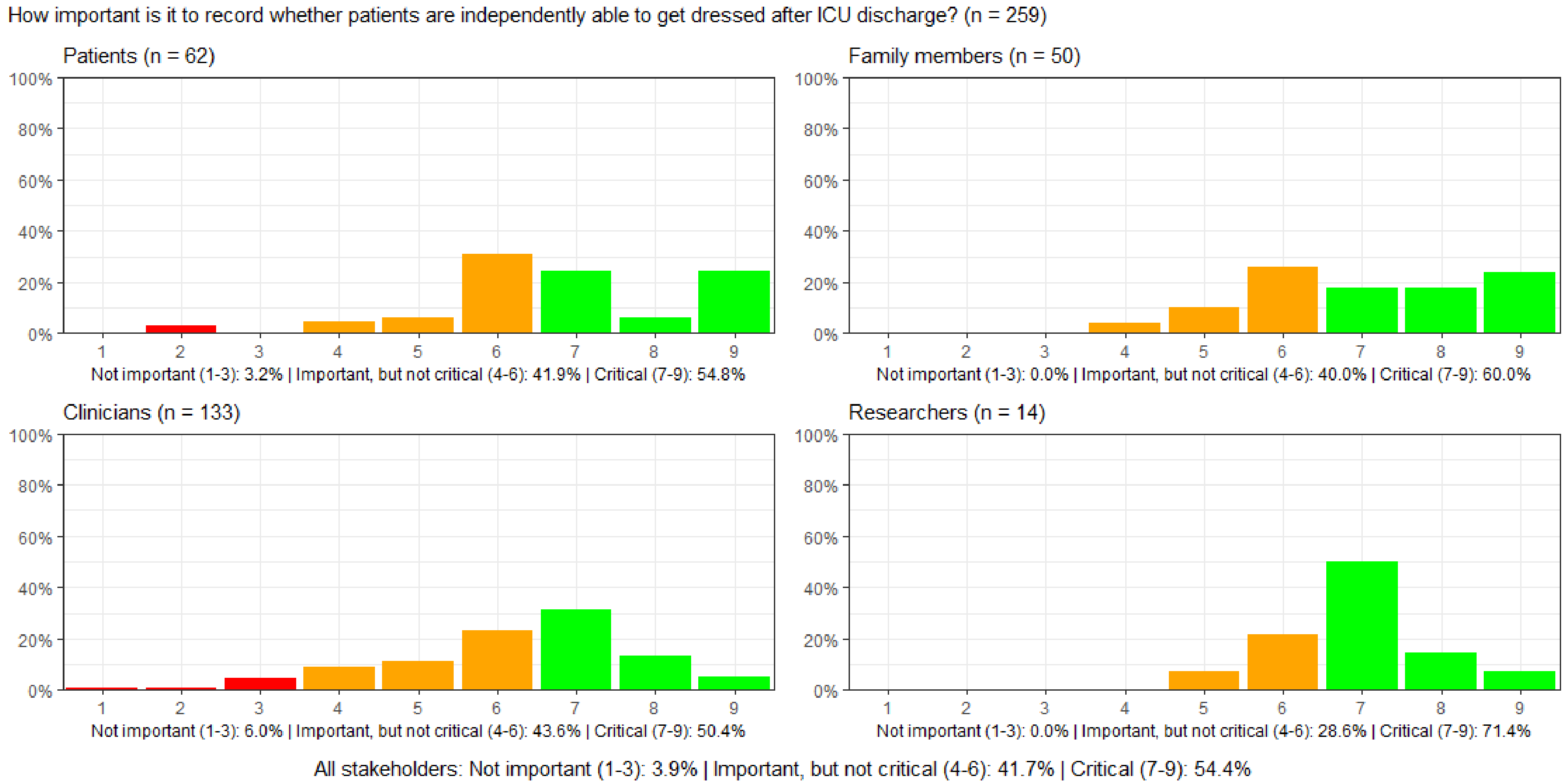

#### Figure S136: How important is it to record whether patients are mobile after ICU discharge?

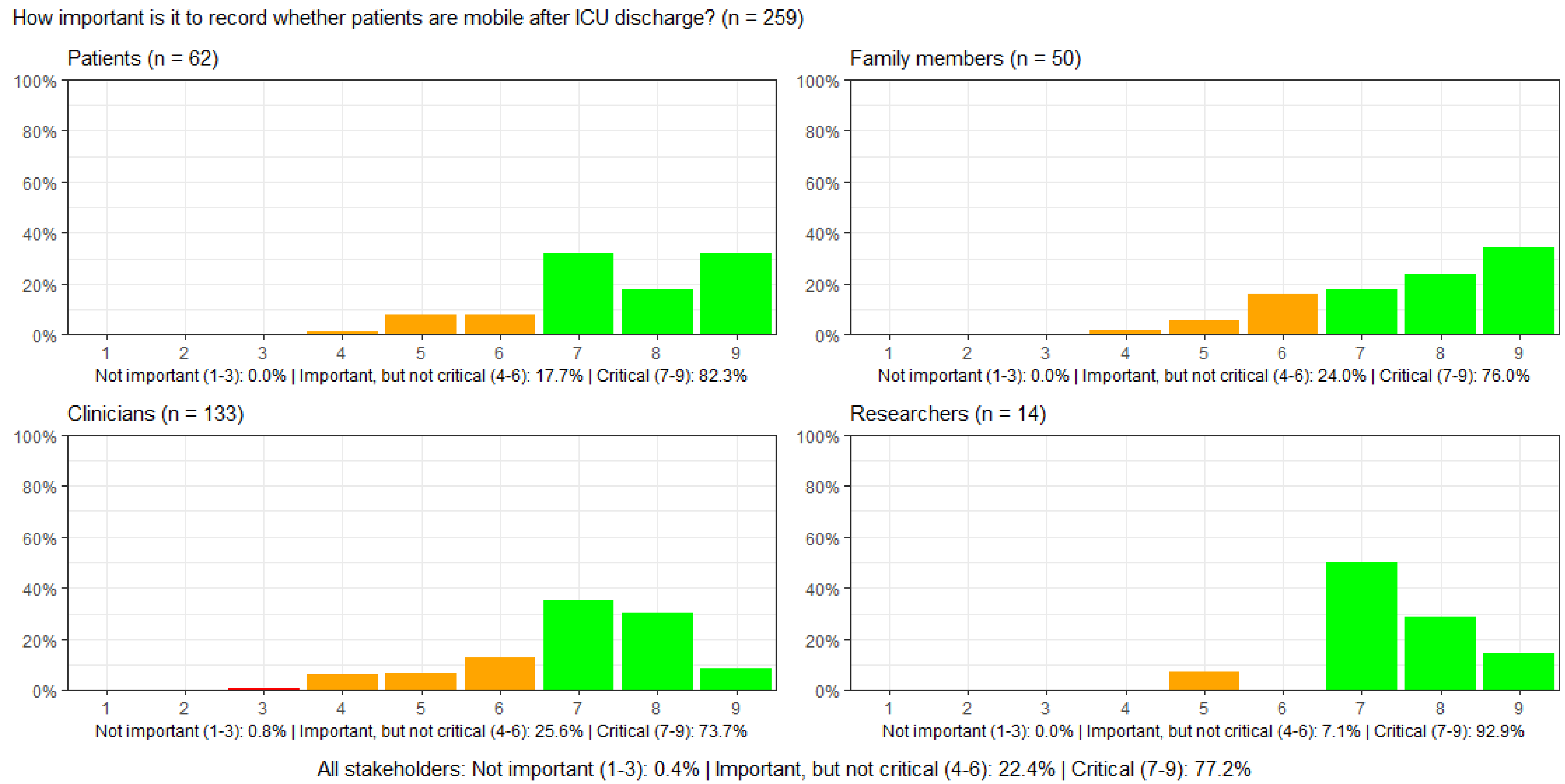

#### Figure S137: How important is it to record whether patients experience newly onset shortness of breath after ICU discharge?

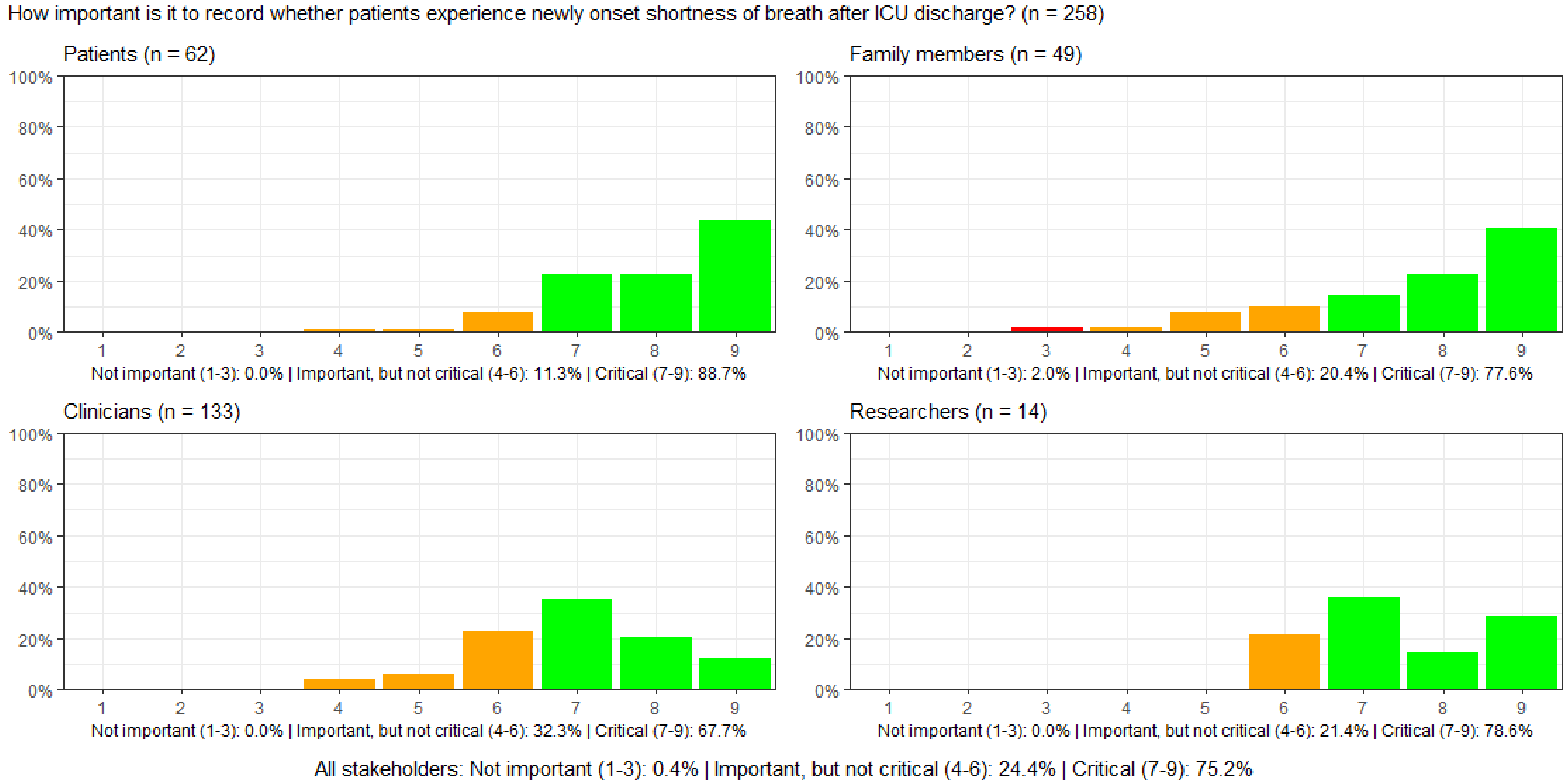

#### Figure S138: How important is it to record whether patients have newly onset trouble speaking after ICU discharge?

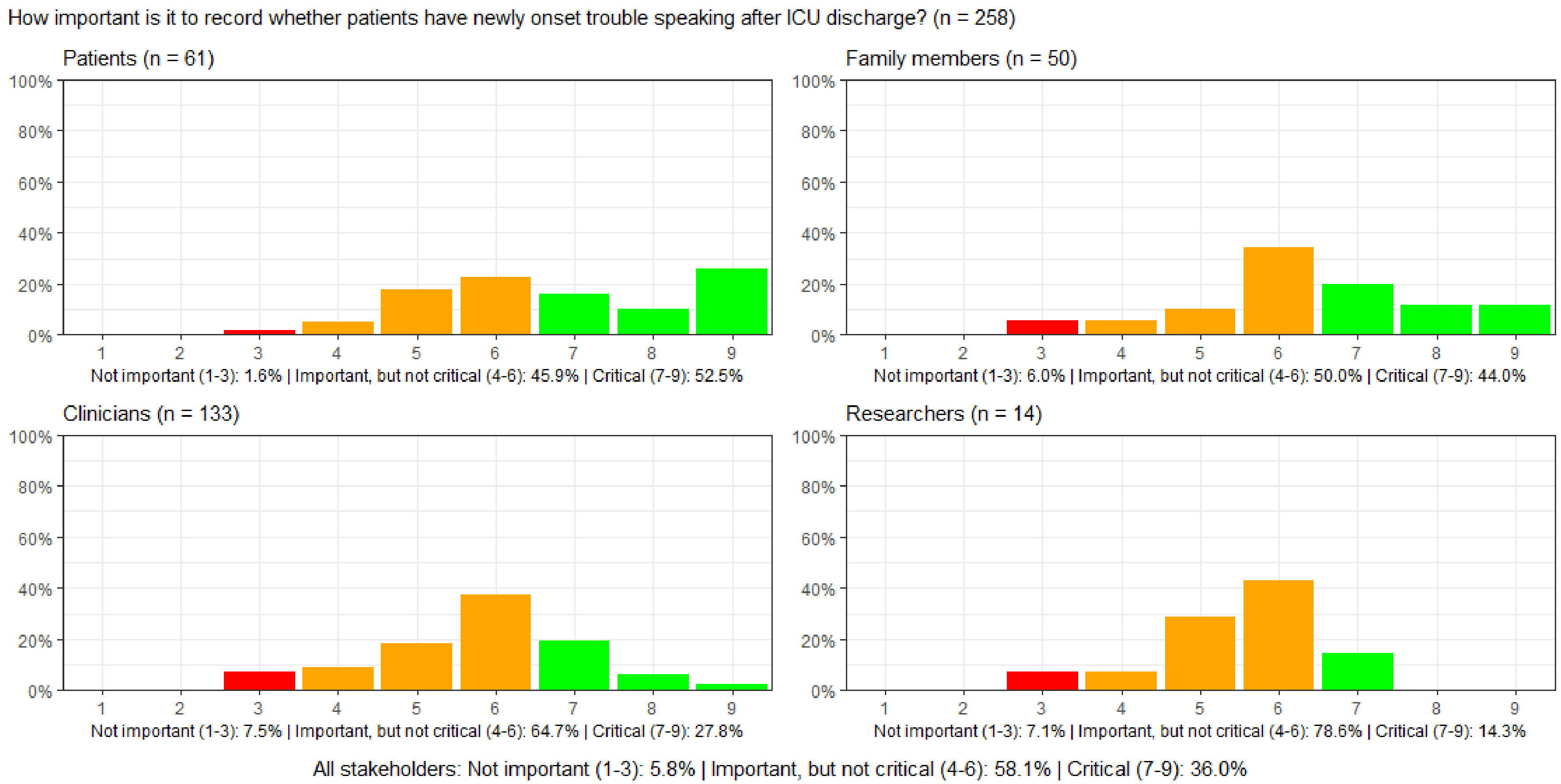

#### Figure S139: How important is it to record whether patients have newly onset difficulty swallowing after ICU discharge?

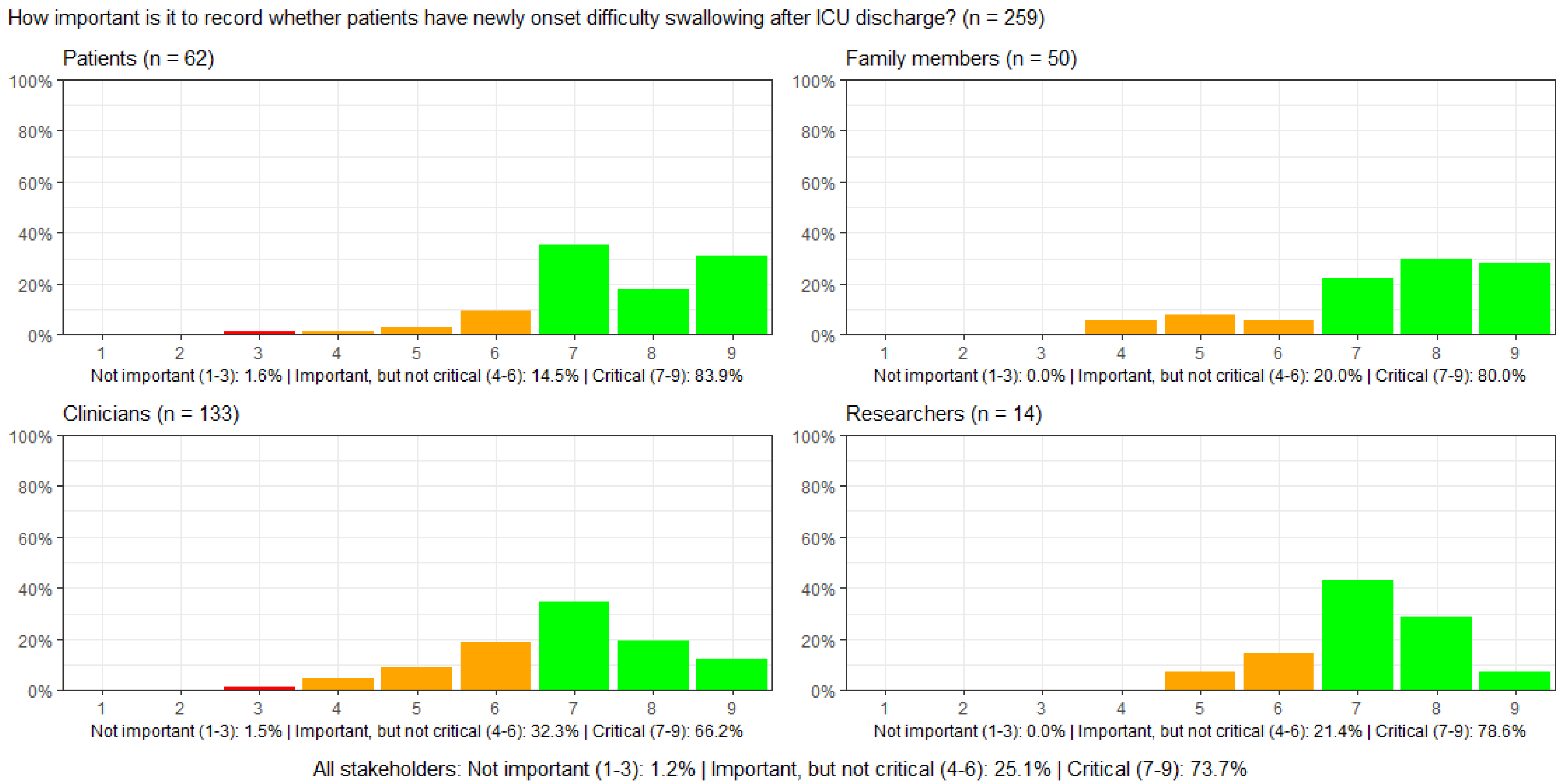

#### Figure S140: How important is it to record patients' bowel function after ICU discharge?

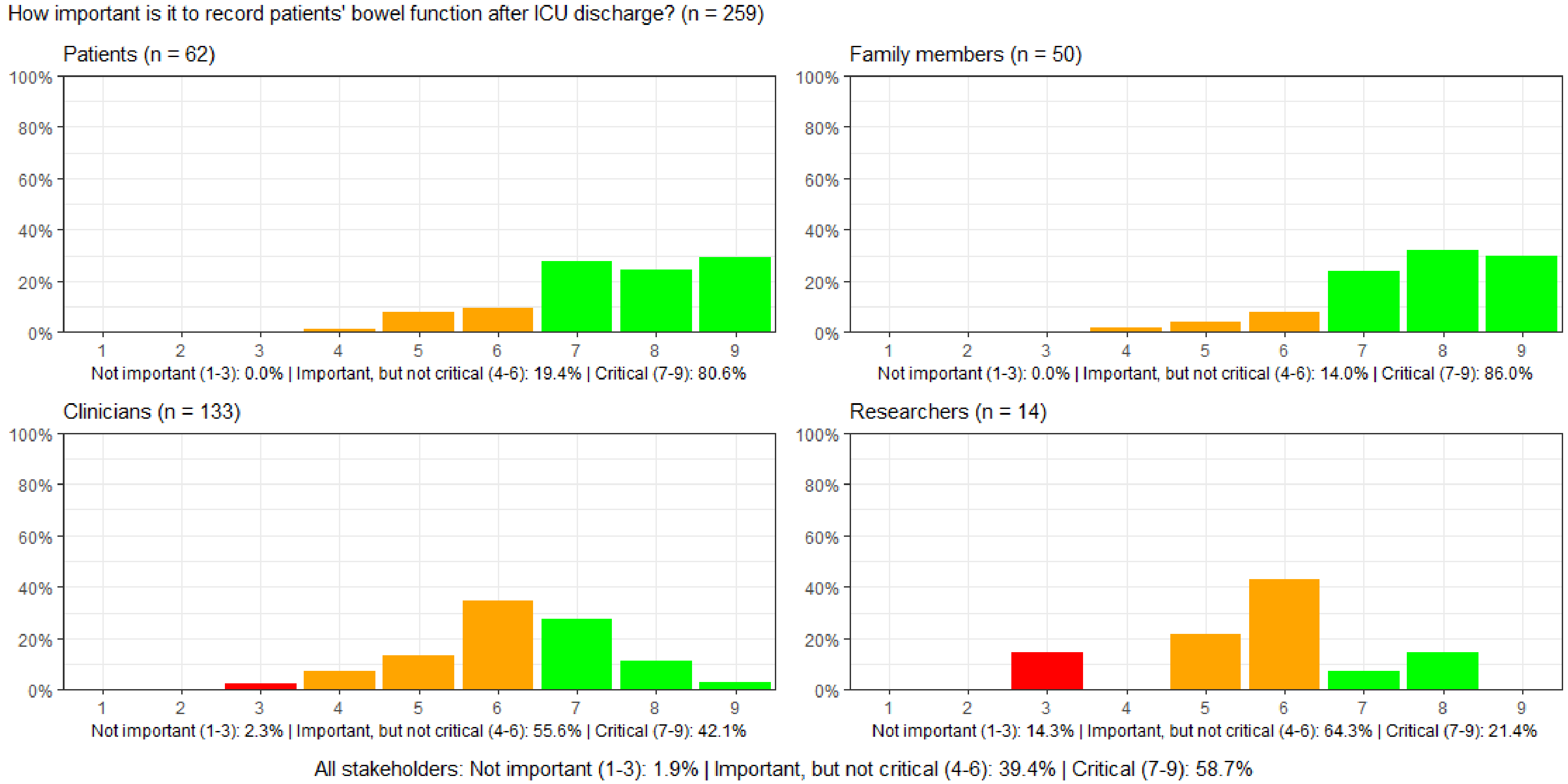

#### Figure S141: How important is it to record patients' urinary function after ICU discharge?

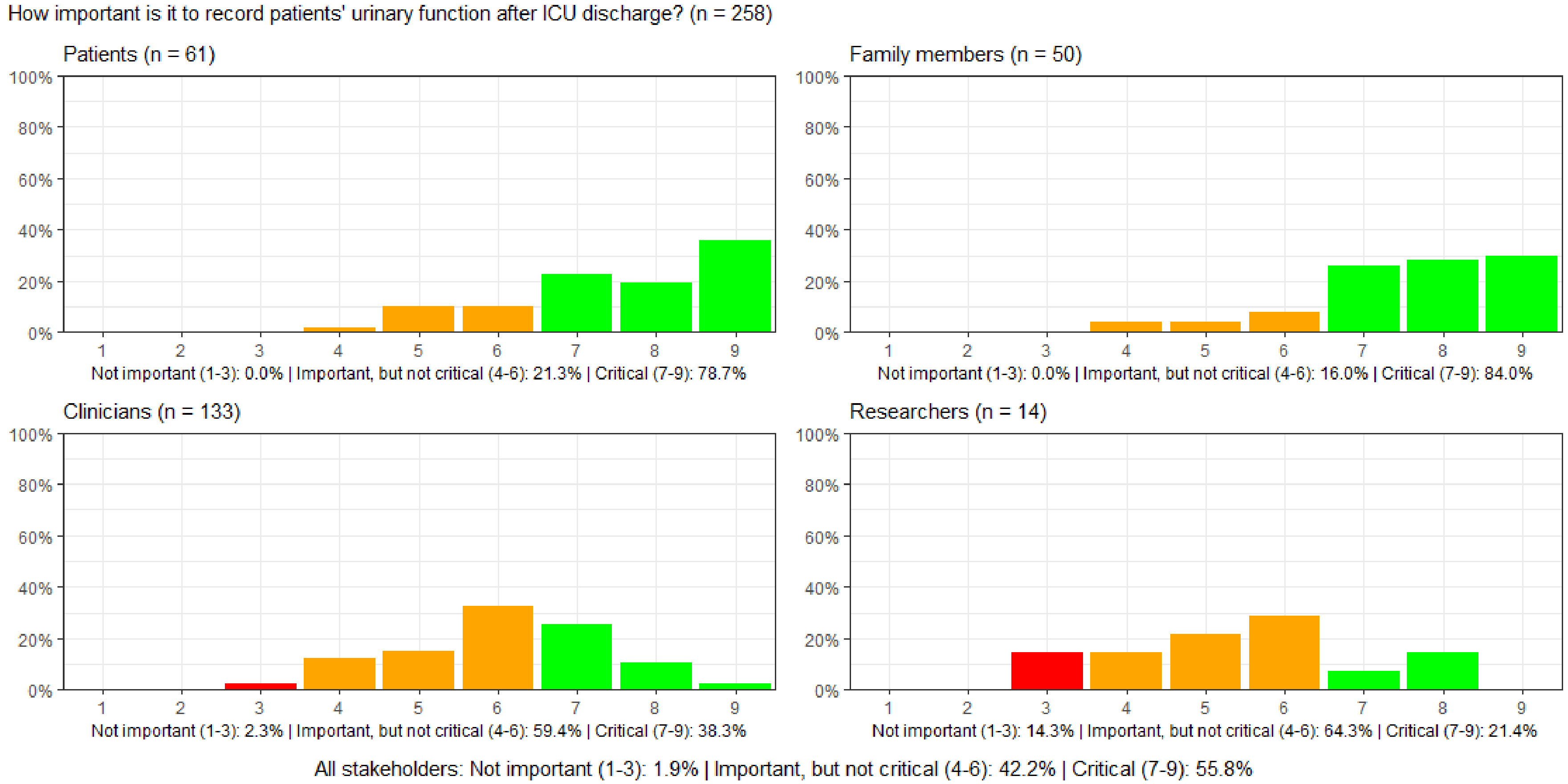

#### Figure S142: How important is it to record the patient's sexual function after ICU discharge?

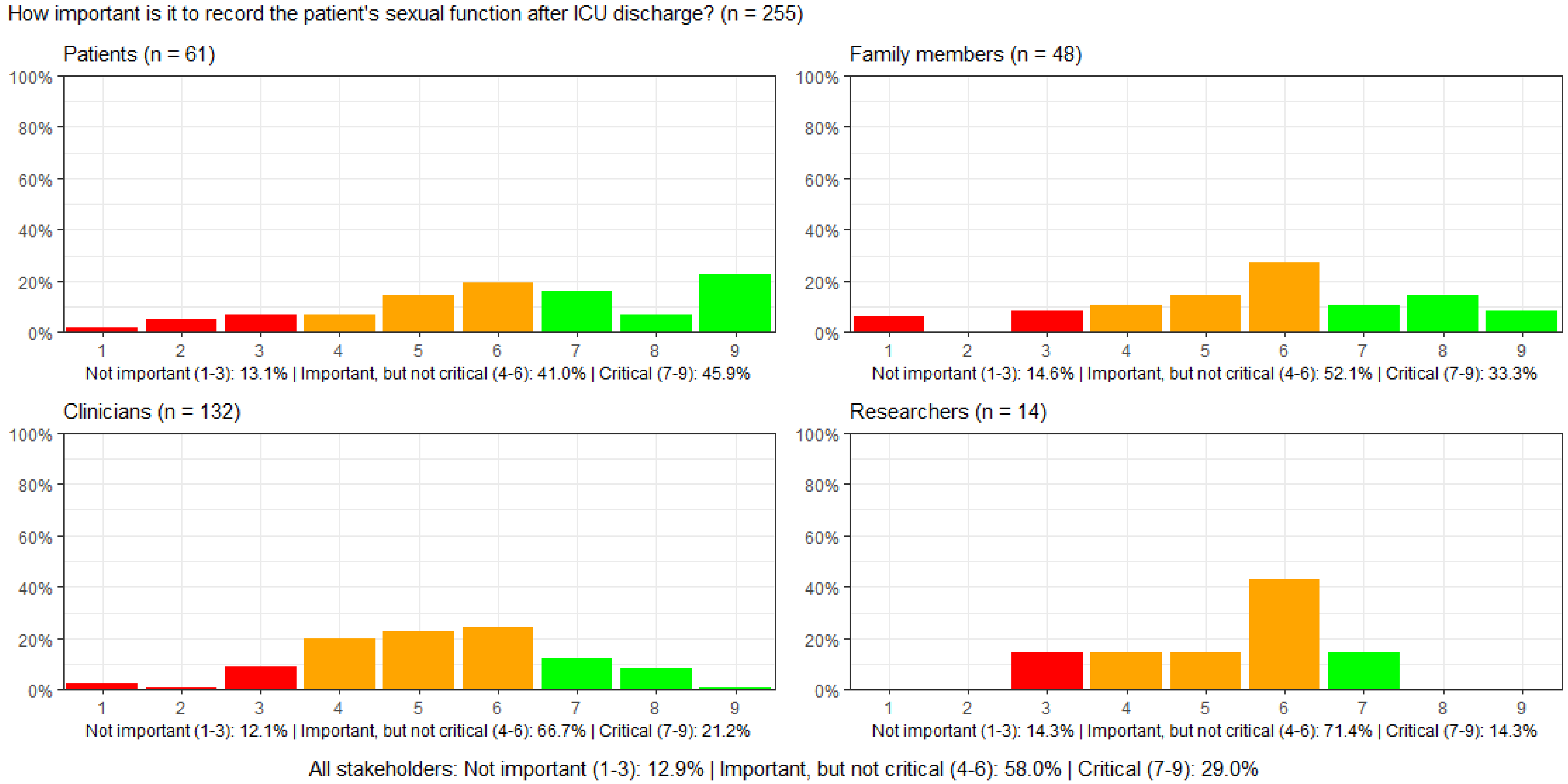

#### Figure S143: How important is it to record patients' senses: vision, hearing, touch, smell, and taste after ICU discharge?

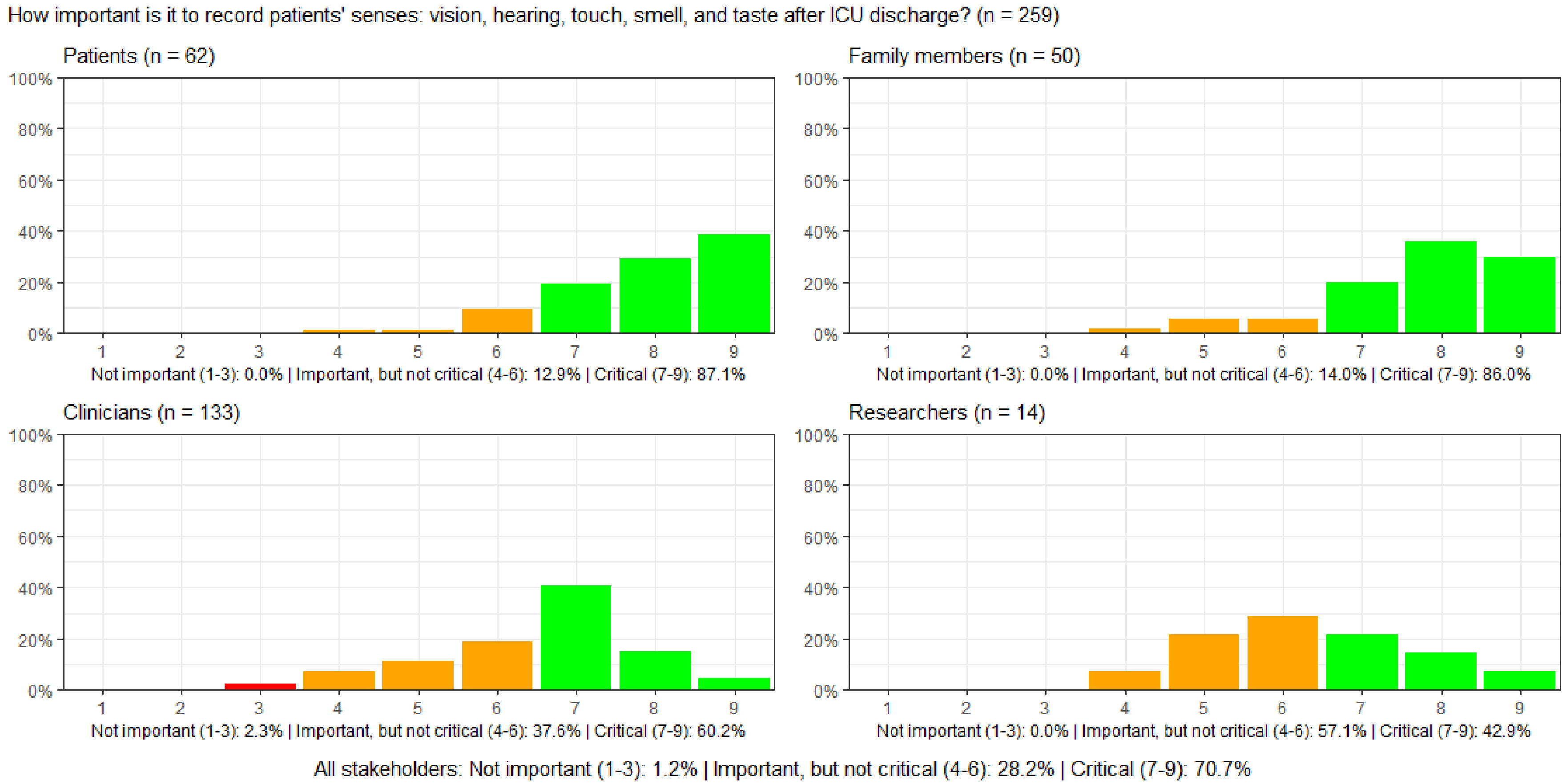

#### Figure S144: How important is it to record whether patients have the physical strength to perform activities of daily living after ICU discharge?

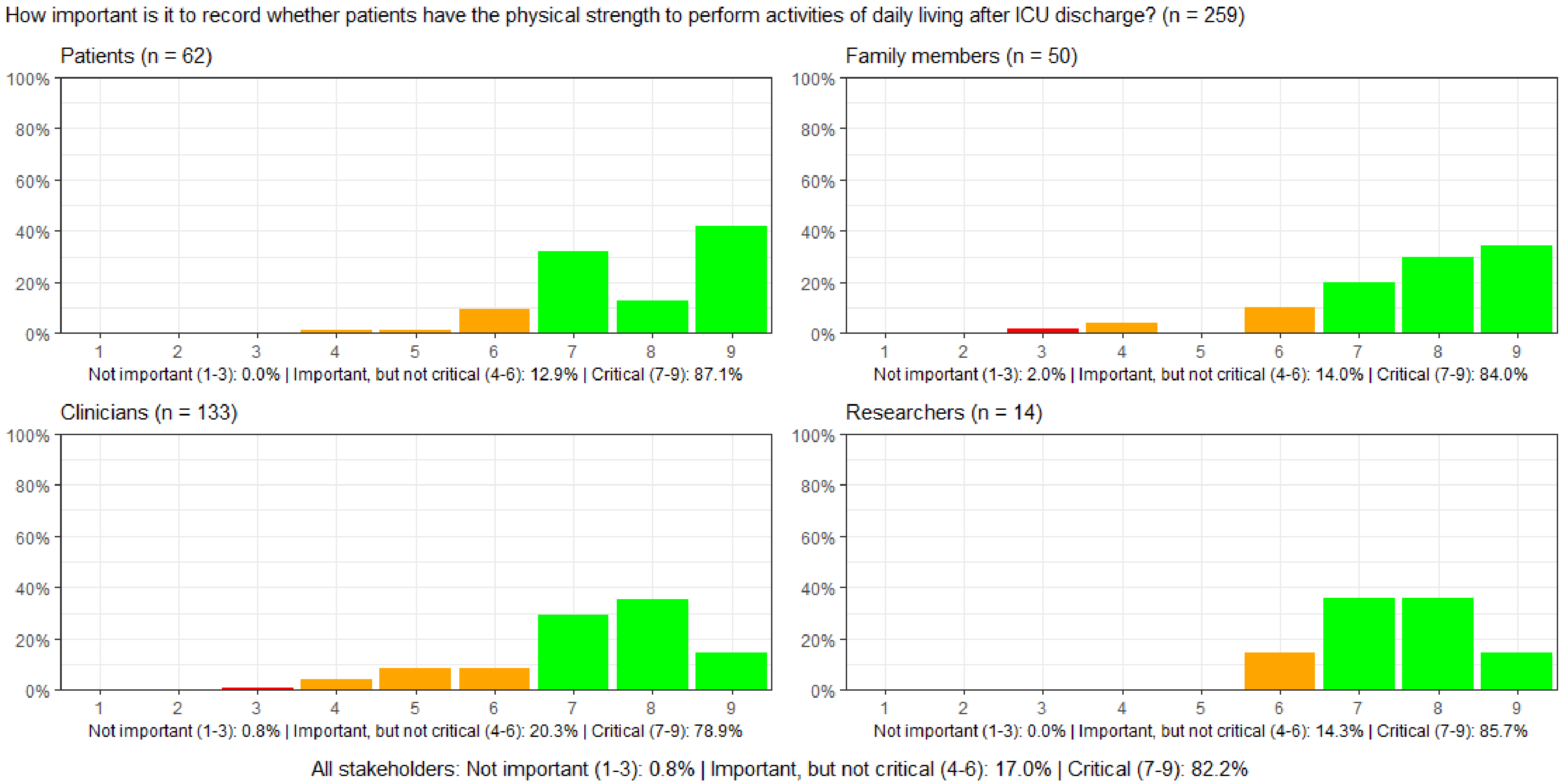

#### Figure S145: How important is it to record patients' mental function (cognitive function) after ICU discharge?

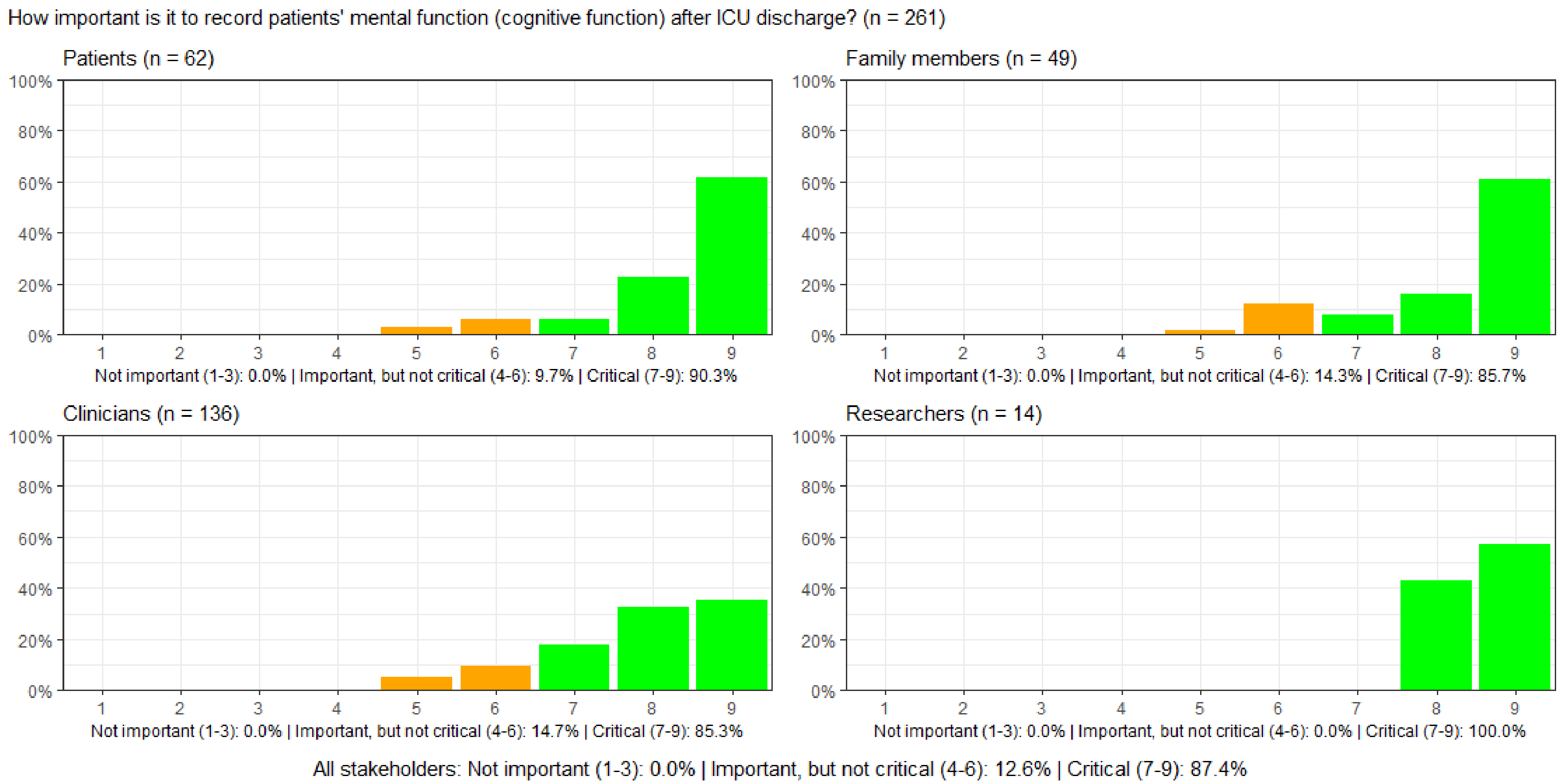

#### Figure S146: How important is it to record whether patients experience loneliness after ICU discharge?

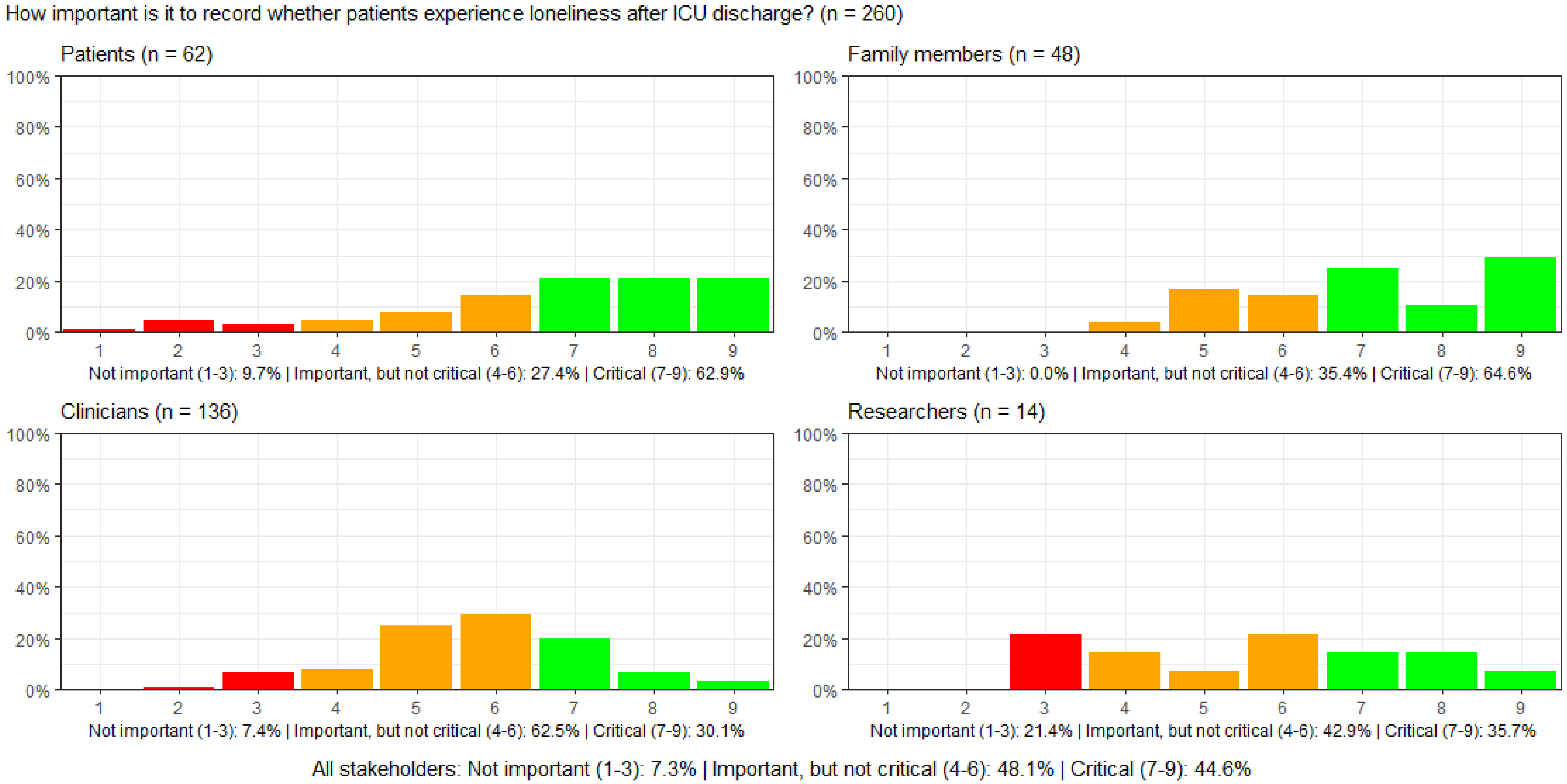

#### Figure S147: How important is it to record whether patients are diagnosed with posttraumatic stress syndrome (PTSD) after ICU discharge?

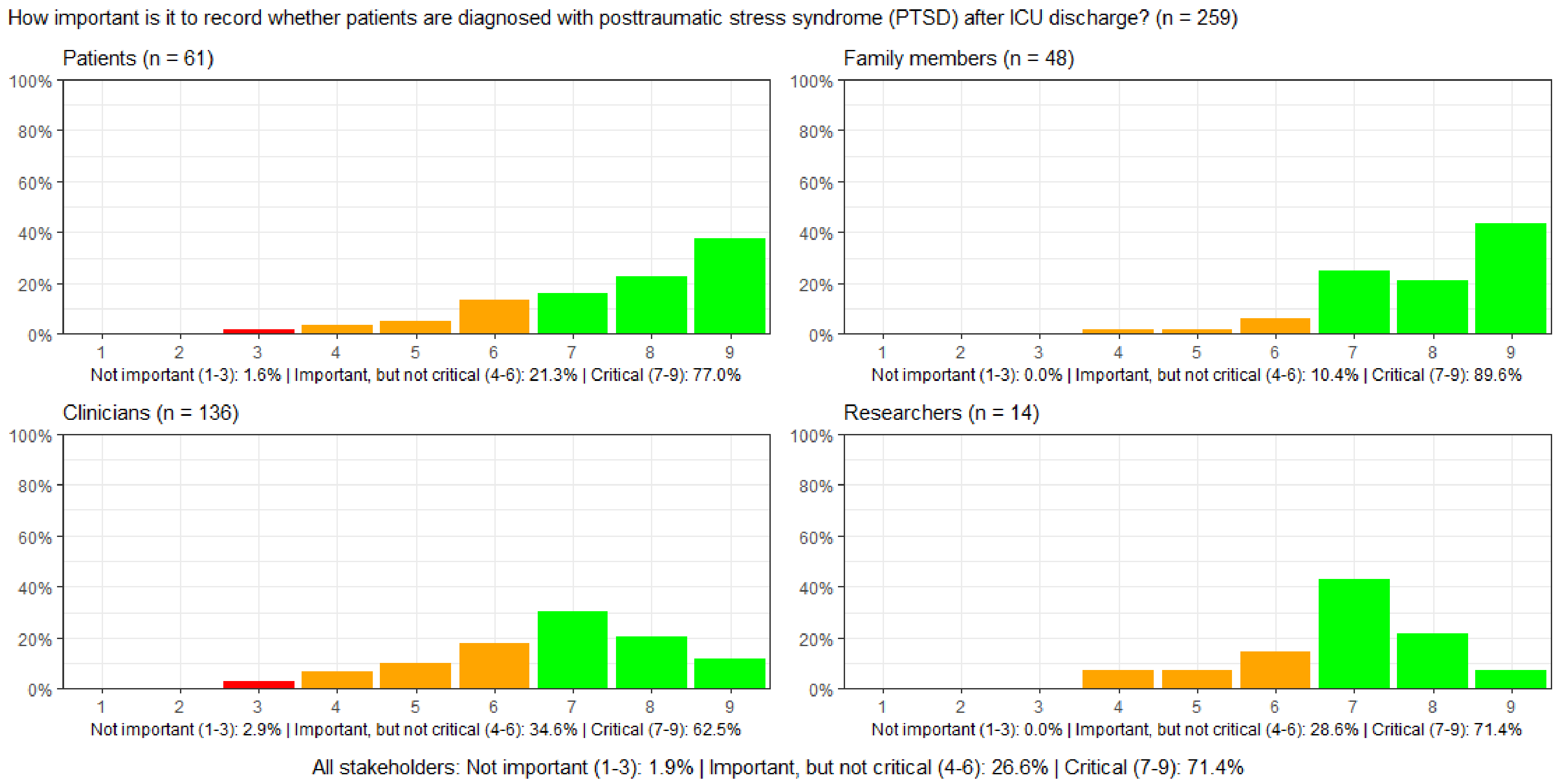

#### Figure S148: How important is it to record whether patients experience anxiety after ICU discharge?

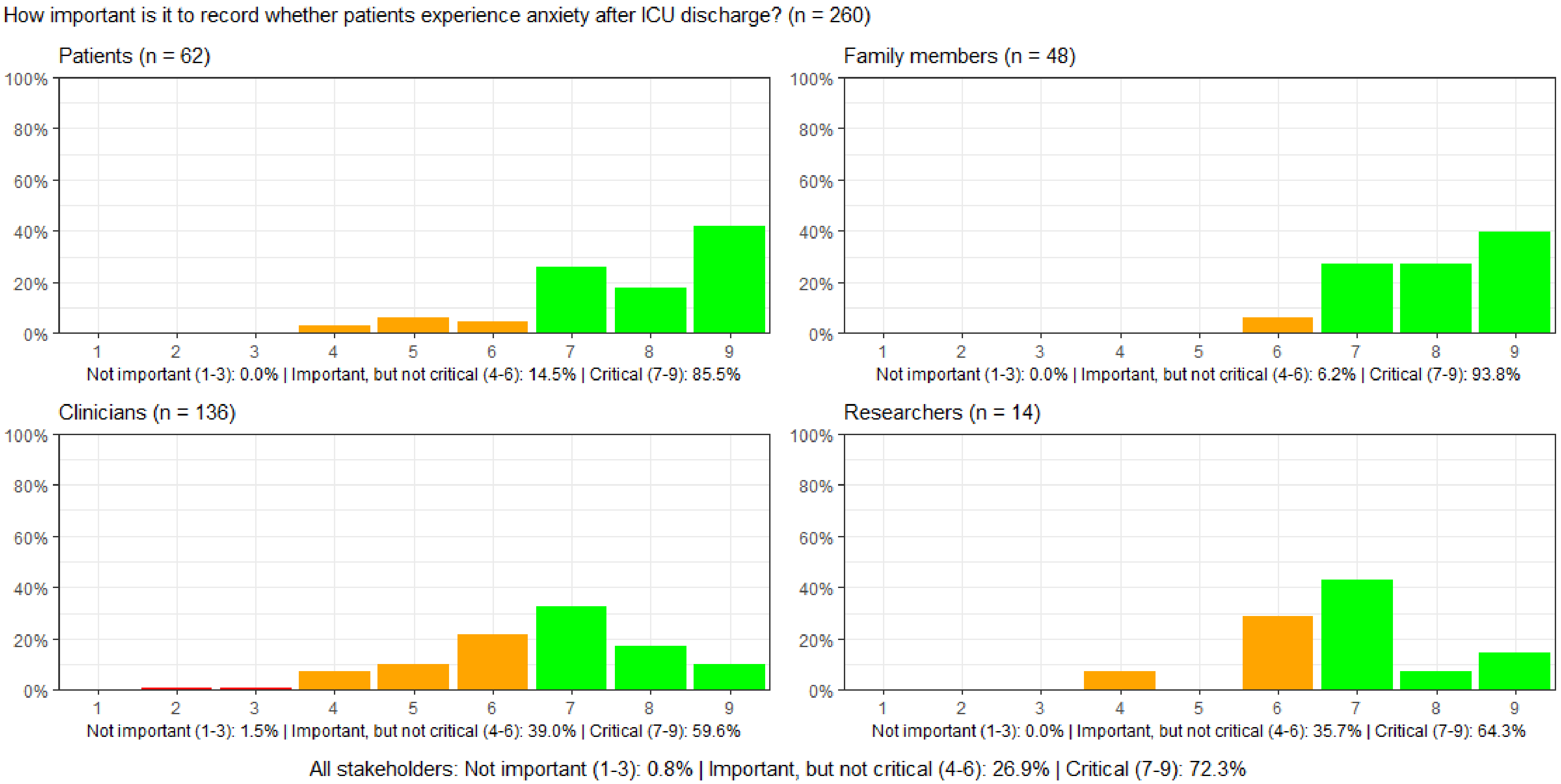

#### Figure S149: How important is it to record whether patients experience depression after ICU discharge?

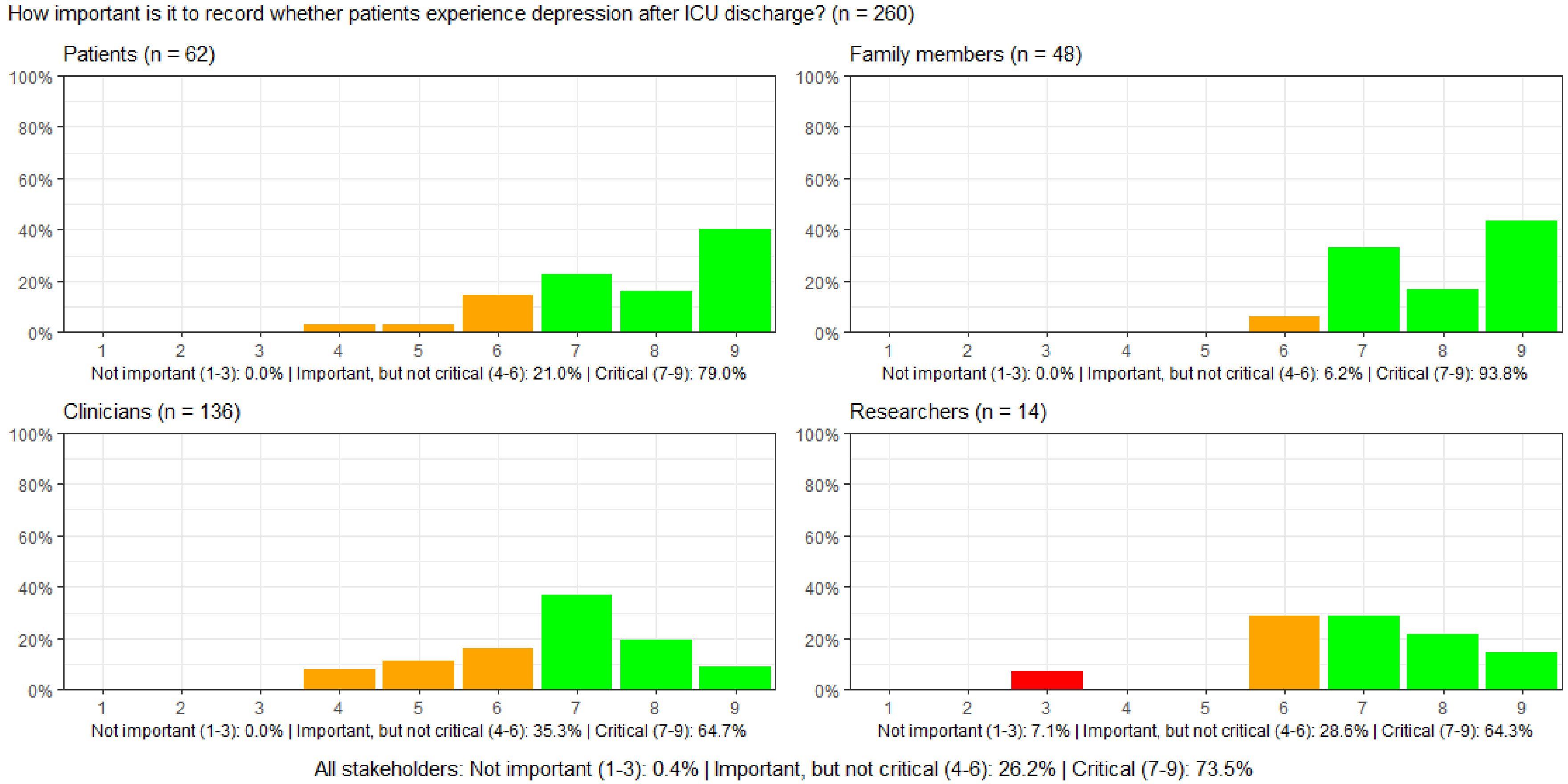

#### Figure S150: How important is it to record whether patients experience nightmares after ICU discharge?

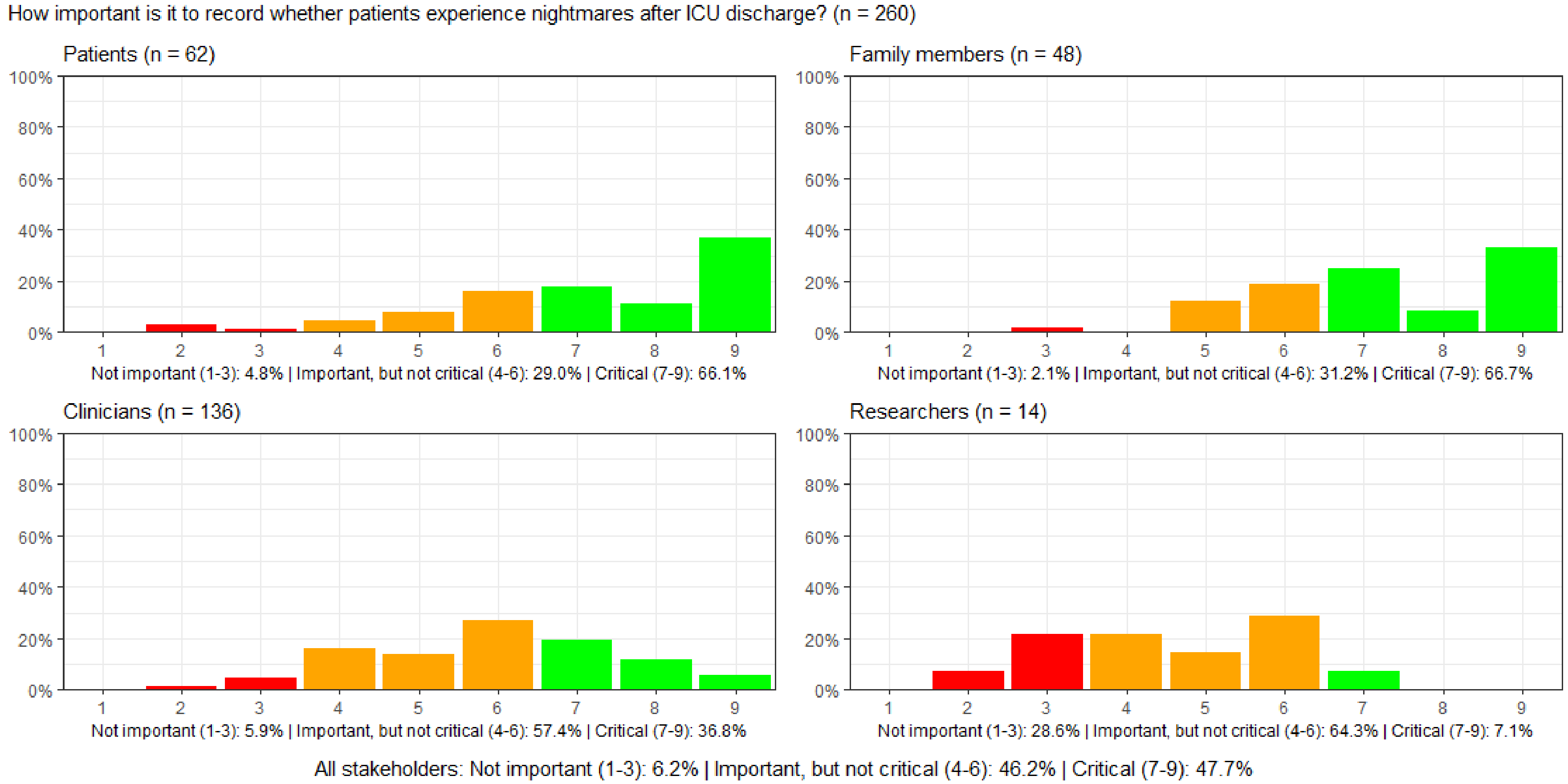

#### Figure S151. Heatmap showing the three Delphi survey round responses for all stakeholders

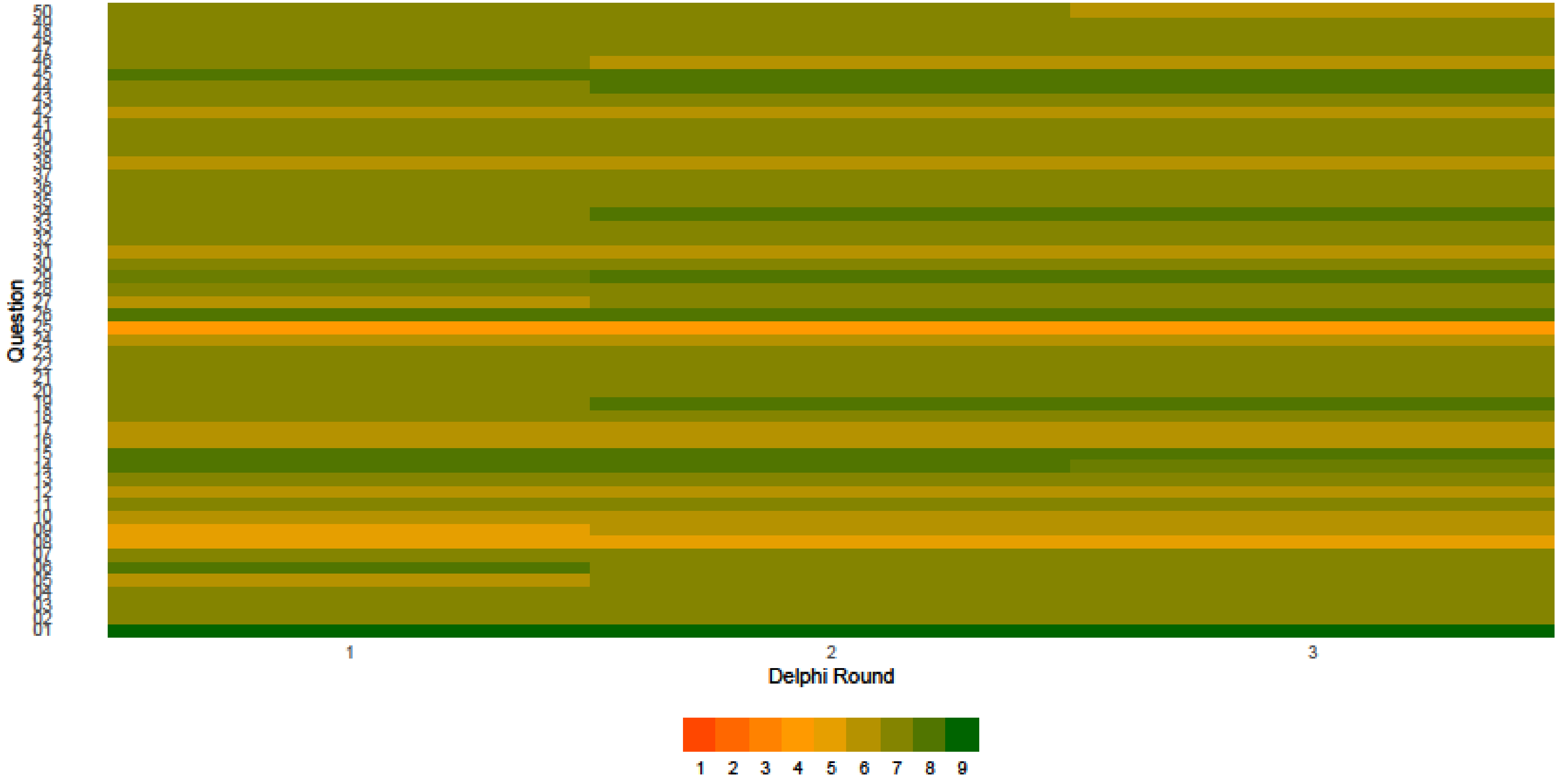


All participants rated the 50 outcomes ("Question") on a Likert scale from 1-9, where 1-3 is 'not important', 4-6 'important, but not critical', and 7-9 'critical' to include this outcome. This was rated for each survey round nr 1, 2, and 3. Greener represents more important and more orange less important.

#### Figure S152. Heatmap showing the three Delphi survey round responses by stakeholder groups

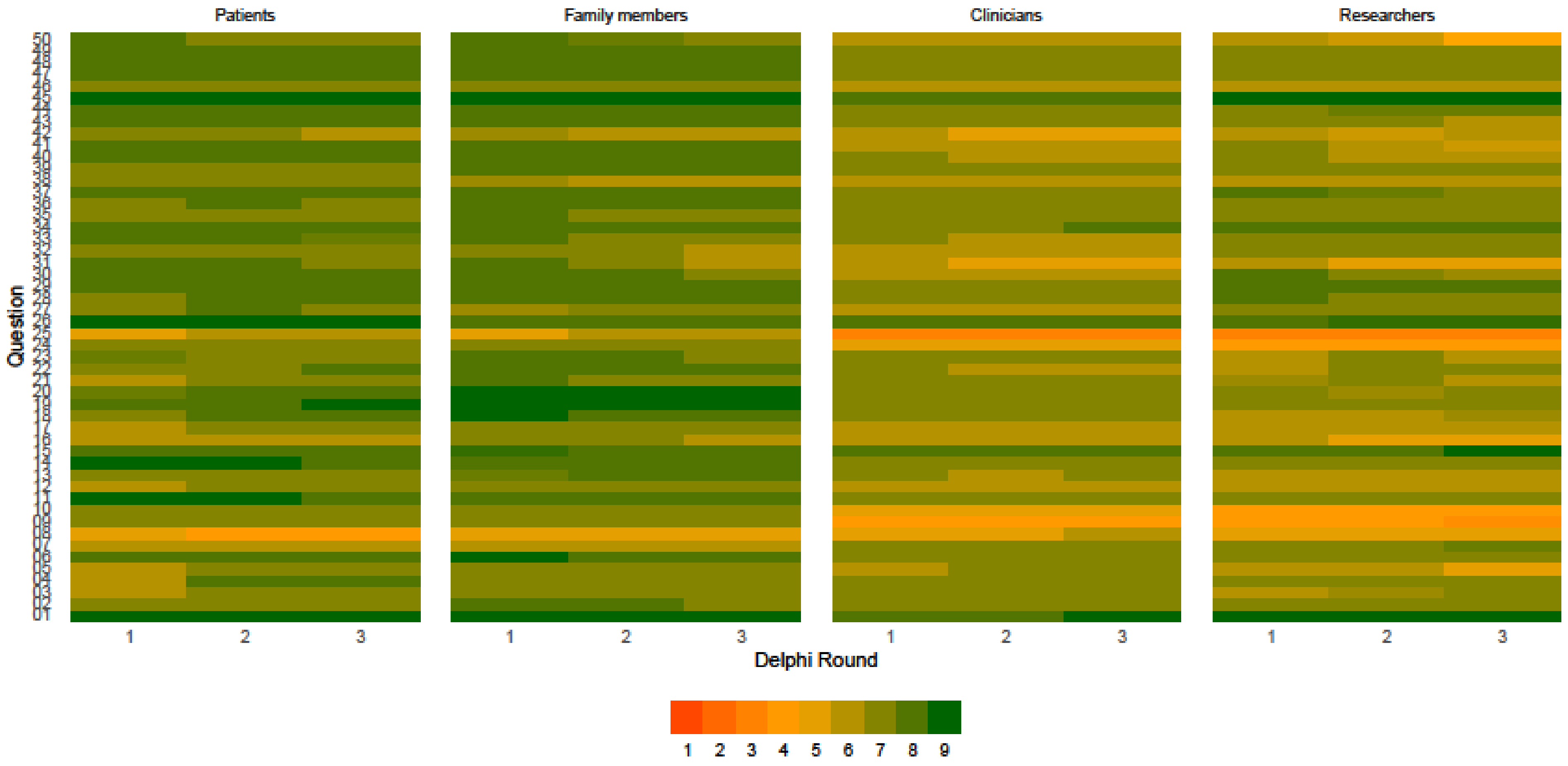


All participants rated the 50 outcomes ("Question") on a Likert scale from 1-9, where 1-3 is 'not important', 4-6 'important, but not critical', and 7-9 'critical' to include this outcome. This was rated for each survey round nr 1, 2, and 3. Greener represents more important and more orange less important.

#### Table S9. Outcomes related to health-related quality of life

| Outcomes | Number of panels (out of 4) who agreed on the importance of each outcome |
|---|---|
| Record patients' outpatient or general practitioner visits related to the long-term effects of intensive care | **2** |
| New onset tiredness or fatigue, unrelieved by sleep or rest, after ICU discharge | **4** |
| New onset pain after ICU discharge | **4** |
| Resume their usual roles in daily life after ICU discharge | **3** |
| Record whether patients after ICU discharge, compared to before, have the same labor market attachment | **2** |
| Independently perform activities of daily living after ICU discharge | **1** |
| Independently able to get dressed after ICU discharge | **1** |
| Mobile after ICU discharge | **1** |
| New onset speaking troubles after ICU discharge | **2** |
| New onset swallowing difficulties ‘after ICU discharge | **1** |
| Bowel function after ICU discharge | **3** |
| Urinary function after ICU discharge | **1** |
| Sexual function after ICU discharge | **1** |
| Senses: vision, hearing, touch, smell, and taste after ICU discharge | **3** |
| Physical strength to perform activities of daily living after ICU discharge | **1** |
| Loneliness after ICU discharge | **2** |
| Anxiety after ICU discharge | **2** |
| Depression after ICU discharge | **2** |

The colours refer to the number of panels who have agreed on the importance of each outcome. Dark green is 4/4 panels, light green 3/4 panels, yellow 2/4 panels, and orange 1/4 panel.

#### Table S10. Results from the international validation meetings across the participating countries

| Country | Initial outcomes[a] | | | | | | Additional outcomes[a] |
|---|---|---|---|---|---|---|---|
| | Survival | Days alive without life support | Days alive without coma or delirium | Days alive out of hospital | HRQoL | Cognitive function | |
| Australia[b] General ICU patient | Adopt | Adapt | Adapt | Adapt | Adopt | Reject | Physical function, ability to return occupation/school, Contribution to society, supporting network |
| Australia[b] Sepsis survivors | Adopt | Adopt | Adapt | Adapt | Adopt | Adapt | |
| Czech Republic | Adopt | Adapt | Adopt | Adopt | Adapt | Adopt | |
| Denmark (Copenhagen)[c] | Adopt | Adopt | Adopt | Adopt | Adopt | Adopt | Wellbeing |
| Denmark (Køge)[d] | Adopt | Adopt | Adopt | Adopt | Adopt | Adopt | Wellbeing |
| Denmark (Kolding)[e] | Adopt | Adopt | Adopt | Adopt | Adopt | Adopt | Wellbeing |
| Denmark (Aalborg)[f] | Adopt | Adopt | Adopt | Adopt | Adopt | Adopt | Wellbeing |
| Finland (Helsinki) | Adopt | Adopt | Adopt | Adopt | Adopt | Adopt | No additional outcomes |
| Finland (Tampere) | Adopt | Adopt | Reject | Adopt | Adopt | Adopt | Wellbeing should be added. |
| Iceland | Adopt | Adopt | Adopt | Adopt | Adopt | Adopt | Readmission |
| India (Chennai)[e] | Adopt | Adopt | Adopt | Adopt | Adopt | Adopt | Financial consequences |
| India (Mumbai) | Adopt | Adopt | Adopt | Reject | Adopt | Adopt | |
| Italy | Adopt | Adopt | Adopt | Adopt | Adopt | Adopt | Wellbeing of patients and families |
| Lithuania | Adopt | Adopt | Adapt | Adopt | Adopt | Adapt | Impact of intervention, |
| Norway | Adopt | Adopt | Reject | Reject | Adopt | Adopt | Physical function and mental health |
| Netherlands | Adopt | Adopt | Adopt | Adopt | Adopt | Adopt | No additional outcomes |
| Poland | Adopt | Adopt | Adopt | Adopt | Adapt | Adapt | No additional outcomes |
| Sweden | Adopt | Adopt | Adopt | Adopt | Adopt | Adopt | Physical function and psychological function |
| Switzerland | Adopt | Adopt | Adopt | Adapt | Adopt | Adopt | Mobility, Independency (self-efficacy in ADL), family outcomes |
| United Kingdom (Cardiff) | Adopt | Adopt | Adapt | Adopt | Adopt | Adopt | No additional outcomes |
| United Kingdom (London) | Adopt | Adopt | Adopt | Adopt | Adopt | Adopt | Mental well-being |
| Total adopted | 21 (100%) | 19 (90%) | 15 (71%) | 16 (76%) | 19 (90%) | 17 (81%) | |
| Total adapted | 0 (0%) | 2 (10%) | 4 (19%) | 3 (14%) | 2 (10%) | 3 (14%) | |
| Total rejected | 0 (0%) | 0 (0%) | 2 (10%) | 2 (10%) | 0 (0%) | 1 (5%) | |

HRQoL: Health-related quality of life, ADL: activities of daily living

Green/Adopt=adopting the outcome, yellow/Adapt=adapting the outcome, and red/Reject=rejecting the outcome.

Number (percentages).

[a] Detailed minutes in Table S10.

[b] Number of participants from different areas of Australia: 1 Aboriginal participants, 11 Western Australia, 5 New South Wales, 4 Victoria, 3 Queensland, and 1 Northern Territory.

[c] The Capitol Region of Denmark

[d] Region Zealand

[e] The Region of Southern Denmark

[f] The North Denmark Region

[e] Indian areas represented: Chennai, Bangalore, Vellore, Pune, and New Delhi

#### Comments to the adjusted core outcomes

**Australia:** full summary in the end of this section.

**Czech Republic:** would like to add ‘non-vegetative state’ to survival as the panel considered that worse than death (may not reflect everyone feelings in the Czech Republic). ‘Free of life support’ was not considered important for patients if comfort, communication, and no dyspnoea feelings were present, why they want to adapt the outcome to ‘free of life support and not dyspnoeic’. With HRQoL the physical component is not as important to patients and families if they can accept, adapt to a condition, that make sense of it or hope for improvement.

**Lithuania:** changed two outcomes from adapt to adopt after the core outcome set has been revised. Regarding ‘free of delirium’ the main challenge with the previous outcome was the definition of coma. No major concerns raised regarding identification of delirium. It is a different outcome from one intended in the initial core outcome set, but it is a very important outcome. Regarding ‘cognitive function’ they were unanimous to its importance, but concerns raised regarding the methodology and tools used for this outcome.

**Netherlands:** after discussing the changes in the core outcome set their research panel agrees with the new set, which covers the most important outcomes.

**Norway:** the majority are against ‘free of delirium’ due to it is impractical and that it is unfortunate omitting specific and separate domains for physical and mental outcomes.

**Sweden:** the only outcome we are hesitant about is ‘out of hospital’. It is financially and clinically important, but perhaps not so important from a patient perspective. However, this might perhaps be addressed in the next step of development

**Switzerland:** finds ‘out of hospital’ okay, however they favour what have been suggested at the consensus meeting; ‘returning to pre-ICU condition’.

**Australian full summary ‘20240118’** for the general ICU patients and sepsis survivors:

##### A. Initial outcomes of the two Australian consumer group meetings

| | Initial core outcomes | | | | | | Additional outcomes |
|---|---|---|---|---|---|---|---|
| | Survival | Days alive without life support | Days alive without coma or delirium | Days alive and out of hospital | Health-Related Quality of Life (HRQoL) | Cognitive function | |
| General ICU patient/Fiona Stanley | Ok | Adapt | Adapt | Adapt | Ok | Reject | Physical function, ability to return occupation/school, Contribution to society, supporting network |
| Sepsis survivors/TGI | Ok | Ok | Adapt | Adapt | Ok | Adapt | |

##### B. Changes proposed by Danish Group based on international feedback

1. SURVIVAL: No change.
2. DAYS ALIVE WITHOUT LIFE SUPPORT: Changed to FREE OF LIFE SUPPORT. Specifics of how to define to be determined.
3. DAYS ALIVE WITHOUT COMA OR DELIRIUM. Changed to FREE OF DELIRIUM. Removal of coma.
4. DAYS ALIVE AND OUT OF HOSPITAL: Changed to OUT OF HOSPITAL.
5. HRQOL: No change.
6. Cognitive function: No change.
7. Additional outcomes: No change (none added).

#### C. <u>Proposed final Australian feedback</u>

| | **Proposed final core outcomes** | | | | | | **Nil Additional outcomes** |
|---|---|---|---|---|---|---|---|
| | **Survival** | **Free of life support** | **Free of delirium** | **Out of hospital** | **HRQoL** | **Cognitive function** | |
| **General ICU patient/Fiona Stanley** | **Ok** | **OK** | **OK** | **Adapt** | **Ok** | **Adapt** | **Adapt** |
| **Sepsis survivors/TGI** | **Ok** | **Ok** | **OK** | **Adapt** | **Ok** | **Adapt** | |

1. SURVIVAL, FREE OF LIFE SUPPORT, FREE OF DELIRIUM and HEALTH RELATED QUALITY OF LIFE: Accept.

2. OUT OF HOSPITAL: <u>Suggest adapt</u> core outcome to include a caveat about return to home and/or previous level of function.

3. COGNITIVE FUNCTION: <u>Suggest adapt</u> core outcome. Add caveat that the impact of cognition on function should be considered in any measure of HEALTH RELATED QUALITY OF LIFE and that cognitive function should not measured as a stand alone core outcome.

4. ADDITIONAL OUTCOMES: <u>Suggest add</u> to Australian core outcomes that physical, cognitive, psychological and social function should be considered in any measure of HEALTH RELATED QUALITY OF LIFE

#### Table S11. Core Outcome Set-STandards for Reporting: The COS-STAR Statement Checklist

| SECTION/TOPIC | ITEM No. | CHECKLIST ITEM | REPORTED ON PAGE NUMBER |
|---|---|---|---|
| TITLE/ABSTRACT | | | |
| Title | 1a | Identify in the title that the paper reports the development of a COS | 1 |
| Abstract | 1b | Provide a structured summary | 1 |
| INTRODUCTION | | | |
| Background and Objectives | 2a | Describe the background and explain the rationale for developing the COS. | 4 |
| | 2b | Describe the specific objectives with reference to developing a COS. | 4 |
| Scope | 3a | Describe the health condition(s) and population(s) covered by the COS. | 4 |
| | 3b | Describe the intervention(s) covered by the COS. | 4 |
| | 3c | Describe the setting(s) in which the COS is to be applied. | 4 |
| METHODS | | | |
| Protocol/Registry Entry | 4 | Indicate where the COS development protocol can be accessed, if available, and/or the study registration details. | 5 |
| Participants | 5 | Describe the rationale for stakeholder groups involved in the COS development process, eligibility criteria for participants from each group, and a description of how the individuals involved were identified. | 5-6 and ESM |
| Information Sources | 6a | Describe the information sources used to identify an initial list of outcomes. | 6 and ESM |
| | 6b | Describe how outcomes were dropped/combined, with reasons (if applicable). | 6 and ESM |
| Consensus Process | 7 | Describe how the consensus process was undertaken. | 6-7 |
| Outcome Scoring | 8 | Describe how outcomes were scored and how scores were summarised. | 6 and ESM |
| Consensus Definition | 9a | Describe the consensus definition. | 6 |
| | 9b | Describe the procedure for determining how outcomes were included or excluded from consideration during the consensus process. | 6 and ESM |
| Ethics and Consent | 10 | Provide a statement regarding the ethics and consent issues for the study. | 7-8 and ESM |
| RESULTS | | | |
| Protocol Deviations | 11 | Describe any changes from the protocol (if applicable), with reasons, and describe what impact these changes have on the results. | 9 |

| | | | |
|---|---|---|---|
| Participants | 12 | Present data on the number and relevant characteristics of the people involved at all stages of COS development. | 9 and Table 1 |
| Outcomes | 13a | List all outcomes considered at the start of the consensus process. | Figure 1 and ESM |
| | 13b | Describe any new outcomes introduced and any outcomes dropped, with reasons, during the consensus process. | 9 |
| COS | 14 | List the outcomes in the final COS. | 10-11 and Figure 3 |
| DISCUSSION | | | |
| Limitations | 15 | Discuss any limitations in the COS development process. | 12-13 |
| Conclusions | 16 | Provide an interpretation of the final COS in the context of other evidence, and implications for future research. | 13 |
| OTHER INFORMATION | | | |
| Funding | 17 | Describe sources of funding/role of funders. | 16 |
| Conflicts of Interest | 18 | Describe any conflicts of interest within the study team and how these were managed. | 16 |

COS: core outcome set

*From: Kirkham JJ, Gorst S, Altman DG, Blazeby JM, Clarke M, Devane D, et al. (2016) Core Outcome Set–STAndards for Reporting: The COS-STAR Statement. PLoS Med 13(10): e1002148.* https://doi.org/10.1371/journal.pmed.1002148 [10].

#### Table S12 Guidance for Reporting Involvement of Patients and the Public short form: The GRIPP2-SF checklist

| Section and topic | Item | Reported on page No |
|---|---|---|
| 1: Aim | Report the aim of PPI in the study | 4 |
| 2: Methods | Provide a clear description of the methods used for PPI in the study | 5 and Figure 1 |
| 3: Study results | Outcomes—Report the results of PPI in the study, including both positive and negative outcomes | ESM 23-32, 33 |
| 4: Discussion and conclusions | Outcomes—Comment on the extent to which PPI influenced the study overall. Describe positive and negative effects | Figure 1 and ESM 232 |
| 5. Reflections/critical perspective | Comment critically on the study, reflecting on the things that went well and those that did not, so others can learn from this experience | ESM 232-233 |

*PPI* patient and public involvement

*From: Staniszewska S, Brett J, Simera I, et al (2017) GRIPP2 reporting checklists: tools to improve reporting of patient and public involvement in research. BMJ j3453.* https://doi.org/10.1136/bmj.j3453 [11].

**GRIPP2-SF Item 4; Discussion and conclusion**

Figure 1 provides an overview of all processes in which the research panels participated. Detailed explanations of these processes are available on page 3 in the ESM.

All research panels involved in the research process received consistent training through a shared slide-show introduction. This training covered the principles of randomised clinical trials, the importance of outcomes in trials and in general, and the validity of a COS repeated across trials [8]. The teaching material was based on a video developed by the COMET initiative – link.

The Danish research panels responded positively to the introduction. Similarly, the international research panels evaluated and commented on the educational material and the process with their own panels (ESM, page 33). Overall, the challenging part was to avoid the discussion of how to define and assess the core outcomes, which will be the next step. Another challenge was to clarify that the core outcomes were intended for use across all conceivable trials in the ICU and therefore it was necessary to adopt a meta-perspective when reflecting on the importance of each of the core outcomes.

**GRIPP2-SF Item 5; Reflections and critical perspective**

The Danish research panels were involved in most of the consensus discussions, except for Step 4 (Figure 1), where the 19 outcomes were discussed and condensed to 6 outcomes. The rationale behind this discussion is detailed on pages 23-32 of the ESM. Step 4 (Figure 1) was the only step with limited involvement, which could have influenced the core outcomes chosen. However, the discussion closely followed the results of the three round Delphi survey and the Step 3 (Figure 1) and discussion across all research panels. Any disagreements or additional outcomes are included in Table S10 in the ESM with further details provided in the rationale behind the Danish COS (ESM, page 23-32) and the summary from the international research panels (ESM, page 33).

Involving all 14 countries in all 6 steps (Figure 1) might have improved the conclusion. Nonetheless, the pragmatic process of involving countries to internationally validate the Danish findings did enhance the conclusion. The pragmatic approach proved to be a functional way to develop the international validation and will strengthen the continued development concerning definitions of the core outcomes, selection of appropriate instruments, and timing of their assessment.
